# MULTITRAIT ANALYSIS EXPANDS GENETIC RISK FACTORS IN CARDIOEMBOLIC STROKE

**DOI:** 10.1101/2021.12.10.21267609

**Authors:** Jara Cárcel-Márquez, Elena Muiño, Cristina Gallego-Fabrega, Natalia Cullell, Miquel Lledós, Laia Llucià-Carol, Tomás Sobrino, Francisco Campos, José Castillo, Marimar Freijo, Juan Francisco Arenillas, Victor Obach, José Álvarez-Sabín, Carlos A. Molina, Marc Ribó, Jordi Jiménez-Conde, Jaume Roquer, Lucia Muñoz-Narbona, Elena Lopez-Cancio, Mònica Millán, Rosa Diaz-Navarro, Cristòfol Vives-Bauza, Gemma Serrano-Heras, Tomás Segura, Laura Ibañez, Laura Heitsch, Pilar Delgado, Rajat Dhar, Jerzy Krupinski, Raquel Delgado-Mederos, Luis Prats-Sánchez, Pol Camps-Renom, Natalia Blay, Lauro Sumoy, Rafael de Cid, Joan Montaner, Carlos Cruchaga, Jin-Moo Lee, Joan Martí-Fàbregas, Israel Férnandez-Cadenas, On behalf GeneStroke Consortium and International Stroke Genetics Consortium

**Affiliations:** Stroke Pharmacogenomics and Genetics Group, Biomedical Research Institute Sant Pau (IIB Sant Pau), Barcelona, Spain; Universitat Autònoma de Barcelona, Departament de Medicina; Stroke Unit, Department of Neurology, Hospital de la Santa Creu i Sant Pau, Barcelona, Spain; Stroke Pharmacogenomics and Genetics Laboratory, Fundación Docència I Recerca Mútua Terrassa, Hospital Mútua Terrassa, Terrassa, Spain; Health Research Institute of Santiago de Compostela (IDIS), Clinical Neurosciences Research Laboratory, Hospital Clínico Universitario de Santiago de Compostela, Santiago de Compostela, Spain; Biocruces-Bizkaia Health Research Institute. Department of Neurology, Bilbao, Spain; Stroke Unit, Department of Neurology, University Hospital of Valladolid, Valladolid, Spain; Department of Neurology, Hospital Clínic de Barcelona, IDIBAPS, Barcelona, Spain; Stroke Unit, Department of Neurology, Hospital Universitari Vall d’Hebron, Barcelona, Spain; Department of Neurology, IMIM-Hospital del Mar; Neurovascular Research Group, IMIM (Institut Hospital del Mar d’Investigacions Mèdiques); Universitat Autònoma de Barcelona/DCEXS-Universitat Pompeu Fabra, Barcelona, Spain; Department of Neurosciences, Hospital Germans Trias I Pujol, Universitat Autònoma de Barcelona, Barcelona, Spain; Departament of Neurology, University Hospital Central de Asturias (HUCA). Oviedo, Spain; Department of Neurology, Son Espases University Hospital, Illes Balears Health Research Institute (IdISBa), Palma, Spain; Department of Neurology, University Hospital of Albacete, Albacete, Spain; Department of Psychiatry, Washington University School of Medicine, Saint Louis, MO, USA; Department of Emergency Medicine, Washington University School of Medicine, Saint Louis, MO, USA; Department of Neurology, Washington University School of Medicine, Saint Louis, MO, USA; Neurovascular Research Laboratory, Vall d’Hebron Institute of Research, Universitat Autònoma de Barcelona, Barcelona, Spain; Neurology Service, Hospital Universitari Mútua Terrassa, Terrassa, Spain; GenomesForLife-GCAT Lab Group, Germans Trias i Pujol Research Institute (IGTP), Badalona, Spain; High Content Genomics and Bioinformatics Unit, Germans Trias i Pujol Research Institute (IGTP), Badalona, Spain; Institute de Biomedicine of Seville, IBiS/Hospital Universitario Virgen del Rocío/CSIC/University of Seville & Department of Neurology, Hospital Universitario Virgen Macarena, Seville, Spain; Neurogenomics and Informatics Center at Washington University in St. Louis

## Abstract

**Background and Purpose:** The genetic architecture of cardioembolic stroke (CES) is still poorly understood. Atrial fibrillation (AF) is the main cause of CES, with which it shares heritability. We aimed to discover novel loci associated with CES by performing a Multitrait Analysis of the GWAS (MTAG) with atrial fibrillation genetic data.

**Methods:** For the MTAG analysis we used the MEGASTROKE cohort, which comprises European patients with CES and controls (n=362,661) and an AF cohort composed of 1,030,836 subjects. Regional genetic pleiotropy of the significant results was explored using an alternative Bayesian approach with GWAS-pairwise method. A replication was performed in an independent European cohort comprising 9,105 subjects using a Genome Wide Association Study (GWAS).

**Results:** MTAG-CES analysis revealed 40 novel and significant loci (p-value<5×10^−8^) associated with CES, four of which had not previously been associated with AF. A significant replication was assessed for eight novel loci: *CAV1, IGF1R, KIAA1755, NEURL1, PRRX1, SYNE2, TEX41* and *WIPF1*, showing a p-value<0.05 in the CES vs controls independent analysis. *KIAA1755*, a locus not previously described associated with AF. Interestingly, 51 AF risk loci were not associated with CES according to GWAS-pairwise analysis. Gene Ontology (GO) analysis revealed that these exclusive AF genes from the 51 loci participate in processes related mainly to cardiac development, whereas genes associated with AF and CES participate mainly in muscle contraction and the conduction of electrical impulses.

**Conclusions:** We found eight new loci associated with CES. In addition, this study provides novel insights into the pathogenesis of CES, highlighting multiple candidate genes for future functional experiments.

## INTRODUCTION

Cardioembolic stroke (CES) represents ≈25% of all ischemic strokes (IS), is responsible for the most severe ones and its prevalence could triple by 2050^1^. The main cause of CES is atrial fibrillation (AF), and the use of anticoagulants in AF patients could prevent ≈70% of strokes^1^, more than with antiplatelet treatment^2^. Moreover, occult paroxysmal AF could be the cause of up to 16% of undetermined strokes^3^. Nevertheless, not everybody with AF will suffer a stroke, and the use of anticoagulation involves an increased risk of major bleeding.

Therefore, understanding the mechanisms that are common to both CES and AF patients in relation to the risk of having a stroke would make it possible to develop specific, more effective therapies with fewer side effects.

To date, there are only eight loci associated with CES: *PITX2*^4^, *ZHFX3*^4^, *RGS7*^4^, *NKX2-5*^4^, *ABO*^4,5^, *GNAO1*^6^, *PHF20*^6^ and locus 5q22.3^6^. However, even the authors doubted the reliability of *RGS7* as a stroke-associated locus. Interestingly, the two most significant susceptibility loci (*PITX2* and *ZFHX3*) for CES are also the two most strongly associated with AF^4,7^; in fact CES and AF have common heritability ^8^.

Multitrait analysis of the GWAS (MTAG) integrates the information contained in correlated traits to boost genetic association signals for each individual trait, providing summary-level statistics^9^. This strategy has been successfully followed to identify novel loci associated with glaucoma^10^, bone mineral density^11^ and small vessel stroke^12^.

Our objective was to identify novel loci associated with CES and understand the biological mechanisms shared between CES and AF by performing a MTAG and additional GWAS analysis.

## METHODS

The data that support the findings of this study are available from the corresponding author upon reasonable request.

### Cohorts’ description

The summary statistics for the Cardioembolic Stroke analysis were obtained from the MEGASTROKE analysis (MEGASTROKE-CES) through the Cerebrovascular Disease Knowledge Portal (http://cerebrovascularportal.org). This cohort was composed of 7,193 CES patients and 355,468 controls of European ancestry. The summary statistics of the Atrial Fibrillation 2018 analysis (AF-2018) were obtained from the GWAS catalog portal (https://www.ebi.ac.uk/gwas/). The AF-2018 cohort was composed of 60,620 AF cases and 970,216 controls. The characteristics of the individuals in both studies are listed in the Online Supplement and publications^4,7^.

### Single Nucleotide Variation quality controls

A series of standard quality controls (QC) was applied to select the single nucleotide variants (SNVs) for the analysis. Variant exclusion criteria: 1) Not common to the summary statistics of the traits, 2) Minor allele frequency lower or equal to 0.01, 3) Missing values, 4) Negative standard error or not a number value, 5) p-value of 0, 6) Not SNVs, 7) Duplicated SNVs, 8) Strand ambiguity, and 8) Inconsistent allele pairs. After QCs, a total of 6,808,676 SNVs were selected (Online Figure I). Locus 15q21.3 prioritized genes *GCOM1* and *MYZAP* from AF-2018 was not evaluated due to absence of the significant SNVs of AF-2018 in the MEGASTROKE-CES analysis.

### Multitrait analysis of genome-wide association studies

We applied multitrait analysis of the GWAS (MTAG)^9^, analyzing MEGASTROKE-CES and AF-2018 summary statistics. We considered associations with CES to be significant if they attained a p-value<5×10^−8^ in the MTAG results for CES (MTAG-CES) and had a p-value<0.05 for association with CES in the MEGASTROKE study. Additionally, we tested whether the results from the MTAG-CES were consistent using an alternative Bayesian approach, to confirm pleiotropy of these loci using GWAS-pairwise^13^. This strategy reveals genomic regions that influence traits separately or pleiotropic regions that influence both traits. We used a posterior probability of pleiotropy (PPA pleiotropy) ≥0.6 to validate regions that influence both traits. Genomic inflation was estimated as lambda.

Loci associated exclusively with each trait were those that were considered significant in the MEGASTROKE-CES or AF-2018 and showed a PPA associated exclusively with one of the two traits ≥0.6 of the region mapped.

### Identification of independent and novel loci associated with CES

Independent loci were defined as those >1 megabase (Mb) apart in physical distance among SNVs with a genome-wide significance threshold of p-value<5×10^−8 7^. Loci were defined as novel when SNVs had an r^2^<0.1 compared with the index SNVs of the loci *PITX2*^4^, *ZFHX3*^4^, *NKX2-5*^4^, *RGS7*^4^, *ABO*^4,5^, *PHF20*^6^, *GNAO1*^6^ and 5q22.3^6^ that were GWAS significant in previous studies.

### Replication stage in an independent European cohort

We performed genome-wide analyses in an independent cohort of 9,105 individuals (3,479 IS and 5,625 controls). IS patients were recruited if they had a measurable neurologic deficit on the NIHSS within 6 hours of the last known asymptomatic status, had been diagnosed with stroke by an experienced neurologist, which had been confirmed by neuroimaging, were over 18 years of age and were recruited via hospital-based studies between 2003 and 2020 in Spain. Controls were subjects without a history of ischemic stroke, aged over 18 years, who declared they were free of neurovascular diseases before recruitment. The control cohort was collected in blood donation at primary care centers in Barcelona and in hospitals throughout Spain. An Institutional Review Board or Ethics Committee approved the study at each participating site. All patients or their relatives provided written informed consent. Further description of the cohorts is present in Online Supplement as well as array information, contribution of hospitals and clinical description (Online Tables I-III).

#### Quality control and imputation

DNA samples were genotyped on commercial arrays from Illumina (San Diego, CA) (Online Table II). Standard QCs were performed using PLINK v1.9 and KING v2.1.3 software. Imputation was performed in the Michigan Imputation Server Pipeline^14^ using Minimac4 And HRC r1.1 2016 panel. Further description of QCs and imputation are present in the Online Supplement.

#### Analysis

Analyses evaluated CES vs controls (CES = 1515, controls = 5626) and AF vs absence of AF (AF cases = 1110, AF controls = 7791). For the GWAS analyses we used an additive genetic model using fastGWA from gcta^15^. Age, sex and the first ten principal components were used as covariates. The results of the CES vs controls analysis were used to evaluate replicability (p-value<0.05) of the index SNVs and those significant SNVs (MTAG-CES p-value<5×10^−8^) in high LD (r^2^>0.8), in the GWAS analysis of CES and to perform a Pearson correlation of the effect sizes (log(OR)) of the MTAG-CES results for the index SNVs of the loci validated by GWAS-pairwise, a strategy used in a previous study^10^. The results of the AF analysis were used to evaluate novel loci associated with AF in the MTAG.

### Functional annotation and prioritization of genes

We used the HaploReg database to determine the functional annotation of the most strongly associated SNVs per locus. We also prioritized biological candidate genes generating a score based on: (1) nearest position to the index variant in a locus; (2) expression of the gene altered by the index variant in the eQTLGen^16^ or GTEx databases; (3) methylation of the gene altered by the index variant in Phenoscanner methylation data^17,18^; (4) whether the gene harbored a protein-altering index variant itself or in high linkage disequilibrium (LD)(r^2^>0.80); (5) whether the gene interacted physically with the index variant in HiC data of cardiomyocytes^19^. For the missense SNVs, we determined the likelihood that amino acid substitution has a deleterious effect on protein function using SIFT.

### Gene set analysis

We conducted a WebGestalt Overrepresentation Analysis of the selected prioritized genes associated in MTAG-CES. Gene Ontology (GO) of biological processes was performed, as well as a Benjamini Hochberg correction of the association p-value. We defined a biological process with a q-value<0.05 as statistically significant.

### Identification of potential drug repurposing targets

Prioritized genes were searched for in the DrugBank database^20^ to find potential candidate drugs for repurposing.

## RESULTS

### MTAG analysis of CES

After QC (Online Figure I), there were 6,808,676 common qualified SNVs from the AF-2018 and MEGASTROKE-CES cohorts. MTAG software revealed mean χ^2^ for AF-2018 and MEGASTROKE-CES in 1.39 and 1.12. The estimated equivalent GWAS sample size of the MTAG analysis for CES was of 861,823 individuals. A Manhattan plot of the MTAG-CES analysis and prioritized genes is shown in Figure 1; little evidence of genomic inflation was observed; lambda = 1.02. The MTAG-CES results revealed a total of 44 associated loci (p-value<5×10^−8^), 40 significant novel loci associated with CES and four previously found (Table 1). All loci previously associated in MEGASTROKE-CES (*ABO, NKX2-5, PITX2* and *ZFHX3*) are associated in this MTAG-CES, with the exception of the locus belonging to *RGS7* gene (top SNV rs146390073 MTAG p-value=0.001, AF-2018 p-value=0.98).

**Figure 1.**
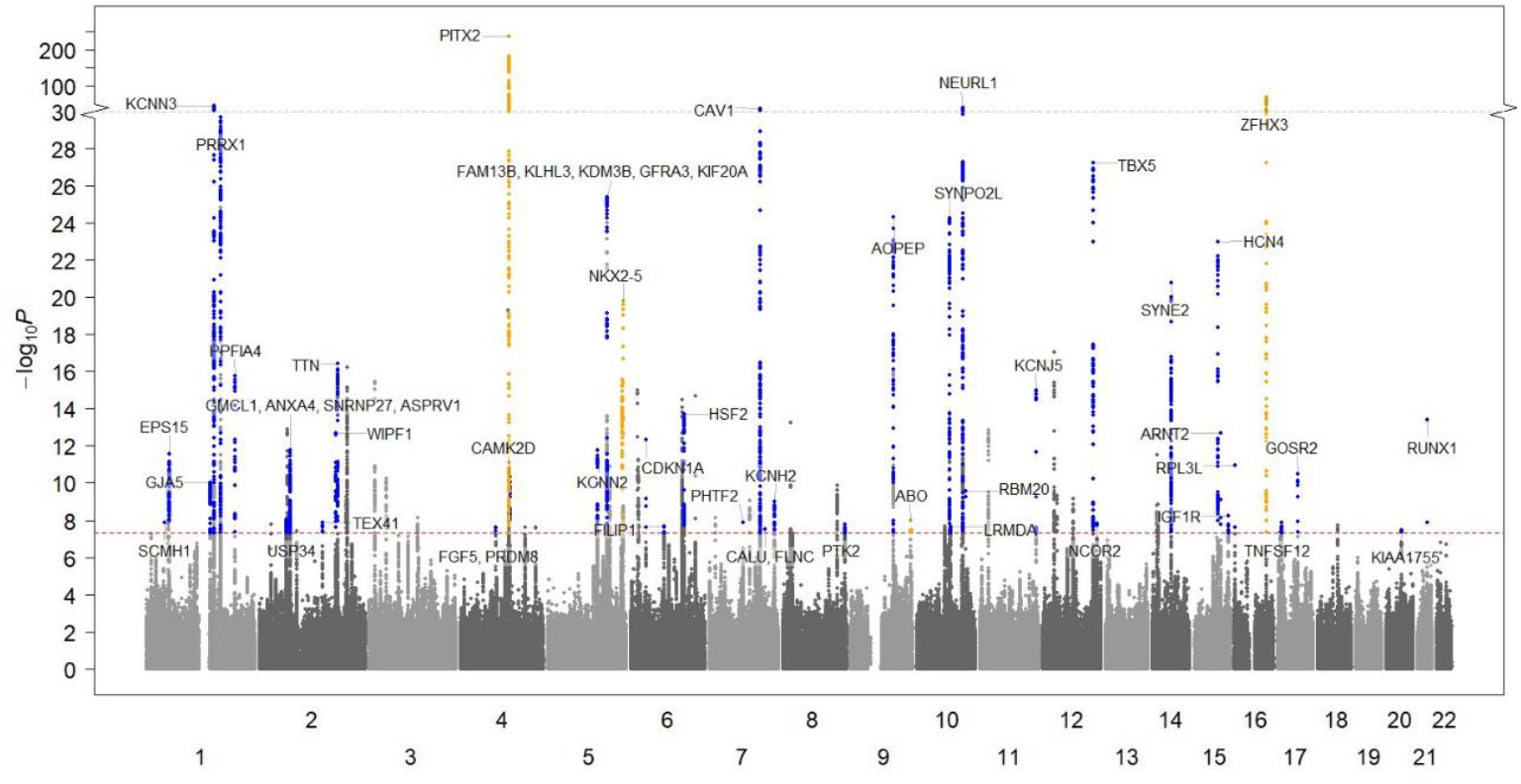
Manhattan plot of MTAG-CES. The X axis represents chromosome location, and the Y axis represents the minus logarithm on base 10 of p-value. The red line represents the GWAS-significance threshold. The novel loci are shown in blue and the established loci in yellow.

**Table 1.**
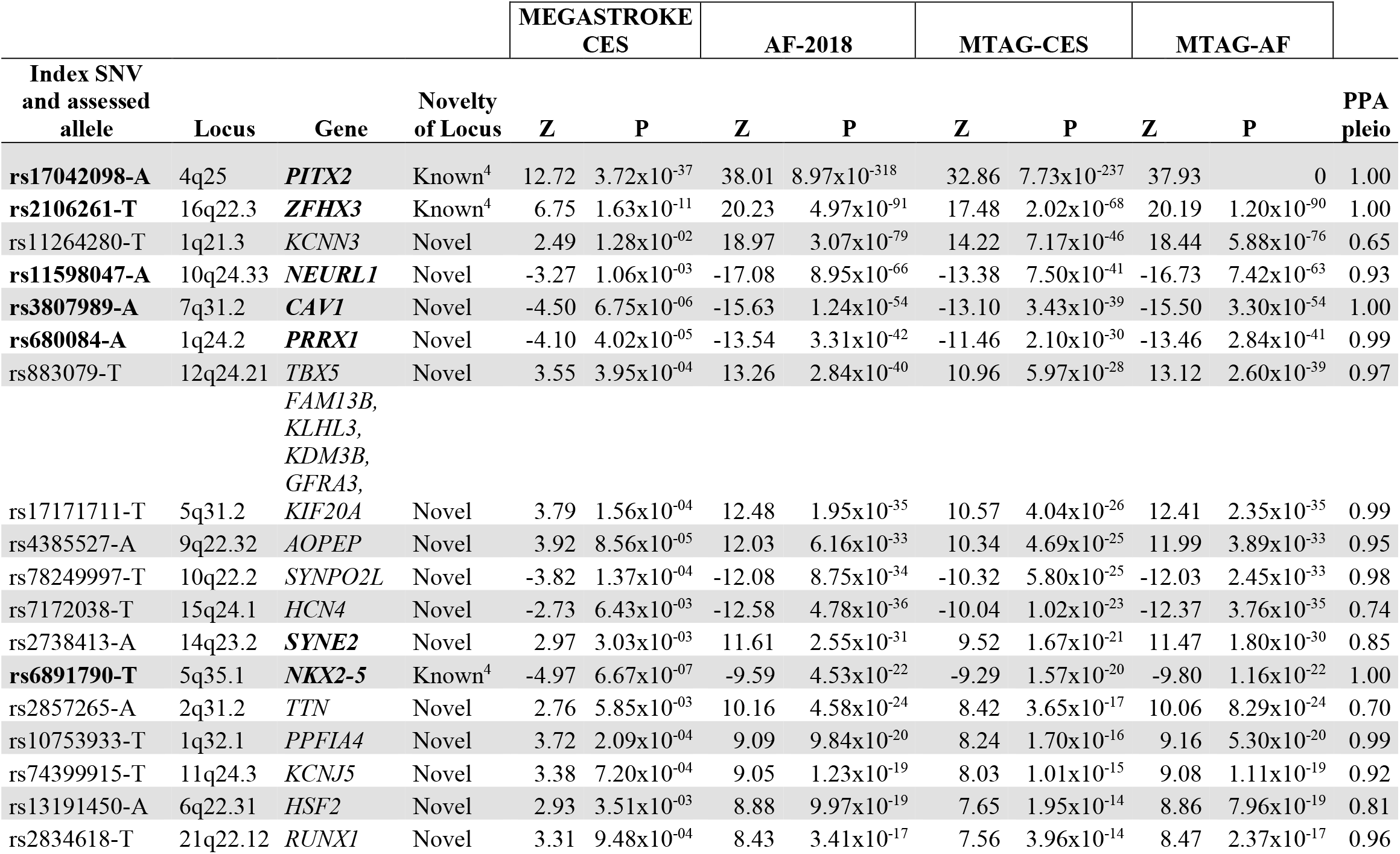
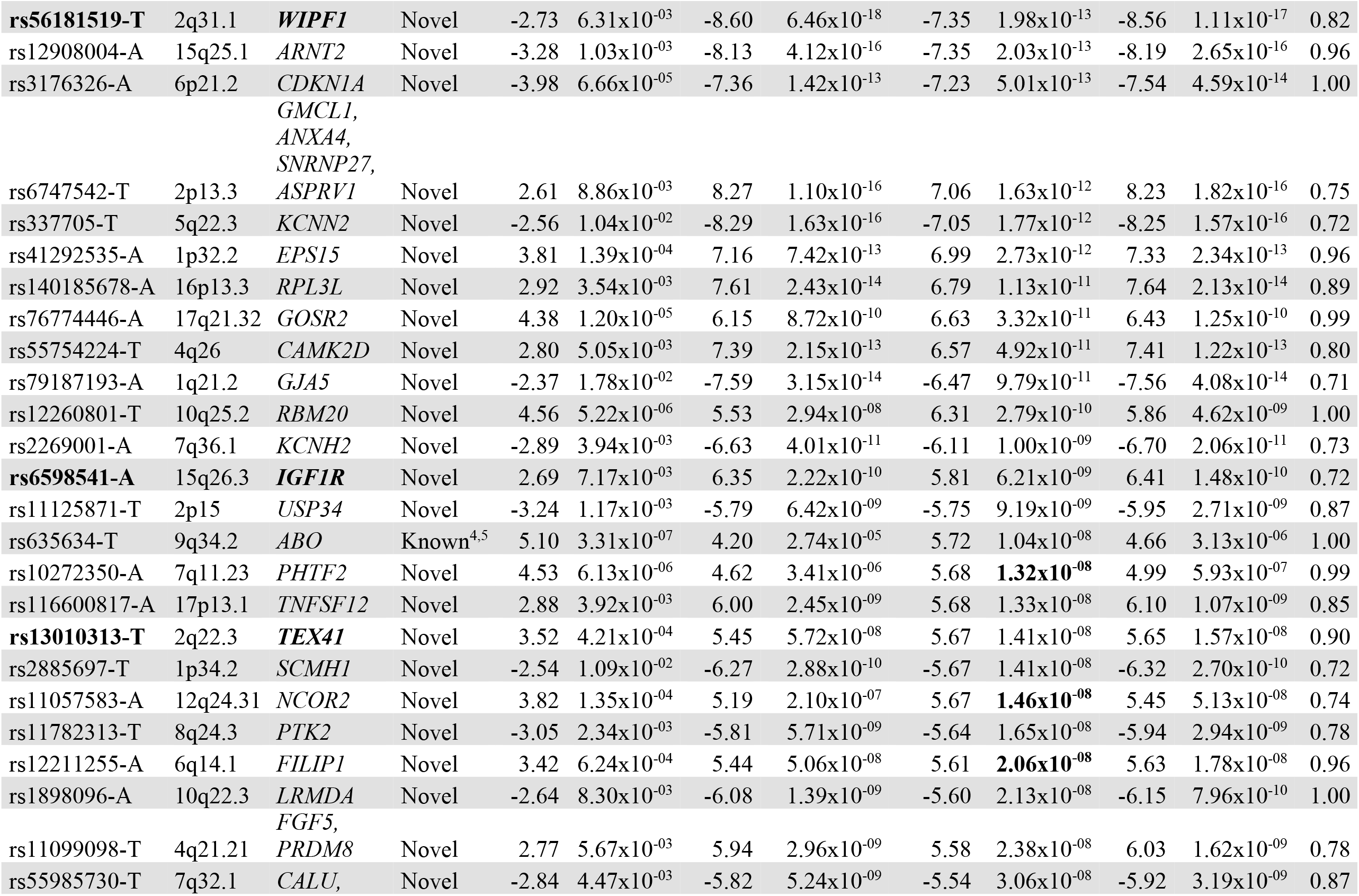
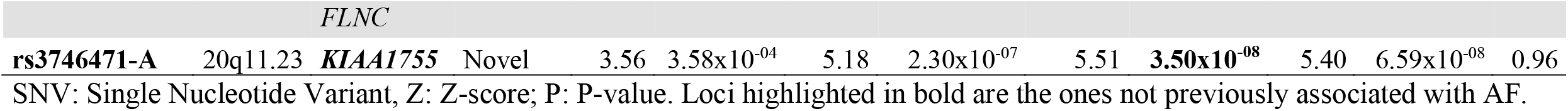
MTAG-CES results of the independent and significant loci.

Other loci found significant in GWAS of CES different from MEGASTROKE, belonged to: *PHF20, GNAO1* and 5q22.3 region. For two of them, the association was more significant in our analysis. For the 5q22.3 region, top SNV rs2169955 MTAG-CES p-value=4.76×10^−7^ vs MEGASTROKE-CES p-value=6.13×10^−3^, and for mapped gene *PHF20*, top SNV rs11697087 MTAG-CES p-value=6.62×10^−5^ vs MEGASTROKE-CES p-value=6.05×10^−4^. *GNAO1* was not evaluated in our study due to absence of the index SNV in AF-2018.

### New candidate loci associated with CES

After gene prioritization, 53 genes were selected from the 44 loci (Online Table IV). Novel loci showed a high degree of functionality of the SNVs as missense variants, eQTL, mQTLs, and HiC physical interaction (Online Tables V-VIII).

Replication analysis was performed in a new cohort of ischemic stroke patients and controls (n = 9,105). Evaluation of the index SNVs and SNVs in high LD revealed 56 SNVs with a p-value<0.05; these SNVs belong to 11 different loci, three already known *PITX2, ZFHX3, NKX2-5* and eight of them novel, prioritized genes: *CAV1, IGF1R, KIAA1755, NEURL1, PRRX1, SYNE2, TEX41* and *WIPF1* (Online Table IX). *NEURL1* locus was already found associated in the all types of stroke analysis of MEGASTROKE but not genome-wide significant in CES analysis ^4^.

Pearson correlation of the effect sizes (log (OR)) between index variants of the significant loci in the MTAG-CES and the CES analysis in an independent cohort was evaluated for 41 of the 44 index variants, due to the poor imputation value (INFO<0.6) of three of them in the Spanish cohort. A significant correlation was obtained; p-value=2.7×10^−8^, cor=0.74 (Figure 4). This association remained after removing the four SNVs from previously known loci (rs17042098, *PITX2*; rs2106261, *ZFHX3*; rs6891790, *NKX2-5* and rs635634, *ABO*) p-value=4.2×10^−7^, cor=0.72.

The top five most significant loci in the MTAG-CES analysis were *PITX2* and *ZFHX3* (previously known loci), and *KCNN3, NEURL1*, and *CAV1* (novel loci), the last two validated in the replication cohort. Interestingly, we found loci not previously described in AF or CES with four prioritized genes: *PHTF2, KIAA1755, NCOR2*, and *FILIP1. FILIP1* locus also reached genome-wide significance in the MTAG results for AF. Functional annotation of the index SNVs revealed rs3746471 as a missense variant of the *KIAA1755* gene coding for R1045W, and it was predicted to be deleterious with a SIFT score of 0.007.

The *FILIP1* index variant was additionally evaluated in the AF GWAS in the independent cohort, revealing a suggestive p-value with consistent direction of the effect of this novel association with AF (rs12211255-A, beta(se)=0.013(0.007), p-value=0.09).

### Biological processes of loci associated with CES and AF and biological processes of loci associated exclusively to AF

GO of biological processes from the Genome-Wide loci of the MTAG-CES analysis revealed 85 enriched gene sets (Figure 2, Online Table X); the top biological processes were: cardiac muscle contraction, actin filament-based movement and cardiac conduction. A DrugBank search for the 53 prioritized genes associated with CES revealed a total of 87 drugs (Online Table XI) that target 10 of the prioritized genes.

**Figure 2.**
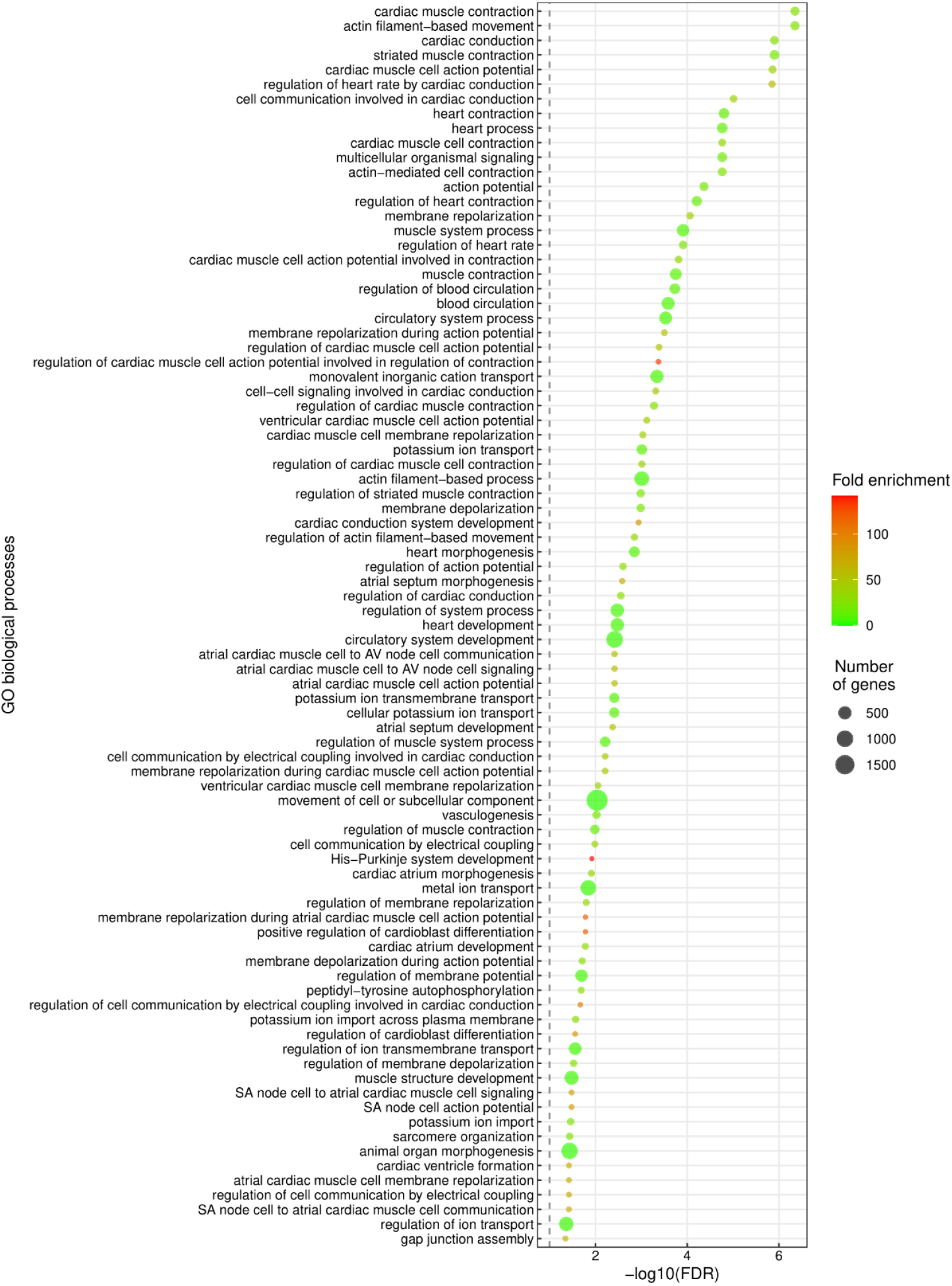
GO Biological processes enriched in CES prioritized gene set.

The study of the 111 AF-2018 significant loci (Online Table XII) using GWAS-pairwise strategy suggests 51 loci that have an exclusive association with AF risk and a lack of association with CES. A biological processes analysis of the genes associated exclusively with AF (Figure 3, Online Table XIII) revealed 25 biological processes exclusive to AF risk and mainly associated with cardiac development processes (Figure 2, Online Table XIV).

**Figure 3.**
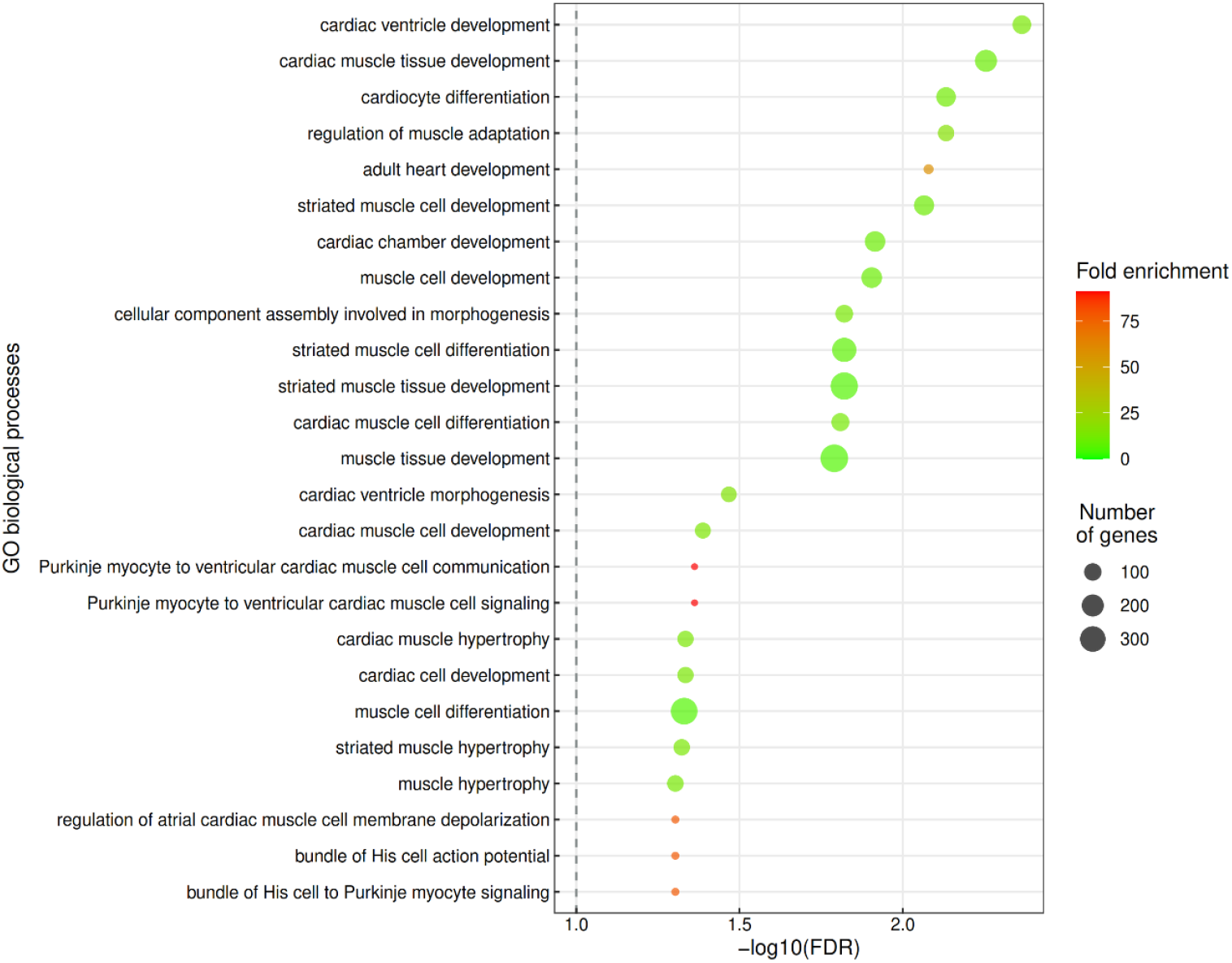
GO Biological processes enriched exclusively in analysis of AF associated genes independently of CES risk.

**Figure 4.**
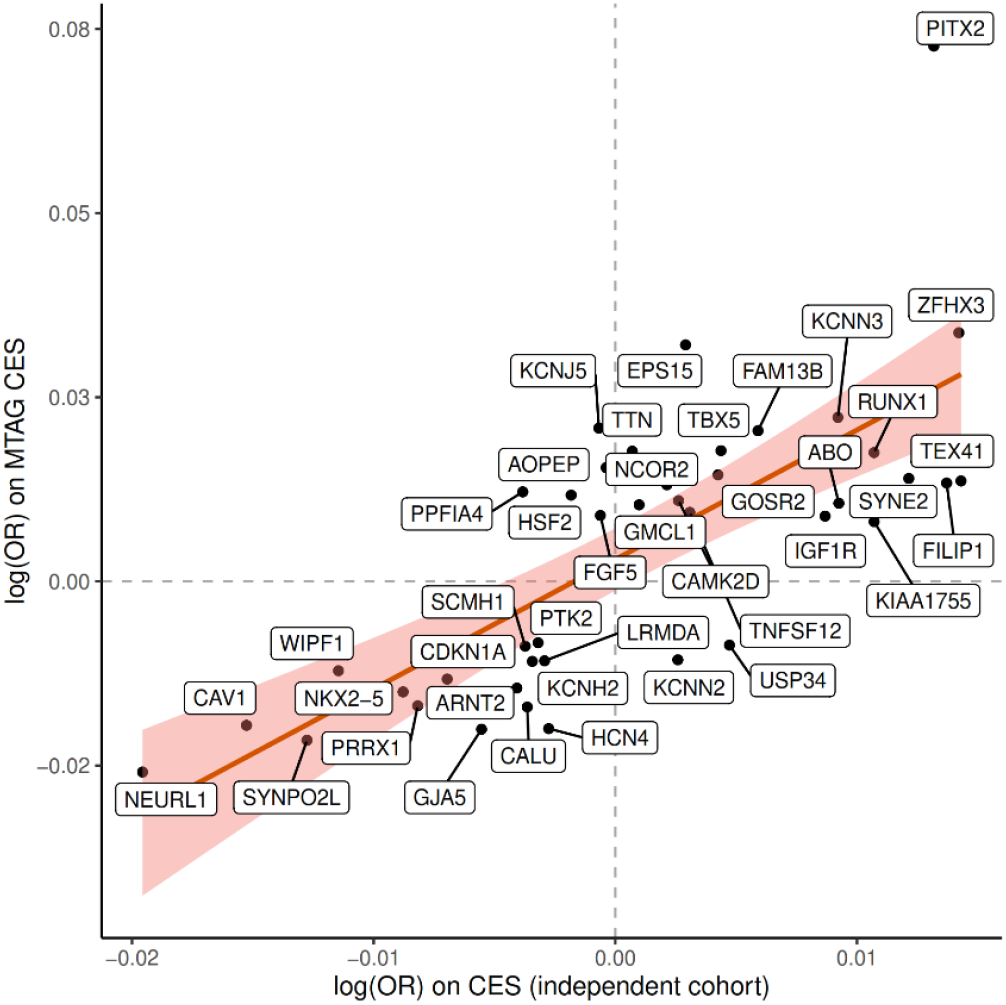
Comparison of the effect sizes (log ORs) for 41 genome-wide significant index SNVs identified from the MTAG-CES analysis versus those in CES analysis of the Spanish independent cohort.

## DISCUSSION

Using a multitrait analysis (MTAG) with the two biggest cohorts of CES^4^ and AF^7^ to date, we expanded the number of loci associated with CES from eight to sixteen. In the MTAG analysis, 44 loci were found associated with CES. Four of them were previously known associations: *PITX2, ZFHX3, NKX2-5* and *ABO*. To explore replicability of the MTAG results, we conducted genome-wide analysis for CES and AF in an independent European cohort comprising 9,105 subjects. We found significant associations (p-value<0.05) of 56 SNVs from 11 loci (eight of them novel associations); prioritized known genes: *PITX2, ZFHX3, NKX2-5*, prioritized novel genes: *CAV1, IGF1R, KIAA1755, NEURL1, PRRX1, SYNE2, TEX41* and *WIPF1*, in the CES analysis.

Moreover, given the small sample size of the replication stage compared with the estimated sample size of the MTAG, we did not expect all the SNVs to be strongly associated, but rather that the odds ratios would be highly concordant, for that we performed Pearson correlation of the effect sizes for the index SNVs in MTAG-CES analysis. The Pearson correlation revealed a positive and significant correlation in this new cohort, showing most of the SNVs in the expected direction of the effect. Following the methodology used in other similar studies^10^, the presence of this significant correlation suggests potential validation of our MTAG-CES results.

Interestingly, *CAV1* has been associated with platelet count and systolic blood pressure, among other traits^21^. Besides, caveolin-1, the protein encoded by *CAV1* in endothelial cells, plays a key role in how gas6 exerts its prothrombotic role in the vasculature^22^. Regarding the other genes, and among other human traits in GWAS, *IGF1R* and *WIPF1* have been found associated with QRS duration^23,24^ and platelet crit^25,26^. *NEURL1* is associated with white matter hyperintensity volume^27^, red blood cell count^28^ and to all types of stroke risk ^4^. *PRRX1, SYNE2* and *TEX41* have been associated with PR interval^29–31^ and *SYNE2* also associated with platelet count^30^. It is worth noting, that of these replicated loci, locus 20q11.23, prioritized gene: *KIAA1755*, has not previously been found associated with AF. The index variant of this locus, rs3746471-A, codes for R1045W amino acid change, predicted to be deleterious according to SIFT. rs3746471-A, has been previously described associated with heart rate^32–34^ and PR interval^34^, and remarkably suggestively associated with stroke infarct volume (p-value=6.80×10^−7^)^35,36^. *KIAA1755* is predicted to encode an uncharacterized protein and is only characterized at the transcriptional level. The transcript is highly expressed in the brain and nerves and is also expressed in the heart.

These parallel associations of the replicated loci highlight the role that these genes could be playing in the risk conferring to CES, via altering AF, platelet count, coagulation cascade and different hematological traits.

Using MTAG strategy, we were also able to analyze the output of AF-2018 enhanced by MEGASTROKE-CES. In this direction, a novel locus, 6q14.1, was found significantly associated with AF. *FILIP1* is the gene prioritized; it encodes for a filamin A binding protein. Interestingly, *FILIP1* has been identified previously as a regulator of myogenesis differentiation, in human cells and in an *in vivo* mouse model^37^. In the replication stage, this SNV was found suggestive (p-value=0.09), highly probable due to the small sample size in comparison to MTAG analysis.

The study of previous known loci associated with CES in the MTAG-CES analysis showed that the *RGS7*^4^ previously genome-wide significant in the MEGASTROKE was not associated with CES in our study. This could be explained by a causal mechanism other than AF in the risk of suffering CES or the possibility of a false positive result, as authors suspected in the MEGASTROKE study given the low allele frequency and poor imputation^4^. The evaluation of loci found in GWAS of CES different from MEGASTROKE (*PHF20, GNAO1* and 5q22.3 region), revealed 5q22.3 and *PHF20* loci with stronger significance in our study. This could be considered a reinforcement of these two signals and their relation to CES risk via AF. Sadly, *GNAO1* was not evaluated due to the lack of the index variant in AF-2018.

Globally, from the 44 loci genome-wide associated in the MTAG-CES, 53 genes were prioritized in our study: the top previously known genes were *PITX2* and *ZFHX3* and other top novel genes were *NEURL1* and *CAV1*. The prioritized genes were involved in biological processes, such as cardiac muscle contraction, actin filament-based movement, cardiac conduction, and striated muscle contraction, which highlights their role in the risk of CES due to AF.

Remarkably, among the 111 loci previously associated with AF, a group of 51 loci had an exclusive genetic association with AF and a lack of association with CES using GWAS-pairwise. Apparently, no statistical differences were observed between the AF loci associated with CES and those with no association. GO of biological processes revealed 43 gene sets that were only enriched by the AF exclusive genes and not present in the biological processes of the 44 loci associated with CES; these biological processes were related mainly to cardiac development. These genes exclusively associated with AF could be important for understanding the risk of CES occurrence, and thereby help to develop more specific prevention drugs or to create better genetic scores for identifying AF patients with high risk of suffering a stroke.

Our study has limitations, as there is a known overlap of individuals between the MEGASTROKE-CES and AF-2018 studies. However, MTAG permits sample overlap between the GWAS summary data of the traits by conducting bivariate LD score regression of the traits^9^. Additionally, the difference in the sample size between the two original studies is a concern, on the other hand MTAG estimation of χ^2^ revealed an scenario expected to be strong against false positives, as tested in the original publication^9^. To control this, we performed a replication analysis in an independent cohort to validate the results obtained. Another limitation is that the MTAG approach relies on the relatively strong assumption that associated variants act on both traits, which may not always be the case for CES and AF. To control this, we only considered SNVs reaching the GWAS significant p-value, showing an association with both traits and having a PPA pleiotropy of the genomic region =>0.6 in the GWAS-pairwise. In addition, we performed a replication step in an independent cohort to further evaluate these loci. Although this replication is objectively weak compared to the MTAG analysis, we hypothesize that more loci found in our study will be validated when an equivalent cohort in the future is analyzed. It is also important to mention that through this strategy we have only found loci associated to CES risk due to AF, and therefore further multitrait analysis should be performed with different traits to uncover the different high-risk sources of CES.

To summarize, in our study, we identified 40 novel loci associated with CES through a multitrait analysis enhanced by AF, and we were able to replicate eight of them in an independent cohort. This study provides novel insights into the pathogenesis of CES, its relation to AF and highlights multiple candidates for use in future functional experiments to identify specific mechanisms conferring a risk of CES that could be targeted therapeutically.

## Data Availability

All data produced in the present study are available upon reasonable request to the authors

## Non-standard Abbreviations and Acronyms

AF: Atrial fibrillation
CES: Cardioembolic stroke
GWAS: Genome-wide association study
IS: Ischemic stroke
MTAG: Multitrait analysis of the GWAS
SNP: Single nucleotide polymorphism
QC: Quality control

## ACKNOWLEDGMENTS

The Genotype-Tissue Expression (GTEx) Project was funded by the Common Fund of the Office of the Director of the National Institutes of Health, and by NCI, NHGRI, NHLBI, NIDA, NIMH, and NINDS. The data used for the analyses described in this manuscript were obtained from the GTEx Portal on 03/30/20. This study uses data generated by the GCAT | Genomes for Life. Cohort study of the Genomes of Catalonia, IGTP. A full list of the investigators who contributed to the generation of the data is available from http://www.genomesforlife.com/. IGTP is part of the CERCA Program/Generalitat de Catalunya.

