## Supplementary material for "MULTITRAIT ANALYSIS EXPANDS GENETIC RISK FACTORS IN CARDIOEMBOLIC STROKE": Online

### ONLINE SUPPLEMENT

#### 1. DETAILED METHODS

##### 1.1. Cohorts used in Multitrait analysis of GWAS (MTAG)

**MEGASTROKE-CES**

For the European ancestry analysis of MEGASTROKE consortium 16 different cohorts were analyzed, comprising up to 34,217 cases of ischemic stroke and 405,111 healthy controls. Stroke was defined according to the World Health Organization (WHO) as rapidly developing signs of focal (or global) disturbance of cerebral function, lasting more than 24 hours or leading to death with no apparent cause other than that of vascular origin. Strokes were defined as ischemic stroke (IS) or intracerebral hemorrhage (ICH) based on clinical and imaging criteria. IS was further subdivided into the following categories mostly using the Trial of Org 10172 in Acute Stroke Treatment (TOAST) criteria): i) large vessel ischemic stroke; ii) cardioembolic ischemic stroke; iii) small vessel ischemic stroke. Specifically, for European analysis of CES they analyzed 7,193 cases of CES and 355,468 healthy controls. Further details of the cohorts in the original publication [1].

**AF-2018 study**

A total of 60,620 cases of AF and 970,216 controls were analyzed, these patients were recruited as part of main cohorts:

HUNT.—The Nord-Trøndelag Health Study (HUNT) is a population-based health survey conducted in the county of Nord-Trøndelag, Norway from 1984 to 2009 [2]. They used a combination of hospital, out-patient, and emergency room discharge diagnoses (ICD-9 and ICD-10) to identify 6,493 atrial fibrillation cases and 63,142 atrial fibrillation-free controls with genotype data. Participation in the HUNT Study is based on informed consent, and the study was approved by the Data Inspectorate and the Regional Ethics Committee for Medical Research in Norway.

deCODE.—The Icelandic atrial fibrillation population consisted of all patients diagnosed with atrial fibrillation (ICD-10 code I48 and ICD-9 code 427.3) at Landspitali, The National University Hospital, in Reykjavik, and Akureyri Hospital (the two largest hospitals in Iceland) from 1987 to 2015. All atrial fibrillation cases, a total of 13,471, were included. Controls were 358,161 Icelanders recruited through different genetic research projects at deCODE genetics, excluding those in the atrial fibrillation cohort. The study was approved by the Icelandic Data Protection Authority and the National Bioethics Committee of Iceland (no. VSNb2015030021).

MGI.—MGI is a hospital-based cohort collected at Michigan Medicine, USA. Atrial fibrillation cases (n = 1,226) were defined as patients with ICD-9 billing code 427.31, and controls were individuals without atrial fibrillation, atrial flutter, or related phenotypes (ICD-9 426–427.99). MGI was reviewed and approved by the Institutional Review Board of the University of Michigan Medical School.

DiscovEHR.—The DiscovEHR collaboration cohort is a hospital-based cohort including 58,124 genotyped individuals of European ancestry from the ongoing MyCode Community Health Initiative of the Geisinger Health System, USA[3]. Atrial fibrillation cases (n = 6,679) were defined as DiscovEHR participants with at least one electronic health record problem list entry or at least two diagnosis code entries for two separate clinical encounters on separate calendar days for ICD-10 I48: atrial fibrillation and flutter. Corresponding controls (n = 41,803) were defined as individuals with no electronic health record diagnosis code entries (problem list or encounter codes) for ICD-10 I48. The Study was approved by the Geisinger Institutional Review Board.

UK Biobank.—The UK Biobank is a population-based cohort collected from multiple sites across the United Kingdom[4]. Cases of atrial fibrillation were selected using ICD-9 and ICD-10 codes for atrial fibrillation or atrial flutter (ICD-9 427.3 and ICD-10 I48). Controls were participants without any ICD-9 or

ICD-10 codes specific for atrial fibrillation, atrial flutter, other cardiac arrhythmias, or conduction disorders.

AFGen Consortium.—Published atrial fibrillation association summary statistics from 31 cohorts representing 17,931 atrial fibrillation cases and 115,142 controls were obtained from the authors.

**1.2 Single Nucleotide Variants (SNVs) quality control.**

A series of standard quality controls were applied to select the single nucleotide variants (SNVs) for the MTAG analysis. Variant exclusion criteria: 1) Not common to the summary statistics of the traits, 2) Minor allele frequency lower or equal to 0.01, 3) Missing values, 4) Negative standard error or not a number value. 5) P-value of 0, 6) Not SNVs, 7) Duplicated SNVs, 8) Strand ambiguity, and 8) Inconsistent allele pairs. After QCs, a total of 6,808,676 SNVs were selected (eFigure 1). Locus 15q21.3 prioritized genes *GCOM1* and *MYZAP* for AF-2018 was not evaluated due to absent of the significant SNVs of AF-2018 in the MEGASTROKE-CES analysis.

**1.3. Replication GWAS in GENERACION cohort**

**Study population**

We performed analysis in GENERACION study cohort (Spain). 9,105 individuals (3,479 IS and 5,626 controls). IS patients were recruited via hospital-based studies between 2003 and 2020. The participants were part of the Genetics of Early Neurological Instability After Ischemic Stroke (GENISIS), Genetic contribution to Functional Outcome and Disability after Stroke (GODS), the Genetic Study in Ischemic Stroke Patients treated with tPA (GenoTPA), the CONtrol ICtus (CONIC), and SEDMAN studies. Controls were subjects without a history of ischemic stroke, aged over 18 years, who declared they were free of neurovascular diseases before recruitment. The control cohort was collected in blood donation at primary care centers in Barcelona and in hospitals throughout Spain as a part of the GCAT, CONtrol ICtus (CONIC), Investigating Silent Stroke in hYpertensives: A magnetic resonance imaging Study (ISSYS) and the Genotyping Recurrence Risk of Stroke (GRECOS) projects. Array information, contribution of hospitals and clinical description of the cohort are present in Online Tables I-III.
The ischemic stroke patients were recruited if they had a measurable neurologic deficit on the NIHSS within 6 hours of the last known asymptomatic status, had been diagnosed with stroke by an experienced neurologist, which had been confirmed by neuroimaging and were over 18 years of age.
These patients were recruited as part of the GENISIS[5], GODS[6], and CONIC[7] projects.
Controls were subjects without a history of ischemic stroke, aged over 18 years, who declared they were free of neurovascular diseases before recruitment. The control cohort was collected in blood donation and primary care centers in Barcelona and in hospitals throughout Spain as a part of the GCAT[8] (Galván-Femenia, I, 2018), CONIC[7], GRECOS[9], and ISSYS[10] projects.

Description of the cohorts included:
GENISIS cohort.—Genetics of Early Neurological Instability after Ischemic Stroke (GENISIS)[5] is an international study currently recruiting patients from four different locations: United States, Finland, Poland, and Spain. The inclusion criteria for the GENISIS study are IS patients (age ≥ 18 years) Collected from 2003 to 2016 with a measurable neurologic deficit on the NIHSS within 6 hours of last known normal. Patients who received endovascular thrombectomy, or for whom consent and/or a blood sample could not be obtained were excluded. For our study we only include Spanish patients. Genotyping was performed with Human Core Exome chip (Illumina®).
GODS cohort.—Genetic contribution to functional Outcome and Disability after Stroke (GODS)[11] project is a study that aim find genetic factors associated to stroke outcome. All participants met the following criteria: (1) European descent, aged >18 years, diagnosis of IS in the anterior vascular territory; (2) assessed

by a neurologist during the acute phase of stroke; (3) initial stroke severity >4, according to the National Institutes of Health Stroke Scale (NIHSS); (4) information on post-stroke functional status at 3 months (or alternatively between 3-6 months); (5) evidence of acute IS in a neuroimaging study; (6) lack of concomitant pathology. Individuals with stroke recurrence during the follow-up period were excluded, as well as, posterior vascular territory and lacunar strokes. Samples were genotyped at the Genetic and Molecular Epidemiology Laboratory of McMaster University (David Braley Research Institute) in Ontario, Canada, with Human Core Exome chip (Illumina®).

CONIC cohort.—CONtrol ICtus (CONIC) study[7] is a national study that recruited controls and IS cases participants in Vall d'Hebron Hospital between 2007 and 2008. All controls were older than 65 years of age and declared free of dementia, neurovascular and/or cardiovascular disease, as evaluated by self-description during a direct interview before recruitment. Subjects with a history of first and/or second-degree neurovascular disorder were also excluded from the study.

The IS cases were admitted to the emergency department of a university hospital who had a documented middle cerebral artery (MCA) occlusion on transcranial Doppler ultrasonography (TCD) and received tPA in a standard 0.9-mg/kg dose (10% bolus, 90% continuous infusion during 1 hour) within 3 hours of symptom onset following National Institute of Neurological Disorders and Stroke (NINDS) recommendations. Cases and controls were genotyped with Human Core Exome chip (Illumina®).

GRECOS cohort.—Genotyping RECurrence Risk Of Stroke (GRECOS)[9] project is a national study that aim find genetic factors associated with the recurrence after stroke. Control participants were selected from relatives of patients (wife or husband, without any consanguinity among cases and controls) and healthy volunteers visiting the same hospital for routine testing. They were >65 years of age and classified as free of neurovascular and cardiovascular history and familial history of stroke by direct interview before recruitment. All samples were genotyped with Human Core Exome chip (Illumina®).

ISSYS cohort.—Investigating Silent Stroke in hYpertensives: A magnetic resonance imaging Study (ISSYS)[10] is an observational prospective study in hypertensive participants to determine the prevalence of silent or magnetic resonance imaging (MRI)–defined brain infarcts and cognitive impairment. This cohort comprises 1000 non-demented individuals, aged 50 to 70 years old, and diagnosed of essential hypertension at least one year before inclusion in the ISSYS study. Those individuals were genotyped with Human Core Exome chip (Illumina®).

GCAT cohort.—GCAT health databank is a collection of health data and samples from participants of the "GCAT/Genomes for Life. Cohort Study of the Genomes of Catalonia Study"[8]. The aim of the GCAT project was study the genetic and environmental factors that lead to the appearance of chronic diseases in the general population. The study is conducted in several waves of data gathering, namely GCAT1, the baseline Survey from 2014-2017 and GCAT2, the GCAT follow-up in the second year. Data collection is done with web-based self-questionnaires, direct interviews, clinical data, and analyses of DNA blood derived samples. Genome-wide genotypes have been generated using Illumina Infinium SNV-bead array technology using the Multi-Ethnic Global (MEGAEX, V.2) consortium array. We used only GCAT1 genotyped patients and we exclude individuals with heart infarct or heart diseases or with non-Caucasian ancestry.

genotPA

Consecutive Caucasian patients with acute ischemic stroke who were admitted to the emergency room and received recombinant tissue-type plasminogen activator (r-tPA) within 4.5 hours of symptom onset were recruited. Patients were enrrolled at Spanish hospitals (Vall d'Hebron University Hospital, Hospital Clinic, Hospital Universitari de Girona Doctor Josep Trueta, Hospital de la Santa Creu i Sant Pau, Hospital Universitari Germans Trias I Pujol, Hospital Universitari del Mar, Hospital de Basurto) between 2002 to 2012. The study protocol was approved by the Ethics Committee of each center, all patients or relatives signed the informed consent.

Patients were identified by medical evaluation at emergency room arrival; stroke diagnosis was performed by trained neurologists and confirmed by neuroimaging. There were no exclusion criteria regarding age, sex or ethnicity. Follow-up CT scan at 24 hours after onset of symptoms or if neurological worsening occurred were performed and was classified according to European Cooperative Acute Stroke Study (ECASS) [12].

SEDMAN cohort. - patients ≥ 18 years old, treated with acenocoumarol or dabigatran for stroke or systemic embolism prevention following the local recommendations. All patients had a stroke or transient ischemic attack (TIA) during the previous 14 days before the initiation of anticoagulation treatment and had a diagnostic of non-valvular atrial fibrillation. Only patients with mild to moderate stroke (less than 2/3 of the vascular territory) with initial Alberta Stroke Program Early CT Score (ASPECTS) in the first CT/MRI > 6 and National Institute of Health Stroke Scale (NIHSS) < 25 were included. All patients had a general condition which allowed the 12 months' follow-up. Only patients with stablished stroke were used in this analysis.

**Quality control and imputation**

DNA samples were genotyped on commercial arrays from Illumina (San Diego, CA) (Online Table II). Quality controls were performed using PLINK v1.9 and KING v2.1.3 software. For all datasets, samples were excluded if there was a mismatch between the genetic and reported sex, genotype call rate lower than 95%, excess or loss of heterozygosity, non-European detected as outliers of 1000 Genomes Project Phase 3 dataset (1000G), duplicate samples or relatedness at a PI-HAT>0.20. SNPs were excluded if call rate lower than 95%, located in non-autosomes, non-biallelic, strand ambiguous, monomorphic or were deviated from Hardy-Weinberg equilibrium (p-value $<10^{-6}$ in controls, $<10^{-10}$ in IS).

Imputation was performed in the Michigan Imputation Server Pipeline[13] using Minimac4. HRC r1.1 2016 (GRCh37/hg19) was the reference panel used, with European population, and for phasing Eagle v2.4 was used. After imputation, we removed SNV with imputation score <0.6 and MAF <1%. SNVs that were not present in at least 95% of the individuals were removed.

**Samples**

DNA for all subjects were obtained from whole blood samples. A total of 10,066 samples were genotyped in the study. After QCs 9,105 samples fulfilled the QC criteria and were not missing for the phenotypes and covariates analyzed: CES, AF, sex, and age.

**Replication-stage in an independent European cohort**

*Post-analysis quality controls*

After analysis using fastGWA we removed variants with minor allele frequency < 1%, minor allele count in cases or controls < 6 and variants that deviated from Hardy Weinberg equilibrium p-value; $< 1x10^{-6}$. Additionally, genomic inflation was estimated as lambda.

*Visual inspection of the analysis*

Manhattan and QQ plots were performed and can be observed in Online Figure II.

#### 2. ONLINE FIGURES

**Online Figure I. Workflow of the SNVs for the AF-2018 and MEGASTROKE-CES datasets**. NaN: Not a number. SNV: Single Nucleotide Variant.

**Online Figure II. Manhatttan and QQ plots con CES and AF analysis in the independent cohort.** The X axis represents chromosome location, and the Y axis represents the minus logarithm on base 10 of p-value (-log 10 (p-value)).

**Online Figure I.** Workflow of the SNVs for the AF-2018 and MEGASTROKE-CES datasets. NaN: Not a number. SNV: Single Nucleotide Variant.

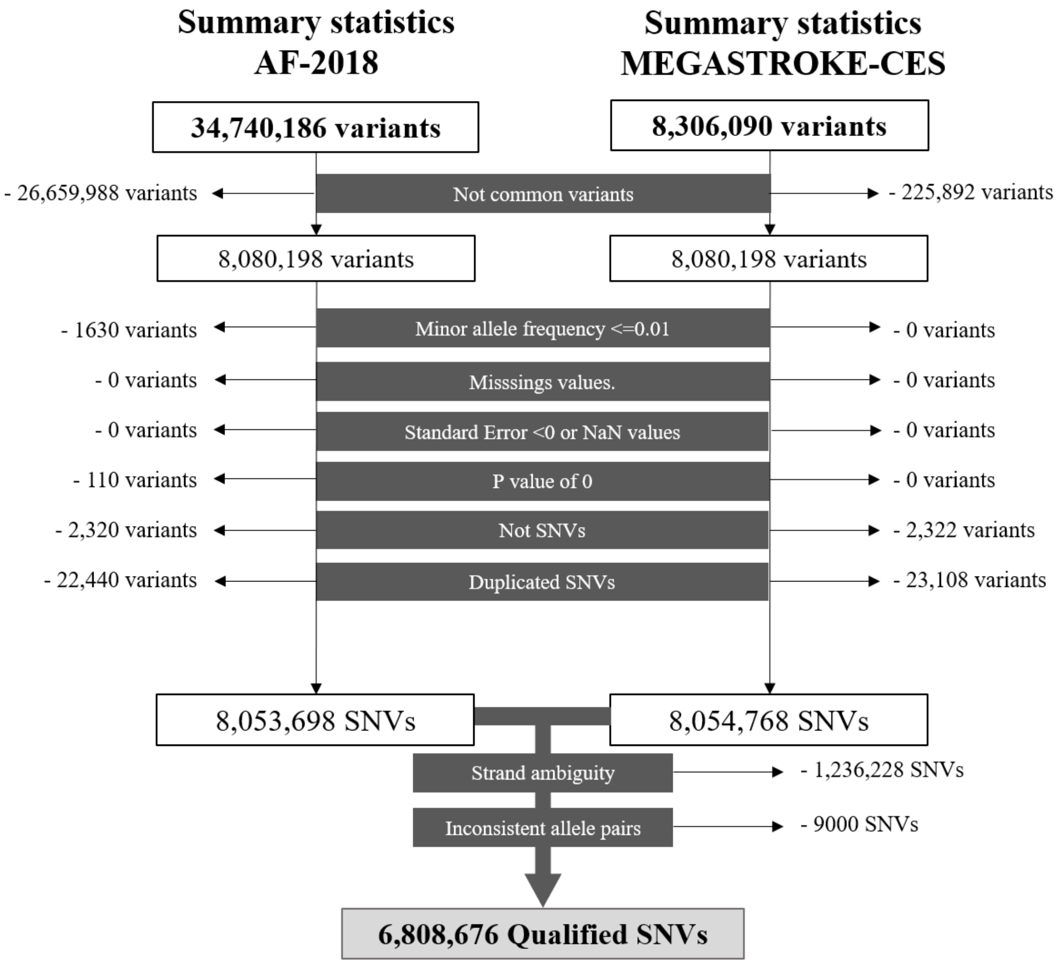

**Online Figure II. Manhatttan and QQ plots con CES and AF analysis in the independent cohort.** The X axis represents chromosome location, and the Y axis represents the minus logarithm on base 10 of p-value (-log 10 (p-value)).

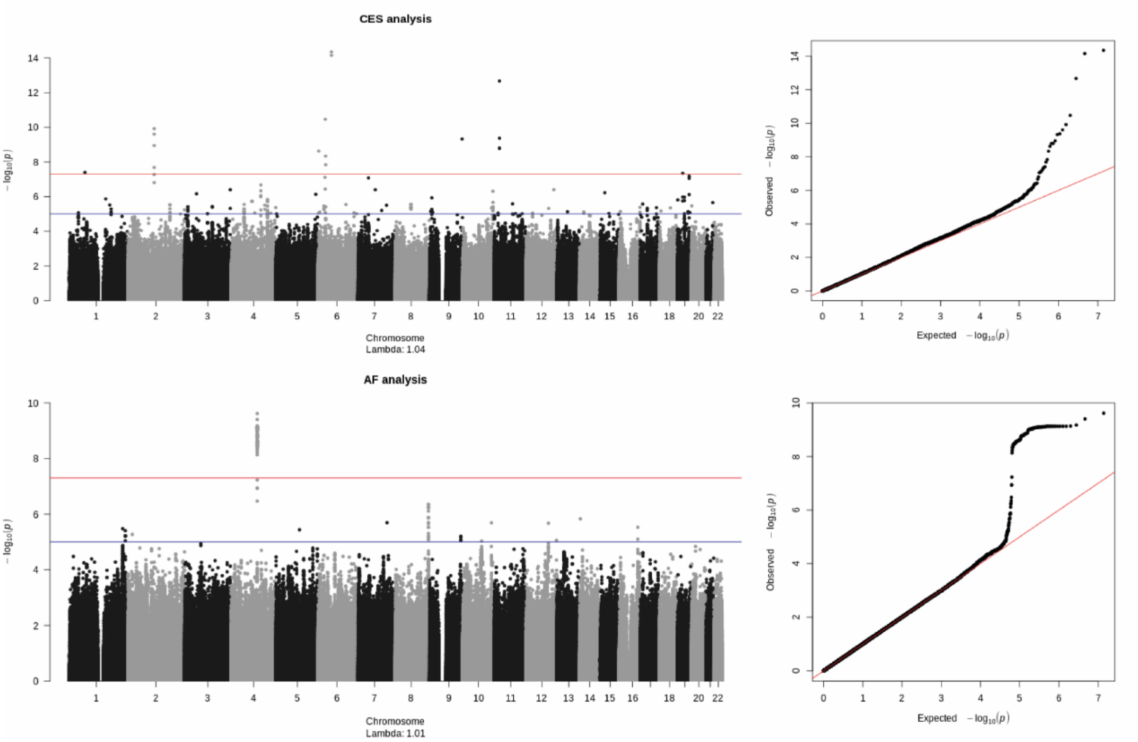

#### 3. ONLINE TABLES

Online tables are annexed in an online data Excel file.

**Online Table I. Detailed number of individuals included form each participant Hospital on the independent cohort.**

**Online Table II**. Detailed number of participants (IS and controls) included from each project. IS: ischemic strokes patients.

**Online Table III. Clinical findings and univariate analysis of the additional cohort.**

**Online Table IV. Genes prioritization of the 44 loci significantly associated with CES in MTAG-CES.**

**Online Table V. Missense variants in high LD (r2 > 0.8) with the 44 locus index variants**. LD information and functional annotation was extracted from HaploReg.

**Online Table VI. CpG-sites modified by the index variants in MTAG-CES.**

**Online Table VII. Significant GTEx eQTLs of the index variants from MTAG-CES.**

**Online Table VIII. Significant eQTLGen eQTLs of the index variants from MTAG-CES**

**Online Table IX. Results of CES analysis on the European independent cohort**. AA: Assesed allele, OA: Other allele, CHR: chromosome, BP: base pairs, SE: Standard Error, B: beta, Z: z-score; P: p-value. Genomic location is in Hg19

**Online Table X. Gene Ontology of biological systems results for the gene sets of prioritized genes associated with a risk of cardioembolic stroke.**

**Online Table XI. DrugBank drugs targeting prioritized genes associated with CES.**

**Online Table XII. Results of MTAG for previous and novel loci associated with AF.** SE: Standard Error, B: beta, Z: z-score; P: p-value.

**Online Table XIII. Gene Ontology of biological systems results for the gene sets of prioritized genes associated with atrial fibrillation risk exclusive.**

**Online Table XIV. Gene Ontology of biological systems results for the gene sets of prioritized genes associated with atrial fibrillation risk that do not overlap with those associated with CES.**

**Online Table I. Detailed number of individuals included form each participant Hospital on the independent cohort.**

| Hospitals (City) | Total | IS | Controls |
|---|---|---|---|
| Hospital Universitari Vall d'Hebron (Barcelona) | 1683 | 1082 | 601 |
| Hospital Clínico Universitario de Santiago (Galicia) | 585 | 585 | |
| Hospital del Mar (Barcelona) | 545 | 539 | 6 |
| Hospital Universitari Germans Tries i Pujol (Badalona) | 353 | 340 | 13 |
| Hospital Universitari Son Espases (Palma de Mallorca) | 253 | 253 | |
| Hospital Universitari Mútua de Terrassa (Terrassa) | 239 | 204 | 35 |
| Complejo Hospitalario Universitario de Albacete (Albacete) | 191 | 191 | |
| Hospital Clinic de Barcelona (Barcelona) | 108 | 108 | |
| Hospital Universitario Virgen del Rocio Y Virgen Macarena (Sevilla) | 71 | 65 | 6 |
| Hospital de la Santa Creu i Sant Pau (Barcelona) | 62 | 60 | 2 |
| Hospital Universitario Río Hortega (Valladolid) | 33 | 26 | 7 |
| Hospital Universitario de Basurto (Bilbao) | 32 | 22 | 10 |
| Hospital Universitario Doctor Josep Trueta (Girona) | 11 | 1 | 10 |
| Hospital Arnau de Vilanova (Lleida) | 3 | 3 | |
| **GCAT Project** | 4936 | | 4936 |
| **TOTAL** | 9105 | 3479 | 5626 |

**Online Table II. Detailed number of participants (IS and controls) included from each project. IS: ischemic strokes patients.**

| Name | Array | Total | IS | Controls |
|---|---|---|---|---|
| GCAT | Illumina® Infimum Multi-Ethnic Global consortium array | 4936 | | 4936 |
| GENISIS | Illumina® Human Core Exome chip | 2187 | 2187 | |
| GODS | Illumina® Human Core Exome chip | 631 | 631 | |
| CONIC | Illumina® Human Core Exome chip | 368 | 179 | 189 |
| ISSYS | Illumina® Human Core Exome chip | 312 | | 312 |
| | Illumina® Human Core Exome chip | 112 | 112 | |
| SEDMAN | Axiom Spain Biobank Array | 202 | 202 | |
| GRECOS | Illumina® Human Core Exome chip | 189 | | 189 |
| GENOTPA | Illumina® Omni 1.5M | 168 | 168 | |
| TOTAL | | 9105 | 3479 | 5626 |

**Online Table III. Clinical findings and univariate analysis of the additional cohort**

| | GENERACION | | | |
|---|---|---|---|---|
| | **Total** (n=9105) | **IS** (n=3479) | **Controls** (n=5626) | **P** |
| **Sex (% male)** | 4434 (49%) | 1915 (55%) | 2519 (45%) | 2.17E-21 |
| **DM (%)** | 1024 (25%) | 914 (26%) | 110 (16%) | 8.87E-09 |
| **HTN (%)** | 3589 (39%) | 2327 (67%) | 1262 (22%) | < 2.2e-16 |
| **DL (%)** | 999 (43%) | 740 (41%) | 259 (49%) | 6.86E-04 |
| **AF (%)** | 1110 (12%) | 1084 (31%) | 26 (0.5%) | < 2.2e-16 |
| **TOAST(%)** | | | | |
| **CES** | | 1515 (44%) | | |
| **LAA** | | 535 (15%) | | |
| **SVO** | | 223 (6%) | | |
| **UND** | | 967 (28%) | | |
| **Age (Years. IQR)** | 58 (49-72) | 75 (66-82) | 52 (46-59) | < 2.2e-16 |

### Online Table IV. Genes prioritization of the 44 loci significantly associated with CES in MTAG-CES.

| Index variant | Locus | Gene | Coding | Nearest | eQTL (eQTLGen or Gtex) | Capture HiC Plotter Choy et al. | mQTL | Combined score | Prioritized genes |
|---|---|---|---|---|---|---|---|---|---|
| rs3746471-A | 20q11.23 | KIAA1755 | 1 | 1 | 1 | | 1 | 4 | KIAA1755 |
| rs55985730-T | 7q32.1 | OPN1SW | | 1 | | 1 | | 2 | CALU, FLNC |
| | | CALU | | | 1 | 1 | 1 | 3 | |
| | | RP11-274B21.4 | | | 1 | | | 1 | |
| | | RP11-274B21.3 | | | 1 | | | 1 | |
| | | RP11-274B21.2 | | | 1 | | | 1 | |
| | | METTL2B | | | 1 | | | 1 | |
| | | RP11-274B21.1 | | | 1 | | | 1 | |
| | | IRF5 | | | 1 | | | 1 | |
| | | RBM28 | | | 1 | | | 1 | |
| | | PRRT4 | | | 1 | | | 1 | |
| | | FLNC | | | 1 | 1 | 1 | 3 | |
| | | FAM71f1 | | | | 1 | | 1 | |
| | | CCDC136 | | | | 1 | | 1 | |
| rs11099098-T | 4q21.21 | FGF5 | | 1 | 1 | | | 2 | FGF5, PRDM8 |
| | | BMP3 | | | 1 | | | 1 | |
| | | PRDM8 | | | 1 | | 1 | 2 | |
| rs1898096-A | 10q22.3 | LRMDA | | 1 | | | 1 | 2 | LRMDA |
| C10orf11 = LRMDA | | PAFAH1B2 | | | 1 | | | 1 | |
| | | ZNF503 | | | 1 | | | 1 | |
| rs12211255-A | 6q14.1 | FILIP1 | | 1 | 1 | 1 | | 3 | FILIP1 |
| | | SENP6 | | | 1 | | | 1 | |
| | | MYO6 | | | 1 | | | 1 | |
| | | TMEM30A | | | 1 | 1 | | 2 | |
| | | COX7A2 | | | | 1 | 1 | 2 | |
| rs11782313-T | 8q24.3 | PTK2 | | 1 | 1 | | 1 | 3 | PTK2 |
| | | CHRAC1 | | | 1 | | | 1 | |
| | | RP11-809O17.1 | | | 1 | | | 1 | |
| | | AGO2 | | | 1 | | 1 | 2 | |
| | | ERICD | | | 1 | | | 1 | |
| rs11057583-A | 12q24.31 | NCOR2 | | 1 | 1 | | 1 | 3 | NCOR2 |
| | | FAM101A | | | | | 1 | 1 | |
| rs2885697-T | 1q34.2 | SCMH1 | | 1 | 1 | | 1 | 3 | SCMH1 |
| | | S100B | | | 1 | | | 1 | |
| | | SPATC1L | | | 1 | | | 1 | |
| | | DIP2A | | | 1 | | | 1 | |
| | | PRMT2 | | | 1 | | | 1 | |
| | | CTPS1 | | | 1 | | 1 | 2 | |
| | | YBEY | | | 1 | | | 1 | |
| | | CITED4 | | | 1 | | | 1 | |
| | | RIMS3 | | | 1 | | | 1 | |
| | | FOXO6 | | | 1 | | | 1 | |
| | | PCNT | | | 1 | | | 1 | |
| | | SLFNL1 | | | 1 | | 1 | 2 | |
| | | RP11-399E6.1 | | | 1 | | | 1 | |
| | | KCNQ4 | | | | | 1 | 1 | |
| rs13010313-T | 2q22.3 | TEX41 | | 1 | | | | 1 | TEX41 |
| rs116600817-A | 17p13.1 | TNFSF12 | | 1 | 1 | | 1 | 3 | TNFSF12 |
| | | SAT2 | | | 1 | | | 1 | |
| | | CD68 | | | 1 | | 1 | 2 | |
| | | AC113189.5 | | | 1 | | | 1 | |
| | | TNFS13 | | | 1 | | | 1 | |
| | | ACAP1 | | | 1 | | | 1 | |
| | | MPDU1 | | | 1 | | 1 | 2 | |
| | | PLSCR3 | | | 1 | | | 1 | |
| | | KCTD11 | | | 1 | | 1 | 2 | |
| | | RP11-186B7.4 | | | 1 | | | 1 | |
| | | CHD3 | | | 1 | | | 1 | |
| | | ZBTB4 | | | 1 | | 1 | 2 | |
| | | CHRNB1 | | | 1 | | 1 | 2 | |
| | | SENP3 | | | 1 | | | 1 | |
| | | CLDN7 | | | 1 | 1 | | 2 | |
| | | POLR2A | | | | 1 | | 1 | |
| | | FGF11 | | | | 1 | 1 | 2 | |
| | | TMEM102 | | | | 1 | 1 | 2 | |
| | | RP11-104H15.7 | | | | 1 | | 1 | |
| | | RP11-104H15.9 | | | | 1 | | 1 | |
| | | RP11-104H15.10 | | | | 1 | | 1 | |
| | | RP11-104H15.8 | | | | 1 | | 1 | |
| | | NLGN2 | | | | 1 | 1 | 2 | |
| | | TMEM256 | | | | 1 | | 1 | |
| | | PLSCR3 | | | | 1 | | 1 | |
| | | TNK1 | | | | 1 | | 1 | |
| | | ACAP1 | | | | 1 | 1 | 2 | |
| | | KCTD11 | | | | 1 | 1 | 2 | |
| | | NEURL4 | | | | 1 | 1 | 2 | |
| | | RP11-542C16.2 | | | | 1 | | 1 | |

| | | | | | | | | | |
|---|---|---|---|---|---|---|---|---|---|
| | | GPS2 | | | | 1 | | 1 | |
| | | EIF5A | | | | 1 | | 1 | |
| | | RP1-4G17.5 | | | | 1 | | 1 | |
| | | ELP5 | | | | 1 | | 1 | |
| | | DVL2 | | | | 1 | | 1 | |
| | | PHF23 | | | | 1 | | 1 | |
| | | GABARAP | | | | 1 | | 1 | |
| | | CTDNEP1 | | | | 1 | | 1 | |
| | | ATP1B2 | | | | | 1 | 1 | |
| | | DNAH2 | | | | | 1 | 1 | |
| | | SHBG | | | | | 1 | 1 | |
| | | SOX15 | | | | | 1 | 1 | |
| rs10272350-A | 7q11.23 | PHTF2 | | 1 | 1 | | | 2 | PHTF2 |
| | | TMEM60 | | | 1 | | | 1 | |
| | | PTPN12 | | | 1 | | | 1 | |
| | | RSBN1L | | | 1 | | | 1 | |
| | | CCDC146 | | | 1 | | | 1 | |
| | | PION | | | 1 | | | 1 | |
| rs635634-T | 9q34.2 | ABO | | 1 | 1 | | 1 | 3 | ABO |
| | | SURF1 | | | 1 | | 1 | 2 | |
| | | ADAMTSL2 | | | | | 1 | 1 | |
| | | SURF6 | | | | | 1 | 1 | |
| | | GBGT1 | | | 1 | | 1 | 2 | |
| | | SURF6 | | | | | 1 | 1 | |
| | | C9orf7 | | | | | 1 | 1 | |
| | | SLC2A6 | | | | | 1 | 1 | |
| | | ADAMTS13 | | | | | 1 | 1 | |
| | | SURF4 | | | | | 1 | 1 | |
| | | TMEM8C | | | | | 1 | 1 | |
| | | RALGDS | | | | | 1 | 1 | |
| | | CACFD1 | | | 1 | | | 1 | |
| | | MED22 | | | 1 | | | 1 | |
| rs11125871-T | 2p15 | USP34 | | 1 | | 1 | 1 | 3 | USP34 |
| | | KIAA1841 | | | 1 | | | 1 | |
| | | AHSA2 | | | 1 | | 1 | 2 | |
| | | ACO16747.3 | | | 1 | | | 1 | |
| | | PUS10 | | | 1 | | | 1 | |
| | | REL | | | 1 | | | 1 | |
| | | PEX13 | | | 1 | | | 1 | |
| | | RP11-493E12.1 | | | 1 | | | 1 | |
| | | C2orf74 | | | 1 | | 1 | 2 | |
| | | XPO1 | | | 1 | | 1 | 2 | |
| | | RP11-355B11.2 | | | 1 | | | 1 | |
| rs6598541-A | 15q26.3 | IGF1R | | 1 | 1 | | 1 | 3 | IGF1R |
| rs2269001-A | 7q36.1 | KCNH2 | | 1 | 1 | | 1 | 3 | KCNH2 |
| | | ABP1 | | | 1 | | 1 | 2 | |
| | | AGAP3 | | | 1 | | 1 | 2 | |
| | | NOS3 | | | | | 1 | 1 | |
| | | TMEM176A | | | | | 1 | 1 | |
| | | TMEM176B | | | | | 1 | 1 | |
| rs12260801-T | 10q25.2 | RBM20 | | 1 | 1 | | | 2 | RBM20 |
| | | BBIP1 | | | 1 | | | 1 | |
| rs79187193-A | 1q21.2 | GJA5 | | 1 | | 1 | 1 | 3 | GJA5 |
| | | NBPF11 | | | 1 | | | 1 | |
| | | CH17-172B3.1 | | | 1 | | | 1 | |
| | | GJA8 | | | | | 1 | 1 | |
| | | PGAP2 | | | | | 1 | 1 | |
| rs55754224-T | 4q26 | CAMK2D | | 1 | 1 | | | 2 | CAMK2D |
| rs76774446-A | 17q21.32 | GOSR2 | | 1 | 1 | | 1 | 3 | GOSR2 |
| | | LRRC37A | | | 1 | | | 1 | |
| | | NSFP1 | | | 1 | | | 1 | |
| | | ARL17B | | | 1 | | 1 | 2 | |
| | | FAM215B | | | 1 | | | 1 | |
| | | NSF | | | 1 | | | 1 | |
| | | TBKP1 | | | 1 | | | 1 | |
| | | WNT3 | | | 1 | | 1 | 2 | |
| | | RPRML | | | 1 | | 1 | 2 | |
| | | LINC01974 | | | 1 | | | 1 | |
| | | OSBPL7 | | | 1 | | | 1 | |
| | | ARL17A | | | | | 1 | 1 | |
| | | WNT9B | | | | | 1 | 1 | |
| rs140185678-A | 16p13.3 | RPL3L | 1 | 1 | | | | 2 | RPL3L |
| | | MEIOB | | | | 1 | | 1 | |
| rs41292535-A | 1p32.2 | EPS15 | | 1 | 1 | | | 2 | EPS15 |
| | | TXNDC12 | | | 1 | | | 1 | |
| | | RNF11 | | | 1 | | | 1 | |
| | | LINC01562 | | | 1 | | | 1 | |

| | | | | | | | | | |
|---|---|---|---|---|---|---|---|---|---|
| | | RP11-296A18.5 | | | 1 | | | 1 | |
| rs337705-T | 5q22.3 | KCNN2 | | 1 | | | | 1 | KCNN2 |
| rs6747542-T | 2p13.3 | GMCL1 | | 1 | 1 | | | 2 | GMCL1, ANXA4, SNRNP27, ASPRV1 |
| | | ANXA4 | | | 1 | | 1 | 2 | |
| | | EMR2 | | | 1 | | | 1 | |
| | | SNRNP27 | | | 1 | | 1 | 2 | |
| | | ZNF333 | | | 1 | | | 1 | |
| | | ASPRV1 | | | 1 | | 1 | 2 | |
| | | PCBP1-AS1 | | | 1 | | | 1 | |
| | | MXD1 | | | 1 | | | 1 | |
| | | C2orf42 | | | 1 | | | 1 | |
| | | FAM136A | | | 1 | | | 1 | |
| | | QKI | | | | | 1 | 1 | |
| rs3176326-A | 6p21.2 | CDKN1A | | 1 | 1 | | 1 | 3 | CDKN1A |
| | | RAB44 | | | 1 | | | 1 | |
| | | KCTD20 | | | 1 | | | 1 | |
| | | ETV7 | | | 1 | | | 1 | |
| | | CPNE5 | | | | 1 | | 1 | |
| rs12908004-A | 15q25.1 | ARNT2 | | 1 | 1 | | | 2 | ARNT2 |
| | | RP11-210M15.2 | | | 1 | | | 1 | |
| | | LINC01314 | | | 1 | | | 1 | |
| | | FAH | | | | | 1 | 1 | |
| rs56181519-T | 2q31.1 | WIPF1 | | 1 | 1 | | 1 | 3 | WIPF1 |
| | | GPR155 | | | 1 | | 1 | 2 | |
| | | WIPF1 | | | 1 | | | 1 | |
| | | RP11-483E17.1 | | | 1 | | | 1 | |
| rs2834618-T | 21q22.12 | RUNX1 | | 1 | | 1 | | 2 | RUNX1 |
| | | SETD4 | | | | 1 | | 1 | |
| | | KCNE1 | | | | | 1 | 1 | |
| rs13191450-A | 6q22.31 | HSF2 | | 1 | 1 | | | 2 | HSF2 |
| | | RP3-425C14.4 | | | 1 | | | 1 | |
| rs74399915-T | 11q24.3 | KCNJ5 | | 1 | 1 | | 1 | 3 | KCNJ5 |
| | | C11orf45 | | | 1 | | 1 | 2 | |
| rs10753933-T | 1q32.1 | PPFIA4 | | 1 | 1 | | 1 | 3 | PPFIA4 |
| | | TMEM183A | | | 1 | | | 1 | |
| | | ADORA1 | | | 1 | | 1 | 2 | |
| | | KLHL12 | | | 1 | | | 1 | |
| | | CHI3L1 | | | 1 | | | 1 | |
| | | MYOG | | | 1 | | 1 | 2 | |
| | | CYB5R1 | | | | | 1 | 1 | |
| | | ADIPOR1 | | | | | 1 | 1 | |
| rs2857265-A | 2q31.2 | TTN | 1 | 1 | 1 | | 1 | 4 | TTN |
| | | TTN-AS1 | | | 1 | | | 1 | |
| | | SESTD1 | | | 1 | | | 1 | |
| | | FKBP7 | | | 1 | | 1 | 2 | |
| | | PLEKHA3 | | | 1 | | 1 | 2 | |
| | | RP11-171I2.1 | | | 1 | | | 1 | |
| | | LIN28 | | | | | 1 | 1 | |
| | | MIR548N | | | | | 1 | 1 | |
| rs6891790-T | 5q35.1 | NKX2-5 | | 1 | 1 | | 1 | 3 | NKX2-5 |
| | | BNIP1 | | | 1 | | 1 | 2 | |
| | | CREBRF | | | 1 | | 1 | 2 | |
| rs2738413-A | 14q23.2 | SYNE2 | | 1 | 1 | | 1 | 3 | SYNE2 |
| | | ESR2 | | | 1 | | 1 | 2 | |
| | | ZBTB25 | | | 1 | | | 1 | |
| | | MTHFD1 | | | 1 | | | 1 | |
| | | PLEKHG3 | | | 1 | | | 1 | |
| | | ZBTB1 | | | 1 | | | 1 | |
| rs7172038-T | 15q24.1 | HCN4 | | 1 | | | 1 | 2 | HCN4 |
| | | NEO1 | | | 1 | | | 1 | |
| rs78249997-T | 10q22.2 | SYNPO2L | 1 | 1 | 1 | | 1 | 4 | SYNPO2L |
| | | NDST2 | | | 1 | | | 1 | |
| | | ECD | | | 1 | | | 1 | |
| | | NUDT13 | | | 1 | | 1 | 2 | |
| | | MRPS16 | | | 1 | | | 1 | |
| | | MRPS16 | | | 1 | | | 1 | |
| | | FAM149B1 | | | 1 | | | 1 | |
| | | RP11-152N13.12 | | | 1 | | | 1 | |
| | | P4HA1 | | | 1 | | | 1 | |
| | | RP11-574K11.5 | | | 1 | | | 1 | |
| | | ADK | | | 1 | | | 1 | |
| | | DNAJC9 | | | 1 | | | 1 | |
| | | RP11-464F9.9 | | | 1 | | | 1 | |
| | | RP11-464F9.1 | | | 1 | | | 1 | |
| | | CAMK2G | | | 1 | | 1 | 2 | |
| | | MYOZ1 | | | 1 | | | 1 | |
| | | FUT11 | | | 1 | 1 | 1 | 3 | |

| | | | | | | | | |
|---|---|---|---|---|---|---|---|---|
| | | RP11-574K11.24 | | 1 | | | 1 | |
| | | BMS1P4 | | 1 | | | 1 | |
| | | ZSWIM8 | | 1 | | | 1 | |
| | | AGAP5 | | 1 | | | 1 | |
| | | USP54 | | 1 | 1 | | 2 | |
| | | DUSP8P5 | | 1 | | | 1 | |
| | | FAM25A | | | | 1 | 1 | |
| | | CHCHD1 | | | | 1 | 1 | |
| | | ASAH2B | | | | 1 | 1 | |
| rs4385527-A | 9q22.32 | AOPEP | 1 | 1 | | 1 | 3 | AOPEP |
| | | HIATL | | 1 | | 1 | 2 | |
| | | ZNF169 | | 1 | | | 1 | |
| | | FANCC | | 1 | | 1 | 2 | |
| | | FBP1 | | | | 1 | 1 | |
| rs17171711-T | 5q31.2 | FAM13B | 1 | 1 | | | 2 | FAM13B, KLHL3, KDM3B, GFRA3, KIF20A |
| | | KLHL3 | | 1 | | 1 | 2 | |
| | | KDM3B | | 1 | | 1 | 2 | |
| | | CTNNA1 | | 1 | | | 1 | |
| | | RP11-325L7.2 | | 1 | | | 1 | |
| | | GFRA3 | | 1 | | 1 | 2 | |
| | | RP11-325L7.1 | | 1 | | | 1 | |
| | | REEP2 | | 1 | | | 1 | |
| | | KIF20A | | 1 | | 1 | 2 | |
| | | CDC23 | | 1 | | | 1 | |
| | | AC034243.1 | | 1 | | | 1 | |
| | | CDC25C | | | | 1 | 1 | |
| rs883079-T | 12q24.21 | TBX5 | 1 | 1 | | 1 | 3 | TBX5 |
| rs680084-A | 1q24.2 | PRRX1 | 1 | 1 | | 1 | 3 | PRRX1 |
| | | PYGB | | 1 | | | 1 | |
| | | LRRFIP2 | | 1 | | | 1 | |
| | | ITGA9 | | 1 | | | 1 | |
| | | NANP | | 1 | | | 1 | |
| | | ABHD12 | | 1 | | | 1 | |
| | | GOLGA4 | | 1 | | | 1 | |
| | | NINL | | 1 | | | 1 | |
| | | MLH1 | | 1 | | | 1 | |
| | | GORAB | | 1 | | | 1 | |
| | | CST7 | | 1 | | | 1 | |
| | | ACSS1 | | 1 | | | 1 | |
| | | C3orf35 | | 1 | | | 1 | |
| | | SELL | | 1 | | | 1 | |
| | | KAT2B | | 1 | | | 1 | |
| | | ENTPD6 | | 1 | | | 1 | |
| | | TAMM41 | | 1 | | | 1 | |
| | | EFHB | | 1 | | | 1 | |
| | | SUCLG2 | | 1 | | | 1 | |
| | | RP1-79C4.4 | | 1 | | | 1 | |
| rs3807989-A | 7q31.2 | CAV1 | 1 | 1 | | 1 | 3 | CAV1 |
| | | CAV2 | | 1 | | 1 | 2 | |
| | | TES | | 1 | | | 1 | |
| | | AC073130.3 | | 1 | | | 1 | |
| | | AC002066.1 | | 1 | | | 1 | |
| | | RP11-667F14.1 | | 1 | | | 1 | |
| rs11598047-A | 10q24.33 | NEURL1 | 1 | 1 | | 1 | 3 | NEURL1 |
| | | USMG5 | | 1 | | | 1 | |
| | | CALHM1 | | | | 1 | 1 | |
| | | SH3PXD2A | | | | 1 | 1 | |
| rs11264280-T | 1q21.3 | KCNN3 | 1 | 1 | | 1 | 3 | KCNN3 |
| | | ADAM15 | | 1 | | | 1 | |
| | | PBXIP1 | | 1 | | 1 | 2 | |
| | | PMVK | | 1 | | 1 | 2 | |
| | | THBS3 | | 1 | | | 1 | |
| | | MUC1 | | 1 | | | 1 | |
| | | RP11-307C12.11 | | 1 | | | 1 | |
| | | SYT11 | | 1 | | | 1 | |
| | | DAP3 | | 1 | | | 1 | |
| | | ZBTB7B | | 1 | | 1 | 2 | |
| | | NCKAP5L | | 1 | | | 1 | |
| | | YY1AP1 | | 1 | | | 1 | |
| | | DCST2 | | 1 | | 1 | 2 | |
| | | DCST1-AS1 | | 1 | | | 1 | |
| | | CRTC2 | | 1 | | | 1 | |
| | | GBA | | 1 | | | 1 | |
| | | MSTO2P | | 1 | | | 1 | |
| | | ADAR | | | 1 | | 1 | |
| | | UBE2Q1 | | | 1 | | 1 | |
| | | CHRNB2 | | | 1 | | 1 | |

| | | | | | | | |
|---|---|---|---|---|---|---|---|
| | | DCST1 | | | 1 | 1 | |
| | | CKS1B | | | 1 | 1 | |
| | | SHC1 | | | 1 | 1 | |
| rs2106261-T | 16q22.3 | ZFHX3 | 1 | | 1 | 2 | ZFHX3 |
| | | HP | | 1 | | 1 | |
| rs17042098-A | 4q25 | PITX2 | 1 | | | 1 | PITX2 |

**Online Table V. Missense variants in high LD (r2 > 0.8) with the 44 locus index variants.** LD information and functional annotation was extracted from HaploReg.

| Locus | Index Variant | Missense Variant | r2 | p-value MTAG-CES | Gene Symbol | Aminoacid change | Transccript ID | SIFT score | SIFT prediction |
|---|---|---|---|---|---|---|---|---|---|
| 20q11.23 | rs3746471 | rs3746471 | 1 | 3.50E-08 | KIAA1755 | R1045W | ENST00000279024 | 0.007 | DELETEREOUS |
| 16p13.3 | rs140185678 | rs140185678 | 1 | 1.13E-11 | RPL3L | A75V | ENST00000268661 | 0.055 | TOLERATED |
| 16p13.3 | rs140185678 | rs140185678 | 1 | 1.13E-11 | RPL3L | A75V | ENST00000565426 | 0.002 | DELETEREOUS |
| 2q31.2 | rs2857265 | rs3829747 | 1 | 1.31E-16 | TTN | R25721H | ENST00000342175 | NA | NA |
| 2q31.2 | rs2857265 | rs3829747 | 1 | 1.31E-16 | TTN | R32026H | ENST00000342992 | NA | NA |
| 2q31.2 | rs2857265 | rs3829747 | 1 | 1.31E-16 | TTN | R25654H | ENST00000359218 | NA | NA |
| 2q31.2 | rs2857265 | rs3829747 | 1 | 1.31E-16 | TTN | R25529H | ENST00000460472 | NA | NA |
| 2q31.2 | rs2857265 | rs3829747 | 1 | 1.31E-16 | TTN | R34594H | ENST00000589042 | NA | NA |
| 2q31.2 | rs2857265 | rs3829747 | 1 | 1.31E-16 | TTN | R32953H | ENST00000591111 | NA | NA |
| 2q31.2 | rs2857265 | rs3731749 | 1 | 3.57E-16 | TTN | R23665H | ENST00000342175 | NA | NA |
| 2q31.2 | rs2857265 | rs3731749 | 1 | 3.57E-16 | TTN | R29970H | ENST00000342992 | NA | NA |
| 2q31.2 | rs2857265 | rs3731749 | 1 | 3.57E-16 | TTN | R23598H | ENST00000359218 | NA | NA |
| 2q31.2 | rs2857265 | rs3731749 | 1 | 3.57E-16 | TTN | R23473H | ENST00000460472 | NA | NA |
| 2q31.2 | rs2857265 | rs3731749 | 1 | 3.57E-16 | TTN | R32538H | ENST00000589042 | NA | NA |
| 2q31.2 | rs2857265 | rs3731749 | 1 | 3.57E-16 | TTN | R30897H | ENST00000591111 | NA | NA |
| 2q31.2 | rs2857265 | rs744426 | 0.96 | 2.74E-16 | TTN | R16074C | ENST00000342175 | NA | NA |
| 2q31.2 | rs2857265 | rs744426 | 0.96 | 2.74E-16 | TTN | R22379C | ENST00000342992 | NA | NA |
| 2q31.2 | rs2857265 | rs744426 | 0.96 | 2.74E-16 | TTN | R16007C | ENST00000359218 | NA | NA |
| 2q31.2 | rs2857265 | rs744426 | 0.96 | 2.74E-16 | TTN | R15882C | ENST00000460472 | NA | NA |
| 2q31.2 | rs2857265 | rs744426 | 0.96 | 2.74E-16 | TTN | R24947C | ENST00000589042 | NA | NA |
| 2q31.2 | rs2857265 | rs744426 | 0.96 | 2.74E-16 | TTN | R23306C | ENST00000591111 | NA | NA |
| 2q31.2 | rs2857265 | rs16866406 | 0.97 | 8.28E-16 | TTN | P10989L | ENST00000342175 | NA | NA |
| 2q31.2 | rs2857265 | rs16866406 | 0.97 | 8.28E-16 | TTN | P17294L | ENST00000342992 | NA | NA |
| 2q31.2 | rs2857265 | rs16866406 | 0.97 | 8.28E-16 | TTN | P10922L | ENST00000359218 | NA | NA |
| 2q31.2 | rs2857265 | rs16866406 | 0.97 | 8.28E-16 | TTN | P10797L | ENST00000460472 | NA | NA |
| 2q31.2 | rs2857265 | rs16866406 | 0.97 | 8.28E-16 | TTN | P19862L | ENST00000589042 | NA | NA |
| 2q31.2 | rs2857265 | rs16866406 | 0.97 | 8.28E-16 | TTN | P18221L | ENST00000591111 | NA | NA |
| 2q31.2 | rs2857265 | rs2288569 | 0.97 | 1.11E-15 | TTN | R10606H | ENST00000342175 | NA | NA |
| 2q31.2 | rs2857265 | rs2288569 | 0.97 | 1.11E-15 | TTN | R16911H | ENST00000342992 | NA | NA |
| 2q31.2 | rs2857265 | rs2288569 | 0.97 | 1.11E-15 | TTN | R10539H | ENST00000359218 | NA | NA |
| 2q31.2 | rs2857265 | rs2288569 | 0.97 | 1.11E-15 | TTN | R10414H | ENST00000460472 | NA | NA |
| 2q31.2 | rs2857265 | rs2288569 | 0.97 | 1.11E-15 | TTN | R19479H | ENST00000589042 | NA | NA |
| 2q31.2 | rs2857265 | rs2288569 | 0.97 | 1.11E-15 | TTN | R17838H | ENST00000591111 | NA | NA |
| 10q22.2 | rs78249997 | rs34163229 | 0.92 | 3.74E-23 | SYNPO2L | S609Y | ENST00000372873 | 0.012 | DELETERIOUS |
| 10q22.2 | rs78249997 | rs34163229 | 0.92 | 3.74E-23 | SYNPO2L | S833Y | ENST00000394810 | 0.017 | DELETERIOUS |
| 10q22.2 | rs78249997 | rs3812629 | 0.92 | 3.74E-23 | SYNPO2L | P483L | ENST00000372873 | 0.034 | DELETERIOUS |
| 10q22.2 | rs78249997 | rs3812629 | 0.92 | 3.74E-23 | SYNPO2L | P707L | ENST00000394810 | 0.036 | DELETERIOUS |
| 10q22.2 | rs78249997 | rs60632610 | 0.93 | NA | SYNPO2L | G2R | ENST00000394810 | 0 | DELETERIOUS |
| 10q22.2 | rs78249997 | rs60632610 | 0.93 | 3.75E-24 | SYNPO2L | G2S | ENST00000394810 | 0 | DELETERIOUS |

#### Online Table VI. CpG-sites modified by the index variants in MTAG-CES.

| SNP | Locus | EA | OA | Marker | Location | Gene | B | SE | P | Tissue | Ancestry | N | Dataset | PMID |
|---|---|---|---|---|---|---|---|---|---|---|---|---|---|---|
| rs2885697 | 1p34.2 | G | T | cg15962197 | chr1:41704537 | SCMH1 | | | 2.81E-49 | Whole blood | European | 3841 | BIOSQTL_methQTL_EUR_2017 | 27918535 |
| rs2885697 | 1p34.2 | G | T | cg20697417 | chr1:41786773 | | | | 6.60E-11 | Whole blood | European | 3841 | BIOSQTL_methQTL_EUR_2017 | 27918535 |
| rs2885697 | 1p34.2 | G | T | cg02290550 | chr1:41487317 | SLFNL1 | 0.3239 | 0.04029 | 3.40E-15 | Cord blood | European | 771 | Gaunt-T_methQTL-Cord_EUR_2016 | 27036880 |
| rs2885697 | 1p34.2 | G | T | cg13367406 | chr1:41481707 | SLFNL1 | | | 1.30E-11 | Whole blood | European | 3841 | BIOSQTL_methQTL_EUR_2017 | 27918535 |
| rs2885697 | 1p34.2 | G | T | cg16733643 | chr1:41575546 | SCMH1 | | | 2.55E-28 | Whole blood | European | 3841 | BIOSQTL_methQTL_EUR_2017 | 27918535 |
| rs2885697 | 1p34.2 | G | T | cg02290550 | chr1:41487317 | SLFNL1 | 0.2993 | 0.04088 | 6.46E-13 | Whole blood | European | 742 | Gaunt-T_methQTL-FOM_EUR_2016 | 27036880 |
| rs2885697 | 1p34.2 | G | T | cg02290550 | chr1:41487317 | SLFNL1 | 0.6008 | 0.1035 | 6.48E-09 | Monocytes | European | 196 | BLUEPRINT_methQTL_EUR_2016 | 27863251 |
| rs2885697 | 1p34.2 | G | T | cg03387723 | chr1:41708464 | SCMH1 | -0.435 | 0.03987 | 5.36E-26 | Whole blood | European | 837 | Gaunt-T_methQTL-15up_EUR_2016 | 27036880 |
| rs2885697 | 1p34.2 | G | T | cg01376402 | chr1:41350185 | | | | 6.06E-23 | Whole blood | European | 3841 | BIOSQTL_methQTL_EUR_2017 | 27918535 |
| rs2885697 | 1p34.2 | G | T | cg03387723 | chr1:41708464 | SCMH1 | -0.3786 | 0.04135 | 4.07E-19 | Whole blood | European | 834 | Gaunt-T_methQTL-F7_EUR_2016 | 27036880 |
| rs2885697 | 1p34.2 | G | T | cg14498674 | chr1:41707677 | SCMH1 | | | 5.60E-13 | Whole blood | European | 3841 | BIOSQTL_methQTL_EUR_2017 | 27918535 |
| rs2885697 | 1p34.2 | G | T | cg02290550 | chr1:41487317 | SLFNL1 | 0.3119 | 0.03756 | 4.02E-16 | Whole blood | European | 837 | Gaunt-T_methQTL-15up_EUR_2016 | 27036880 |
| rs2885697 | 1p34.2 | G | T | cg02290550 | chr1:41487317 | SLFNL1 | 0.2604 | 0.0418 | 7.70E-10 | Whole blood | European | 837 | Gaunt-T_methQTL-Antenatal_EUR_2016 | 27036880 |
| rs2885697 | 1p34.2 | G | T | cg02331769 | chr1:41487964 | SLFNL1 | | | 2.89E-09 | Whole blood | European | 3841 | BIOSQTL_methQTL_EUR_2017 | 27918535 |
| rs2885697 | 1p34.2 | G | T | cg11152463 | chr1:41350430 | | | | 3.12E-11 | Whole blood | European | 3841 | BIOSQTL_methQTL_EUR_2017 | 27918535 |
| rs2885697 | 1p34.2 | G | T | cg07906262 | chr1:41303546 | KCNQ4 | | | 6.40E-09 | Whole blood | European | 3841 | BIOSQTL_methQTL_EUR_2017 | 27918535 |
| rs2885697 | 1p34.2 | G | T | cg13472192 | chr1:41349990 | | | | 1.88E-13 | Whole blood | European | 3841 | BIOSQTL_methQTL_EUR_2017 | 27918535 |
| rs2885697 | 1p34.2 | G | T | cg03387723 | chr1:41708488 | SCMH1 | | | 2.43E-193 | Whole blood | European | 3841 | BIOSQTL_methQTL_EUR_2017 | 27918535 |
| rs2885697 | 1p34.2 | G | T | cg02290550 | chr1:41487317 | SLFNL1 | 0.3352 | 0.03738 | 1.97E-18 | Whole blood | European | 834 | Gaunt-T_methQTL-F7_EUR_2016 | 27036880 |
| rs2885697 | 1p34.2 | G | T | cg03387723 | chr1:41708464 | SCMH1 | -0.379 | 0.03987 | 2.46E-20 | Cord blood | European | 771 | Gaunt-T_methQTL-Cord_EUR_2016 | 27036880 |
| rs2885697 | 1p34.2 | G | T | cg03387723 | chr1:41708464 | SCMH1 | -0.4942 | 0.04399 | 3.84E-27 | Whole blood | European | 742 | Gaunt-T_methQTL-FOM_EUR_2016 | 27036880 |
| rs2885697 | 1p34.2 | G | T | cg02290550 | chr1:41487341 | SLFNL1 | | | 1.93E-82 | Whole blood | European | 3841 | BIOSQTL_methQTL_EUR_2017 | 27918535 |
| rs2885697 | 1p34.2 | G | T | cg03387723 | chr1:41708464 | SCMH1 | -0.4336 | 0.04243 | 4.63E-23 | Whole blood | European | 837 | Gaunt-T_methQTL-Antenatal_EUR_2016 | 27036880 |
| rs2885697 | 1p34.2 | G | T | cg22005565 | chr1:41487461 | SLFNL1 | | | 1.04E-11 | Whole blood | European | 3841 | BIOSQTL_methQTL_EUR_2017 | 27918535 |
| rs2885697 | 1p34.2 | G | T | cg06248701 | chr1:41350316 | | | | 4.49E-23 | Whole blood | European | 3841 | BIOSQTL_methQTL_EUR_2017 | 27918535 |
| rs2885697 | 1p34.2 | G | T | cg24141382 | chr1:41446702 | CTPS | | | 7.06E-12 | Whole blood | European | 3841 | BIOSQTL_methQTL_EUR_2017 | 27918535 |
| rs2885697 | 1p34.2 | G | T | cg08870646 | chr1:41350207 | | | | 3.18E-09 | Whole blood | European | 3841 | BIOSQTL_methQTL_EUR_2017 | 27918535 |
| rs2885697 | 1p34.2 | G | T | cg20276780 | chr1:41303332 | KCNQ4 | | | 4.48E-09 | Whole blood | European | 3841 | BIOSQTL_methQTL_EUR_2017 | 27918535 |
| rs79187193 | 1q21.2 | A | G | cg26191449 | chr1:147245602 | GJA5 | | | 1.90E-16 | Whole blood | European | 3841 | BIOSQTL_methQTL_EUR_2017 | 27918535 |
| rs79187193 | 1q21.2 | A | G | cg20042248 | chr11:3829760 | PGAP2 | 0.4993 | 0.08885 | 2.69E-08 | Whole blood | European | 837 | Gaunt-T_methQTL-Antenatal_EUR_2016 | 27036880 |
| rs79187193 | 1q21.2 | A | G | cg08307963 | chr1:147245461 | GJA5 | | | 2.99E-11 | Whole blood | European | 3841 | BIOSQTL_methQTL_EUR_2017 | 27918535 |

| | | | | | | | | | | | | | | |
|---|---|---|---|---|---|---|---|---|---|---|---|---|---|---|
| rs79187193 | 1q21.2 | A | G | cg10503840 | chr1:147373596 | GJA8 | | | 1.58E-35 | Whole blood | European | 3841 | BIOSQTL_methQTL_EUR_2017 | 27918535 |
| rs79187193 | 1q21.2 | A | G | cg02103822 | chr1:147246353 | GJA5 | | | 2.21E-34 | Whole blood | European | 3841 | BIOSQTL_methQTL_EUR_2017 | 27918535 |
| rs79187193 | 1q21.2 | A | G | cg17941330 | chr1:147245470 | GJA5 | | | 3.46E-37 | Whole blood | European | 3841 | BIOSQTL_methQTL_EUR_2017 | 27918535 |
| rs79187193 | 1q21.2 | A | G | cg11135724 | chr1:147309371 | | | | 2.94E-20 | Whole blood | European | 3841 | BIOSQTL_methQTL_EUR_2017 | 27918535 |
| rs11264280 | 1q21.3 | C | T | cg09359103 | chr1:154839909 | KCNN3 | -0.591 | 0.0405 | 1.41E-42 | Whole blood | European | 742 | Gaunt-T_methQTL-FOM_EUR_2016 | 27036880 |
| rs11264280 | 1q21.3 | C | T | cg18315680 | chr1:154833093 | KCNN3 | | | 8.54E-122 | Whole blood | European | 3841 | BIOSQTL_methQTL_EUR_2017 | 27918535 |
| rs11264280 | 1q21.3 | C | T | cg06221963 | chr1:154839813 | KCNN3 | -0.6544 | 0.04452 | 2.72E-43 | Cord blood | European | 771 | Gaunt-T_methQTL-Cord_EUR_2016 | 27036880 |
| rs11264280 | 1q21.3 | C | T | cg06221963 | chr1:154839789 | KCNN3 | | | 9.17E-237 | Whole blood | European | 3841 | BIOSQTL_methQTL_EUR_2017 | 27918535 |
| rs11264280 | 1q21.3 | C | T | cg09359103 | chr1:154839909 | KCNN3 | -0.5817 | 0.04571 | 8.31E-34 | Cord blood | European | 771 | Gaunt-T_methQTL-Cord_EUR_2016 | 27036880 |
| rs11264280 | 1q21.3 | C | T | cg06742440 | chr1:154842741 | KCNN3 | | | 6.64E-21 | Whole blood | European | 3841 | BIOSQTL_methQTL_EUR_2017 | 27918535 |
| rs11264280 | 1q21.3 | C | T | cg19233405 | chr1:154988745 | ZBTB7B | | | 3.06E-28 | Whole blood | European | 3841 | BIOSQTL_methQTL_EUR_2017 | 27918535 |
| rs11264280 | 1q21.3 | C | T | cg10503610 | chr1:154833121 | KCNN3 | 0.2637 | 0.04565 | 1.11E-08 | Cord blood | European | 771 | Gaunt-T_methQTL-Cord_EUR_2016 | 27036880 |
| rs11264280 | 1q21.3 | C | T | cg10503610 | chr1:154833145 | KCNN3 | | | 1.38E-91 | Whole blood | European | 3841 | BIOSQTL_methQTL_EUR_2017 | 27918535 |
| rs11264280 | 1q21.3 | C | T | cg12436631 | chr1:155007014 | DCST1;DCST2 | 0.266 | 0.0431 | 1.09E-09 | Whole blood | European | 837 | Gaunt-T_methQTL-Antenatal_EUR_2016 | 27036880 |
| rs11264280 | 1q21.3 | C | T | cg20065217 | chr1:154913283 | | | | 4.75E-33 | Whole blood | European | 3841 | BIOSQTL_methQTL_EUR_2017 | 27918535 |
| rs11264280 | 1q21.3 | C | T | cg24348479 | chr1:155007299 | DCST1;DCST2 | | | 1.25E-08 | Whole blood | European | 3841 | BIOSQTL_methQTL_EUR_2017 | 27918535 |
| rs11264280 | 1q21.3 | C | T | cg14980020 | chr1:154992238 | DCST2 | | | 1.94E-20 | Whole blood | European | 3841 | BIOSQTL_methQTL_EUR_2017 | 27918535 |
| rs11264280 | 1q21.3 | C | T | cg06418219 | chr1:154948305 | SHC1;CKS1B | -0.8298 | 0.03535 | 2.55E-92 | Cord blood | European | 771 | Gaunt-T_methQTL-Cord_EUR_2016 | 27036880 |
| rs11264280 | 1q21.3 | C | T | cg11902458 | chr1:154842722 | KCNN3 | | | 3.99E-15 | Whole blood | European | 3841 | BIOSQTL_methQTL_EUR_2017 | 27918535 |
| rs11264280 | 1q21.3 | C | T | cg06221963 | chr1:154839813 | KCNN3 | -0.6769 | 0.04124 | 4.45E-52 | Whole blood | European | 837 | Gaunt-T_methQTL-Antenatal_EUR_2016 | 27036880 |
| rs11264280 | 1q21.3 | C | T | cg12436631 | chr1:155007014 | DCST1;DCST2 | 0.3156 | 0.04539 | 7.30E-12 | Whole blood | European | 834 | Gaunt-T_methQTL-F7_EUR_2016 | 27036880 |
| rs11264280 | 1q21.3 | C | T | cg16318349 | chr1:154917283 | PBXIP1 | | | 3.97E-76 | Whole blood | European | 3841 | BIOSQTL_methQTL_EUR_2017 | 27918535 |
| rs11264280 | 1q21.3 | C | T | cg09359103 | chr1:154839909 | KCNN3 | -0.6657 | 0.04254 | 1.42E-48 | Whole blood | European | 834 | Gaunt-T_methQTL-F7_EUR_2016 | 27036880 |
| rs11264280 | 1q21.3 | C | T | cg16296829 | chr1:154832511 | KCNN3 | | | 1.77E-26 | Whole blood | European | 3841 | BIOSQTL_methQTL_EUR_2017 | 27918535 |
| rs11264280 | 1q21.3 | C | T | cg15204354 | chr1:155006935 | DCST1;DCST2 | | | 3.36E-36 | Whole blood | European | 3841 | BIOSQTL_methQTL_EUR_2017 | 27918535 |
| rs11264280 | 1q21.3 | C | T | cg27332243 | chr1:154842743 | KCNN3 | | | 1.16E-13 | Whole blood | European | 3841 | BIOSQTL_methQTL_EUR_2017 | 27918535 |
| rs11264280 | 1q21.3 | C | T | cg09359103 | chr1:154839909 | KCNN3 | -0.6664 | 0.04241 | 6.20E-49 | Whole blood | European | 837 | Gaunt-T_methQTL-15up_EUR_2016 | 27036880 |
| rs11264280 | 1q21.3 | C | T | cg18315680 | chr1:154833117 | KCNN3 | 0.287 | 0.0465 | 1.09E-09 | Cord blood | European | 771 | Gaunt-T_methQTL-Cord_EUR_2016 | 27036880 |
| rs11264280 | 1q21.3 | C | T | cg06221963 | chr1:154839813 | KCNN3 | -0.672 | 0.04246 | 1.51E-49 | Whole blood | European | 837 | Gaunt-T_methQTL-15up_EUR_2016 | 27036880 |
| rs11264280 | 1q21.3 | C | T | cg06221963 | chr1:154839813 | KCNN3 | -0.6653 | 0.04307 | 1.64E-47 | Whole blood | European | 834 | Gaunt-T_methQTL-F7_EUR_2016 | 27036880 |
| rs11264280 | 1q21.3 | C | T | cg16680214 | chr1:154839959 | KCNN3 | | | 6.88E-168 | Whole blood | European | 3841 | BIOSQTL_methQTL_EUR_2017 | 27918535 |

| | | | | | | | | | | | | | | |
|---|---|---|---|---|---|---|---|---|---|---|---|---|---|---|
| rs11264280 | 1q21.3 | C | T | cg18315680 | chr1:154833117 | KCNN3 | 0.3065 | 0.04376 | 5.47E-12 | Whole blood | European | 837 | Gaunt-T_methQTL-Antenatal_EUR_2016 | 27036880 |
| rs11264280 | 1q21.3 | C | T | cg18315680 | chr1:154833117 | KCNN3 | 0.2678 | 0.04178 | 2.41E-10 | Whole blood | European | 837 | Gaunt-T_methQTL-15up_EUR_2016 | 27036880 |
| rs11264280 | 1q21.3 | C | T | cg09359103 | chr1:154839909 | KCNN3 | -0.6824 | 0.04057 | 3.16E-54 | Whole blood | European | 837 | Gaunt-T_methQTL-Antenatal_EUR_2016 | 27036880 |
| rs11264280 | 1q21.3 | C | T | cg09359103 | chr1:154839933 | KCNN3 | | | 2.18E-220 | Whole blood | European | 3841 | BIOSQTL_methQTL_EUR_2017 | 27918535 |
| rs11264280 | 1q21.3 | C | T | cg21596858 | chr1:155007230 | DCST1;DCST2 | | | 9.00E-53 | Whole blood | European | 3841 | BIOSQTL_methQTL_EUR_2017 | 27918535 |
| rs11264280 | 1q21.3 | C | T | cg06221963 | chr1:154839813 | KCNN3 | -0.6074 | 0.04046 | 1.17E-44 | Whole blood | European | 742 | Gaunt-T_methQTL-FOM_EUR_2016 | 27036880 |
| rs11264280 | 1q21.3 | C | T | cg01178854 | chr1:154913768 | | | | 1.44E-12 | Whole blood | European | 3841 | BIOSQTL_methQTL_EUR_2017 | 27918535 |
| rs11264280 | 1q21.3 | C | T | cg13396821 | chr1:154812928 | KCNN3 | | | 5.14E-12 | Whole blood | European | 3841 | BIOSQTL_methQTL_EUR_2017 | 27918535 |
| rs11264280 | 1q21.3 | C | T | cg06418219 | chr1:154948305 | SHC1;CKS1B | -0.7646 | 0.03352 | 1.28E-87 | Whole blood | European | 742 | Gaunt-T_methQTL-FOM_EUR_2016 | 27036880 |
| rs11264280 | 1q21.3 | C | T | cg12436631 | chr1:155006990 | DCST1;DCST2 | | | 1.74E-55 | Whole blood | European | 3841 | BIOSQTL_methQTL_EUR_2017 | 27918535 |
| rs11264280 | 1q21.3 | C | T | cg23803022 | chr1:154908805 | PMVK | | | 3.52E-18 | Whole blood | European | 3841 | BIOSQTL_methQTL_EUR_2017 | 27918535 |
| rs11264280 | 1q21.3 | C | T | cg18315680 | chr1:154833117 | KCNN3 | 0.2721 | 0.04354 | 6.59E-10 | Whole blood | European | 834 | Gaunt-T_methQTL-F7_EUR_2016 | 27036880 |
| rs11264280 | 1q21.3 | C | T | cg06418219 | chr1:154948305 | SHC1;CKS1B | -0.8508 | 0.03462 | 1.02E-100 | Whole blood | European | 834 | Gaunt-T_methQTL-F7_EUR_2016 | 27036880 |
| rs11264280 | 1q21.3 | C | T | cg10503610 | chr1:154833121 | KCNN3 | 0.2592 | 0.04034 | 2.32E-10 | Whole blood | European | 837 | Gaunt-T_methQTL-Antenatal_EUR_2016 | 27036880 |
| rs11264280 | 1q21.3 | C | T | cg08472142 | chr1:155007190 | DCST1;DCST2 | | | 9.59E-39 | Whole blood | European | 3841 | BIOSQTL_methQTL_EUR_2017 | 27918535 |
| rs11264280 | 1q21.3 | C | T | cg24250549 | chr1:154909264 | PMVK | | | 4.45E-55 | Whole blood | European | 3841 | BIOSQTL_methQTL_EUR_2017 | 27918535 |
| rs11264280 | 1q21.3 | C | T | cg10503610 | chr1:154833121 | KCNN3 | 0.2244 | 0.04037 | 3.69E-08 | Whole blood | European | 837 | Gaunt-T_methQTL-15up_EUR_2016 | 27036880 |
| rs11264280 | 1q21.3 | C | T | cg06418219 | chr1:154948305 | SHC1;CKS1B | -0.8337 | 0.03376 | 2.10E-99 | Whole blood | European | 837 | Gaunt-T_methQTL-Antenatal_EUR_2016 | 27036880 |
| rs11264280 | 1q21.3 | C | T | cg06418219 | chr1:154948305 | SHC1;CKS1B | -0.8306 | 0.03505 | 2.44E-95 | Whole blood | European | 837 | Gaunt-T_methQTL-15up_EUR_2016 | 27036880 |
| rs11264280 | 1q21.3 | C | T | cg03351412 | chr1:154909275 | PMVK | | | 1.86E-47 | Whole blood | European | 3841 | BIOSQTL_methQTL_EUR_2017 | 27918535 |
| rs680084 | 1q24.2 | A | G | cg09010107 | chr1:170638831 | PRRX1 | | | 1.08E-38 | Whole blood | European | 3841 | BIOSQTL_methQTL_EUR_2017 | 27918535 |
| rs680084 | 1q24.2 | A | G | cg22693806 | chr1:170631823 | PRRX1 | | | 2.54E-28 | Whole blood | European | 3841 | BIOSQTL_methQTL_EUR_2017 | 27918535 |
| rs680084 | 1q24.2 | A | G | cg09010107 | chr1:170638807 | PRRX1 | -0.2503 | 0.04302 | 8.77E-09 | Cord blood | European | 771 | Gaunt-T_methQTL-Cord_EUR_2016 | 27036880 |
| rs680084 | 1q24.2 | A | G | cg22371492 | chr1:170637820 | PRRX1 | | | 1.02E-10 | Whole blood | European | 3841 | BIOSQTL_methQTL_EUR_2017 | 27918535 |
| rs680084 | 1q24.2 | A | G | cg03350186 | chr1:170633484 | PRRX1 | | | 4.45E-13 | Whole blood | European | 3841 | BIOSQTL_methQTL_EUR_2017 | 27918535 |
| rs680084 | 1q24.2 | A | G | cg09010107 | chr1:170638807 | PRRX1 | -0.2767 | 0.04323 | 2.57E-10 | Whole blood | European | 837 | Gaunt-T_methQTL-15up_EUR_2016 | 27036880 |
| rs680084 | 1q24.2 | A | G | cg26669793 | chr1:170637624 | PRRX1 | | | 2.08E-10 | Whole blood | European | 3841 | BIOSQTL_methQTL_EUR_2017 | 27918535 |
| rs10753933 | 1q32.1 | G | T | cg00805880 | chr1:203045044 | PPFIA4 | -0.273 | 0.0457 | 3.58E-09 | Whole blood | European | 837 | Gaunt-T_methQTL-Antenatal_EUR_2016 | 27036880 |
| rs10753933 | 1q32.1 | G | T | cg15493970 | chr1:203046861 | PPFIA4 | | | 4.87E-12 | Whole blood | European | 3841 | BIOSQTL_methQTL_EUR_2017 | 27918535 |
| rs10753933 | 1q32.1 | G | T | cg17862152 | chr1:203044906 | PPFIA4 | | | 3.76E-15 | Whole blood | European | 3841 | BIOSQTL_methQTL_EUR_2017 | 27918535 |
| rs10753933 | 1q32.1 | G | T | cg05652480 | chr1:203016359 | | | | 1.21E-12 | Whole blood | European | 3841 | BIOSQTL_methQTL_EUR_2017 | 27918535 |

| | | | | | | | | | | | | | | |
|---|---|---|---|---|---|---|---|---|---|---|---|---|---|---|
| rs10753933 | 1q32.1 | G | T | cg00805880 | chr1:203045044 | PPFIA4 | -0.3221 | 0.04428 | 8.96E-13 | Whole blood | European | 742 | Gaunt-T_methQTL-FOM_EUR_2016 | 27036880 |
| rs10753933 | 1q32.1 | G | T | cg16731083 | chr1:203086962 | | 0.2579 | 0.04337 | 4.22E-09 | Whole blood | European | 742 | Gaunt-T_methQTL-FOM_EUR_2016 | 27036880 |
| rs10753933 | 1q32.1 | G | T | cg18811731 | chr1:203086869 | | 0.248 | 0.04278 | 9.55E-09 | Whole blood | European | 834 | Gaunt-T_methQTL-F7_EUR_2016 | 27036880 |
| rs10753933 | 1q32.1 | G | T | cg18811731 | chr1:203086869 | | 0.2609 | 0.04191 | 7.58E-10 | Whole blood | European | 837 | Gaunt-T_methQTL-15up_EUR_2016 | 27036880 |
| rs10753933 | 1q32.1 | G | T | cg09396704 | chr1:203045253 | PPFIA4 | | | 4.51E-184 | Whole blood | European | 3841 | BIOSQTL_methQTL_EUR_2017 | 27918535 |
| rs10753933 | 1q32.1 | G | T | cg09396704 | chr1:203045229 | PPFIA4 | -0.5313 | 0.0964 | 3.52E-08 | Monocytes | European | 196 | BLUEPRINT_methQTL_EUR_2016 | 27863251 |
| rs10753933 | 1q32.1 | G | T | cg11656175 | chr1:203040847 | PPFIA4 | | | 8.76E-44 | Whole blood | European | 3841 | BIOSQTL_methQTL_EUR_2017 | 27918535 |
| rs10753933 | 1q32.1 | G | T | cg12096325 | chr1:202996565 | | | | 1.21E-14 | Whole blood | European | 3841 | BIOSQTL_methQTL_EUR_2017 | 27918535 |
| rs10753933 | 1q32.1 | G | T | cg18169013 | chr1:203030744 | PPFIA4 | | | 5.58E-71 | Whole blood | European | 3841 | BIOSQTL_methQTL_EUR_2017 | 27918535 |
| rs10753933 | 1q32.1 | G | T | cg16731083 | chr1:203086962 | | 0.279 | 0.04183 | 4.69E-11 | Whole blood | European | 834 | Gaunt-T_methQTL-F7_EUR_2016 | 27036880 |
| rs10753933 | 1q32.1 | G | T | cg10952234 | chr1:203020593 | PPFIA4 | | | 3.82E-20 | Whole blood | European | 3841 | BIOSQTL_methQTL_EUR_2017 | 27918535 |
| rs10753933 | 1q32.1 | G | T | cg09396704 | chr1:203045229 | PPFIA4 | -0.5623 | 0.096 | 4.77E-09 | Neutrophils | European | 197 | BLUEPRINT_methQTL_EUR_2016 | 27863251 |
| rs10753933 | 1q32.1 | G | T | cg09396704 | chr1:203045229 | PPFIA4 | -0.5807 | 0.03995 | 1.80E-42 | Cord blood | European | 771 | Gaunt-T_methQTL-Cord_EUR_2016 | 27036880 |
| rs10753933 | 1q32.1 | G | T | cg18275051 | chr1:202937138 | CYB5R1 | | | 3.78E-08 | Whole blood | European | 3841 | BIOSQTL_methQTL_EUR_2017 | 27918535 |
| rs10753933 | 1q32.1 | G | T | cg04566392 | chr1:202927049 | ADIPOR1 | | | 3.15E-08 | Whole blood | European | 3841 | BIOSQTL_methQTL_EUR_2017 | 27918535 |
| rs10753933 | 1q32.1 | G | T | cg20142535 | chr1:203097604 | ADORA1 | | | 1.16E-43 | Whole blood | European | 3841 | BIOSQTL_methQTL_EUR_2017 | 27918535 |
| rs10753933 | 1q32.1 | G | T | cg16819803 | chr1:202996151 | | | | 6.55E-11 | Whole blood | European | 3841 | BIOSQTL_methQTL_EUR_2017 | 27918535 |
| rs10753933 | 1q32.1 | G | T | cg23098069 | chr1:203055483 | MYOG | | | 1.35E-24 | Whole blood | European | 3841 | BIOSQTL_methQTL_EUR_2017 | 27918535 |
| rs10753933 | 1q32.1 | G | T | cg18811731 | chr1:203086869 | | 0.2759 | 0.04322 | 3.06E-10 | Whole blood | European | 742 | Gaunt-T_methQTL-FOM_EUR_2016 | 27036880 |
| rs10753933 | 1q32.1 | G | T | cg18811731 | chr1:203086869 | | 0.2942 | 0.04556 | 1.88E-10 | Whole blood | European | 837 | Gaunt-T_methQTL-Antenatal_EUR_2016 | 27036880 |
| rs10753933 | 1q32.1 | G | T | cg16731083 | chr1:203086938 | | | | 9.39E-110 | Whole blood | European | 3841 | BIOSQTL_methQTL_EUR_2017 | 27918535 |
| rs10753933 | 1q32.1 | G | T | cg18811731 | chr1:203086845 | | | | 4.29E-68 | Whole blood | European | 3841 | BIOSQTL_methQTL_EUR_2017 | 27918535 |
| rs10753933 | 1q32.1 | G | T | cg09396704 | chr1:203045229 | PPFIA4 | -0.5613 | 0.04181 | 4.88E-37 | Whole blood | European | 837 | Gaunt-T_methQTL-Antenatal_EUR_2016 | 27036880 |
| rs10753933 | 1q32.1 | G | T | cg00805880 | chr1:203045044 | PPFIA4 | -0.3463 | 0.04343 | 5.10E-15 | Whole blood | European | 834 | Gaunt-T_methQTL-F7_EUR_2016 | 27036880 |
| rs10753933 | 1q32.1 | G | T | cg14640592 | chr1:203044702 | PPFIA4 | | | 8.82E-16 | Whole blood | European | 3841 | BIOSQTL_methQTL_EUR_2017 | 27918535 |
| rs10753933 | 1q32.1 | G | T | cg18169013 | chr1:203030768 | PPFIA4 | -0.2359 | 0.0407 | 1.01E-08 | Whole blood | European | 742 | Gaunt-T_methQTL-FOM_EUR_2016 | 27036880 |
| rs10753933 | 1q32.1 | G | T | cg16731083 | chr1:203086962 | | 0.5295 | 0.0969 | 4.64E-08 | Monocytes | European | 196 | BLUEPRINT_methQTL_EUR_2016 | 27863251 |
| rs10753933 | 1q32.1 | G | T | cg09396704 | chr1:203045229 | PPFIA4 | -0.5266 | 0.04153 | 1.78E-33 | Whole blood | European | 742 | Gaunt-T_methQTL-FOM_EUR_2016 | 27036880 |
| rs10753933 | 1q32.1 | G | T | cg21997465 | chr1:203055686 | MYOG | | | 3.57E-10 | Whole blood | European | 3841 | BIOSQTL_methQTL_EUR_2017 | 27918535 |
| rs10753933 | 1q32.1 | G | T | cg09396704 | chr1:203045229 | PPFIA4 | -0.5849 | 0.04219 | 1.72E-39 | Whole blood | European | 837 | Gaunt-T_methQTL-15up_EUR_2016 | 27036880 |
| rs10753933 | 1q32.1 | G | T | cg22126171 | chr1:203047854 | PPFIA4 | | | 7.61E-29 | Whole blood | European | 3841 | BIOSQTL_methQTL_EUR_2017 | 27918535 |

| | | | | | | | | | | | | | | |
|---|---|---|---|---|---|---|---|---|---|---|---|---|---|---|
| rs10753933 | 1q32.1 | G | T | cg09396704 | chr1:203045229 | PPFIA4 | -0.5817 | 0.04013 | 1.31E-42 | Whole blood | European | 834 | Gaunt-T_methQTL-F7_EUR_2016 | 27036880 |
| rs10753933 | 1q32.1 | G | T | cg00805880 | chr1:203045044 | PPFIA4 | -0.3257 | 0.04401 | 3.31E-13 | Whole blood | European | 837 | Gaunt-T_methQTL-15up_EUR_2016 | 27036880 |
| rs10753933 | 1q32.1 | G | T | cg10901806 | chr1:203055398 | MYOG | | | 2.72E-39 | Whole blood | European | 3841 | BIOSQTL_methQTL_EUR_2017 | 27918535 |
| rs10753933 | 1q32.1 | G | T | cg00805880 | chr1:203045068 | PPFIA4 | | | 2.04E-53 | Whole blood | European | 3841 | BIOSQTL_methQTL_EUR_2017 | 27918535 |
| rs10753933 | 1q32.1 | G | T | cg00805880 | chr1:203045044 | PPFIA4 | -0.3284 | 0.04318 | 8.25E-14 | Cord blood | European | 771 | Gaunt-T_methQTL-Cord_EUR_2016 | 27036880 |
| rs10753933 | 1q32.1 | G | T | cg16731083 | chr1:203086962 | | 0.3408 | 0.04395 | 2.86E-14 | Whole blood | European | 837 | Gaunt-T_methQTL-Antenatal_EUR_2016 | 27036880 |
| rs10753933 | 1q32.1 | G | T | cg03900565 | chr1:203031839 | PPFIA4 | | | 7.72E-52 | Whole blood | European | 3841 | BIOSQTL_methQTL_EUR_2017 | 27918535 |
| rs10753933 | 1q32.1 | G | T | cg14084623 | chr1:203024403 | PPFIA4 | | | 3.01E-26 | Whole blood | European | 3841 | BIOSQTL_methQTL_EUR_2017 | 27918535 |
| rs11125871 | 2p15 | C | T | cg26147678 | chr2:61371904 | C2orf74 | | | 1.11E-12 | Whole blood | European | 3841 | BIOSQTL_methQTL_EUR_2017 | 27918535 |
| rs11125871 | 2p15 | C | T | cg02054108 | chr2:61607478 | USP34 | 0.6241 | 0.1031 | 1.41E-09 | Neutrophils | European | 197 | BLUEPRINT_methQTL_EUR_2016 | 27863251 |
| rs11125871 | 2p15 | C | T | cg10580144 | chr2:61372292 | C2orf74 | | | 1.90E-54 | Whole blood | European | 3841 | BIOSQTL_methQTL_EUR_2017 | 27918535 |
| rs11125871 | 2p15 | C | T | cg15711740 | chr2:61764176 | XPO1 | -0.2106 | 0.03688 | 1.55E-08 | Whole blood | European | 834 | Gaunt-T_methQTL-F7_EUR_2016 | 27036880 |
| rs11125871 | 2p15 | C | T | cg22446208 | chr2:61388665 | C2orf74 | -0.2732 | 0.04107 | 5.24E-11 | Whole blood | European | 834 | Gaunt-T_methQTL-F7_EUR_2016 | 27036880 |
| rs11125871 | 2p15 | C | T | cg01648237 | chr2:61372090 | C2orf74 | | | 3.35E-18 | Whole blood | European | 3841 | BIOSQTL_methQTL_EUR_2017 | 27918535 |
| rs11125871 | 2p15 | C | T | cg06949933 | chr2:61406515 | AHSA2 | | | 8.83E-53 | Whole blood | European | 3841 | BIOSQTL_methQTL_EUR_2017 | 27918535 |
| rs11125871 | 2p15 | C | T | cg02054108 | chr2:61607454 | USP34 | | | 2.40E-71 | Whole blood | European | 3841 | BIOSQTL_methQTL_EUR_2017 | 27918535 |
| rs11125871 | 2p15 | C | T | cg02054108 | chr2:61607478 | USP34 | 0.1906 | 0.02662 | 1.76E-12 | Whole blood | European | 834 | Gaunt-T_methQTL-F7_EUR_2016 | 27036880 |
| rs11125871 | 2p15 | C | T | cg03313140 | chr2:61403635 | AHSA2 | | | 3.30E-25 | Whole blood | European | 3841 | BIOSQTL_methQTL_EUR_2017 | 27918535 |
| rs11125871 | 2p15 | C | T | cg16430332 | chr2:61389197 | C2orf74 | | | 4.48E-09 | Whole blood | European | 3841 | BIOSQTL_methQTL_EUR_2017 | 27918535 |
| rs11125871 | 2p15 | C | T | cg02057644 | chr2:61400528 | | | | 1.20E-15 | Whole blood | European | 3841 | BIOSQTL_methQTL_EUR_2017 | 27918535 |
| rs11125871 | 2p15 | C | T | cg04541421 | chr2:61370740 | C2orf74 | | | 2.29E-14 | Whole blood | European | 3841 | BIOSQTL_methQTL_EUR_2017 | 27918535 |
| rs11125871 | 2p15 | C | T | cg12310231 | chr2:61661196 | USP34 | | | 2.21E-09 | Whole blood | European | 3841 | BIOSQTL_methQTL_EUR_2017 | 27918535 |
| rs11125871 | 2p15 | C | T | cg02054108 | chr2:61607478 | USP34 | 0.2344 | 0.03309 | 3.19E-12 | Whole blood | European | 837 | Gaunt-T_methQTL-Antenatal_EUR_2016 | 27036880 |
| rs6747542 | 2p13.3 | C | T | cg22792910 | chr2:70009327 | ANXA4 | | | 1.10E-09 | Whole blood | European | 3841 | BIOSQTL_methQTL_EUR_2017 | 27918535 |
| rs6747542 | 2p13.3 | C | T | cg02498382 | chr2:70120550 | SNRNP27 | -0.2072 | 0.03592 | 1.12E-08 | Whole blood | European | 834 | Gaunt-T_methQTL-F7_EUR_2016 | 27036880 |
| rs6747542 | 2p13.3 | C | T | cg22117468 | chr2:70176317 | | | | 1.98E-08 | Whole blood | European | 3841 | BIOSQTL_methQTL_EUR_2017 | 27918535 |
| rs6747542 | 2p13.3 | C | T | cg02498382 | chr2:70120550 | SNRNP27 | -0.5462 | 0.0935 | 5.19E-09 | Monocytes | European | 196 | BLUEPRINT_methQTL_EUR_2016 | 27863251 |
| rs6747542 | 2p13.3 | C | T | cg15107349 | chr2:70121099 | SNRNP27 | -0.1957 | 0.03417 | 1.42E-08 | Whole blood | European | 834 | Gaunt-T_methQTL-F7_EUR_2016 | 27036880 |
| rs6747542 | 2p13.3 | C | T | cg02498382 | chr2:70120550 | SNRNP27 | -0.2188 | 0.03719 | 5.95E-09 | Whole blood | European | 837 | Gaunt-T_methQTL-Antenatal_EUR_2016 | 27036880 |
| rs6747542 | 2p13.3 | C | T | cg25332238 | chr2:70122357 | SNRNP27 | | | 1.65E-12 | Whole blood | European | 3841 | BIOSQTL_methQTL_EUR_2017 | 27918535 |
| rs6747542 | 2p13.3 | C | T | cg13874290 | - | ANXA4 | -1.109 | 0.0761 | 4.22E-48 | Neutrophils | European | 197 | BLUEPRINT_methQTL_EUR_2016 | 27863251 |

| | | | | | | | | | | | | | | |
|---|---|---|---|---|---|---|---|---|---|---|---|---|---|---|
| rs6747542 | 2p13.3 | C | T | cg05298696 | chr2:70188691 | ASPRV1 | 0.2632 | 0.03909 | 3.08E-11 | Whole blood | European | 837 | Gaunt-T_methQTL-15up_EUR_2016 | 27036880 |
| rs6747542 | 2p13.3 | C | T | cg18052967 | chr6:163951256 | QKI | 0.1717 | 0.02933 | 6.90E-09 | Whole blood | European | 834 | Gaunt-T_methQTL-F7_EUR_2016 | 27036880 |
| rs6747542 | 2p13.3 | C | T | cg12518775 | chr2:69970318 | ANXA4 | | | 1.75E-28 | Whole blood | European | 3841 | BIOSQTL_methQTL_EUR_2017 | 27918535 |
| rs6747542 | 2p13.3 | C | T | cg01726273 | chr2:70360150 | | -0.2627 | 0.0419 | 6.09E-10 | Whole blood | European | 837 | Gaunt-T_methQTL-Antenatal_EUR_2016 | 27036880 |
| rs6747542 | 2p13.3 | C | T | cg14608581 | chr2:70122216 | SNRNP27 | | | 3.39E-14 | Whole blood | European | 3841 | BIOSQTL_methQTL_EUR_2017 | 27918535 |
| rs6747542 | 2p13.3 | C | T | cg02498382 | chr2:70120550 | SNRNP27 | -0.253 | 0.03414 | 3.10E-13 | Whole blood | European | 837 | Gaunt-T_methQTL-15up_EUR_2016 | 27036880 |
| rs6747542 | 2p13.3 | C | T | cg01726273 | chr2:70360150 | | -0.2317 | 0.03633 | 3.17E-10 | Whole blood | European | 742 | Gaunt-T_methQTL-FOM_EUR_2016 | 27036880 |
| rs6747542 | 2p13.3 | C | T | cg05298696 | chr2:70188667 | ASPRV1 | | | 2.58E-45 | Whole blood | European | 3841 | BIOSQTL_methQTL_EUR_2017 | 27918535 |
| rs6747542 | 2p13.3 | C | T | cg13874290 | - | ANXA4 | -1.123 | 0.0925 | 6.59E-34 | T cells | European | 132 | BLUEPRINT_methQTL_EUR_2016 | 27863251 |
| rs6747542 | 2p13.3 | C | T | cg05298696 | chr2:70188691 | ASPRV1 | 0.2635 | 0.0385 | 1.49E-11 | Whole blood | European | 834 | Gaunt-T_methQTL-F7_EUR_2016 | 27036880 |
| rs6747542 | 2p13.3 | C | T | cg05298696 | chr2:70188691 | ASPRV1 | 0.2475 | 0.03782 | 1.13E-10 | Whole blood | European | 742 | Gaunt-T_methQTL-FOM_EUR_2016 | 27036880 |
| rs6747542 | 2p13.3 | C | T | cg08792812 | chr2:70009225 | ANXA4 | | | 4.67E-19 | Whole blood | European | 3841 | BIOSQTL_methQTL_EUR_2017 | 27918535 |
| rs6747542 | 2p13.3 | C | T | cg02498382 | chr2:70120526 | SNRNP27 | | | 4.21E-149 | Whole blood | European | 3841 | BIOSQTL_methQTL_EUR_2017 | 27918535 |
| rs6747542 | 2p13.3 | C | T | cg02498382 | chr2:70120550 | SNRNP27 | -0.2611 | 0.03724 | 5.31E-12 | Whole blood | European | 742 | Gaunt-T_methQTL-FOM_EUR_2016 | 27036880 |
| rs6747542 | 2p13.3 | C | T | cg04333863 | chr2:70121129 | SNRNP27 | -0.1788 | 0.02917 | 1.40E-09 | Cord blood | European | 771 | Gaunt-T_methQTL-Cord_EUR_2016 | 27036880 |
| rs6747542 | 2p13.3 | C | T | cg01438537 | chr2:70348979 | | | | 8.33E-14 | Whole blood | European | 3841 | BIOSQTL_methQTL_EUR_2017 | 27918535 |
| rs6747542 | 2p13.3 | C | T | cg12430078 | chr2:70121712 | SNRNP27 | | | 3.84E-08 | Whole blood | European | 3841 | BIOSQTL_methQTL_EUR_2017 | 27918535 |
| rs6747542 | 2p13.3 | C | T | cg06635952 | chr2:70025893 | ANXA4 | | | 3.23E-41 | Whole blood | European | 3841 | BIOSQTL_methQTL_EUR_2017 | 27918535 |
| rs6747542 | 2p13.3 | C | T | cg09868768 | chr2:70188629 | ASPRV1 | | | 2.21E-14 | Whole blood | European | 3841 | BIOSQTL_methQTL_EUR_2017 | 27918535 |
| rs6747542 | 2p13.3 | C | T | cg04333863 | chr2:70121129 | SNRNP27 | -0.1975 | 0.02838 | 6.97E-12 | Whole blood | European | 837 | Gaunt-T_methQTL-15up_EUR_2016 | 27036880 |
| rs6747542 | 2p13.3 | C | T | cg13874290 | - | ANXA4 | -1.138 | 0.0741 | 3.40E-53 | Monocytes | European | 196 | BLUEPRINT_methQTL_EUR_2016 | 27863251 |
| rs13010313 | 2q22.3 | C | T | cg27635453 | chr2:145747901 | | | | 3.58E-14 | Whole blood | European | 3841 | BIOSQTL_methQTL_EUR_2017 | 27918535 |
| rs13010313 | 2q22.3 | C | T | cg05704183 | chr2:145764914 | | | | 9.67E-24 | Whole blood | European | 3841 | BIOSQTL_methQTL_EUR_2017 | 27918535 |
| rs13010313 | 2q22.3 | C | T | cg07679370 | chr2:145795773 | | | | 2.51E-14 | Whole blood | European | 3841 | BIOSQTL_methQTL_EUR_2017 | 27918535 |
| rs56181519 | 2q31.1 | C | T | cg09989631 | chr2:175543290 | WIPF1 | | | 1.76E-11 | Whole blood | European | 3841 | BIOSQTL_methQTL_EUR_2017 | 27918535 |
| rs56181519 | 2q31.1 | C | T | cg27337902 | chr2:175581979 | | | | 9.78E-29 | Whole blood | European | 3841 | BIOSQTL_methQTL_EUR_2017 | 27918535 |
| rs56181519 | 2q31.1 | C | T | cg00405190 | chr2:175545814 | WIPF1 | | | 9.11E-09 | Whole blood | European | 3841 | BIOSQTL_methQTL_EUR_2017 | 27918535 |
| rs56181519 | 2q31.1 | C | T | cg10037068 | chr2:175500048 | WIPF1 | | | 3.46E-28 | Whole blood | European | 3841 | BIOSQTL_methQTL_EUR_2017 | 27918535 |
| rs56181519 | 2q31.1 | C | T | cg12153668 | chr2:175560812 | | | | 6.53E-10 | Whole blood | European | 3841 | BIOSQTL_methQTL_EUR_2017 | 27918535 |
| rs56181519 | 2q31.1 | C | T | cg13651986 | chr2:175548471 | WIPF1 | | | 1.70E-12 | Whole blood | European | 3841 | BIOSQTL_methQTL_EUR_2017 | 27918535 |
| rs56181519 | 2q31.1 | C | T | cg01356950 | chr2:175546690 | WIPF1 | | | 4.22E-42 | Whole blood | European | 3841 | BIOSQTL_methQTL_EUR_2017 | 27918535 |

| | | | | | | | | | | | | | | |
|---|---|---|---|---|---|---|---|---|---|---|---|---|---|---|
| rs56181519 | 2q31.1 | C | T | cg25586410 | chr2:175350597 | GPR155 | | | 1.16E-08 | Whole blood | European | 3841 | BIOSQTL_methQTL_EUR_2017 | 27918535 |
| rs2857265 | 2q31.2 | G | A | cg18634211 | chr1:26737262 | LIN28 | -0.5044 | 0.06482 | 2.31E-14 | Cord blood | European | 771 | Gaunt-T_methQTL-Cord_EUR_2016 | 27036880 |
| rs2857265 | 2q31.2 | G | A | cg17740434 | chr2:179388064 | MIR548N | -0.5055 | 0.0642 | 1.18E-14 | Whole blood | European | 837 | Gaunt-T_methQTL-Antenatal_EUR_2016 | 27036880 |
| rs2857265 | 2q31.2 | G | A | cg14011486 | chr1:26737247 | LIN28 | -0.6767 | 0.06234 | 1.28E-25 | Whole blood | European | 837 | Gaunt-T_methQTL-Antenatal_EUR_2016 | 27036880 |
| rs2857265 | 2q31.2 | G | A | cg15609237 | chr2:179395885 | TTN;MIR548N | -0.2884 | 0.04477 | 2.06E-10 | Cord blood | European | 771 | Gaunt-T_methQTL-Cord_EUR_2016 | 27036880 |
| rs2857265 | 2q31.2 | G | A | cg15609237 | chr2:179395885 | TTN;MIR548N | -0.4024 | 0.0515 | 1.84E-14 | Whole blood | European | 837 | Gaunt-T_methQTL-Antenatal_EUR_2016 | 27036880 |
| rs2857265 | 2q31.2 | G | A | cg15609237 | chr2:179395885 | TTN;MIR548N | -0.234 | 0.04087 | 1.43E-08 | Whole blood | European | 837 | Gaunt-T_methQTL-15up_EUR_2016 | 27036880 |
| rs2857265 | 2q31.2 | G | A | cg14011486 | chr1:26737247 | LIN28 | -0.6629 | 0.06142 | 1.64E-25 | Whole blood | European | 837 | Gaunt-T_methQTL-15up_EUR_2016 | 27036880 |
| rs2857265 | 2q31.2 | G | A | cg08736813 | chr2:179396190 | TTN;MIR548N | | | 6.99E-41 | Whole blood | European | 3841 | BIOSQTL_methQTL_EUR_2017 | 27918535 |
| rs2857265 | 2q31.2 | G | A | cg15609237 | chr2:179395885 | TTN;MIR548N | -0.9277 | 0.1505 | 7.15E-10 | Neutrophils | European | 197 | BLUEPRINT_methQTL_EUR_2016 | 27863251 |
| rs2857265 | 2q31.2 | G | A | cg14011486 | chr1:26737247 | LIN28 | -0.6013 | 0.06208 | 5.18E-21 | Cord blood | European | 771 | Gaunt-T_methQTL-Cord_EUR_2016 | 27036880 |
| rs2857265 | 2q31.2 | G | A | cg10087519 | chr2:179397507 | TTN;MIR548N | | | 4.35E-11 | Whole blood | European | 3841 | BIOSQTL_methQTL_EUR_2017 | 27918535 |
| rs2857265 | 2q31.2 | G | A | cg09915519 | chr2:179397537 | TTN;MIR548N | | | 1.08E-33 | Whole blood | European | 3841 | BIOSQTL_methQTL_EUR_2017 | 27918535 |
| rs2857265 | 2q31.2 | G | A | cg08691235 | chr2:179395959 | TTN;MIR548N | | | 1.58E-18 | Whole blood | European | 3841 | BIOSQTL_methQTL_EUR_2017 | 27918535 |
| rs2857265 | 2q31.2 | G | A | cg04220104 | chr2:179387877 | MIR548N | | | 2.49E-12 | Whole blood | European | 3841 | BIOSQTL_methQTL_EUR_2017 | 27918535 |
| rs2857265 | 2q31.2 | G | A | cg17740434 | chr2:179388064 | MIR548N | -0.3603 | 0.06225 | 1.01E-08 | Whole blood | European | 837 | Gaunt-T_methQTL-15up_EUR_2016 | 27036880 |
| rs2857265 | 2q31.2 | G | A | cg18634211 | chr1:26737262 | LIN28 | -0.6258 | 0.06465 | 5.61E-21 | Whole blood | European | 837 | Gaunt-T_methQTL-Antenatal_EUR_2016 | 27036880 |
| rs2857265 | 2q31.2 | G | A | cg14011486 | chr1:26737247 | LIN28 | -0.5886 | 0.05925 | 4.73E-22 | Whole blood | European | 834 | Gaunt-T_methQTL-F7_EUR_2016 | 27036880 |
| rs2857265 | 2q31.2 | G | A | cg17740434 | chr2:179388088 | MIR548N | | | 4.01E-28 | Whole blood | European | 3841 | BIOSQTL_methQTL_EUR_2017 | 27918535 |
| rs2857265 | 2q31.2 | G | A | cg18634211 | chr1:26737262 | LIN28 | -0.567 | 0.06473 | 1.33E-17 | Whole blood | European | 742 | Gaunt-T_methQTL-FOM_EUR_2016 | 27036880 |
| rs2857265 | 2q31.2 | G | A | cg24560920 | chr2:179344411 | FKBP7;PLEKHA3;MIR548N | | | 3.19E-10 | Whole blood | European | 3841 | BIOSQTL_methQTL_EUR_2017 | 27918535 |
| rs2857265 | 2q31.2 | G | A | cg17740434 | chr2:179388064 | MIR548N | -0.4399 | 0.05935 | 3.07E-13 | Whole blood | European | 834 | Gaunt-T_methQTL-F7_EUR_2016 | 27036880 |
| rs2857265 | 2q31.2 | G | A | cg18634211 | chr1:26737262 | LIN28 | -0.5169 | 0.05798 | 3.03E-18 | Whole blood | European | 834 | Gaunt-T_methQTL-F7_EUR_2016 | 27036880 |
| rs2857265 | 2q31.2 | G | A | cg19906284 | chr2:179395888 | TTN;MIR548N | | | 2.99E-38 | Whole blood | European | 3841 | BIOSQTL_methQTL_EUR_2017 | 27918535 |
| rs2857265 | 2q31.2 | G | A | cg03991736 | chr2:179345179 | PLEKHA3;MIR548N | | | 5.79E-10 | Whole blood | European | 3841 | BIOSQTL_methQTL_EUR_2017 | 27918535 |
| rs2857265 | 2q31.2 | G | A | cg19906284 | chr2:179395912 | TTN;MIR548N | -0.9254 | 0.1506 | 8.02E-10 | Neutrophils | European | 197 | BLUEPRINT_methQTL_EUR_2016 | 27863251 |
| rs2857265 | 2q31.2 | G | A | cg19916659 | chr2:179387907 | MIR548N | | | 2.28E-10 | Whole blood | European | 3841 | BIOSQTL_methQTL_EUR_2017 | 27918535 |
| rs2857265 | 2q31.2 | G | A | cg14011486 | chr1:26737247 | LIN28 | -0.7328 | 0.0662 | 1.86E-26 | Whole blood | European | 742 | Gaunt-T_methQTL-FOM_EUR_2016 | 27036880 |
| rs2857265 | 2q31.2 | G | A | cg18634211 | chr1:26737262 | LIN28 | -0.5021 | 0.05961 | 1.60E-16 | Whole blood | European | 837 | Gaunt-T_methQTL-15up_EUR_2016 | 27036880 |
| rs2857265 | 2q31.2 | G | A | cg17740434 | chr2:179388064 | MIR548N | -0.4599 | 0.0666 | 1.08E-11 | Whole blood | European | 742 | Gaunt-T_methQTL-FOM_EUR_2016 | 27036880 |
| rs2857265 | 2q31.2 | G | A | cg15609237 | chr2:179395885 | TTN;MIR548N | -0.2531 | 0.03832 | 7.56E-11 | Whole blood | European | 742 | Gaunt-T_methQTL-FOM_EUR_2016 | 27036880 |

| | | | | | | | | | | | | | | |
|---|---|---|---|---|---|---|---|---|---|---|---|---|---|---|
| rs2857265 | 2q31.2 | G | A | cg15609237 | chr2:179395909 | TTN;MIR548N | | | 2.00E-51 | Whole blood | European | 3841 | BIOSQTL_methQTL_EUR_2017 | 27918535 |
| rs11099098 | 4q21.21 | G | T | cg10752158 | chr4:81184998 | | | | 1.73E-18 | Whole blood | European | 3841 | BIOSQTL_methQTL_EUR_2017 | 27918535 |
| rs11099098 | 4q21.21 | G | T | cg10880006 | chr4:81122663 | PRDM8;PRDM8 | | | 3.85E-08 | Whole blood | European | 3841 | BIOSQTL_methQTL_EUR_2017 | 27918535 |
| rs11099098 | 4q21.21 | G | T | cg12337525 | chr4:81185204 | | | | 7.91E-20 | Whole blood | European | 3841 | BIOSQTL_methQTL_EUR_2017 | 27918535 |
| rs11099098 | 4q21.21 | G | T | cg12635937 | chr4:81184949 | | | | 1.67E-34 | Whole blood | European | 3841 | BIOSQTL_methQTL_EUR_2017 | 27918535 |
| rs17042098 | 4q25 | A | G | cg03587884 | chr4:111642122 | | | | 6.63E-30 | Whole blood | European | 3841 | BIOSQTL_methQTL_EUR_2017 | 27918535 |
| rs17171711 | 5q31.2 | C | T | cg19880597 | chr5:136965606 | KLHL3 | -0.3245 | 0.05435 | 3.49E-09 | Whole blood | European | 834 | Gaunt-T_methQTL-F7_EUR_2016 | 27036880 |
| rs17171711 | 5q31.2 | C | T | cg07848042 | chr5:137667509 | CDC25C | -0.2403 | 0.0436 | 4.87E-08 | Whole blood | European | 837 | Gaunt-T_methQTL-Antenatal_EUR_2016 | 27036880 |
| rs17171711 | 5q31.2 | C | T | cg19880597 | chr5:136965606 | KLHL3 | -0.3106 | 0.05419 | 1.43E-08 | Whole blood | European | 837 | Gaunt-T_methQTL-Antenatal_EUR_2016 | 27036880 |
| rs17171711 | 5q31.2 | C | T | cg03684156 | chr5:137514661 | BRD8;KIF20A | -0.3448 | 0.05 | 1.15E-11 | Whole blood | European | 742 | Gaunt-T_methQTL-FOM_EUR_2016 | 27036880 |
| rs17171711 | 5q31.2 | C | T | cg09817016 | chr5:137690920 | KDM3B | -0.4327 | 0.05365 | 2.84E-15 | Whole blood | European | 837 | Gaunt-T_methQTL-Antenatal_EUR_2016 | 27036880 |
| rs17171711 | 5q31.2 | C | T | cg19880597 | chr5:136965606 | KLHL3 | -0.293 | 0.05266 | 3.68E-08 | Whole blood | European | 742 | Gaunt-T_methQTL-FOM_EUR_2016 | 27036880 |
| rs17171711 | 5q31.2 | C | T | cg22657492 | chr5:137593464 | GFRA3 | | | 4.03E-39 | Whole blood | European | 3841 | BIOSQTL_methQTL_EUR_2017 | 27918535 |
| rs17171711 | 5q31.2 | C | T | cg09817016 | chr5:137690920 | KDM3B | -0.3295 | 0.04796 | 1.37E-11 | Whole blood | European | 742 | Gaunt-T_methQTL-FOM_EUR_2016 | 27036880 |
| rs17171711 | 5q31.2 | C | T | cg09817016 | chr5:137690920 | KDM3B | -0.3721 | 0.05171 | 1.39E-12 | Whole blood | European | 837 | Gaunt-T_methQTL-15up_EUR_2016 | 27036880 |
| rs17171711 | 5q31.2 | C | T | cg11518421 | chr5:137667529 | CDC25C | -0.3158 | 0.05282 | 3.46E-09 | Whole blood | European | 837 | Gaunt-T_methQTL-Antenatal_EUR_2016 | 27036880 |
| rs17171711 | 5q31.2 | C | T | cg19880597 | chr5:136965606 | KLHL3 | -0.3892 | 0.0602 | 1.81E-10 | Cord blood | European | 771 | Gaunt-T_methQTL-Cord_EUR_2016 | 27036880 |
| rs17171711 | 5q31.2 | C | T | cg03684156 | chr5:137514661 | BRD8;KIF20A | -0.3626 | 0.04844 | 1.81E-13 | Whole blood | European | 837 | Gaunt-T_methQTL-15up_EUR_2016 | 27036880 |
| rs17171711 | 5q31.2 | C | T | cg03684156 | chr5:137514637 | BRD8;KIF20A | | | 1.26E-17 | Whole blood | European | 3841 | BIOSQTL_methQTL_EUR_2017 | 27918535 |
| rs17171711 | 5q31.2 | C | T | cg03684156 | chr5:137514661 | BRD8;KIF20A | -0.3087 | 0.04666 | 6.88E-11 | Whole blood | European | 837 | Gaunt-T_methQTL-Antenatal_EUR_2016 | 27036880 |
| rs17171711 | 5q31.2 | C | T | cg09817016 | chr5:137690920 | KDM3B | -0.3745 | 0.05059 | 3.28E-13 | Whole blood | European | 834 | Gaunt-T_methQTL-F7_EUR_2016 | 27036880 |
| rs17171711 | 5q31.2 | C | T | cg19880597 | chr5:136965606 | KLHL3 | -0.3746 | 0.05269 | 2.50E-12 | Whole blood | European | 837 | Gaunt-T_methQTL-15up_EUR_2016 | 27036880 |
| rs17171711 | 5q31.2 | C | T | cg03684156 | chr5:137514661 | BRD8;KIF20A | -0.3748 | 0.04746 | 9.06E-15 | Whole blood | European | 834 | Gaunt-T_methQTL-F7_EUR_2016 | 27036880 |
| rs6891790 | 5q35.1 | G | T | cg01753208 | chr5:172666071 | | -0.434 | 0.04515 | 1.09E-20 | Whole blood | European | 742 | Gaunt-T_methQTL-FOM_EUR_2016 | 27036880 |
| rs6891790 | 5q35.1 | G | T | cg12825773 | chr5:172662463 | NKX2-5 | 0.3161 | 0.04636 | 1.87E-11 | Whole blood | European | 837 | Gaunt-T_methQTL-Antenatal_EUR_2016 | 27036880 |
| rs6891790 | 5q35.1 | G | T | cg06974592 | chr5:172665173 | | | | 7.72E-10 | Whole blood | European | 3841 | BIOSQTL_methQTL_EUR_2017 | 27918535 |
| rs6891790 | 5q35.1 | G | T | cg12825773 | chr5:172662463 | NKX2-5 | 0.3356 | 0.04463 | 1.44E-13 | Whole blood | European | 834 | Gaunt-T_methQTL-F7_EUR_2016 | 27036880 |
| rs6891790 | 5q35.1 | G | T | cg12825773 | chr5:172662463 | NKX2-5 | 0.3216 | 0.04726 | 2.05E-11 | Cord blood | European | 771 | Gaunt-T_methQTL-Cord_EUR_2016 | 27036880 |
| rs6891790 | 5q35.1 | G | T | cg01753208 | chr5:172666071 | | -0.8492 | 0.0995 | 1.38E-17 | Monocytes | European | 196 | BLUEPRINT_methQTL_EUR_2016 | 27863251 |
| rs6891790 | 5q35.1 | G | T | cg13021417 | chr5:172640615 | | 0.2094 | 0.03564 | 6.12E-09 | Whole blood | European | 834 | Gaunt-T_methQTL-F7_EUR_2016 | 27036880 |
| rs6891790 | 5q35.1 | G | T | cg02692390 | chr5:172657441 | | | | 6.24E-14 | Whole blood | European | 3841 | BIOSQTL_methQTL_EUR_2017 | 27918535 |

| | | | | | | | | | | | | | | |
|---|---|---|---|---|---|---|---|---|---|---|---|---|---|---|
| rs6891790 | 5q35.1 | G | T | cg14980667 | chr5:172666228 | | -0.2888 | 0.04161 | 7.93E-12 | Whole blood | European | 834 | Gaunt-T_methQTL-F7_EUR_2016 | 27036880 |
| rs6891790 | 5q35.1 | G | T | cg18512156 | chr5:172661946 | NKX2-5 | | | 3.41E-10 | Whole blood | European | 3841 | BIOSQTL_methQTL_EUR_2017 | 27918535 |
| rs6891790 | 5q35.1 | G | T | cg13021417 | chr5:172640615 | | 0.2151 | 0.03517 | 1.48E-09 | Whole blood | European | 837 | Gaunt-T_methQTL-15up_EUR_2016 | 27036880 |
| rs6891790 | 5q35.1 | G | T | cg19423543 | chr5:172655972 | | | | 1.75E-24 | Whole blood | European | 3841 | BIOSQTL_methQTL_EUR_2017 | 27918535 |
| rs6891790 | 5q35.1 | G | T | cg14980667 | chr5:172666228 | | -0.2661 | 0.04578 | 9.12E-09 | Whole blood | European | 742 | Gaunt-T_methQTL-FOM_EUR_2016 | 27036880 |
| rs6891790 | 5q35.1 | G | T | cg13021417 | chr5:172640615 | | 0.2459 | 0.03588 | 1.48E-11 | Cord blood | European | 771 | Gaunt-T_methQTL-Cord_EUR_2016 | 27036880 |
| rs6891790 | 5q35.1 | G | T | cg12825773 | chr5:172662487 | NKX2-5 | | | 9.36E-114 | Whole blood | European | 3841 | BIOSQTL_methQTL_EUR_2017 | 27918535 |
| rs6891790 | 5q35.1 | G | T | cg13570101 | chr5:172661835 | NKX2-5 | | | 4.19E-30 | Whole blood | European | 3841 | BIOSQTL_methQTL_EUR_2017 | 27918535 |
| rs6891790 | 5q35.1 | G | T | cg08418980 | chr5:172665424 | | | | 2.07E-68 | Whole blood | European | 3841 | BIOSQTL_methQTL_EUR_2017 | 27918535 |
| rs6891790 | 5q35.1 | G | T | cg13021417 | chr5:172640615 | | 0.2334 | 0.03333 | 5.62E-12 | Whole blood | European | 742 | Gaunt-T_methQTL-FOM_EUR_2016 | 27036880 |
| rs6891790 | 5q35.1 | G | T | cg13004182 | chr5:172665683 | | | | 1.17E-39 | Whole blood | European | 3841 | BIOSQTL_methQTL_EUR_2017 | 27918535 |
| rs6891790 | 5q35.1 | G | T | cg01753208 | chr5:172666071 | | -0.6284 | 0.1042 | 1.65E-09 | Neutrophils | European | 197 | BLUEPRINT_methQTL_EUR_2016 | 27863251 |
| rs6891790 | 5q35.1 | G | T | cg01753208 | chr5:172666071 | | -0.4812 | 0.04313 | 5.01E-27 | Whole blood | European | 834 | Gaunt-T_methQTL-F7_EUR_2016 | 27036880 |
| rs6891790 | 5q35.1 | G | T | cg01753208 | chr5:172666071 | | -0.8472 | 0.1199 | 1.61E-12 | T cells | European | 132 | BLUEPRINT_methQTL_EUR_2016 | 27863251 |
| rs6891790 | 5q35.1 | G | T | cg14980667 | chr5:172666204 | | | | 8.14E-208 | Whole blood | European | 3841 | BIOSQTL_methQTL_EUR_2017 | 27918535 |
| rs6891790 | 5q35.1 | G | T | cg03219908 | chr5:172659488 | NKX2-5 | | | 5.14E-16 | Whole blood | European | 3841 | BIOSQTL_methQTL_EUR_2017 | 27918535 |
| rs6891790 | 5q35.1 | G | T | cg18839504 | chr5:172665095 | | | | 4.23E-34 | Whole blood | European | 3841 | BIOSQTL_methQTL_EUR_2017 | 27918535 |
| rs6891790 | 5q35.1 | G | T | cg14980667 | chr5:172666228 | | -0.2801 | 0.04572 | 1.43E-09 | Cord blood | European | 771 | Gaunt-T_methQTL-Cord_EUR_2016 | 27036880 |
| rs6891790 | 5q35.1 | G | T | cg12825773 | chr5:172662463 | NKX2-5 | 0.3211 | 0.04485 | 1.77E-12 | Whole blood | European | 837 | Gaunt-T_methQTL-15up_EUR_2016 | 27036880 |
| rs6891790 | 5q35.1 | G | T | cg19506433 | - | | 0.6238 | 0.1049 | 2.74E-09 | Monocytes | European | 196 | BLUEPRINT_methQTL_EUR_2016 | 27863251 |
| rs6891790 | 5q35.1 | G | T | cg08814105 | chr5:172662106 | NKX2-5 | | | 1.17E-19 | Whole blood | European | 3841 | BIOSQTL_methQTL_EUR_2017 | 27918535 |
| rs6891790 | 5q35.1 | G | T | cg15982655 | chr5:172571401 | BNIP1 | | | 1.32E-16 | Whole blood | European | 3841 | BIOSQTL_methQTL_EUR_2017 | 27918535 |
| rs6891790 | 5q35.1 | G | T | cg14980667 | chr5:172666228 | | -0.2975 | 0.04329 | 1.33E-11 | Whole blood | European | 837 | Gaunt-T_methQTL-Antenatal_EUR_2016 | 27036880 |
| rs6891790 | 5q35.1 | G | T | cg08386832 | chr5:172482831 | C5orf41 | | | 1.21E-15 | Whole blood | European | 3841 | BIOSQTL_methQTL_EUR_2017 | 27918535 |
| rs6891790 | 5q35.1 | G | T | cg14980667 | chr5:172666228 | | -0.7043 | 0.1236 | 1.21E-08 | T cells | European | 132 | BLUEPRINT_methQTL_EUR_2016 | 27863251 |
| rs6891790 | 5q35.1 | G | T | cg01753208 | chr5:172666071 | | -0.4271 | 0.04593 | 1.21E-19 | Whole blood | European | 837 | Gaunt-T_methQTL-15up_EUR_2016 | 27036880 |
| rs6891790 | 5q35.1 | G | T | cg01753208 | chr5:172666095 | | | | 4.75E-306 | Whole blood | European | 3841 | BIOSQTL_methQTL_EUR_2017 | 27918535 |
| rs6891790 | 5q35.1 | G | T | cg22761308 | chr5:172571414 | BNIP1 | | | 2.06E-09 | Whole blood | European | 3841 | BIOSQTL_methQTL_EUR_2017 | 27918535 |
| rs6891790 | 5q35.1 | G | T | cg07798980 | chr5:172659265 | NKX2-5 | | | 3.82E-09 | Whole blood | European | 3841 | BIOSQTL_methQTL_EUR_2017 | 27918535 |
| rs6891790 | 5q35.1 | G | T | cg11330740 | chr5:172660860 | NKX2-5 | | | 7.46E-80 | Whole blood | European | 3841 | BIOSQTL_methQTL_EUR_2017 | 27918535 |
| rs6891790 | 5q35.1 | G | T | cg14980667 | chr5:172666228 | | -0.5885 | 0.105 | 2.11E-08 | Neutrophils | European | 197 | BLUEPRINT_methQTL_EUR_2016 | 27863251 |

| | | | | | | | | | | | | | | |
|---|---|---|---|---|---|---|---|---|---|---|---|---|---|---|
| rs6891790 | 5q35.1 | G | T | cg01753208 | chr5:172666071 | | -0.3668 | 0.04582 | 4.46E-15 | Whole blood | European | 837 | Gaunt-T_methQTL-Antenatal_EUR_2016 | 27036880 |
| rs6891790 | 5q35.1 | G | T | cg03219908 | chr5:172659512 | NKX2-5 | 0.3116 | 0.04596 | 2.29E-11 | Whole blood | European | 837 | Gaunt-T_methQTL-15up_EUR_2016 | 27036880 |
| rs6891790 | 5q35.1 | G | T | cg19506433 | - | | 0.7017 | 0.124 | 1.54E-08 | T cells | European | 132 | BLUEPRINT_methQTL_EUR_2016 | 27863251 |
| rs6891790 | 5q35.1 | G | T | cg10895922 | chr5:172662291 | NKX2-5 | | | 3.02E-10 | Whole blood | European | 3841 | BIOSQTL_methQTL_EUR_2017 | 27918535 |
| rs6891790 | 5q35.1 | G | T | cg19506433 | chr5:172670494 | | | | 2.20E-148 | Whole blood | European | 3841 | BIOSQTL_methQTL_EUR_2017 | 27918535 |
| rs6891790 | 5q35.1 | G | T | cg14980667 | chr5:172666228 | | -0.3833 | 0.0435 | 7.15E-18 | Whole blood | European | 837 | Gaunt-T_methQTL-15up_EUR_2016 | 27036880 |
| rs6891790 | 5q35.1 | G | T | cg03625415 | chr5:172662717 | NKX2-5 | | | 4.38E-11 | Whole blood | European | 3841 | BIOSQTL_methQTL_EUR_2017 | 27918535 |
| rs6891790 | 5q35.1 | G | T | cg21165027 | chr5:172663808 | NKX2-5 | | | 7.79E-10 | Whole blood | European | 3841 | BIOSQTL_methQTL_EUR_2017 | 27918535 |
| rs6891790 | 5q35.1 | G | T | cg24767237 | chr5:172662748 | NKX2-5 | | | 1.12E-12 | Whole blood | European | 3841 | BIOSQTL_methQTL_EUR_2017 | 27918535 |
| rs6891790 | 5q35.1 | G | T | cg11330740 | chr5:172660884 | NKX2-5 | -0.2682 | 0.04648 | 1.15E-08 | Whole blood | European | 837 | Gaunt-T_methQTL-Antenatal_EUR_2016 | 27036880 |
| rs6891790 | 5q35.1 | G | T | cg09143673 | chr5:172664415 | | | | 1.50E-10 | Whole blood | European | 3841 | BIOSQTL_methQTL_EUR_2017 | 27918535 |
| rs6891790 | 5q35.1 | G | T | cg12825773 | chr5:172662463 | NKX2-5 | 0.2733 | 0.04798 | 1.76E-08 | Whole blood | European | 742 | Gaunt-T_methQTL-FOM_EUR_2016 | 27036880 |
| rs3176326 | 6p21.2 | A | G | cg02730678 | chr6:36641554 | | -0.9649 | 0.1154 | 6.25E-17 | Monocytes | European | 196 | BLUEPRINT_methQTL_EUR_2016 | 27863251 |
| rs3176326 | 6p21.2 | A | G | cg03714916 | chr6:36645910 | CDKN1A | | | 1.25E-107 | Whole blood | European | 3841 | BIOSQTL_methQTL_EUR_2017 | 27918535 |
| rs3176326 | 6p21.2 | A | G | cg15474579 | chr6:36645812 | CDKN1A | -0.476 | 0.05107 | 1.20E-19 | Cord blood | European | 771 | Gaunt-T_methQTL-Cord_EUR_2016 | 27036880 |
| rs3176326 | 6p21.2 | A | G | cg15474579 | chr6:36645812 | CDKN1A | -0.7215 | 0.1201 | 1.87E-09 | Neutrophils | European | 197 | BLUEPRINT_methQTL_EUR_2016 | 27863251 |
| rs3176326 | 6p21.2 | A | G | cg11920449 | - | CDKN1A | -1.086 | 0.1108 | 1.13E-22 | Monocytes | European | 196 | BLUEPRINT_methQTL_EUR_2016 | 27863251 |
| rs3176326 | 6p21.2 | A | G | cg02730678 | chr6:36641554 | | -0.4587 | 0.05044 | 6.92E-19 | Whole blood | European | 834 | Gaunt-T_methQTL-F7_EUR_2016 | 27036880 |
| rs3176326 | 6p21.2 | A | G | cg15474579 | chr6:36645812 | CDKN1A | -0.767 | 0.119 | 1.15E-10 | Monocytes | European | 196 | BLUEPRINT_methQTL_EUR_2016 | 27863251 |
| rs3176326 | 6p21.2 | A | G | cg02730678 | chr6:36641530 | | | | 9.54E-46 | Whole blood | European | 3841 | BIOSQTL_methQTL_EUR_2017 | 27918535 |
| rs3176326 | 6p21.2 | A | G | cg24425727 | - | CDKN1A | -0.8371 | 0.1408 | 2.73E-09 | T cells | European | 132 | BLUEPRINT_methQTL_EUR_2016 | 27863251 |
| rs3176326 | 6p21.2 | A | G | cg03714916 | chr6:36645886 | CDKN1A | -0.4326 | 0.05129 | 1.66E-16 | Cord blood | European | 771 | Gaunt-T_methQTL-Cord_EUR_2016 | 27036880 |
| rs3176326 | 6p21.2 | A | G | cg15474579 | chr6:36645812 | CDKN1A | -0.4778 | 0.04855 | 1.08E-21 | Whole blood | European | 837 | Gaunt-T_methQTL-15up_EUR_2016 | 27036880 |
| rs3176326 | 6p21.2 | A | G | cg15474579 | chr6:36645812 | CDKN1A | -0.3857 | 0.04675 | 7.18E-16 | Whole blood | European | 742 | Gaunt-T_methQTL-FOM_EUR_2016 | 27036880 |
| rs3176326 | 6p21.2 | A | G | cg08179530 | chr6:36648295 | CDKN1A | 0.6667 | 0.1203 | 3.03E-08 | Neutrophils | European | 197 | BLUEPRINT_methQTL_EUR_2016 | 27863251 |
| rs3176326 | 6p21.2 | A | G | cg02730678 | chr6:36641554 | | -0.5538 | 0.05038 | 2.43E-26 | Whole blood | European | 837 | Gaunt-T_methQTL-15up_EUR_2016 | 27036880 |
| rs3176326 | 6p21.2 | A | G | cg03032677 | chr6:36648794 | CDKN1A | 0.4339 | 0.05085 | 6.65E-17 | Whole blood | European | 837 | Gaunt-T_methQTL-15up_EUR_2016 | 27036880 |
| rs3176326 | 6p21.2 | A | G | cg08179530 | chr6:36648295 | CDKN1A | 0.8921 | 0.1167 | 2.13E-14 | Monocytes | European | 196 | BLUEPRINT_methQTL_EUR_2016 | 27863251 |
| rs3176326 | 6p21.2 | A | G | cg11920449 | - | CDKN1A | -0.7857 | 0.1413 | 2.68E-08 | T cells | European | 132 | BLUEPRINT_methQTL_EUR_2016 | 27863251 |
| rs3176326 | 6p21.2 | A | G | cg03032677 | chr6:36648794 | CDKN1A | 0.4116 | 0.04874 | 1.35E-16 | Whole blood | European | 834 | Gaunt-T_methQTL-F7_EUR_2016 | 27036880 |
| rs3176326 | 6p21.2 | A | G | cg02730678 | chr6:36641554 | | -0.5452 | 0.05106 | 6.78E-25 | Whole blood | European | 837 | Gaunt-T_methQTL-Antenatal_EUR_2016 | 27036880 |

| | | | | | | | | | | | | | | |
|---|---|---|---|---|---|---|---|---|---|---|---|---|---|---|
| rs3176326 | 6p21.2 | A | G | cg24425727 | - | CDKN1A | -1.081 | 0.111 | 2.10E-22 | Neutrophils | European | 197 | BLUEPRINT_methQTL_EUR_2016 | 27863251 |
| rs3176326 | 6p21.2 | A | G | cg24425727 | - | CDKN1A | -1.049 | 0.112 | 7.38E-21 | Monocytes | European | 196 | BLUEPRINT_methQTL_EUR_2016 | 27863251 |
| rs3176326 | 6p21.2 | A | G | cg18206424 | chr6:36629615 | | | | 2.75E-16 | Whole blood | European | 3841 | BIOSQTL_methQTL_EUR_2017 | 27918535 |
| rs3176326 | 6p21.2 | A | G | cg02730678 | chr6:36641554 | | -0.5192 | 0.05189 | 3.09E-22 | Cord blood | European | 771 | Gaunt-T_methQTL-Cord_EUR_2016 | 27036880 |
| rs3176326 | 6p21.2 | A | G | cg03032677 | chr6:36648794 | CDKN1A | 0.5013 | 0.05177 | 5.88E-21 | Whole blood | European | 742 | Gaunt-T_methQTL-FOM_EUR_2016 | 27036880 |
| rs3176326 | 6p21.2 | A | G | cg03032677 | chr6:36648794 | CDKN1A | 0.3533 | 0.05042 | 5.28E-12 | Cord blood | European | 771 | Gaunt-T_methQTL-Cord_EUR_2016 | 27036880 |
| rs3176326 | 6p21.2 | A | G | cg08179530 | chr6:36648271 | CDKN1A | | | 7.77E-113 | Whole blood | European | 3841 | BIOSQTL_methQTL_EUR_2017 | 27918535 |
| rs3176326 | 6p21.2 | A | G | cg03032677 | chr6:36648818 | CDKN1A | | | 1.07E-142 | Whole blood | European | 3841 | BIOSQTL_methQTL_EUR_2017 | 27918535 |
| rs3176326 | 6p21.2 | A | G | cg03032677 | chr6:36648794 | CDKN1A | 0.5024 | 0.05094 | 1.14E-21 | Whole blood | European | 837 | Gaunt-T_methQTL-Antenatal_EUR_2016 | 27036880 |
| rs3176326 | 6p21.2 | A | G | cg15474579 | chr6:36645812 | CDKN1A | -0.4568 | 0.05069 | 1.60E-18 | Whole blood | European | 837 | Gaunt-T_methQTL-Antenatal_EUR_2016 | 27036880 |
| rs3176326 | 6p21.2 | A | G | cg05389236 | chr6:36635063 | | | | 1.63E-22 | Whole blood | European | 3841 | BIOSQTL_methQTL_EUR_2017 | 27918535 |
| rs3176326 | 6p21.2 | A | G | cg15474579 | chr6:36645812 | CDKN1A | -0.476 | 0.04768 | 3.06E-22 | Whole blood | European | 834 | Gaunt-T_methQTL-F7_EUR_2016 | 27036880 |
| rs3176326 | 6p21.2 | A | G | cg02730678 | chr6:36641554 | | -0.9291 | 0.1152 | 7.21E-16 | Neutrophils | European | 197 | BLUEPRINT_methQTL_EUR_2016 | 27863251 |
| rs3176326 | 6p21.2 | A | G | cg12231373 | chr6:36628079 | | | | 4.45E-09 | Whole blood | European | 3841 | BIOSQTL_methQTL_EUR_2017 | 27918535 |
| rs3176326 | 6p21.2 | A | G | cg15474579 | chr6:36645788 | CDKN1A | | | 3.97E-221 | Whole blood | European | 3841 | BIOSQTL_methQTL_EUR_2017 | 27918535 |
| rs3176326 | 6p21.2 | A | G | cg03714916 | chr6:36645886 | CDKN1A | -0.2925 | 0.04766 | 1.30E-09 | Whole blood | European | 837 | Gaunt-T_methQTL-15up_EUR_2016 | 27036880 |
| rs3176326 | 6p21.2 | A | G | cg02730678 | chr6:36641554 | | -0.5277 | 0.05197 | 9.22E-23 | Whole blood | European | 742 | Gaunt-T_methQTL-FOM_EUR_2016 | 27036880 |
| rs3176326 | 6p21.2 | A | G | cg15474579 | chr6:36645812 | CDKN1A | -0.852 | 0.1395 | 1.02E-09 | T cells | European | 132 | BLUEPRINT_methQTL_EUR_2016 | 27863251 |
| rs3176326 | 6p21.2 | A | G | cg11920449 | - | CDKN1A | -1.073 | 0.1113 | 5.29E-22 | Neutrophils | European | 197 | BLUEPRINT_methQTL_EUR_2016 | 27863251 |
| rs3176326 | 6p21.2 | A | G | cg15106294 | chr6:36626068 | | | | 4.92E-12 | Whole blood | European | 3841 | BIOSQTL_methQTL_EUR_2017 | 27918535 |
| rs12211255 | 6q14.1 | A | C | cg19367436 | chr6:75953959 | COX7A2 | | | 3.21E-10 | Whole blood | European | 3841 | BIOSQTL_methQTL_EUR_2017 | 27918535 |
| rs12211255 | 6q14.1 | A | C | cg06809298 | chr6:75953877 | COX7A2 | | | 1.42E-15 | Whole blood | European | 3841 | BIOSQTL_methQTL_EUR_2017 | 27918535 |
| rs12211255 | 6q14.1 | A | C | cg25716013 | chr6:75954029 | COX7A2 | | | 9.89E-16 | Whole blood | European | 3841 | BIOSQTL_methQTL_EUR_2017 | 27918535 |
| rs3807989 | 7q31.2 | A | G | cg00917413 | chr7:116138591 | CAV2 | -0.2897 | 0.04468 | 1.60E-10 | Whole blood | European | 837 | Gaunt-T_methQTL-Antenatal_EUR_2016 | 27036880 |
| rs3807989 | 7q31.2 | A | G | cg16553024 | chr7:116138462 | CAV2 | -0.2924 | 0.04555 | 2.30E-10 | Whole blood | European | 837 | Gaunt-T_methQTL-15up_EUR_2016 | 27036880 |
| rs3807989 | 7q31.2 | A | G | cg12739419 | chr7:116140617 | CAV2 | | | 3.73E-170 | Whole blood | European | 3841 | BIOSQTL_methQTL_EUR_2017 | 27918535 |
| rs3807989 | 7q31.2 | A | G | cg16553024 | chr7:116138462 | CAV2 | -0.2732 | 0.04542 | 2.68E-09 | Whole blood | European | 834 | Gaunt-T_methQTL-F7_EUR_2016 | 27036880 |
| rs3807989 | 7q31.2 | A | G | cg05232628 | chr7:116200160 | CAV1 | | | 3.19E-34 | Whole blood | European | 3841 | BIOSQTL_methQTL_EUR_2017 | 27918535 |
| rs3807989 | 7q31.2 | A | G | cg07838272 | chr7:116164694 | CAV1 | | | 2.28E-08 | Whole blood | European | 3841 | BIOSQTL_methQTL_EUR_2017 | 27918535 |
| rs3807989 | 7q31.2 | A | G | cg12739419 | chr7:116140593 | CAV2 | 0.4873 | 0.04132 | 8.41E-30 | Whole blood | European | 837 | Gaunt-T_methQTL-15up_EUR_2016 | 27036880 |
| rs3807989 | 7q31.2 | A | G | cg00917413 | chr7:116138591 | CAV2 | -0.2684 | 0.04318 | 8.12E-10 | Whole blood | European | 834 | Gaunt-T_methQTL-F7_EUR_2016 | 27036880 |

| | | | | | | | | | | | | | | |
|---|---|---|---|---|---|---|---|---|---|---|---|---|---|---|
| rs3807989 | 7q31.2 | A | G | cg00917413 | chr7:116138591 | CAV2 | -0.3209 | 0.04425 | 9.39E-13 | Whole blood | European | 837 | Gaunt-T_methQTL-15up_EUR_2016 | 27036880 |
| rs3807989 | 7q31.2 | A | G | cg16260298 | chr7:116140366 | CAV2 | | | 5.92E-15 | Whole blood | European | 3841 | BIOSQTL_methQTL_EUR_2017 | 27918535 |
| rs3807989 | 7q31.2 | A | G | cg00917413 | chr7:116138591 | CAV2 | -0.3277 | 0.04573 | 1.81E-12 | Cord blood | European | 771 | Gaunt-T_methQTL-Cord_EUR_2016 | 27036880 |
| rs3807989 | 7q31.2 | A | G | cg18887458 | chr7:115995958 | | | | 1.51E-19 | Whole blood | European | 3841 | BIOSQTL_methQTL_EUR_2017 | 27918535 |
| rs3807989 | 7q31.2 | A | G | cg12739419 | chr7:116140593 | CAV2 | 0.4198 | 0.04206 | 3.87E-22 | Whole blood | European | 837 | Gaunt-T_methQTL-Antenatal_EUR_2016 | 27036880 |
| rs3807989 | 7q31.2 | A | G | cg12739419 | chr7:116140593 | CAV2 | 0.4732 | 0.03862 | 7.45E-32 | Whole blood | European | 834 | Gaunt-T_methQTL-F7_EUR_2016 | 27036880 |
| rs3807989 | 7q31.2 | A | G | cg12739419 | chr7:116140593 | CAV2 | 0.3321 | 0.04361 | 8.09E-14 | Whole blood | European | 742 | Gaunt-T_methQTL-FOM_EUR_2016 | 27036880 |
| rs3807989 | 7q31.2 | A | G | cg04474049 | chr7:116164509 | CAV1 | | | 1.36E-53 | Whole blood | European | 3841 | BIOSQTL_methQTL_EUR_2017 | 27918535 |
| rs3807989 | 7q31.2 | A | G | cg16553024 | chr7:116138438 | CAV2 | | | 2.86E-18 | Whole blood | European | 3841 | BIOSQTL_methQTL_EUR_2017 | 27918535 |
| rs3807989 | 7q31.2 | A | G | cg01265597 | chr7:116164657 | CAV1 | | | 6.80E-12 | Whole blood | European | 3841 | BIOSQTL_methQTL_EUR_2017 | 27918535 |
| rs3807989 | 7q31.2 | A | G | cg12739419 | chr7:116140593 | CAV2 | 0.3701 | 0.04359 | 1.05E-16 | Cord blood | European | 771 | Gaunt-T_methQTL-Cord_EUR_2016 | 27036880 |
| rs3807989 | 7q31.2 | A | G | cg00917413 | chr7:116138615 | CAV2 | | | 1.20E-43 | Whole blood | European | 3841 | BIOSQTL_methQTL_EUR_2017 | 27918535 |
| rs3807989 | 7q31.2 | A | G | cg25274503 | chr7:116140128 | CAV2 | -0.2516 | 0.04087 | 1.16E-09 | Whole blood | European | 834 | Gaunt-T_methQTL-F7_EUR_2016 | 27036880 |
| rs3807989 | 7q31.2 | A | G | cg12739419 | chr7:116140593 | CAV2 | 0.6773 | 0.0994 | 9.64E-12 | Monocytes | European | 196 | BLUEPRINT_methQTL_EUR_2016 | 27863251 |
| rs3807989 | 7q31.2 | A | G | cg17469978 | chr7:116164680 | CAV1 | | | 8.51E-14 | Whole blood | European | 3841 | BIOSQTL_methQTL_EUR_2017 | 27918535 |
| rs3807989 | 7q31.2 | A | G | cg12771187 | chr7:116152034 | | | | 4.96E-45 | Whole blood | European | 3841 | BIOSQTL_methQTL_EUR_2017 | 27918535 |
| rs3807989 | 7q31.2 | A | G | cg12739419 | chr7:116140593 | CAV2 | 0.6773 | 0.1167 | 6.56E-09 | T cells | European | 132 | BLUEPRINT_methQTL_EUR_2016 | 27863251 |
| rs3807989 | 7q31.2 | A | G | cg25274503 | chr7:116140152 | CAV2 | | | 1.30E-09 | Whole blood | European | 3841 | BIOSQTL_methQTL_EUR_2017 | 27918535 |
| rs55985730 | 7q32.1 | G | T | cg10826733 | chr7:128482561 | FLNC | -0.7709 | 0.0854 | 1.41E-18 | Whole blood | European | 837 | Gaunt-T_methQTL-Antenatal_EUR_2016 | 27036880 |
| rs55985730 | 7q32.1 | G | T | cg13951589 | chr7:128482263 | FLNC | | | 3.57E-56 | Whole blood | European | 3841 | BIOSQTL_methQTL_EUR_2017 | 27918535 |
| rs55985730 | 7q32.1 | G | T | cg18693656 | chr7:128482715 | FLNC | | | 4.46E-30 | Whole blood | European | 3841 | BIOSQTL_methQTL_EUR_2017 | 27918535 |
| rs55985730 | 7q32.1 | G | T | cg25664034 | chr7:128469867 | FLNC | | | 3.34E-12 | Whole blood | European | 3841 | BIOSQTL_methQTL_EUR_2017 | 27918535 |
| rs55985730 | 7q32.1 | G | T | cg10826733 | chr7:128482561 | FLNC | -0.886 | 0.09002 | 1.31E-21 | Cord blood | European | 771 | Gaunt-T_methQTL-Cord_EUR_2016 | 27036880 |
| rs55985730 | 7q32.1 | G | T | cg25303427 | chr7:128482679 | FLNC | -0.504 | 0.09007 | 3.09E-08 | Whole blood | European | 742 | Gaunt-T_methQTL-FOM_EUR_2016 | 27036880 |
| rs55985730 | 7q32.1 | G | T | cg25303427 | chr7:128482679 | FLNC | -0.6325 | 0.08989 | 4.37E-12 | Cord blood | European | 771 | Gaunt-T_methQTL-Cord_EUR_2016 | 27036880 |
| rs55985730 | 7q32.1 | G | T | cg14678430 | chr7:128494126 | FLNC | | | 2.39E-10 | Whole blood | European | 3841 | BIOSQTL_methQTL_EUR_2017 | 27918535 |
| rs55985730 | 7q32.1 | G | T | cg15031685 | chr7:128494036 | FLNC | | | 3.47E-19 | Whole blood | European | 3841 | BIOSQTL_methQTL_EUR_2017 | 27918535 |
| rs55985730 | 7q32.1 | G | T | cg03379100 | chr7:128494201 | FLNC | | | 4.44E-11 | Whole blood | European | 3841 | BIOSQTL_methQTL_EUR_2017 | 27918535 |
| rs55985730 | 7q32.1 | G | T | cg10826733 | chr7:128482561 | FLNC | -0.8159 | 0.08346 | 1.94E-21 | Whole blood | European | 834 | Gaunt-T_methQTL-F7_EUR_2016 | 27036880 |
| rs55985730 | 7q32.1 | G | T | cg10826733 | chr7:128482561 | FLNC | -0.7238 | 0.08698 | 3.51E-16 | Whole blood | European | 837 | Gaunt-T_methQTL-15up_EUR_2016 | 27036880 |
| rs55985730 | 7q32.1 | G | T | cg10826733 | chr7:128482561 | FLNC | -0.6506 | 0.08704 | 2.19E-13 | Whole blood | European | 742 | Gaunt-T_methQTL-FOM_EUR_2016 | 27036880 |

| | | | | | | | | | | | | | | |
|---|---|---|---|---|---|---|---|---|---|---|---|---|---|---|
| rs55985730 | 7q32.1 | G | T | cg25303427 | chr7:128482703 | FLNC | | | 4.12E-41 | Whole blood | European | 3841 | BIOSQTL_methQTL_EUR_2017 | 27918535 |
| rs55985730 | 7q32.1 | G | T | cg10826733 | chr7:128482537 | FLNC | | | 8.15E-72 | Whole blood | European | 3841 | BIOSQTL_methQTL_EUR_2017 | 27918535 |
| rs55985730 | 7q32.1 | G | T | cg27045724 | chr7:128379184 | CALU | | | 3.57E-11 | Whole blood | European | 3841 | BIOSQTL_methQTL_EUR_2017 | 27918535 |
| rs55985730 | 7q32.1 | G | T | cg25303427 | chr7:128482679 | FLNC | -0.5829 | 0.08727 | 4.36E-11 | Whole blood | European | 837 | Gaunt-T_methQTL-15up_EUR_2016 | 27036880 |
| rs2269001 | 7q36.1 | A | G | cg05998486 | chr7:150645068 | KCNH2 | | | 1.38E-12 | Whole blood | European | 3841 | BIOSQTL_methQTL_EUR_2017 | 27918535 |
| rs2269001 | 7q36.1 | A | G | cg05726600 | chr7:150498594 | TMEM176A;TMEM176B | 0.3143 | 0.05181 | 1.98E-09 | Whole blood | European | 834 | Gaunt-T_methQTL-F7_EUR_2016 | 27036880 |
| rs2269001 | 7q36.1 | A | G | cg10636020 | chr7:150656979 | KCNH2 | -0.8257 | 0.1192 | 4.30E-12 | Neutrophils | European | 197 | BLUEPRINT_methQTL_EUR_2016 | 27863251 |
| rs2269001 | 7q36.1 | A | G | cg24576940 | chr7:150648307 | KCNH2 | | | 4.02E-23 | Whole blood | European | 3841 | BIOSQTL_methQTL_EUR_2017 | 27918535 |
| rs2269001 | 7q36.1 | A | G | cg10636020 | chr7:150656979 | KCNH2 | -1.02 | 0.1356 | 5.48E-14 | T cells | European | 132 | BLUEPRINT_methQTL_EUR_2016 | 27863251 |
| rs2269001 | 7q36.1 | A | G | cg07998558 | chr7:150648521 | KCNH2 | | | 8.64E-12 | Whole blood | European | 3841 | BIOSQTL_methQTL_EUR_2017 | 27918535 |
| rs2269001 | 7q36.1 | A | G | cg10636020 | chr7:150656979 | KCNH2 | -0.7634 | 0.04918 | 2.02E-47 | Whole blood | European | 837 | Gaunt-T_methQTL-Antenatal_EUR_2016 | 27036880 |
| rs2269001 | 7q36.1 | A | G | cg10636020 | chr7:150656979 | KCNH2 | -0.5646 | 0.04864 | 7.99E-29 | Cord blood | European | 771 | Gaunt-T_methQTL-Cord_EUR_2016 | 27036880 |
| rs2269001 | 7q36.1 | A | G | cg10636020 | chr7:150656979 | KCNH2 | -0.7022 | 0.04828 | 7.01E-43 | Whole blood | European | 837 | Gaunt-T_methQTL-15up_EUR_2016 | 27036880 |
| rs2269001 | 7q36.1 | A | G | cg01559770 | chr7:150644459 | KCNH2 | 0.2914 | 0.05273 | 4.49E-08 | Whole blood | European | 837 | Gaunt-T_methQTL-Antenatal_EUR_2016 | 27036880 |
| rs2269001 | 7q36.1 | A | G | cg20665259 | chr7:150659402 | KCNH2 | -0.2754 | 0.04848 | 1.90E-08 | Whole blood | European | 837 | Gaunt-T_methQTL-Antenatal_EUR_2016 | 27036880 |
| rs2269001 | 7q36.1 | A | G | cg06221570 | chr7:150558353 | ABP1 | | | 4.54E-20 | Whole blood | European | 3841 | BIOSQTL_methQTL_EUR_2017 | 27918535 |
| rs2269001 | 7q36.1 | A | G | cg08359956 | chr7:150498229 | TMEM176A;TMEM176B | | | 1.59E-19 | Whole blood | European | 3841 | BIOSQTL_methQTL_EUR_2017 | 27918535 |
| rs2269001 | 7q36.1 | A | G | cg05726600 | chr7:150498570 | TMEM176A;TMEM176B | | | 4.59E-35 | Whole blood | European | 3841 | BIOSQTL_methQTL_EUR_2017 | 27918535 |
| rs2269001 | 7q36.1 | A | G | cg10454121 | chr7:150675883 | KCNH2 | | | 2.21E-22 | Whole blood | European | 3841 | BIOSQTL_methQTL_EUR_2017 | 27918535 |
| rs2269001 | 7q36.1 | A | G | cg20665259 | chr7:150659426 | KCNH2 | | | 1.95E-87 | Whole blood | European | 3841 | BIOSQTL_methQTL_EUR_2017 | 27918535 |
| rs2269001 | 7q36.1 | A | G | cg20665259 | chr7:150659402 | KCNH2 | -0.2599 | 0.04715 | 4.89E-08 | Whole blood | European | 742 | Gaunt-T_methQTL-FOM_EUR_2016 | 27036880 |
| rs2269001 | 7q36.1 | A | G | cg01559770 | chr7:150644459 | KCNH2 | 0.3385 | 0.05054 | 4.18E-11 | Whole blood | European | 742 | Gaunt-T_methQTL-FOM_EUR_2016 | 27036880 |
| rs2269001 | 7q36.1 | A | G | cg26385222 | chr7:150497101 | TMEM176A;TMEM176B | | | 9.20E-11 | Whole blood | European | 3841 | BIOSQTL_methQTL_EUR_2017 | 27918535 |
| rs2269001 | 7q36.1 | A | G | cg01559770 | chr7:150644459 | KCNH2 | 0.3886 | 0.04659 | 3.04E-16 | Whole blood | European | 837 | Gaunt-T_methQTL-15up_EUR_2016 | 27036880 |
| rs2269001 | 7q36.1 | A | G | cg01559770 | chr7:150644459 | KCNH2 | 0.3528 | 0.04518 | 1.74E-14 | Whole blood | European | 834 | Gaunt-T_methQTL-F7_EUR_2016 | 27036880 |
| rs2269001 | 7q36.1 | A | G | cg10636020 | chr7:150656979 | KCNH2 | -0.7762 | 0.05099 | 1.03E-45 | Whole blood | European | 742 | Gaunt-T_methQTL-FOM_EUR_2016 | 27036880 |
| rs2269001 | 7q36.1 | A | G | cg01539854 | chr7:150655085 | KCNH2 | | | 1.56E-09 | Whole blood | European | 3841 | BIOSQTL_methQTL_EUR_2017 | 27918535 |
| rs2269001 | 7q36.1 | A | G | cg24830730 | chr7:150653219 | KCNH2 | | | 2.87E-16 | Whole blood | European | 3841 | BIOSQTL_methQTL_EUR_2017 | 27918535 |
| rs2269001 | 7q36.1 | A | G | cg17402325 | chr7:150644842 | KCNH2 | | | 4.24E-17 | Whole blood | European | 3841 | BIOSQTL_methQTL_EUR_2017 | 27918535 |
| rs2269001 | 7q36.1 | A | G | cg03964111 | chr7:150498517 | TMEM176A;TMEM176B | | | 2.86E-23 | Whole blood | European | 3841 | BIOSQTL_methQTL_EUR_2017 | 27918535 |
| rs2269001 | 7q36.1 | A | G | cg10636020 | chr7:150656979 | KCNH2;KCNH2 | -0.8295 | 0.1195 | 3.81E-12 | Monocytes | European | 196 | BLUEPRINT_methQTL_EUR_2016 | 27863251 |

| | | | | | | | | | | | | | | |
|---|---|---|---|---|---|---|---|---|---|---|---|---|---|---|
| rs2269001 | 7q36.1 | A | G | cg10636020 | chr7:150656979 | KCNH2;KCNH2 | -0.8071 | 0.04769 | 1.95E-55 | Whole blood | European | 834 | Gaunt-T_methQTL-F7_EUR_2016 | 27036880 |
| rs2269001 | 7q36.1 | A | G | cg17694719 | chr7:150554891 | ABP1 | | | 3.73E-09 | Whole blood | European | 3841 | BIOSQTL_methQTL_EUR_2017 | 27918535 |
| rs2269001 | 7q36.1 | A | G | cg10409299 | chr7:150498867 | TMEM176A;TMEM176B | | | 2.30E-26 | Whole blood | European | 3841 | BIOSQTL_methQTL_EUR_2017 | 27918535 |
| rs2269001 | 7q36.1 | A | G | cg01559770 | chr7:150644435 | KCNH2 | | | 3.52E-70 | Whole blood | European | 3841 | BIOSQTL_methQTL_EUR_2017 | 27918535 |
| rs2269001 | 7q36.1 | A | G | cg24032393 | chr7:150706331 | NOS3 | | | 1.68E-08 | Whole blood | European | 3841 | BIOSQTL_methQTL_EUR_2017 | 27918535 |
| rs2269001 | 7q36.1 | A | G | cg10636020 | chr7:150657003 | KCNH2 | | | 3.68E-284 | Whole blood | European | 3841 | BIOSQTL_methQTL_EUR_2017 | 27918535 |
| rs2269001 | 7q36.1 | A | G | cg21901847 | chr7:150704976 | NOS3 | | | 1.91E-18 | Whole blood | European | 3841 | BIOSQTL_methQTL_EUR_2017 | 27918535 |
| rs2269001 | 7q36.1 | A | G | cg19752686 | chr7:150811751 | AGAP3 | | | 8.50E-09 | Whole blood | European | 3841 | BIOSQTL_methQTL_EUR_2017 | 27918535 |
| rs2269001 | 7q36.1 | A | G | cg11857646 | chr7:150644607 | KCNH2 | | | 8.51E-22 | Whole blood | European | 3841 | BIOSQTL_methQTL_EUR_2017 | 27918535 |
| rs2269001 | 7q36.1 | A | G | cg14991769 | chr7:150497133 | TMEM176A;TMEM176B | | | 3.93E-08 | Whole blood | European | 3841 | BIOSQTL_methQTL_EUR_2017 | 27918535 |
| rs11782313 | 8q24.3 | C | T | cg10996527 | chr8:141994633 | PTK2 | -0.2517 | 0.03794 | 5.90E-11 | Whole blood | European | 837 | Gaunt-T_methQTL-15up_EUR_2016 | 27036880 |
| rs11782313 | 8q24.3 | C | T | cg07891862 | chr8:141774629 | PTK2 | | | 5.69E-31 | Whole blood | European | 3841 | BIOSQTL_methQTL_EUR_2017 | 27918535 |
| rs11782313 | 8q24.3 | C | T | cg14396066 | chr8:142010253 | PTK2 | 0.3199 | 0.03735 | 6.00E-17 | Whole blood | European | 837 | Gaunt-T_methQTL-Antenatal_EUR_2016 | 27036880 |
| rs11782313 | 8q24.3 | C | T | cg14396066 | chr8:142010253 | PTK2 | 0.3008 | 0.0343 | 9.82E-18 | Whole blood | European | 837 | Gaunt-T_methQTL-15up_EUR_2016 | 27036880 |
| rs11782313 | 8q24.3 | C | T | cg26074100 | chr8:141568712 | EIF2C2 | -0.3794 | 0.04177 | 7.53E-19 | Whole blood | European | 834 | Gaunt-T_methQTL-F7_EUR_2016 | 27036880 |
| rs11782313 | 8q24.3 | C | T | cg26074100 | chr8:141568712 | EIF2C2 | -0.415 | 0.04237 | 1.62E-21 | Whole blood | European | 837 | Gaunt-T_methQTL-15up_EUR_2016 | 27036880 |
| rs11782313 | 8q24.3 | C | T | cg10996527 | chr8:141994633 | PTK2 | -0.2335 | 0.03812 | 1.44E-09 | Cord blood | European | 771 | Gaunt-T_methQTL-Cord_EUR_2016 | 27036880 |
| rs11782313 | 8q24.3 | C | T | cg10996527 | chr8:141994633 | PTK2 | -0.2009 | 0.03565 | 2.40E-08 | Whole blood | European | 834 | Gaunt-T_methQTL-F7_EUR_2016 | 27036880 |
| rs11782313 | 8q24.3 | C | T | cg22822656 | chr8:142015196 | PTK2 | | | 3.61E-258 | Whole blood | European | 3841 | BIOSQTL_methQTL_EUR_2017 | 27918535 |
| rs11782313 | 8q24.3 | C | T | cg22822656 | - | PTK2 | 0.59 | 0.0936 | 2.89E-10 | Monocytes | European | 196 | BLUEPRINT_methQTL_EUR_2016 | 27863251 |
| rs11782313 | 8q24.3 | C | T | cg22822656 | - | | 1.028 | 0.0803 | 1.59E-37 | Neutrophils | European | 197 | BLUEPRINT_methQTL_EUR_2016 | 27863251 |
| rs11782313 | 8q24.3 | C | T | cg14396066 | chr8:142010253 | PTK2 | 0.3181 | 0.0379 | 2.24E-16 | Cord blood | European | 771 | Gaunt-T_methQTL-Cord_EUR_2016 | 27036880 |
| rs11782313 | 8q24.3 | C | T | cg14396066 | chr8:142010253 | PTK2 | 0.2934 | 0.03905 | 1.66E-13 | Whole blood | European | 742 | Gaunt-T_methQTL-FOM_EUR_2016 | 27036880 |
| rs11782313 | 8q24.3 | C | T | cg26291848 | chr8:141608316 | EIF2C2 | 0.6552 | 0.1168 | 2.02E-08 | T cells | European | 132 | BLUEPRINT_methQTL_EUR_2016 | 27863251 |
| rs11782313 | 8q24.3 | C | T | cg25998745 | chr8:142028649 | | | | 3.06E-94 | Whole blood | European | 3841 | BIOSQTL_methQTL_EUR_2017 | 27918535 |
| rs11782313 | 8q24.3 | C | T | cg26074100 | chr8:141568712 | EIF2C2 | -0.4199 | 0.04367 | 9.83E-21 | Whole blood | European | 837 | Gaunt-T_methQTL-Antenatal_EUR_2016 | 27036880 |
| rs11782313 | 8q24.3 | C | T | cg26074100 | chr8:141568712 | EIF2C2 | -0.3594 | 0.04415 | 1.59E-15 | Cord blood | European | 771 | Gaunt-T_methQTL-Cord_EUR_2016 | 27036880 |
| rs11782313 | 8q24.3 | C | T | cg09914555 | chr8:142094765 | | | | 4.05E-16 | Whole blood | European | 3841 | BIOSQTL_methQTL_EUR_2017 | 27918535 |
| rs11782313 | 8q24.3 | C | T | cg08444833 | chr8:141574162 | EIF2C2 | -0.2477 | 0.04467 | 3.95E-08 | Whole blood | European | 837 | Gaunt-T_methQTL-15up_EUR_2016 | 27036880 |
| rs11782313 | 8q24.3 | C | T | cg26074100 | chr8:141568712 | EIF2C2 | -0.3695 | 0.04361 | 1.28E-16 | Whole blood | European | 742 | Gaunt-T_methQTL-FOM_EUR_2016 | 27036880 |
| rs11782313 | 8q24.3 | C | T | cg08444833 | chr8:141574162 | EIF2C2 | -0.2596 | 0.0452 | 1.34E-08 | Cord blood | European | 771 | Gaunt-T_methQTL-Cord_EUR_2016 | 27036880 |

| | | | | | | | | | | | | | | |
|---|---|---|---|---|---|---|---|---|---|---|---|---|---|---|
| rs11782313 | 8q24.3 | C | T | cg26291848 | chr8:141608316 | EIF2C2 | 0.2456 | 0.04054 | 2.08E-09 | Whole blood | European | 837 | Gaunt-T_methQTL-15up_EUR_2016 | 27036880 |
| rs11782313 | 8q24.3 | C | T | cg14396066 | chr8:142010253 | PTK2 | 0.2769 | 0.03374 | 8.48E-16 | Whole blood | European | 834 | Gaunt-T_methQTL-F7_EUR_2016 | 27036880 |
| rs11782313 | 8q24.3 | C | T | cg26291848 | chr8:141608316 | EIF2C2 | 0.2543 | 0.04381 | 9.45E-09 | Cord blood | European | 771 | Gaunt-T_methQTL-Cord_EUR_2016 | 27036880 |
| rs11782313 | 8q24.3 | C | T | cg10996527 | chr8:141994657 | PTK2 | | | 4.01E-14 | Whole blood | European | 3841 | BIOSQTL_methQTL_EUR_2017 | 27918535 |
| rs11782313 | 8q24.3 | C | T | cg14774290 | chr8:141934276 | PTK2 | | | 1.96E-21 | Whole blood | European | 3841 | BIOSQTL_methQTL_EUR_2017 | 27918535 |
| rs4385527 | 9q22.32 | G | A | cg13740339 | chr9:97400733 | FBP1 | | | 7.04E-15 | Whole blood | European | 3841 | BIOSQTL_methQTL_EUR_2017 | 27918535 |
| rs4385527 | 9q22.32 | G | A | cg13792694 | chr9:97865256 | FANCC | | | 4.92E-10 | Whole blood | European | 3841 | BIOSQTL_methQTL_EUR_2017 | 27918535 |
| rs4385527 | 9q22.32 | G | A | cg03456213 | chr9:97767978 | C9orf3 | | | 9.44E-20 | Whole blood | European | 3841 | BIOSQTL_methQTL_EUR_2017 | 27918535 |
| rs4385527 | 9q22.32 | G | A | cg04523069 | chr9:97136363 | HIATL1 | -0.2845 | 0.04321 | 8.09E-11 | Whole blood | European | 834 | Gaunt-T_methQTL-F7_EUR_2016 | 27036880 |
| rs4385527 | 9q22.32 | G | A | cg19305934 | chr9:97811111 | C9orf3 | | | 1.41E-08 | Whole blood | European | 3841 | BIOSQTL_methQTL_EUR_2017 | 27918535 |
| rs635634 | 9q34.2 | T | C | cg11879188 | chr9:136149908 | ABO | 0.7484 | 0.05055 | 8.79E-44 | Whole blood | European | 837 | Gaunt-T_methQTL-Antenatal_EUR_2016 | 27036880 |
| rs635634 | 9q34.2 | T | C | cg18089000 | chr9:136039682 | GBGT1 | | | 1.12E-23 | Whole blood | European | 3841 | BIOSQTL_methQTL_EUR_2017 | 27918535 |
| rs635634 | 9q34.2 | T | C | cg24267699 | chr9:136151359 | ABO | 0.801 | 0.05025 | 2.13E-49 | Whole blood | European | 742 | Gaunt-T_methQTL-FOM_EUR_2016 | 27036880 |
| rs635634 | 9q34.2 | T | C | cg07241568 | chr9:136150823 | ABO | 0.5333 | 0.04416 | 7.60E-31 | Whole blood | European | 837 | Gaunt-T_methQTL-Antenatal_EUR_2016 | 27036880 |
| rs635634 | 9q34.2 | T | C | cg01263574 | chr9:136390837 | TMEM8C | | | 2.31E-08 | Whole blood | European | 3841 | BIOSQTL_methQTL_EUR_2017 | 27918535 |
| rs635634 | 9q34.2 | T | C | cg13465995 | chr9:136203363 | SURF6 | | | 2.14E-09 | Whole blood | European | 3841 | BIOSQTL_methQTL_EUR_2017 | 27918535 |
| rs635634 | 9q34.2 | T | C | cg07241568 | chr9:136150823 | ABO | 0.4795 | 0.04303 | 5.66E-27 | Whole blood | European | 834 | Gaunt-T_methQTL-F7_EUR_2016 | 27036880 |
| rs635634 | 9q34.2 | T | C | cg14440550 | chr9:136131118 | ABO | 0.4023 | 0.05282 | 7.16E-14 | Whole blood | European | 834 | Gaunt-T_methQTL-F7_EUR_2016 | 27036880 |
| rs635634 | 9q34.2 | T | C | cg11879188 | chr9:136149908 | ABO | 0.9268 | 0.1123 | 1.57E-16 | Monocytes | European | 196 | BLUEPRINT_methQTL_EUR_2016 | 27863251 |
| rs635634 | 9q34.2 | T | C | cg13040392 | chr9:136075369 | | | | 1.28E-13 | Whole blood | European | 3841 | BIOSQTL_methQTL_EUR_2017 | 27918535 |
| rs635634 | 9q34.2 | T | C | cg13506600 | chr9:136150361 | ABO | 0.6896 | 0.1175 | 4.39E-09 | Monocytes | European | 196 | BLUEPRINT_methQTL_EUR_2016 | 27863251 |
| rs635634 | 9q34.2 | T | C | cg13506600 | chr9:136150361 | ABO | 0.422 | 0.05363 | 1.11E-14 | Whole blood | European | 834 | Gaunt-T_methQTL-F7_EUR_2016 | 27036880 |
| rs635634 | 9q34.2 | T | C | cg02257517 | chr9:136340286 | SLC2A6 | | | 3.71E-08 | Whole blood | European | 3841 | BIOSQTL_methQTL_EUR_2017 | 27918535 |
| rs635634 | 9q34.2 | T | C | cg13980863 | chr9:136117807 | | | | 3.17E-16 | Whole blood | European | 3841 | BIOSQTL_methQTL_EUR_2017 | 27918535 |
| rs635634 | 9q34.2 | T | C | cg07241568 | chr9:136150823 | ABO | 0.5575 | 0.04478 | 9.28E-33 | Whole blood | European | 837 | Gaunt-T_methQTL-15up_EUR_2016 | 27036880 |
| rs635634 | 9q34.2 | T | C | cg13850847 | chr9:136131715 | ABO | | | 2.85E-164 | Whole blood | European | 3841 | BIOSQTL_methQTL_EUR_2017 | 27918535 |
| rs635634 | 9q34.2 | T | C | cg13531387 | chr9:136078681 | | | | 3.26E-78 | Whole blood | European | 3841 | BIOSQTL_methQTL_EUR_2017 | 27918535 |
| rs635634 | 9q34.2 | T | C | cg24267699 | chr9:136151359 | ABO | 0.9075 | 0.04731 | 3.25E-67 | Whole blood | European | 837 | Gaunt-T_methQTL-Antenatal_EUR_2016 | 27036880 |
| rs635634 | 9q34.2 | T | C | cg24267699 | chr9:136151359 | ABO | 1.187 | 0.1047 | 9.06E-30 | Neutrophils | European | 197 | BLUEPRINT_methQTL_EUR_2016 | 27863251 |
| rs635634 | 9q34.2 | T | C | cg13506600 | chr9:136150361 | ABO | 0.5314 | 0.05619 | 4.11E-20 | Whole blood | European | 742 | Gaunt-T_methQTL-FOM_EUR_2016 | 27036880 |
| rs635634 | 9q34.2 | T | C | cg13506600 | chr9:136150361 | ABO | -0.466 | 0.05333 | 1.48E-17 | Cord blood | European | 771 | Gaunt-T_methQTL-Cord_EUR_2016 | 27036880 |

| | | | | | | | | | | | | | | |
|---|---|---|---|---|---|---|---|---|---|---|---|---|---|---|
| rs635634 | 9q34.2 | T | C | cg14442518 | chr9:136102519 | | | | 1.03E-24 | Whole blood | European | 3841 | BIOSQTL_methQTL_EUR_2017 | 27918535 |
| rs635634 | 9q34.2 | T | C | cg13506600 | chr9:136150361 | ABO | 0.6761 | 0.05071 | 1.34E-36 | Whole blood | European | 837 | Gaunt-T_methQTL-Antenatal_EUR_2016 | 27036880 |
| rs635634 | 9q34.2 | T | C | cg24267699 | chr9:136151359 | ABO | 1.199 | 0.1045 | 1.75E-30 | Monocytes | European | 196 | BLUEPRINT_methQTL_EUR_2016 | 27863251 |
| rs635634 | 9q34.2 | T | C | cg06818865 | chr9:136151958 | ABO | 0.3268 | 0.05181 | 4.78E-10 | Whole blood | European | 837 | Gaunt-T_methQTL-Antenatal_EUR_2016 | 27036880 |
| rs635634 | 9q34.2 | T | C | cg11879188 | chr9:136149908 | ABO | 0.6529 | 0.05086 | 3.36E-34 | Whole blood | European | 742 | Gaunt-T_methQTL-FOM_EUR_2016 | 27036880 |
| rs635634 | 9q34.2 | T | C | cg07241568 | chr9:136150823 | ABO | 0.8078 | 0.1154 | 2.55E-12 | Monocytes | European | 196 | BLUEPRINT_methQTL_EUR_2016 | 27863251 |
| rs635634 | 9q34.2 | T | C | cg13531387 | chr9:136078657 | | -0.3067 | 0.04824 | 3.37E-10 | Whole blood | European | 834 | Gaunt-T_methQTL-F7_EUR_2016 | 27036880 |
| rs635634 | 9q34.2 | T | C | cg13531387 | chr9:136078657 | | -0.3185 | 0.05156 | 1.07E-09 | Whole blood | European | 742 | Gaunt-T_methQTL-FOM_EUR_2016 | 27036880 |
| rs635634 | 9q34.2 | T | C | cg14653977 | chr9:136038716 | GBGT1 | | | 9.45E-14 | Whole blood | European | 3841 | BIOSQTL_methQTL_EUR_2017 | 27918535 |
| rs635634 | 9q34.2 | T | C | cg24267699 | chr9:136151359 | ABO | 0.8585 | 0.0474 | 4.17E-62 | Whole blood | European | 837 | Gaunt-T_methQTL-15up_EUR_2016 | 27036880 |
| rs635634 | 9q34.2 | T | C | cg22535403 | chr9:136150032 | ABO | 1.151 | 0.106 | 1.91E-27 | Neutrophils | European | 197 | BLUEPRINT_methQTL_EUR_2016 | 27863251 |
| rs635634 | 9q34.2 | T | C | cg13620499 | chr9:136353126 | | 0.3346 | 0.05416 | 1.02E-09 | Whole blood | European | 837 | Gaunt-T_methQTL-15up_EUR_2016 | 27036880 |
| rs635634 | 9q34.2 | T | C | cg13506600 | chr9:136150361 | ABO | 0.5464 | 0.0534 | 3.17E-23 | Whole blood | European | 837 | Gaunt-T_methQTL-15up_EUR_2016 | 27036880 |
| rs635634 | 9q34.2 | T | C | cg14440550 | chr9:136131118 | ABO | 0.3718 | 0.05601 | 6.19E-11 | Whole blood | European | 742 | Gaunt-T_methQTL-FOM_EUR_2016 | 27036880 |
| rs635634 | 9q34.2 | T | C | cg11879188 | chr9:136149908 | ABO | 0.9592 | 0.1114 | 7.48E-18 | Neutrophils | European | 197 | BLUEPRINT_methQTL_EUR_2016 | 27863251 |
| rs635634 | 9q34.2 | T | C | cg07241568 | chr9:136150823 | ABO | 0.9026 | 0.1324 | 9.36E-12 | T cells | European | 132 | BLUEPRINT_methQTL_EUR_2016 | 27863251 |
| rs635634 | 9q34.2 | T | C | cg07241568 | chr9:136150823 | ABO | 0.4601 | 0.04666 | 1.25E-21 | Whole blood | European | 742 | Gaunt-T_methQTL-FOM_EUR_2016 | 27036880 |
| rs635634 | 9q34.2 | T | C | cg13850847 | chr9:136131739 | ABO | 0.7553 | 0.1149 | 5.00E-11 | Neutrophils | European | 197 | BLUEPRINT_methQTL_EUR_2016 | 27863251 |
| rs635634 | 9q34.2 | T | C | cg00017475 | chr9:136131683 | ABO | | | 2.37E-11 | Whole blood | European | 3841 | BIOSQTL_methQTL_EUR_2017 | 27918535 |
| rs635634 | 9q34.2 | T | C | cg14296235 | chr9:136182191 | | | | 4.52E-10 | Whole blood | European | 3841 | BIOSQTL_methQTL_EUR_2017 | 27918535 |
| rs635634 | 9q34.2 | T | C | cg14560040 | chr9:136203329 | SURF6 | | | 3.24E-16 | Whole blood | European | 3841 | BIOSQTL_methQTL_EUR_2017 | 27918535 |
| rs635634 | 9q34.2 | T | C | cg14636261 | chr9:136133604 | ABO | | | 3.00E-15 | Whole blood | European | 3841 | BIOSQTL_methQTL_EUR_2017 | 27918535 |
| rs635634 | 9q34.2 | T | C | cg00878953 | chr9:136129899 | | | | 5.14E-33 | Whole blood | European | 3841 | BIOSQTL_methQTL_EUR_2017 | 27918535 |
| rs635634 | 9q34.2 | T | C | cg22535403 | chr9:136150032 | ABO | 0.8639 | 0.04729 | 5.62E-63 | Whole blood | European | 837 | Gaunt-T_methQTL-15up_EUR_2016 | 27036880 |
| rs635634 | 9q34.2 | T | C | cg14440550 | chr9:136131142 | ABO | | | 1.23E-84 | Whole blood | European | 3841 | BIOSQTL_methQTL_EUR_2017 | 27918535 |
| rs635634 | 9q34.2 | T | C | cg22535403 | chr9:136150032 | ABO | 0.8667 | 0.04822 | 1.72E-60 | Whole blood | European | 837 | Gaunt-T_methQTL-Antenatal_EUR_2016 | 27036880 |
| rs635634 | 9q34.2 | T | C | cg24267699 | chr9:136151359 | ABO | 0.8918 | 0.04628 | 1.05E-68 | Whole blood | European | 834 | Gaunt-T_methQTL-F7_EUR_2016 | 27036880 |
| rs635634 | 9q34.2 | T | C | cg07241568 | chr9:136150823 | ABO | 0.319 | 0.04766 | 4.21E-11 | Cord blood | European | 771 | Gaunt-T_methQTL-Cord_EUR_2016 | 27036880 |
| rs635634 | 9q34.2 | T | C | cg13947653 | chr9:136343819 | SLC2A6 | | | 6.09E-13 | Whole blood | European | 3841 | BIOSQTL_methQTL_EUR_2017 | 27918535 |
| rs635634 | 9q34.2 | T | C | cg06818865 | chr9:136151982 | ABO | | | 7.60E-81 | Whole blood | European | 3841 | BIOSQTL_methQTL_EUR_2017 | 27918535 |
| rs635634 | 9q34.2 | T | C | cg13963044 | chr9:136335053 | C9orf7 | | | 5.36E-15 | Whole blood | European | 3841 | BIOSQTL_methQTL_EUR_2017 | 27918535 |

| | | | | | | | | | | | | | | |
|---|---|---|---|---|---|---|---|---|---|---|---|---|---|---|
| rs635634 | 9q34.2 | T | C | cg13660174 | chr9:136238416 | SURF4 | | | 2.99E-16 | Whole blood | European | 3841 | BIOSQTL_methQTL_EUR_2017 | 27918535 |
| rs635634 | 9q34.2 | T | C | cg13850847 | chr9:136131739 | ABO | 0.3966 | 0.05276 | 1.58E-13 | Whole blood | European | 837 | Gaunt-T_methQTL-Antenatal_EUR_2016 | 27036880 |
| rs635634 | 9q34.2 | T | C | cg13966005 | chr9:136128023 | | | | 2.79E-13 | Whole blood | European | 3841 | BIOSQTL_methQTL_EUR_2017 | 27918535 |
| rs635634 | 9q34.2 | T | C | cg22535403 | chr9:136150032 | ABO | 1.142 | 0.1065 | 8.32E-27 | Monocytes | European | 196 | BLUEPRINT_methQTL_EUR_2016 | 27863251 |
| rs635634 | 9q34.2 | T | C | cg13850847 | chr9:136131739 | ABO | 0.414 | 0.05075 | 1.25E-15 | Whole blood | European | 834 | Gaunt-T_methQTL-F7_EUR_2016 | 27036880 |
| rs635634 | 9q34.2 | T | C | cg22535403 | chr9:136150032 | ABO | 0.8983 | 0.04975 | 1.14E-60 | Whole blood | European | 742 | Gaunt-T_methQTL-FOM_EUR_2016 | 27036880 |
| rs635634 | 9q34.2 | T | C | cg13506600 | chr9:136150361 | ABO | 0.8699 | 0.114 | 2.34E-14 | Neutrophils | European | 197 | BLUEPRINT_methQTL_EUR_2016 | 27863251 |
| rs635634 | 9q34.2 | T | C | cg21160290 | chr9:136149941 | ABO | 1.131 | 0.1066 | 2.75E-26 | Monocytes | European | 196 | BLUEPRINT_methQTL_EUR_2016 | 27863251 |
| rs635634 | 9q34.2 | T | C | cg21160290 | chr9:136149941 | ABO | 0.8696 | 0.04977 | 1.67E-57 | Whole blood | European | 742 | Gaunt-T_methQTL-FOM_EUR_2016 | 27036880 |
| rs635634 | 9q34.2 | T | C | cg24267699 | chr9:136151359 | ABO | 0.7124 | 0.04797 | 4.72E-44 | Cord blood | European | 771 | Gaunt-T_methQTL-Cord_EUR_2016 | 27036880 |
| rs635634 | 9q34.2 | T | C | cg24267699 | chr9:136151359 | ABO | 0.9675 | 0.1302 | 1.09E-13 | T cells | European | 132 | BLUEPRINT_methQTL_EUR_2016 | 27863251 |
| rs635634 | 9q34.2 | T | C | cg14440550 | chr9:136131118 | ABO | 0.3974 | 0.05488 | 1.01E-12 | Whole blood | European | 837 | Gaunt-T_methQTL-15up_EUR_2016 | 27036880 |
| rs635634 | 9q34.2 | T | C | cg14271713 | chr9:136153846 | | -0.3363 | 0.05259 | 2.68E-10 | Whole blood | European | 837 | Gaunt-T_methQTL-15up_EUR_2016 | 27036880 |
| rs635634 | 9q34.2 | T | C | cg21160290 | chr9:136149941 | ABO | 0.8537 | 0.1337 | 1.71E-10 | T cells | European | 132 | BLUEPRINT_methQTL_EUR_2016 | 27863251 |
| rs635634 | 9q34.2 | T | C | cg21160290 | chr9:136149941 | ABO | 0.8999 | 0.04845 | 8.39E-64 | Whole blood | European | 837 | Gaunt-T_methQTL-Antenatal_EUR_2016 | 27036880 |
| rs635634 | 9q34.2 | T | C | cg14271713 | chr9:136153822 | | | | 2.15E-94 | Whole blood | European | 3841 | BIOSQTL_methQTL_EUR_2017 | 27918535 |
| rs635634 | 9q34.2 | T | C | cg13850847 | chr9:136131739 | ABO | 0.2996 | 0.05281 | 2.00E-08 | Whole blood | European | 742 | Gaunt-T_methQTL-FOM_EUR_2016 | 27036880 |
| rs635634 | 9q34.2 | T | C | cg00115654 | chr9:136294896 | ADAMTS13 | | | 6.94E-11 | Whole blood | European | 3841 | BIOSQTL_methQTL_EUR_2017 | 27918535 |
| rs635634 | 9q34.2 | T | C | cg21202204 | chr9:136398141 | ADAMTSL2 | | | 1.98E-09 | Whole blood | European | 3841 | BIOSQTL_methQTL_EUR_2017 | 27918535 |
| rs635634 | 9q34.2 | T | C | cg13568213 | chr9:136387259 | TMEM8C | | | 3.01E-12 | Whole blood | European | 3841 | BIOSQTL_methQTL_EUR_2017 | 27918535 |
| rs635634 | 9q34.2 | T | C | cg12020464 | chr9:136131159 | ABO | | | 9.27E-16 | Whole blood | European | 3841 | BIOSQTL_methQTL_EUR_2017 | 27918535 |
| rs635634 | 9q34.2 | T | C | cg21160290 | chr9:136149941 | ABO | 0.8544 | 0.04751 | 2.40E-61 | Whole blood | European | 834 | Gaunt-T_methQTL-F7_EUR_2016 | 27036880 |
| rs635634 | 9q34.2 | T | C | cg13822791 | chr9:136293255 | ADAMTS13 | | | 1.52E-16 | Whole blood | European | 3841 | BIOSQTL_methQTL_EUR_2017 | 27918535 |
| rs635634 | 9q34.2 | T | C | cg21160290 | chr9:136149941 | ABO | 0.8556 | 0.04841 | 1.22E-59 | Whole blood | European | 837 | Gaunt-T_methQTL-15up_EUR_2016 | 27036880 |
| rs635634 | 9q34.2 | T | C | cg18718102 | chr9:136203313 | SURF6 | | | 9.77E-34 | Whole blood | European | 3841 | BIOSQTL_methQTL_EUR_2017 | 27918535 |
| rs635634 | 9q34.2 | T | C | cg13850847 | chr9:136131739 | ABO | 0.6999 | 0.1167 | 2.03E-09 | Monocytes | European | 196 | BLUEPRINT_methQTL_EUR_2016 | 27863251 |
| rs635634 | 9q34.2 | T | C | cg13620499 | chr9:136353102 | | | | 3.56E-83 | Whole blood | European | 3841 | BIOSQTL_methQTL_EUR_2017 | 27918535 |
| rs635634 | 9q34.2 | T | C | cg11879188 | chr9:136149884 | ABO | | | 5.66E-235 | Whole blood | European | 3841 | BIOSQTL_methQTL_EUR_2017 | 27918535 |
| rs635634 | 9q34.2 | T | C | cg22535403 | chr9:136150032 | ABO | 0.8514 | 0.04729 | 1.86E-61 | Whole blood | European | 834 | Gaunt-T_methQTL-F7_EUR_2016 | 27036880 |
| rs635634 | 9q34.2 | T | C | cg14271713 | chr9:136153846 | | -0.3317 | 0.04993 | 5.53E-11 | Whole blood | European | 834 | Gaunt-T_methQTL-F7_EUR_2016 | 27036880 |
| rs635634 | 9q34.2 | T | C | cg07241568 | chr9:136150847 | ABO | | | 2.96E-219 | Whole blood | European | 3841 | BIOSQTL_methQTL_EUR_2017 | 27918535 |

| | | | | | | | | | | | | | | |
|---|---|---|---|---|---|---|---|---|---|---|---|---|---|---|
| rs635634 | 9q34.2 | T | C | cg22535403 | chr9:136150032 | ABO | 0.9124 | 0.1321 | 5.02E-12 | T cells | European | 132 | BLUEPRINT_methQTL_EUR_2016 | 27863251 |
| rs635634 | 9q34.2 | T | C | cg13506600 | chr9:136150385 | ABO | | | 5.09E-215 | Whole blood | European | 3841 | BIOSQTL_methQTL_EUR_2017 | 27918535 |
| rs635634 | 9q34.2 | T | C | cg03474926 | chr9:136023383 | RALGDS | | | 1.27E-24 | Whole blood | European | 3841 | BIOSQTL_methQTL_EUR_2017 | 27918535 |
| rs635634 | 9q34.2 | T | C | cg14271713 | chr9:136153846 | | -0.7308 | 0.1163 | 3.33E-10 | Monocytes | European | 196 | BLUEPRINT_methQTL_EUR_2016 | 27863251 |
| rs635634 | 9q34.2 | T | C | cg13850847 | chr9:136131739 | ABO | 0.3805 | 0.05302 | 1.57E-12 | Whole blood | European | 837 | Gaunt-T_methQTL-15up_EUR_2016 | 27036880 |
| rs635634 | 9q34.2 | T | C | cg11879188 | chr9:136149908 | ABO | 0.5255 | 0.0494 | 7.22E-25 | Whole blood | European | 834 | Gaunt-T_methQTL-F7_EUR_2016 | 27036880 |
| rs635634 | 9q34.2 | T | C | cg11879188 | chr9:136149908 | ABO | 0.5395 | 0.05288 | 4.10E-23 | Whole blood | European | 837 | Gaunt-T_methQTL-15up_EUR_2016 | 27036880 |
| rs635634 | 9q34.2 | T | C | cg13620499 | chr9:136353126 | | 0.3673 | 0.05539 | 6.30E-11 | Cord blood | European | 771 | Gaunt-T_methQTL-Cord_EUR_2016 | 27036880 |
| rs635634 | 9q34.2 | T | C | cg07241568 | chr9:136150823 | ABO | 0.7856 | 0.1152 | 9.02E-12 | Neutrophils | European | 197 | BLUEPRINT_methQTL_EUR_2016 | 27863251 |
| rs635634 | 9q34.2 | T | C | cg06818865 | chr9:136151958 | ABO | 0.7732 | 0.1155 | 2.20E-11 | Neutrophils | European | 197 | BLUEPRINT_methQTL_EUR_2016 | 27863251 |
| rs635634 | 9q34.2 | T | C | cg21160290 | chr9:136149941 | ABO | 1.123 | 0.1066 | 5.98E-26 | Neutrophils | European | 197 | BLUEPRINT_methQTL_EUR_2016 | 27863251 |
| rs635634 | 9q34.2 | T | C | cg13620499 | chr9:136353126 | | 0.317 | 0.05401 | 6.34E-09 | Whole blood | European | 834 | Gaunt-T_methQTL-F7_EUR_2016 | 27036880 |
| rs78249997 | 10q22.2 | C | T | cg07699608 | chr10:75541558 | CHCHD1 | -0.4895 | 0.04534 | 2.12E-25 | Cord blood | European | 771 | Gaunt-T_methQTL-Cord_EUR_2016 | 27036880 |
| rs78249997 | 10q22.2 | C | T | cg23083424 | chr10:75415875 | SYNPO2L | 0.643 | 0.04719 | 2.49E-38 | Whole blood | European | 837 | Gaunt-T_methQTL-15up_EUR_2016 | 27036880 |
| rs78249997 | 10q22.2 | C | T | cg09521743 | - | SYNPO2L | 0.7683 | 0.1286 | 2.34E-09 | Neutrophils | European | 197 | BLUEPRINT_methQTL_EUR_2016 | 27863251 |
| rs78249997 | 10q22.2 | C | T | cg13918328 | chr10:52500089 | ASAH2B | 0.3099 | 0.05604 | 4.28E-08 | Whole blood | European | 834 | Gaunt-T_methQTL-F7_EUR_2016 | 27036880 |
| rs78249997 | 10q22.2 | C | T | cg02286717 | chr10:75415704 | SYNPO2L | 1.284 | 0.1155 | 1.02E-28 | Monocytes | European | 196 | BLUEPRINT_methQTL_EUR_2016 | 27863251 |
| rs78249997 | 10q22.2 | C | T | cg02286717 | chr10:75415704 | SYNPO2L | 1.024 | 0.04473 | 1.80E-90 | Whole blood | European | 837 | Gaunt-T_methQTL-15up_EUR_2016 | 27036880 |
| rs78249997 | 10q22.2 | C | T | cg04320760 | chr10:75533115 | FUT11 | | | 3.07E-42 | Whole blood | European | 3841 | BIOSQTL_methQTL_EUR_2017 | 27918535 |
| rs78249997 | 10q22.2 | C | T | cg04320760 | chr10:75533139 | FUT11 | -0.3615 | 0.05996 | 2.57E-09 | Cord blood | European | 771 | Gaunt-T_methQTL-Cord_EUR_2016 | 27036880 |
| rs78249997 | 10q22.2 | C | T | cg09521743 | - | SYNPO2L | 0.8187 | 0.1281 | 1.64E-10 | Monocytes | European | 196 | BLUEPRINT_methQTL_EUR_2016 | 27863251 |
| rs78249997 | 10q22.2 | C | T | cg17411546 | chr10:75410026 | SYNPO2L | -0.3554 | 0.04445 | 4.30E-15 | Whole blood | European | 834 | Gaunt-T_methQTL-F7_EUR_2016 | 27036880 |
| rs78249997 | 10q22.2 | C | T | cg02286717 | chr10:75415704 | SYNPO2L | 1.325 | 0.1141 | 3.35E-31 | Neutrophils | European | 197 | BLUEPRINT_methQTL_EUR_2016 | 27863251 |
| rs78249997 | 10q22.2 | C | T | cg23083424 | chr10:75415875 | SYNPO2L | 0.8834 | 0.1265 | 2.85E-12 | Neutrophils | European | 197 | BLUEPRINT_methQTL_EUR_2016 | 27863251 |
| rs78249997 | 10q22.2 | C | T | cg19442545 | chr10:75533407 | FUT11 | | | 4.42E-115 | Whole blood | European | 3841 | BIOSQTL_methQTL_EUR_2017 | 27918535 |
| rs78249997 | 10q22.2 | C | T | cg02286717 | chr10:75415704 | SYNPO2L | 1.021 | 0.04442 | 3.44E-89 | Whole blood | European | 837 | Gaunt-T_methQTL-Antenatal_EUR_2016 | 27036880 |
| rs78249997 | 10q22.2 | C | T | cg07699608 | chr10:75541558 | CHCHD1 | -0.4238 | 0.04918 | 3.40E-17 | Whole blood | European | 837 | Gaunt-T_methQTL-15up_EUR_2016 | 27036880 |
| rs78249997 | 10q22.2 | C | T | cg17411546 | chr10:75410026 | SYNPO2L | -0.2601 | 0.04326 | 2.84E-09 | Cord blood | European | 771 | Gaunt-T_methQTL-Cord_EUR_2016 | 27036880 |
| rs78249997 | 10q22.2 | C | T | cg07699608 | chr10:75541558 | CHCHD1 | -0.4472 | 0.04509 | 5.45E-22 | Whole blood | European | 834 | Gaunt-T_methQTL-F7_EUR_2016 | 27036880 |
| rs78249997 | 10q22.2 | C | T | cg13918328 | chr10:52500089 | ASAH2B | 0.3678 | 0.05661 | 1.40E-10 | Whole blood | European | 837 | Gaunt-T_methQTL-15up_EUR_2016 | 27036880 |
| rs78249997 | 10q22.2 | C | T | cg02286717 | chr10:75415704 | SYNPO2L | 1.314 | 0.141 | 1.20E-20 | T cells | European | 132 | BLUEPRINT_methQTL_EUR_2016 | 27863251 |

| | | | | | | | | | | | | | | |
|---|---|---|---|---|---|---|---|---|---|---|---|---|---|---|
| rs78249997 | 10q22.2 | C | T | cg23083424 | chr10:75415875 | SYNPO2L | 0.6744 | 0.05029 | 5.73E-37 | Whole blood | European | 837 | Gaunt-T_methQTL-Antenatal_EUR_2016 | 27036880 |
| rs78249997 | 10q22.2 | C | T | cg13918328 | chr10:52500089 | ASAH2B | 0.4247 | 0.05895 | 1.41E-12 | Whole blood | European | 837 | Gaunt-T_methQTL-Antenatal_EUR_2016 | 27036880 |
| rs78249997 | 10q22.2 | C | T | cg05697785 | chr10:75416784 | SYNPO2L | | | 1.05E-64 | Whole blood | European | 3841 | BIOSQTL_methQTL_EUR_2017 | 27918535 |
| rs78249997 | 10q22.2 | C | T | cg19154600 | chr10:75415868 | SYNPO2L | 0.5594 | 0.05252 | 8.51E-25 | Whole blood | European | 837 | Gaunt-T_methQTL-Antenatal_EUR_2016 | 27036880 |
| rs78249997 | 10q22.2 | C | T | cg15291476 | chr10:88779109 | FAM25A | -0.2648 | 0.0435 | 1.83E-09 | Whole blood | European | 742 | Gaunt-T_methQTL-FOM_EUR_2016 | 27036880 |
| rs78249997 | 10q22.2 | C | T | cg23083424 | chr10:75415851 | SYNPO2L | | | 1.54E-249 | Whole blood | European | 3841 | BIOSQTL_methQTL_EUR_2017 | 27918535 |
| rs78249997 | 10q22.2 | C | T | cg04833713 | chr10:74871078 | NUDT13 | -0.3067 | 0.05484 | 3.03E-08 | Whole blood | European | 834 | Gaunt-T_methQTL-F7_EUR_2016 | 27036880 |
| rs78249997 | 10q22.2 | C | T | cg17411546 | chr10:75410026 | SYNPO2L | -0.2996 | 0.04446 | 2.97E-11 | Whole blood | European | 837 | Gaunt-T_methQTL-15up_EUR_2016 | 27036880 |
| rs78249997 | 10q22.2 | C | T | cg23231163 | chr10:75533350 | FUT11 | -0.3972 | 0.06156 | 1.95E-10 | Cord blood | European | 771 | Gaunt-T_methQTL-Cord_EUR_2016 | 27036880 |
| rs78249997 | 10q22.2 | C | T | cg19154600 | chr10:75415868 | SYNPO2L | 0.8836 | 0.1267 | 3.13E-12 | Neutrophils | European | 197 | BLUEPRINT_methQTL_EUR_2016 | 27863251 |
| rs78249997 | 10q22.2 | C | T | cg19154600 | chr10:75415868 | SYNPO2L | 0.5994 | 0.05102 | 1.34E-29 | Whole blood | European | 834 | Gaunt-T_methQTL-F7_EUR_2016 | 27036880 |
| rs78249997 | 10q22.2 | C | T | cg23083424 | chr10:75415875 | SYNPO2L | 0.5266 | 0.04719 | 7.87E-27 | Whole blood | European | 742 | Gaunt-T_methQTL-FOM_EUR_2016 | 27036880 |
| rs78249997 | 10q22.2 | C | T | cg19154600 | chr10:75415868 | SYNPO2L | 0.4339 | 0.04761 | 7.40E-19 | Whole blood | European | 742 | Gaunt-T_methQTL-FOM_EUR_2016 | 27036880 |
| rs78249997 | 10q22.2 | C | T | cg15838173 | chr10:75533400 | FUT11 | -0.4125 | 0.06078 | 2.36E-11 | Whole blood | European | 742 | Gaunt-T_methQTL-FOM_EUR_2016 | 27036880 |
| rs78249997 | 10q22.2 | C | T | cg15838173 | chr10:75533376 | FUT11 | | | 5.94E-88 | Whole blood | European | 3841 | BIOSQTL_methQTL_EUR_2017 | 27918535 |
| rs78249997 | 10q22.2 | C | T | cg09521743 | chr10:75415776 | SYNPO2L | | | 7.55E-97 | Whole blood | European | 3841 | BIOSQTL_methQTL_EUR_2017 | 27918535 |
| rs78249997 | 10q22.2 | C | T | cg23083424 | chr10:75415875 | SYNPO2L | 1.067 | 0.15 | 1.14E-12 | T cells | European | 132 | BLUEPRINT_methQTL_EUR_2016 | 27863251 |
| rs78249997 | 10q22.2 | C | T | cg23231163 | chr10:75533350 | FUT11 | -0.3679 | 0.05514 | 4.63E-11 | Whole blood | European | 834 | Gaunt-T_methQTL-F7_EUR_2016 | 27036880 |
| rs78249997 | 10q22.2 | C | T | cg04717534 | chr10:75633028 | CAMK2G | | | 5.76E-10 | Whole blood | European | 3841 | BIOSQTL_methQTL_EUR_2017 | 27918535 |
| rs78249997 | 10q22.2 | C | T | cg19154600 | chr10:75415868 | SYNPO2L | 0.4609 | 0.05052 | 5.31E-19 | Whole blood | European | 837 | Gaunt-T_methQTL-15up_EUR_2016 | 27036880 |
| rs78249997 | 10q22.2 | C | T | cg23577033 | chr10:75636885 | | | | 1.26E-11 | Whole blood | European | 3841 | BIOSQTL_methQTL_EUR_2017 | 27918535 |
| rs78249997 | 10q22.2 | C | T | cg02286717 | chr10:75415704 | SYNPO2L | 0.9619 | 0.04451 | 1.12E-80 | Whole blood | European | 742 | Gaunt-T_methQTL-FOM_EUR_2016 | 27036880 |
| rs78249997 | 10q22.2 | C | T | cg19442545 | - | FUT11 | -0.7069 | 0.1294 | 4.69E-08 | Monocytes | European | 196 | BLUEPRINT_methQTL_EUR_2016 | 27863251 |
| rs78249997 | 10q22.2 | C | T | cg23083424 | chr10:75415875 | SYNPO2L | 0.6728 | 0.05042 | 5.97E-37 | Whole blood | European | 834 | Gaunt-T_methQTL-F7_EUR_2016 | 27036880 |
| rs78249997 | 10q22.2 | C | T | cg17411546 | chr10:75410026 | SYNPO2L | -0.2993 | 0.04611 | 1.55E-10 | Whole blood | European | 742 | Gaunt-T_methQTL-FOM_EUR_2016 | 27036880 |
| rs78249997 | 10q22.2 | C | T | cg07512258 | chr10:75619396 | CAMK2G | | | 5.71E-25 | Whole blood | European | 3841 | BIOSQTL_methQTL_EUR_2017 | 27918535 |
| rs78249997 | 10q22.2 | C | T | cg19154600 | chr10:75415868 | SYNPO2L | 0.494 | 0.05446 | 9.71E-19 | Cord blood | European | 771 | Gaunt-T_methQTL-Cord_EUR_2016 | 27036880 |
| rs78249997 | 10q22.2 | C | T | cg07699608 | chr10:75541558 | CHCHD1 | -0.545 | 0.0471 | 1.22E-28 | Whole blood | European | 837 | Gaunt-T_methQTL-Antenatal_EUR_2016 | 27036880 |
| rs78249997 | 10q22.2 | C | T | cg06211744 | chr10:75389209 | | | | 3.84E-09 | Whole blood | European | 3841 | BIOSQTL_methQTL_EUR_2017 | 27918535 |
| rs78249997 | 10q22.2 | C | T | cg15838173 | chr10:75533400 | FUT11 | -0.3993 | 0.06108 | 1.14E-10 | Cord blood | European | 771 | Gaunt-T_methQTL-Cord_EUR_2016 | 27036880 |
| rs78249997 | 10q22.2 | C | T | cg13918328 | chr10:52500089 | ASAH2B | 0.3766 | 0.06115 | 1.18E-09 | Cord blood | European | 771 | Gaunt-T_methQTL-Cord_EUR_2016 | 27036880 |

| | | | | | | | | | | | | | | |
|---|---|---|---|---|---|---|---|---|---|---|---|---|---|---|
| rs78249997 | 10q22.2 | C | T | cg15291476 | chr10:88779109 | FAM25A | -0.2708 | 0.04481 | 2.37E-09 | Whole blood | European | 837 | Gaunt-T_methQTL-Antenatal_EUR_2016 | 27036880 |
| rs78249997 | 10q22.2 | C | T | cg00564723 | chr10:75632090 | CAMK2G | | | 4.09E-09 | Whole blood | European | 3841 | BIOSQTL_methQTL_EUR_2017 | 27918535 |
| rs78249997 | 10q22.2 | C | T | cg07699608 | chr10:75541558 | CHCHD1 | -0.4703 | 0.04715 | 4.52E-22 | Whole blood | European | 742 | Gaunt-T_methQTL-FOM_EUR_2016 | 27036880 |
| rs78249997 | 10q22.2 | C | T | cg23231163 | chr10:75533350 | FUT11 | -0.4377 | 0.05858 | 2.18E-13 | Whole blood | European | 837 | Gaunt-T_methQTL-Antenatal_EUR_2016 | 27036880 |
| rs78249997 | 10q22.2 | C | T | cg25725418 | chr10:75639855 | | | | 3.13E-12 | Whole blood | European | 3841 | BIOSQTL_methQTL_EUR_2017 | 27918535 |
| rs78249997 | 10q22.2 | C | T | cg17411546 | chr10:75410026 | SYNPO2L | -0.3233 | 0.04374 | 3.87E-13 | Whole blood | European | 837 | Gaunt-T_methQTL-Antenatal_EUR_2016 | 27036880 |
| rs78249997 | 10q22.2 | C | T | cg23231163 | chr10:75533350 | FUT11 | -0.3874 | 0.05516 | 4.91E-12 | Whole blood | European | 742 | Gaunt-T_methQTL-FOM_EUR_2016 | 27036880 |
| rs78249997 | 10q22.2 | C | T | cg19154600 | chr10:75415844 | SYNPO2L | | | 3.20E-186 | Whole blood | European | 3841 | BIOSQTL_methQTL_EUR_2017 | 27918535 |
| rs78249997 | 10q22.2 | C | T | cg02286717 | chr10:75415704 | SYNPO2L | 1.026 | 0.04478 | 5.52E-89 | Cord blood | European | 771 | Gaunt-T_methQTL-Cord_EUR_2016 | 27036880 |
| rs78249997 | 10q22.2 | C | T | cg15838173 | chr10:75533400 | FUT11 | -0.5473 | 0.05841 | 8.13E-20 | Whole blood | European | 837 | Gaunt-T_methQTL-Antenatal_EUR_2016 | 27036880 |
| rs78249997 | 10q22.2 | C | T | cg23231163 | chr10:75533326 | FUT11 | | | 2.57E-67 | Whole blood | European | 3841 | BIOSQTL_methQTL_EUR_2017 | 27918535 |
| rs78249997 | 10q22.2 | C | T | cg23083424 | chr10:75415875 | SYNPO2L | 0.4539 | 0.05139 | 6.89E-18 | Cord blood | European | 771 | Gaunt-T_methQTL-Cord_EUR_2016 | 27036880 |
| rs78249997 | 10q22.2 | C | T | cg15838173 | chr10:75533400 | FUT11 | -0.3583 | 0.05556 | 1.91E-10 | Whole blood | European | 834 | Gaunt-T_methQTL-F7_EUR_2016 | 27036880 |
| rs78249997 | 10q22.2 | C | T | cg02286717 | chr10:75415704 | SYNPO2L | 1.046 | 0.04038 | 6.91E-109 | Whole blood | European | 834 | Gaunt-T_methQTL-F7_EUR_2016 | 27036880 |
| rs78249997 | 10q22.2 | C | T | cg12317414 | chr10:75407967 | SYNPO2L | | | 2.89E-16 | Whole blood | European | 3841 | BIOSQTL_methQTL_EUR_2017 | 27918535 |
| rs78249997 | 10q22.2 | C | T | cg18704363 | chr10:75387446 | | | | 1.40E-14 | Whole blood | European | 3841 | BIOSQTL_methQTL_EUR_2017 | 27918535 |
| rs78249997 | 10q22.2 | C | T | cg19154600 | chr10:75415868 | SYNPO2L | 0.8608 | 0.1553 | 2.96E-08 | T cells | European | 132 | BLUEPRINT_methQTL_EUR_2016 | 27863251 |
| rs1898096 | 10q22.3 | G | A | cg05395704 | chr10:77955799 | C10orf11 | | | 2.88E-16 | Whole blood | European | 3841 | BIOSQTL_methQTL_EUR_2017 | 27918535 |
| rs1898096 | 10q22.3 | G | A | cg03458695 | chr10:78163679 | C10orf11 | | | 5.14E-09 | Whole blood | European | 3841 | BIOSQTL_methQTL_EUR_2017 | 27918535 |
| rs11598047 | 10q24.33 | A | G | cg12477923 | chr10:105365624 | SH3PXD2A | | | 1.43E-52 | Whole blood | European | 3841 | BIOSQTL_methQTL_EUR_2017 | 27918535 |
| rs11598047 | 10q24.33 | A | G | cg00453258 | chr10:105219196 | CALHM1 | | | 5.63E-13 | Whole blood | European | 3841 | BIOSQTL_methQTL_EUR_2017 | 27918535 |
| rs11598047 | 10q24.33 | A | G | cg26550162 | chr10:105344774 | NEURL | | | 9.43E-30 | Whole blood | European | 3841 | BIOSQTL_methQTL_EUR_2017 | 27918535 |
| rs11598047 | 10q24.33 | A | G | cg12662887 | chr10:105343920 | NEURL | -0.3593 | 0.05514 | 1.30E-10 | Cord blood | European | 771 | Gaunt-T_methQTL-Cord_EUR_2016 | 27036880 |
| rs11598047 | 10q24.33 | A | G | cg19946982 | chr10:105315418 | NEURL | | | 8.07E-10 | Whole blood | European | 3841 | BIOSQTL_methQTL_EUR_2017 | 27918535 |
| rs11598047 | 10q24.33 | A | G | cg23476898 | chr10:105331410 | NEURL | | | 1.38E-10 | Whole blood | European | 3841 | BIOSQTL_methQTL_EUR_2017 | 27918535 |
| rs11598047 | 10q24.33 | A | G | cg14475478 | chr10:105303742 | NEURL | | | 2.90E-12 | Whole blood | European | 3841 | BIOSQTL_methQTL_EUR_2017 | 27918535 |
| rs11598047 | 10q24.33 | A | G | cg18636003 | chr10:105319021 | NEURL | | | 1.74E-75 | Whole blood | European | 3841 | BIOSQTL_methQTL_EUR_2017 | 27918535 |
| rs11598047 | 10q24.33 | A | G | cg11819799 | chr10:105452315 | SH3PXD2A | | | 2.65E-09 | Whole blood | European | 3841 | BIOSQTL_methQTL_EUR_2017 | 27918535 |
| rs11598047 | 10q24.33 | A | G | cg18636003 | chr10:105319045 | NEURL | -0.3404 | 0.05818 | 7.03E-09 | Whole blood | European | 837 | Gaunt-T_methQTL-15up_EUR_2016 | 27036880 |
| rs11598047 | 10q24.33 | A | G | cg18636003 | chr10:105319045 | NEURL | -0.3398 | 0.05681 | 3.46E-09 | Whole blood | European | 742 | Gaunt-T_methQTL-FOM_EUR_2016 | 27036880 |
| rs11598047 | 10q24.33 | A | G | cg05359750 | chr10:105362008 | SH3PXD2A | | | 3.95E-08 | Whole blood | European | 3841 | BIOSQTL_methQTL_EUR_2017 | 27918535 |

| | | | | | | | | | | | | | | |
|---|---|---|---|---|---|---|---|---|---|---|---|---|---|---|
| rs11598047 | 10q24.33 | A | G | cg07969475 | chr10:105341914 | NEURL | 0.2941 | 0.05062 | 9.34E-09 | Whole blood | European | 742 | Gaunt-T_methQTL-FOM_EUR_2016 | 27036880 |
| rs11598047 | 10q24.33 | A | G | cg03422944 | chr10:105344723 | NEURL | -0.4097 | 0.06211 | 7.48E-11 | Whole blood | European | 834 | Gaunt-T_methQTL-F7_EUR_2016 | 27036880 |
| rs11598047 | 10q24.33 | A | G | cg12662887 | chr10:105343920 | NEURL | -0.334 | 0.05642 | 4.91E-09 | Whole blood | European | 742 | Gaunt-T_methQTL-FOM_EUR_2016 | 27036880 |
| rs11598047 | 10q24.33 | A | G | cg12662887 | chr10:105343920 | NEURL | -0.4631 | 0.05542 | 3.08E-16 | Whole blood | European | 837 | Gaunt-T_methQTL-Antenatal_EUR_2016 | 27036880 |
| rs11598047 | 10q24.33 | A | G | cg19007269 | chr10:105420525 | SH3PXD2A | | | 7.30E-10 | Whole blood | European | 3841 | BIOSQTL_methQTL_EUR_2017 | 27918535 |
| rs11598047 | 10q24.33 | A | G | cg12662887 | chr10:105343920 | NEURL | -0.3671 | 0.05336 | 1.19E-11 | Whole blood | European | 837 | Gaunt-T_methQTL-15up_EUR_2016 | 27036880 |
| rs11598047 | 10q24.33 | A | G | cg10831642 | chr10:105378320 | SH3PXD2A | | | 2.11E-31 | Whole blood | European | 3841 | BIOSQTL_methQTL_EUR_2017 | 27918535 |
| rs11598047 | 10q24.33 | A | G | cg07969475 | chr10:105341914 | NEURL | 0.3015 | 0.05453 | 4.44E-08 | Whole blood | European | 837 | Gaunt-T_methQTL-Antenatal_EUR_2016 | 27036880 |
| rs11598047 | 10q24.33 | A | G | cg04688330 | chr10:105451778 | SH3PXD2A | | | 1.31E-23 | Whole blood | European | 3841 | BIOSQTL_methQTL_EUR_2017 | 27918535 |
| rs11598047 | 10q24.33 | A | G | cg17426192 | chr10:105344771 | NEURL | | | 7.50E-61 | Whole blood | European | 3841 | BIOSQTL_methQTL_EUR_2017 | 27918535 |
| rs11598047 | 10q24.33 | A | G | cg07969475 | chr10:105341914 | NEURL | 0.2971 | 0.04823 | 1.12E-09 | Whole blood | European | 837 | Gaunt-T_methQTL-15up_EUR_2016 | 27036880 |
| rs11598047 | 10q24.33 | A | G | cg12662887 | chr10:105343920 | NEURL | -0.51 | 0.05374 | 2.34E-20 | Whole blood | European | 834 | Gaunt-T_methQTL-F7_EUR_2016 | 27036880 |
| rs11598047 | 10q24.33 | A | G | cg16311536 | chr10:105344150 | NEURL | | | 5.33E-72 | Whole blood | European | 3841 | BIOSQTL_methQTL_EUR_2017 | 27918535 |
| rs11598047 | 10q24.33 | A | G | cg17426192 | chr10:105344747 | NEURL | -0.3555 | 0.06076 | 7.01E-09 | Whole blood | European | 837 | Gaunt-T_methQTL-15up_EUR_2016 | 27036880 |
| rs11598047 | 10q24.33 | A | G | cg00126946 | chr10:105363282 | SH3PXD2A | | | 5.44E-12 | Whole blood | European | 3841 | BIOSQTL_methQTL_EUR_2017 | 27918535 |
| rs11598047 | 10q24.33 | A | G | cg17426192 | chr10:105344747 | NEURL | -0.3759 | 0.0616 | 1.61E-09 | Whole blood | European | 834 | Gaunt-T_methQTL-F7_EUR_2016 | 27036880 |
| rs11598047 | 10q24.33 | A | G | cg12662887 | chr10:105343896 | NEURL | | | 1.77E-137 | Whole blood | European | 3841 | BIOSQTL_methQTL_EUR_2017 | 27918535 |
| rs11598047 | 10q24.33 | A | G | cg20386580 | chr10:105344783 | NEURL | | | 2.26E-37 | Whole blood | European | 3841 | BIOSQTL_methQTL_EUR_2017 | 27918535 |
| rs11598047 | 10q24.33 | A | G | cg03422944 | chr10:105344747 | NEURL | | | 7.28E-68 | Whole blood | European | 3841 | BIOSQTL_methQTL_EUR_2017 | 27918535 |
| rs11598047 | 10q24.33 | A | G | cg07709475 | chr10:105423582 | SH3PXD2A | | | 3.04E-47 | Whole blood | European | 3841 | BIOSQTL_methQTL_EUR_2017 | 27918535 |
| rs11598047 | 10q24.33 | A | G | cg07969475 | chr10:105341890 | NEURL | | | 2.13E-189 | Whole blood | European | 3841 | BIOSQTL_methQTL_EUR_2017 | 27918535 |
| rs74399915 | 11q24.3 | C | T | cg03340701 | chr11:128693825 | | | | 2.93E-15 | Whole blood | European | 3841 | BIOSQTL_methQTL_EUR_2017 | 27918535 |
| rs74399915 | 11q24.3 | C | T | cg10430742 | chr11:128695517 | | | | 1.06E-12 | Whole blood | European | 3841 | BIOSQTL_methQTL_EUR_2017 | 27918535 |
| rs74399915 | 11q24.3 | C | T | cg07690882 | chr11:128694279 | | | | 2.14E-34 | Whole blood | European | 3841 | BIOSQTL_methQTL_EUR_2017 | 27918535 |
| rs74399915 | 11q24.3 | C | T | cg20933297 | chr11:128694364 | | | | 4.12E-36 | Whole blood | European | 3841 | BIOSQTL_methQTL_EUR_2017 | 27918535 |
| rs74399915 | 11q24.3 | C | T | cg09813060 | chr11:128694269 | | | | 6.31E-34 | Whole blood | European | 3841 | BIOSQTL_methQTL_EUR_2017 | 27918535 |
| rs74399915 | 11q24.3 | C | T | cg13019868 | chr11:128694077 | | | | 3.97E-30 | Whole blood | European | 3841 | BIOSQTL_methQTL_EUR_2017 | 27918535 |
| rs74399915 | 11q24.3 | C | T | cg04452906 | chr11:128760633 | KCNJ5 | | | 1.49E-12 | Whole blood | European | 3841 | BIOSQTL_methQTL_EUR_2017 | 27918535 |
| rs74399915 | 11q24.3 | C | T | cg15509024 | chr11:128693937 | | | | 7.11E-25 | Whole blood | European | 3841 | BIOSQTL_methQTL_EUR_2017 | 27918535 |
| rs74399915 | 11q24.3 | C | T | cg09318763 | chr11:128798541 | | | | 4.39E-14 | Whole blood | European | 3841 | BIOSQTL_methQTL_EUR_2017 | 27918535 |
| rs74399915 | 11q24.3 | C | T | cg20128672 | chr11:128774625 | C11orf45;KCNJ5 | | | 8.97E-10 | Whole blood | European | 3841 | BIOSQTL_methQTL_EUR_2017 | 27918535 |

| | | | | | | | | | | | | | | |
|---|---|---|---|---|---|---|---|---|---|---|---|---|---|---|
| rs74399915 | 11q24.3 | C | T | cg22755534 | chr11:128796466 | | | | 3.91E-10 | Whole blood | European | 3841 | BIOSQTL_methQTL_EUR_2017 | 27918535 |
| rs74399915 | 11q24.3 | C | T | cg16777618 | chr11:128694160 | | | | 5.18E-33 | Whole blood | European | 3841 | BIOSQTL_methQTL_EUR_2017 | 27918535 |
| rs74399915 | 11q24.3 | C | T | cg05078611 | chr11:128781358 | KCNJ5 | | | 6.72E-56 | Whole blood | European | 3841 | BIOSQTL_methQTL_EUR_2017 | 27918535 |
| rs74399915 | 11q24.3 | C | T | cg09971562 | chr11:128694655 | | | | 1.16E-13 | Whole blood | European | 3841 | BIOSQTL_methQTL_EUR_2017 | 27918535 |
| rs74399915 | 11q24.3 | C | T | cg17297696 | chr11:128693701 | | | | 1.56E-12 | Whole blood | European | 3841 | BIOSQTL_methQTL_EUR_2017 | 27918535 |
| rs883079 | 12q24.21 | T | C | cg10233830 | chr12:114701389 | | | | 7.18E-56 | Whole blood | European | 3841 | BIOSQTL_methQTL_EUR_2017 | 27918535 |
| rs883079 | 12q24.21 | T | C | cg10233830 | chr12:114701413 | | 0.3524 | 0.05012 | 4.56E-12 | Cord blood | European | 771 | Gaunt-T_methQTL-Cord_EUR_2016 | 27036880 |
| rs883079 | 12q24.21 | T | C | cg10233830 | chr12:114701413 | | 0.3591 | 0.04747 | 1.03E-13 | Whole blood | European | 837 | Gaunt-T_methQTL-15up_EUR_2016 | 27036880 |
| rs883079 | 12q24.21 | T | C | cg23530906 | chr12:114708279 | | | | 7.16E-20 | Whole blood | European | 3841 | BIOSQTL_methQTL_EUR_2017 | 27918535 |
| rs883079 | 12q24.21 | T | C | cg10233830 | chr12:114701413 | | 0.388 | 0.04667 | 3.76E-16 | Whole blood | European | 834 | Gaunt-T_methQTL-F7_EUR_2016 | 27036880 |
| rs883079 | 12q24.21 | T | C | cg10233830 | chr12:114701413 | | 0.3952 | 0.04802 | 8.09E-16 | Whole blood | European | 837 | Gaunt-T_methQTL-Antenatal_EUR_2016 | 27036880 |
| rs883079 | 12q24.21 | T | C | cg04672622 | chr12:114724281 | | | | 7.92E-11 | Whole blood | European | 3841 | BIOSQTL_methQTL_EUR_2017 | 27918535 |
| rs883079 | 12q24.21 | T | C | cg10233830 | chr12:114701413 | | 0.3671 | 0.04964 | 3.84E-13 | Whole blood | European | 742 | Gaunt-T_methQTL-FOM_EUR_2016 | 27036880 |
| rs883079 | 12q24.21 | T | C | cg02011607 | chr12:114792569 | TBX5 | | | 7.86E-24 | Whole blood | European | 3841 | BIOSQTL_methQTL_EUR_2017 | 27918535 |
| rs883079 | 12q24.21 | T | C | cg14126466 | chr12:114830039 | TBX5 | | | 4.57E-09 | Whole blood | European | 3841 | BIOSQTL_methQTL_EUR_2017 | 27918535 |
| rs11057583 | 12q24.31 | A | G | cg19781462 | chr12:124744261 | | 0.7428 | 0.07438 | 3.74E-22 | Whole blood | European | 837 | Gaunt-T_methQTL-Antenatal_EUR_2016 | 27036880 |
| rs11057583 | 12q24.31 | A | G | cg07227671 | chr12:124780924 | FAM101A | -0.4358 | 0.06886 | 4.01E-10 | Whole blood | European | 834 | Gaunt-T_methQTL-F7_EUR_2016 | 27036880 |
| rs11057583 | 12q24.31 | A | G | cg19781462 | chr12:124744261 | | 0.5871 | 0.06771 | 2.19E-17 | Whole blood | European | 837 | Gaunt-T_methQTL-15up_EUR_2016 | 27036880 |
| rs11057583 | 12q24.31 | A | G | cg07227671 | chr12:124780900 | FAM101A | | | 1.07E-37 | Whole blood | European | 3841 | BIOSQTL_methQTL_EUR_2017 | 27918535 |
| rs11057583 | 12q24.31 | A | G | cg19781462 | chr12:124744261 | | 0.5631 | 0.07048 | 4.97E-15 | Cord blood | European | 771 | Gaunt-T_methQTL-Cord_EUR_2016 | 27036880 |
| rs11057583 | 12q24.31 | A | G | cg27504292 | chr12:124779508 | FAM101A | | | 3.34E-19 | Whole blood | European | 3841 | BIOSQTL_methQTL_EUR_2017 | 27918535 |
| rs11057583 | 12q24.31 | A | G | cg25098479 | chr12:124763800 | | 0.6198 | 0.06722 | 3.02E-19 | Whole blood | European | 742 | Gaunt-T_methQTL-FOM_EUR_2016 | 27036880 |
| rs11057583 | 12q24.31 | A | G | cg19781462 | chr12:124744261 | | 0.5397 | 0.07382 | 6.90E-13 | Whole blood | European | 742 | Gaunt-T_methQTL-FOM_EUR_2016 | 27036880 |
| rs11057583 | 12q24.31 | A | G | cg19781462 | chr12:124744237 | | | | 2.34E-89 | Whole blood | European | 3841 | BIOSQTL_methQTL_EUR_2017 | 27918535 |
| rs11057583 | 12q24.31 | A | G | cg18049099 | chr12:124832532 | NCOR2 | | | 3.36E-08 | Whole blood | European | 3841 | BIOSQTL_methQTL_EUR_2017 | 27918535 |
| rs11057583 | 12q24.31 | A | G | cg25098479 | chr12:124763800 | | 0.5316 | 0.06387 | 3.94E-16 | Whole blood | European | 837 | Gaunt-T_methQTL-Antenatal_EUR_2016 | 27036880 |
| rs11057583 | 12q24.31 | A | G | cg25098479 | chr12:124763800 | | 0.6415 | 0.05809 | 1.47E-26 | Whole blood | European | 837 | Gaunt-T_methQTL-15up_EUR_2016 | 27036880 |
| rs11057583 | 12q24.31 | A | G | cg25098479 | chr12:124763800 | | 0.5485 | 0.06319 | 2.34E-17 | Cord blood | European | 771 | Gaunt-T_methQTL-Cord_EUR_2016 | 27036880 |
| rs11057583 | 12q24.31 | A | G | cg25098479 | chr12:124763800 | | 0.5605 | 0.05846 | 1.00E-20 | Whole blood | European | 834 | Gaunt-T_methQTL-F7_EUR_2016 | 27036880 |
| rs11057583 | 12q24.31 | A | G | cg19781462 | chr12:124744261 | | 0.6508 | 0.067 | 3.33E-21 | Whole blood | European | 834 | Gaunt-T_methQTL-F7_EUR_2016 | 27036880 |
| rs11057583 | 12q24.31 | A | G | cg25463779 | chr12:124773260 | FAM101A | | | 2.51E-22 | Whole blood | European | 3841 | BIOSQTL_methQTL_EUR_2017 | 27918535 |

| | | | | | | | | | | | | | | |
|---|---|---|---|---|---|---|---|---|---|---|---|---|---|---|
| rs11057583 | 12q24.31 | A | G | cg26814987 | chr12:124773692 | FAM101A | | | 4.71E-50 | Whole blood | European | 3841 | BIOSQTL_methQTL_EUR_2017 | 27918535 |
| rs11057583 | 12q24.31 | A | G | cg04257064 | chr12:124835415 | NCOR2 | | | 3.86E-26 | Whole blood | European | 3841 | BIOSQTL_methQTL_EUR_2017 | 27918535 |
| rs11057583 | 12q24.31 | A | G | cg25098479 | chr12:124763824 | | | | 2.50E-157 | Whole blood | European | 3841 | BIOSQTL_methQTL_EUR_2017 | 27918535 |
| rs2738413 | 14q23.2 | A | G | cg00851532 | chr14:64680245 | SYNE2 | 0.2094 | 0.03477 | 2.59E-09 | Whole blood | European | 837 | Gaunt-T_methQTL-15up_EUR_2016 | 27036880 |
| rs2738413 | 14q23.2 | A | G | cg23250157 | chr14:64679961 | SYNE2 | -0.2341 | 0.04235 | 4.48E-08 | Cord blood | European | 771 | Gaunt-T_methQTL-Cord_EUR_2016 | 27036880 |
| rs2738413 | 14q23.2 | A | G | cg23250157 | chr14:64679985 | SYNE2 | | | 2.61E-180 | Whole blood | European | 3841 | BIOSQTL_methQTL_EUR_2017 | 27918535 |
| rs2738413 | 14q23.2 | A | G | cg09993711 | chr14:64746129 | ESR2 | 0.2261 | 0.03805 | 4.36E-09 | Whole blood | European | 742 | Gaunt-T_methQTL-FOM_EUR_2016 | 27036880 |
| rs2738413 | 14q23.2 | A | G | cg00851532 | chr14:64680245 | SYNE2 | 0.1809 | 0.03202 | 2.23E-08 | Whole blood | European | 834 | Gaunt-T_methQTL-F7_EUR_2016 | 27036880 |
| rs2738413 | 14q23.2 | A | G | cg21029932 | chr14:64805413 | ESR2 | | | 7.23E-14 | Whole blood | European | 3841 | BIOSQTL_methQTL_EUR_2017 | 27918535 |
| rs2738413 | 14q23.2 | A | G | cg23250157 | chr14:64679961 | SYNE2 | -0.2388 | 0.0391 | 1.60E-09 | Whole blood | European | 837 | Gaunt-T_methQTL-Antenatal_EUR_2016 | 27036880 |
| rs2738413 | 14q23.2 | A | G | cg00851532 | chr14:64680245 | SYNE2 | 0.2004 | 0.03639 | 4.98E-08 | Whole blood | European | 837 | Gaunt-T_methQTL-Antenatal_EUR_2016 | 27036880 |
| rs2738413 | 14q23.2 | A | G | cg09993711 | chr14:64746153 | ESR2 | | | 1.51E-12 | Whole blood | European | 3841 | BIOSQTL_methQTL_EUR_2017 | 27918535 |
| rs2738413 | 14q23.2 | A | G | cg00851532 | chr14:64680245 | SYNE2 | 0.1902 | 0.03323 | 1.50E-08 | Whole blood | European | 742 | Gaunt-T_methQTL-FOM_EUR_2016 | 27036880 |
| rs2738413 | 14q23.2 | A | G | cg23284245 | chr14:64757679 | ESR2 | 0.2447 | 0.03481 | 4.59E-12 | Whole blood | European | 837 | Gaunt-T_methQTL-Antenatal_EUR_2016 | 27036880 |
| rs2738413 | 14q23.2 | A | G | cg21174375 | - | SYNE2 | -0.5507 | 0.096 | 9.83E-09 | Monocytes | European | 196 | BLUEPRINT_methQTL_EUR_2016 | 27863251 |
| rs2738413 | 14q23.2 | A | G | cg19583967 | chr14:64805968 | ESR2 | | | 7.85E-13 | Whole blood | European | 3841 | BIOSQTL_methQTL_EUR_2017 | 27918535 |
| rs2738413 | 14q23.2 | A | G | cg09268963 | chr14:64692326 | SYNE2 | | | 7.92E-13 | Whole blood | European | 3841 | BIOSQTL_methQTL_EUR_2017 | 27918535 |
| rs2738413 | 14q23.2 | A | G | cg20019356 | chr14:64764192 | ESR2 | | | 1.31E-24 | Whole blood | European | 3841 | BIOSQTL_methQTL_EUR_2017 | 27918535 |
| rs2738413 | 14q23.2 | A | G | cg23284245 | chr14:64757703 | ESR2 | | | 6.86E-116 | Whole blood | European | 3841 | BIOSQTL_methQTL_EUR_2017 | 27918535 |
| rs2738413 | 14q23.2 | A | G | cg18371424 | chr14:64654283 | SYNE2 | | | 4.10E-31 | Whole blood | European | 3841 | BIOSQTL_methQTL_EUR_2017 | 27918535 |
| rs2738413 | 14q23.2 | A | G | cg10018167 | chr14:64663659 | SYNE2 | | | 8.83E-57 | Whole blood | European | 3841 | BIOSQTL_methQTL_EUR_2017 | 27918535 |
| rs2738413 | 14q23.2 | A | G | cg23284245 | chr14:64757679 | ESR2 | 0.2467 | 0.03399 | 9.91E-13 | Whole blood | European | 742 | Gaunt-T_methQTL-FOM_EUR_2016 | 27036880 |
| rs2738413 | 14q23.2 | A | G | cg21174375 | chr14:64681201 | SYNE2 | | | 5.62E-85 | Whole blood | European | 3841 | BIOSQTL_methQTL_EUR_2017 | 27918535 |
| rs7172038 | 15q24.1 | G | T | cg10576051 | chr15:73661652 | HCN4 | -0.3818 | 0.05952 | 2.49E-10 | Whole blood | European | 837 | Gaunt-T_methQTL-Antenatal_EUR_2016 | 27036880 |
| rs7172038 | 15q24.1 | G | T | cg06071033 | chr15:73662282 | HCN4 | | | 9.18E-12 | Whole blood | European | 3841 | BIOSQTL_methQTL_EUR_2017 | 27918535 |
| rs7172038 | 15q24.1 | G | T | cg01796676 | chr15:73680308 | | | | 1.25E-47 | Whole blood | European | 3841 | BIOSQTL_methQTL_EUR_2017 | 27918535 |
| rs7172038 | 15q24.1 | G | T | cg08318283 | chr15:73611226 | | | | 3.42E-26 | Whole blood | European | 3841 | BIOSQTL_methQTL_EUR_2017 | 27918535 |
| rs7172038 | 15q24.1 | G | T | cg19184771 | chr15:73664342 | | | | 9.40E-16 | Whole blood | European | 3841 | BIOSQTL_methQTL_EUR_2017 | 27918535 |
| rs7172038 | 15q24.1 | G | T | cg06757333 | chr15:73655217 | HCN4 | -0.5128 | 0.04598 | 5.10E-27 | Whole blood | European | 837 | Gaunt-T_methQTL-15up_EUR_2016 | 27036880 |
| rs7172038 | 15q24.1 | G | T | cg10576051 | chr15:73661652 | HCN4 | -0.3193 | 0.05591 | 1.56E-08 | Whole blood | European | 834 | Gaunt-T_methQTL-F7_EUR_2016 | 27036880 |
| rs7172038 | 15q24.1 | G | T | cg11288274 | chr15:73646400 | HCN4 | | | 1.38E-18 | Whole blood | European | 3841 | BIOSQTL_methQTL_EUR_2017 | 27918535 |

| | | | | | | | | | | | | | | |
|---|---|---|---|---|---|---|---|---|---|---|---|---|---|---|
| rs7172038 | 15q24.1 | G | T | cg01211349 | chr15:73661745 | HCN4 | | | 7.41E-09 | Whole blood | European | 3841 | BIOSQTL_methQTL_EUR_2017 | 27918535 |
| rs7172038 | 15q24.1 | G | T | cg06757333 | chr15:73655217 | HCN4 | -0.5439 | 0.04899 | 1.15E-26 | Cord blood | European | 771 | Gaunt-T_methQTL-Cord_EUR_2016 | 27036880 |
| rs7172038 | 15q24.1 | G | T | cg06757333 | chr15:73655193 | HCN4 | | | 2.93E-103 | Whole blood | European | 3841 | BIOSQTL_methQTL_EUR_2017 | 27918535 |
| rs7172038 | 15q24.1 | G | T | cg06757333 | chr15:73655217 | HCN4 | -0.4718 | 0.04512 | 3.97E-24 | Whole blood | European | 834 | Gaunt-T_methQTL-F7_EUR_2016 | 27036880 |
| rs7172038 | 15q24.1 | G | T | cg06757333 | chr15:73655217 | HCN4 | -0.4695 | 0.04943 | 2.70E-20 | Whole blood | European | 837 | Gaunt-T_methQTL-Antenatal_EUR_2016 | 27036880 |
| rs7172038 | 15q24.1 | G | T | cg06757333 | chr15:73655217 | HCN4 | -0.4074 | 0.0502 | 2.00E-15 | Whole blood | European | 742 | Gaunt-T_methQTL-FOM_EUR_2016 | 27036880 |
| rs12908004 | 15q25.1 | A | G | cg18254123 | chr15:80544685 | | | | 2.22E-10 | Whole blood | European | 3841 | BIOSQTL_methQTL_EUR_2017 | 27918535 |
| rs12908004 | 15q25.1 | A | G | cg16887183 | chr15:80658073 | | | | 2.67E-18 | Whole blood | European | 3841 | BIOSQTL_methQTL_EUR_2017 | 27918535 |
| rs12908004 | 15q25.1 | A | G | cg05018460 | chr15:80688054 | | | | 3.86E-14 | Whole blood | European | 3841 | BIOSQTL_methQTL_EUR_2017 | 27918535 |
| rs12908004 | 15q25.1 | A | G | cg27663476 | chr15:80444432 | FAH | | | 4.63E-08 | Whole blood | European | 3841 | BIOSQTL_methQTL_EUR_2017 | 27918535 |
| rs12908004 | 15q25.1 | A | G | cg21603534 | chr15:80694026 | | | | 2.78E-08 | Whole blood | European | 3841 | BIOSQTL_methQTL_EUR_2017 | 27918535 |
| rs6598541 | 15q26.3 | A | G | cg03437748 | chr15:99193271 | IGF1R | | | 5.85E-19 | Whole blood | European | 3841 | BIOSQTL_methQTL_EUR_2017 | 27918535 |
| rs6598541 | 15q26.3 | A | G | cg27438980 | chr15:99195167 | IGF1R | | | 9.12E-09 | Whole blood | European | 3841 | BIOSQTL_methQTL_EUR_2017 | 27918535 |
| rs6598541 | 15q26.3 | A | G | cg05169988 | chr15:99204912 | IGF1R | -0.2083 | 0.03731 | 3.27E-08 | Whole blood | European | 837 | Gaunt-T_methQTL-Antenatal_EUR_2016 | 27036880 |
| rs6598541 | 15q26.3 | A | G | cg05169988 | chr15:99204936 | IGF1R | | | 8.21E-36 | Whole blood | European | 3841 | BIOSQTL_methQTL_EUR_2017 | 27918535 |
| rs6598541 | 15q26.3 | A | G | cg13297560 | chr15:99320030 | IGF1R | | | 1.03E-73 | Whole blood | European | 3841 | BIOSQTL_methQTL_EUR_2017 | 27918535 |
| rs2106261 | 16q22.3 | T | C | cg15712310 | chr16:73100814 | | | | 1.84E-08 | Whole blood | European | 3841 | BIOSQTL_methQTL_EUR_2017 | 27918535 |
| rs2106261 | 16q22.3 | T | C | cg09780180 | chr16:73097879 | | | | 2.54E-13 | Whole blood | European | 3841 | BIOSQTL_methQTL_EUR_2017 | 27918535 |
| rs2106261 | 16q22.3 | T | C | cg01744822 | chr16:73100510 | | -0.3449 | 0.0613 | 2.51E-08 | Whole blood | European | 834 | Gaunt-T_methQTL-F7_EUR_2016 | 27036880 |
| rs2106261 | 16q22.3 | T | C | cg03805182 | chr16:73078047 | ZFHX3 | | | 3.63E-12 | Whole blood | European | 3841 | BIOSQTL_methQTL_EUR_2017 | 27918535 |
| rs2106261 | 16q22.3 | T | C | cg06618356 | chr16:73097388 | | | | 4.97E-35 | Whole blood | European | 3841 | BIOSQTL_methQTL_EUR_2017 | 27918535 |
| rs2106261 | 16q22.3 | T | C | cg01744822 | chr16:73100510 | | -0.3386 | 0.06066 | 3.24E-08 | Whole blood | European | 837 | Gaunt-T_methQTL-15up_EUR_2016 | 27036880 |
| rs2106261 | 16q22.3 | T | C | cg01744822 | chr16:73100534 | | | | 1.16E-37 | Whole blood | European | 3841 | BIOSQTL_methQTL_EUR_2017 | 27918535 |
| rs2106261 | 16q22.3 | T | C | cg03463523 | chr16:73099893 | | | | 1.11E-15 | Whole blood | European | 3841 | BIOSQTL_methQTL_EUR_2017 | 27918535 |
| rs2106261 | 16q22.3 | T | C | cg06984903 | chr16:73100450 | | | | 2.92E-14 | Whole blood | European | 3841 | BIOSQTL_methQTL_EUR_2017 | 27918535 |
| rs2106261 | 16q22.3 | T | C | cg16791832 | chr16:73102267 | | | | 3.31E-117 | Whole blood | European | 3841 | BIOSQTL_methQTL_EUR_2017 | 27918535 |
| rs116600817 | 17p13.1 | A | G | cg00004421 | chr17:7475079 | EIF4A1;SENP3 | -0.2896 | 0.04103 | 3.79E-12 | Cord blood | European | 771 | Gaunt-T_methQTL-Cord_EUR_2016 | 27036880 |
| rs116600817 | 17p13.1 | A | G | cg00004421 | chr17:7475079 | EIF4A1;SENP3 | -0.2769 | 0.04284 | 1.74E-10 | Whole blood | European | 837 | Gaunt-T_methQTL-15up_EUR_2016 | 27036880 |
| rs116600817 | 17p13.1 | A | G | cg00004421 | chr17:7475079 | EIF4A1;SENP3 | -0.3276 | 0.04544 | 1.39E-12 | Whole blood | European | 742 | Gaunt-T_methQTL-FOM_EUR_2016 | 27036880 |
| rs116600817 | 17p13.1 | A | G | cg00004421 | chr17:7475079 | EIF4A1;SENP3 | -0.2619 | 0.04037 | 1.51E-10 | Whole blood | European | 834 | Gaunt-T_methQTL-F7_EUR_2016 | 27036880 |
| rs116600817 | 17p13.1 | A | G | cg01557754 | chr17:7342661 | FGF11 | 0.2847 | 0.04081 | 6.18E-12 | Whole blood | European | 834 | Gaunt-T_methQTL-F7_EUR_2016 | 27036880 |

| | | | | | | | | | | | | | | |
|---|---|---|---|---|---|---|---|---|---|---|---|---|---|---|
| rs116600817 | 17p13.1 | A | G | cg00031162 | chr17:7453353 | TNFSF12;TNFSF13 | | | 1.56E-08 | Whole blood | European | 3841 | BIOSQTL_methQTL_EUR_2017 | 27918535 |
| rs116600817 | 17p13.1 | A | G | cg00053373 | chr17:7554851 | ATP1B2 | | | 5.34E-09 | Whole blood | European | 3841 | BIOSQTL_methQTL_EUR_2017 | 27918535 |
| rs116600817 | 17p13.1 | A | G | cg17892169 | chr17:7452644 | TNFSF12;TNFSF13 | 0.7579 | 0.1041 | 3.28E-13 | Monocytes | European | 196 | BLUEPRINT_methQTL_EUR_2016 | 27863251 |
| rs116600817 | 17p13.1 | A | G | cg17892169 | chr17:7452644 | TNFSF12;TNFSF13 | 0.6807 | 0.1058 | 1.22E-10 | Neutrophils | European | 197 | BLUEPRINT_methQTL_EUR_2016 | 27863251 |
| rs116600817 | 17p13.1 | A | G | cg17892169 | chr17:7452644 | TNFSF12;TNFSF13 | 0.3963 | 0.03547 | 4.11E-27 | Whole blood | European | 837 | Gaunt-T_methQTL-15up_EUR_2016 | 27036880 |
| rs116600817 | 17p13.1 | A | G | cg17892169 | chr17:7452644 | TNFSF12;TNFSF13 | 0.4186 | 0.04225 | 7.39E-22 | Cord blood | European | 771 | Gaunt-T_methQTL-Cord_EUR_2016 | 27036880 |
| rs116600817 | 17p13.1 | A | G | cg03461559 | chr17:7318896 | NLGN2 | | | 1.56E-30 | Whole blood | European | 3841 | BIOSQTL_methQTL_EUR_2017 | 27918535 |
| rs116600817 | 17p13.1 | A | G | cg17892169 | chr17:7452644 | TNFSF12;TNFSF13 | 0.3887 | 0.03696 | 2.28E-24 | Whole blood | European | 834 | Gaunt-T_methQTL-F7_EUR_2016 | 27036880 |
| rs116600817 | 17p13.1 | A | G | cg17892169 | chr17:7452644 | TNFSF12;TNFSF13 | 0.5438 | 0.04225 | 1.92E-34 | Whole blood | European | 837 | Gaunt-T_methQTL-Antenatal_EUR_2016 | 27036880 |
| rs116600817 | 17p13.1 | A | G | cg10509001 | chr17:7553848 | ATP1B2 | | | 4.97E-30 | Whole blood | European | 3841 | BIOSQTL_methQTL_EUR_2017 | 27918535 |
| rs116600817 | 17p13.1 | A | G | cg22637538 | chr17:7348303 | CHRNB1 | | | 2.39E-18 | Whole blood | European | 3841 | BIOSQTL_methQTL_EUR_2017 | 27918535 |
| rs116600817 | 17p13.1 | A | G | cg03666441 | chr17:7254695 | KCTD11;ACAP1 | | | 3.77E-15 | Whole blood | European | 3841 | BIOSQTL_methQTL_EUR_2017 | 27918535 |
| rs116600817 | 17p13.1 | A | G | cg02550691 | chr17:7255712 | KCTD11 | | | 1.86E-16 | Whole blood | European | 3841 | BIOSQTL_methQTL_EUR_2017 | 27918535 |
| rs116600817 | 17p13.1 | A | G | cg05514680 | chr17:7461532 | TNFSF12;TNFSF13 | | | 1.48E-10 | Whole blood | European | 3841 | BIOSQTL_methQTL_EUR_2017 | 27918535 |
| rs116600817 | 17p13.1 | A | G | cg08090557 | chr17:7464064 | TNFSF12;TNFSF13;SENP3 | | | 7.86E-45 | Whole blood | European | 3841 | BIOSQTL_methQTL_EUR_2017 | 27918535 |
| rs116600817 | 17p13.1 | A | G | cg12418947 | chr17:7255013 | KCTD11 | | | 1.20E-10 | Whole blood | European | 3841 | BIOSQTL_methQTL_EUR_2017 | 27918535 |
| rs116600817 | 17p13.1 | A | G | cg09566763 | chr17:7553736 | ATP1B2 | | | 1.99E-10 | Whole blood | European | 3841 | BIOSQTL_methQTL_EUR_2017 | 27918535 |
| rs116600817 | 17p13.1 | A | G | cg09663193 | chr17:7486845 | MPDU1 | | | 6.70E-25 | Whole blood | European | 3841 | BIOSQTL_methQTL_EUR_2017 | 27918535 |
| rs116600817 | 17p13.1 | A | G | cg12045829 | chr17:7452902 | TNFSF12;TNFSF13 | | | 1.75E-37 | Whole blood | European | 3841 | BIOSQTL_methQTL_EUR_2017 | 27918535 |
| rs116600817 | 17p13.1 | A | G | cg05872923 | chr17:7461236 | TNFSF12;TNFSF13 | | | 4.53E-08 | Whole blood | European | 3841 | BIOSQTL_methQTL_EUR_2017 | 27918535 |
| rs116600817 | 17p13.1 | A | G | cg08324090 | chr17:7253213 | ACAP1 | | | 1.76E-18 | Whole blood | European | 3841 | BIOSQTL_methQTL_EUR_2017 | 27918535 |
| rs116600817 | 17p13.1 | A | G | cg13829089 | chr17:7460714 | TNFSF12;TNFSF13 | | | 1.08E-16 | Whole blood | European | 3841 | BIOSQTL_methQTL_EUR_2017 | 27918535 |
| rs116600817 | 17p13.1 | A | G | cg12239264 | chr17:7452067 | TNFSF12;TNFSF13 | | | 3.14E-36 | Whole blood | European | 3841 | BIOSQTL_methQTL_EUR_2017 | 27918535 |
| rs116600817 | 17p13.1 | A | G | cg01557754 | chr17:7342661 | FGF11 | 0.3879 | 0.0464 | 2.97E-16 | Cord blood | European | 771 | Gaunt-T_methQTL-Cord_EUR_2016 | 27036880 |
| rs116600817 | 17p13.1 | A | G | cg23161218 | chr17:7461614 | TNFSF12;TNFSF13 | | | 9.16E-10 | Whole blood | European | 3841 | BIOSQTL_methQTL_EUR_2017 | 27918535 |
| rs116600817 | 17p13.1 | A | G | cg25000600 | chr17:7621569 | DNAH2 | | | 5.08E-32 | Whole blood | European | 3841 | BIOSQTL_methQTL_EUR_2017 | 27918535 |
| rs116600817 | 17p13.1 | A | G | cg01229452 | chr17:7350258 | CHRNB1 | | | 2.49E-14 | Whole blood | European | 3841 | BIOSQTL_methQTL_EUR_2017 | 27918535 |
| rs116600817 | 17p13.1 | A | G | cg02676175 | chr17:7253696 | KCTD11;ACAP1 | | | 1.81E-18 | Whole blood | European | 3841 | BIOSQTL_methQTL_EUR_2017 | 27918535 |
| rs116600817 | 17p13.1 | A | G | cg05164926 | chr17:7255624 | KCTD11 | 0.2706 | 0.04551 | 4.05E-09 | Whole blood | European | 837 | Gaunt-T_methQTL-15up_EUR_2016 | 27036880 |
| rs116600817 | 17p13.1 | A | G | cg05164926 | chr17:7255648 | KCTD11 | | | 5.73E-16 | Whole blood | European | 3841 | BIOSQTL_methQTL_EUR_2017 | 27918535 |
| rs116600817 | 17p13.1 | A | G | cg07451297 | chr17:7486898 | MPDU1 | | | 1.73E-27 | Whole blood | European | 3841 | BIOSQTL_methQTL_EUR_2017 | 27918535 |

| | | | | | | | | | | | | | | |
|---|---|---|---|---|---|---|---|---|---|---|---|---|---|---|
| rs116600817 | 17p13.1 | A | G | cg06236279 | chr17:7617310 | | | | 5.60E-10 | Whole blood | European | 3841 | BIOSQTL_methQTL_EUR_2017 | 27918535 |
| rs116600817 | 17p13.1 | A | G | cg07639694 | chr17:7254885 | KCTD11 | | | 1.04E-15 | Whole blood | European | 3841 | BIOSQTL_methQTL_EUR_2017 | 27918535 |
| rs116600817 | 17p13.1 | A | G | cg14782678 | chr17:7339697 | TMEM102 | | | 2.05E-14 | Whole blood | European | 3841 | BIOSQTL_methQTL_EUR_2017 | 27918535 |
| rs116600817 | 17p13.1 | A | G | cg01557754 | chr17:7342685 | FGF11 | | | 2.69E-52 | Whole blood | European | 3841 | BIOSQTL_methQTL_EUR_2017 | 27918535 |
| rs116600817 | 17p13.1 | A | G | cg01557754 | chr17:7342661 | FGF11 | 0.3178 | 0.042 | 1.02E-13 | Whole blood | European | 837 | Gaunt-T_methQTL-15up_EUR_2016 | 27036880 |
| rs116600817 | 17p13.1 | A | G | cg17892169 | chr17:7452644 | TNFSF12;TNFSF13 | 0.3913 | 0.03767 | 1.10E-23 | Whole blood | European | 742 | Gaunt-T_methQTL-FOM_EUR_2016 | 27036880 |
| rs116600817 | 17p13.1 | A | G | cg01557754 | chr17:7342661 | FGF11 | 0.3453 | 0.04564 | 1.10E-13 | Whole blood | European | 837 | Gaunt-T_methQTL-Antenatal_EUR_2016 | 27036880 |
| rs116600817 | 17p13.1 | A | G | cg01557754 | chr17:7342661 | FGF11 | 0.2863 | 0.04533 | 4.66E-10 | Whole blood | European | 742 | Gaunt-T_methQTL-FOM_EUR_2016 | 27036880 |
| rs116600817 | 17p13.1 | A | G | cg25581039 | chr17:7350857 | CHRNB1 | | | 5.64E-11 | Whole blood | European | 3841 | BIOSQTL_methQTL_EUR_2017 | 27918535 |
| rs116600817 | 17p13.1 | A | G | cg13358186 | chr17:7461751 | TNFSF12;TNFSF13 | | | 1.33E-19 | Whole blood | European | 3841 | BIOSQTL_methQTL_EUR_2017 | 27918535 |
| rs116600817 | 17p13.1 | A | G | cg06931941 | chr17:7486746 | MPDU1 | | | 2.00E-20 | Whole blood | European | 3841 | BIOSQTL_methQTL_EUR_2017 | 27918535 |
| rs116600817 | 17p13.1 | A | G | cg21468274 | chr17:7339859 | TMEM102 | | | 2.82E-11 | Whole blood | European | 3841 | BIOSQTL_methQTL_EUR_2017 | 27918535 |
| rs116600817 | 17p13.1 | A | G | cg10479431 | chr17:7461445 | TNFSF12;TNFSF13 | | | 7.78E-14 | Whole blood | European | 3841 | BIOSQTL_methQTL_EUR_2017 | 27918535 |
| rs116600817 | 17p13.1 | A | G | cg25900902 | chr17:7253364 | ACAP1 | | | 4.72E-19 | Whole blood | European | 3841 | BIOSQTL_methQTL_EUR_2017 | 27918535 |
| rs116600817 | 17p13.1 | A | G | cg19440264 | chr17:7350389 | CHRNB1 | | | 2.05E-10 | Whole blood | European | 3841 | BIOSQTL_methQTL_EUR_2017 | 27918535 |
| rs116600817 | 17p13.1 | A | G | cg24517323 | chr17:7227588 | NEURL4 | | | 1.38E-10 | Whole blood | European | 3841 | BIOSQTL_methQTL_EUR_2017 | 27918535 |
| rs116600817 | 17p13.1 | A | G | cg16673904 | chr17:7521972 | SHBG | | | 1.02E-20 | Whole blood | European | 3841 | BIOSQTL_methQTL_EUR_2017 | 27918535 |
| rs116600817 | 17p13.1 | A | G | cg06812840 | chr17:7492500 | SOX15 | | | 1.69E-09 | Whole blood | European | 3841 | BIOSQTL_methQTL_EUR_2017 | 27918535 |
| rs116600817 | 17p13.1 | A | G | cg17379325 | chr17:7555435 | ATP1B2 | | | 5.49E-11 | Whole blood | European | 3841 | BIOSQTL_methQTL_EUR_2017 | 27918535 |
| rs116600817 | 17p13.1 | A | G | cg14824495 | chr17:7492777 | SOX15 | | | 5.97E-16 | Whole blood | European | 3841 | BIOSQTL_methQTL_EUR_2017 | 27918535 |
| rs116600817 | 17p13.1 | A | G | cg13098960 | chr17:7494175 | SOX15 | | | 3.59E-10 | Whole blood | European | 3841 | BIOSQTL_methQTL_EUR_2017 | 27918535 |
| rs116600817 | 17p13.1 | A | G | cg13546736 | chr17:7486882 | MPDU1 | | | 8.18E-29 | Whole blood | European | 3841 | BIOSQTL_methQTL_EUR_2017 | 27918535 |
| rs116600817 | 17p13.1 | A | G | cg17788227 | chr17:7347121 | FGF11;CHRNB1 | | | 5.79E-14 | Whole blood | European | 3841 | BIOSQTL_methQTL_EUR_2017 | 27918535 |
| rs116600817 | 17p13.1 | A | G | cg21808406 | chr17:7482451 | CD68 | | | 3.89E-09 | Whole blood | European | 3841 | BIOSQTL_methQTL_EUR_2017 | 27918535 |
| rs116600817 | 17p13.1 | A | G | cg07711099 | chr17:7621599 | DNAH2 | | | 1.95E-35 | Whole blood | European | 3841 | BIOSQTL_methQTL_EUR_2017 | 27918535 |
| rs116600817 | 17p13.1 | A | G | cg17107156 | chr17:7350268 | CHRNB1 | | | 9.46E-20 | Whole blood | European | 3841 | BIOSQTL_methQTL_EUR_2017 | 27918535 |
| rs116600817 | 17p13.1 | A | G | cg07148458 | chr17:7254301 | KCTD11;ACAP1 | 0.2911 | 0.04832 | 2.53E-09 | Whole blood | European | 837 | Gaunt-T_methQTL-15up_EUR_2016 | 27036880 |
| rs116600817 | 17p13.1 | A | G | cg22084563 | chr17:7255022 | KCTD11 | | | 1.11E-12 | Whole blood | European | 3841 | BIOSQTL_methQTL_EUR_2017 | 27918535 |
| rs116600817 | 17p13.1 | A | G | cg15792487 | chr17:7348292 | CHRNB1 | | | 1.20E-11 | Whole blood | European | 3841 | BIOSQTL_methQTL_EUR_2017 | 27918535 |
| rs116600817 | 17p13.1 | A | G | cg07168214 | chr17:7380136 | ZBTB4 | | | 6.59E-29 | Whole blood | European | 3841 | BIOSQTL_methQTL_EUR_2017 | 27918535 |
| rs116600817 | 17p13.1 | A | G | cg07925670 | chr17:7253513 | ACAP1 | | | 1.43E-19 | Whole blood | European | 3841 | BIOSQTL_methQTL_EUR_2017 | 27918535 |

| | | | | | | | | | | | | | |
|---|---|---|---|---|---|---|---|---|---|---|---|---|---|
| rs116600817 | 17p13.1 | A | G | cg18884137 | chr17:7348466 | CHRNB1 | | | 1.46E-20 | Whole blood | European | 3841 | BIOSQTL_methQTL_EUR_2017 | 27918535 |
| rs116600817 | 17p13.1 | A | G | cg24128434 | chr17:7623107 | DNAH2 | | | 1.27E-17 | Whole blood | European | 3841 | BIOSQTL_methQTL_EUR_2017 | 27918535 |
| rs116600817 | 17p13.1 | A | G | cg24479590 | chr17:7348298 | CHRNB1 | | | 1.01E-16 | Whole blood | European | 3841 | BIOSQTL_methQTL_EUR_2017 | 27918535 |
| rs116600817 | 17p13.1 | A | G | cg14050954 | chr17:7239550 | ACAP1 | | | 1.02E-12 | Whole blood | European | 3841 | BIOSQTL_methQTL_EUR_2017 | 27918535 |
| rs116600817 | 17p13.1 | A | G | cg17892169 | chr17:7452668 | TNFSF12;TNFSF13 | | | 1.94E-138 | Whole blood | European | 3841 | BIOSQTL_methQTL_EUR_2017 | 27918535 |
| rs76774446 | 17q21.32 | A | C | cg26909813 | chr17:45057218 | RPRML | 0.4497 | 0.06003 | 1.96E-13 | Whole blood | European | 742 | Gaunt-T_methQTL-FOM_EUR_2016 | 27036880 |
| rs76774446 | 17q21.32 | A | C | cg26170930 | chr17:45020334 | | | | 1.47E-24 | Whole blood | European | 3841 | BIOSQTL_methQTL_EUR_2017 | 27918535 |
| rs76774446 | 17q21.32 | A | C | cg13665025 | chr17:45056837 | RPRML | 0.7121 | 0.05672 | 3.04E-33 | Whole blood | European | 837 | Gaunt-T_methQTL-15up_EUR_2016 | 27036880 |
| rs76774446 | 17q21.32 | A | C | cg08010546 | chr17:45060311 | | | | 1.01E-49 | Whole blood | European | 3841 | BIOSQTL_methQTL_EUR_2017 | 27918535 |
| rs76774446 | 17q21.32 | A | C | cg13665025 | chr17:45056837 | RPRML | 0.7184 | 0.05644 | 4.90E-34 | Whole blood | European | 834 | Gaunt-T_methQTL-F7_EUR_2016 | 27036880 |
| rs76774446 | 17q21.32 | A | C | cg13665025 | chr17:45056837 | RPRML | 0.8346 | 0.0569 | 3.98E-43 | Cord blood | European | 771 | Gaunt-T_methQTL-Cord_EUR_2016 | 27036880 |
| rs76774446 | 17q21.32 | A | C | cg20769177 | chr17:44928492 | WNT9B | | | 9.12E-22 | Whole blood | European | 3841 | BIOSQTL_methQTL_EUR_2017 | 27918535 |
| rs76774446 | 17q21.32 | A | C | cg16759221 | chr17:45003001 | GOSR2 | | | 3.73E-34 | Whole blood | European | 3841 | BIOSQTL_methQTL_EUR_2017 | 27918535 |
| rs76774446 | 17q21.32 | A | C | cg12311600 | chr17:44439469 | ARL17A;ARL17B | 0.3395 | 0.05713 | 4.14E-09 | Whole blood | European | 834 | Gaunt-T_methQTL-F7_EUR_2016 | 27036880 |
| rs76774446 | 17q21.32 | A | C | cg06096245 | chr17:45018442 | GOSR2 | | | 3.99E-16 | Whole blood | European | 3841 | BIOSQTL_methQTL_EUR_2017 | 27918535 |
| rs76774446 | 17q21.32 | A | C | cg17881353 | chr17:44928186 | WNT9B | | | 2.58E-17 | Whole blood | European | 3841 | BIOSQTL_methQTL_EUR_2017 | 27918535 |
| rs76774446 | 17q21.32 | A | C | cg26909813 | chr17:45057218 | RPRML | 0.341 | 0.05934 | 1.31E-08 | Cord blood | European | 771 | Gaunt-T_methQTL-Cord_EUR_2016 | 27036880 |
| rs76774446 | 17q21.32 | A | C | cg13665025 | chr17:45056861 | RPRML | | | 7.38E-134 | Whole blood | European | 3841 | BIOSQTL_methQTL_EUR_2017 | 27918535 |
| rs76774446 | 17q21.32 | A | C | cg08358943 | chr17:44889560 | WNT3 | | | 3.23E-35 | Whole blood | European | 3841 | BIOSQTL_methQTL_EUR_2017 | 27918535 |
| rs76774446 | 17q21.32 | A | C | cg07993743 | chr17:44928264 | WNT9B | | | 7.53E-20 | Whole blood | European | 3841 | BIOSQTL_methQTL_EUR_2017 | 27918535 |
| rs76774446 | 17q21.32 | A | C | cg07142555 | chr17:45056905 | RPRML | | | 2.92E-20 | Whole blood | European | 3841 | BIOSQTL_methQTL_EUR_2017 | 27918535 |
| rs76774446 | 17q21.32 | A | C | cg12311600 | chr17:44439469 | ARL17A;ARL17B | 0.3531 | 0.05558 | 3.48E-10 | Whole blood | European | 837 | Gaunt-T_methQTL-15up_EUR_2016 | 27036880 |
| rs76774446 | 17q21.32 | A | C | cg26909813 | chr17:45057218 | RPRML | 0.4851 | 0.05672 | 5.74E-17 | Whole blood | European | 834 | Gaunt-T_methQTL-F7_EUR_2016 | 27036880 |
| rs76774446 | 17q21.32 | A | C | cg24434387 | chr17:45051716 | | | | 7.33E-17 | Whole blood | European | 3841 | BIOSQTL_methQTL_EUR_2017 | 27918535 |
| rs76774446 | 17q21.32 | A | C | cg26226202 | chr17:44928528 | WNT9B | | | 1.19E-13 | Whole blood | European | 3841 | BIOSQTL_methQTL_EUR_2017 | 27918535 |
| rs76774446 | 17q21.32 | A | C | cg13665025 | chr17:45056837 | RPRML | 0.801 | 0.05791 | 5.50E-39 | Whole blood | European | 837 | Gaunt-T_methQTL-Antenatal_EUR_2016 | 27036880 |
| rs76774446 | 17q21.32 | A | C | cg13665025 | chr17:45056837 | RPRML | 0.787 | 0.05866 | 6.82E-37 | Whole blood | European | 742 | Gaunt-T_methQTL-FOM_EUR_2016 | 27036880 |
| rs76774446 | 17q21.32 | A | C | cg26909813 | chr17:45057218 | RPRML | 0.4145 | 0.0578 | 1.64E-12 | Whole blood | European | 837 | Gaunt-T_methQTL-15up_EUR_2016 | 27036880 |
| rs76774446 | 17q21.32 | A | C | cg13550419 | chr17:44898486 | | | | 2.28E-16 | Whole blood | European | 3841 | BIOSQTL_methQTL_EUR_2017 | 27918535 |
| rs76774446 | 17q21.32 | A | C | cg01440864 | chr17:44892111 | WNT3 | | | 3.47E-19 | Whole blood | European | 3841 | BIOSQTL_methQTL_EUR_2017 | 27918535 |
| rs76774446 | 17q21.32 | A | C | cg24703533 | chr17:45055294 | | | | 7.88E-13 | Whole blood | European | 3841 | BIOSQTL_methQTL_EUR_2017 | 27918535 |

| | | | | | | | | | | | | | | |
|---|---|---|---|---|---|---|---|---|---|---|---|---|---|---|
| rs76774446 | 17q21.32 | A | C | cg15125338 | chr17:44899370 | | | | 3.04E-12 | Whole blood | European | 3841 | BIOSQTL_methQTL_EUR_2017 | 27918535 |
| rs76774446 | 17q21.32 | A | C | cg26909813 | chr17:45057218 | RPRML | 0.4829 | 0.05982 | 2.69E-15 | Whole blood | European | 837 | Gaunt-T_methQTL-Antenatal_EUR_2016 | 27036880 |
| rs3746471 | 20q11.23 | A | G | cg14003035 | chr20:36889388 | KIAA1755 | | | 5.14E-11 | Whole blood | European | 3841 | BIOSQTL_methQTL_EUR_2017 | 27918535 |
| rs3746471 | 20q11.23 | A | G | cg20472707 | chr20:36889314 | KIAA1755 | | | 9.58E-09 | Whole blood | European | 3841 | BIOSQTL_methQTL_EUR_2017 | 27918535 |
| rs3746471 | 20q11.23 | A | G | cg02379107 | chr20:36889402 | KIAA1755 | | | 4.09E-09 | Whole blood | European | 3841 | BIOSQTL_methQTL_EUR_2017 | 27918535 |
| rs2834618 | 21q22.12 | G | T | cg01183122 | chr21:35884324 | KCNE1 | | | 7.06E-20 | Whole blood | European | 3841 | BIOSQTL_methQTL_EUR_2017 | 27918535 |
| rs2834618 | 21q22.12 | G | T | cg02471848 | chr21:35990178 | | | | 1.12E-09 | Whole blood | European | 3841 | BIOSQTL_methQTL_EUR_2017 | 27918535 |
| rs2834618 | 21q22.12 | G | T | cg21088514 | chr21:35884352 | KCNE1 | | | 1.19E-18 | Whole blood | European | 3841 | BIOSQTL_methQTL_EUR_2017 | 27918535 |

### Online Table VII. Significant GTEx eQTLs of the index variants from MTAG-CES

| SNP | Locus | Variant.Id | NES | Pvalue | Gene | GeneSymbol | Tissue |
|---|---|---|---|---|---|---|---|
| rs10753933 | 1q32.1 | chr1_203057086_T_G_b38 | 0.47 | 1.20E-30 | ENSG00000143847.15 | PPFIA4 | Whole Blood |
| rs10753933 | 1q32.1 | chr1_203057086_T_G_b38 | 0.16 | 1.10E-07 | ENSG00000143847.15 | PPFIA4 | Artery - Aorta |
| rs10753933 | 1q32.1 | chr1_203057086_T_G_b38 | -0.12 | 3.40E-05 | ENSG00000122180.4 | MYOG | Muscle - Skeletal |
| rs11099098 | 4q21.21 | chr4_80248758_G_T_b38 | 0.62 | 1.10E-07 | ENSG00000138675.16 | FGF5 | Kidney - Cortex |
| rs11099098 | 4q21.21 | chr4_80248758_G_T_b38 | 0.32 | 6.90E-05 | ENSG00000152785.6 | BMP3 | Brain - Cerebellum |
| rs11099098 | 4q21.21 | chr4_80248758_G_T_b38 | -0.2 | 0.00013 | ENSG00000152784.15 | PRDM8 | Testis |
| rs11125871 | 2p15 | chr2_61242991_C_T_b38 | -0.63 | 1.40E-57 | ENSG00000237651.6 | C2orf74 | Muscle - Skeletal |
| rs11125871 | 2p15 | chr2_61242991_C_T_b38 | 0.92 | 2.60E-55 | ENSG00000212978.6 | AC016747.3 | Testis |
| rs11125871 | 2p15 | chr2_61242991_C_T_b38 | -0.7 | 2.40E-47 | ENSG00000237651.6 | C2orf74 | Artery - Tibial |
| rs11125871 | 2p15 | chr2_61242991_C_T_b38 | -0.69 | 3.40E-47 | ENSG00000237651.6 | C2orf74 | Adipose - Subcutaneous |
| rs11125871 | 2p15 | chr2_61242991_C_T_b38 | -0.66 | 8.70E-47 | ENSG00000237651.6 | C2orf74 | Thyroid |
| rs11125871 | 2p15 | chr2_61242991_C_T_b38 | -0.64 | 1.20E-44 | ENSG00000237651.6 | C2orf74 | Nerve - Tibial |
| rs11125871 | 2p15 | chr2_61242991_C_T_b38 | -0.74 | 2.30E-42 | ENSG00000237651.6 | C2orf74 | Esophagus - Mucosa |
| rs11125871 | 2p15 | chr2_61242991_C_T_b38 | 0.36 | 2.30E-42 | ENSG00000173209.22 | AHSA2 | Whole Blood |
| rs11125871 | 2p15 | chr2_61242991_C_T_b38 | -0.76 | 1.60E-41 | ENSG00000237651.6 | C2orf74 | Cells - Cultured fibroblasts |
| rs11125871 | 2p15 | chr2_61242991_C_T_b38 | -0.58 | 2.60E-40 | ENSG00000237651.6 | C2orf74 | Skin - Sun Exposed (Lower leg) |
| rs11125871 | 2p15 | chr2_61242991_C_T_b38 | -0.75 | 3.10E-39 | ENSG00000237651.6 | C2orf74 | Esophagus - Muscularis |
| rs11125871 | 2p15 | chr2_61242991_C_T_b38 | -0.6 | 6.60E-39 | ENSG00000237651.6 | C2orf74 | Lung |
| rs11125871 | 2p15 | chr2_61242991_C_T_b38 | -0.66 | 8.20E-36 | ENSG00000237651.6 | C2orf74 | Adipose - Visceral (Omentum) |
| rs11125871 | 2p15 | chr2_61242991_C_T_b38 | -0.56 | 3.00E-32 | ENSG00000237651.6 | C2orf74 | Skin - Not Sun Exposed (Suprapubic) |
| rs11125871 | 2p15 | chr2_61242991_C_T_b38 | -0.66 | 1.20E-30 | ENSG00000237651.6 | C2orf74 | Breast - Mammary Tissue |
| rs11125871 | 2p15 | chr2_61242991_C_T_b38 | -0.71 | 3.00E-29 | ENSG00000237651.6 | C2orf74 | Artery - Aorta |
| rs11125871 | 2p15 | chr2_61242991_C_T_b38 | -0.69 | 4.30E-28 | ENSG00000237651.6 | C2orf74 | Stomach |
| rs11125871 | 2p15 | chr2_61242991_C_T_b38 | -0.61 | 2.80E-26 | ENSG00000237651.6 | C2orf74 | Colon - Sigmoid |
| rs11125871 | 2p15 | chr2_61242991_C_T_b38 | -0.51 | 5.30E-25 | ENSG00000237651.6 | C2orf74 | Heart - Left Ventricle |
| rs11125871 | 2p15 | chr2_61242991_C_T_b38 | -0.37 | 3.80E-24 | ENSG00000237651.6 | C2orf74 | Colon - Transverse |
| rs11125871 | 2p15 | chr2_61242991_C_T_b38 | -0.55 | 5.70E-24 | ENSG00000237651.6 | C2orf74 | Heart - Atrial Appendage |
| rs11125871 | 2p15 | chr2_61242991_C_T_b38 | -0.67 | 2.60E-23 | ENSG00000237651.6 | C2orf74 | Esophagus - Gastroesophageal Junction |
| rs11125871 | 2p15 | chr2_61242991_C_T_b38 | -0.68 | 1.70E-20 | ENSG00000237651.6 | C2orf74 | Pancreas |
| rs11125871 | 2p15 | chr2_61242991_C_T_b38 | -0.8 | 2.40E-20 | ENSG00000237651.6 | C2orf74 | Spleen |
| rs11125871 | 2p15 | chr2_61242991_C_T_b38 | -0.69 | 5.60E-20 | ENSG00000237651.6 | C2orf74 | Adrenal Gland |
| rs11125871 | 2p15 | chr2_61242991_C_T_b38 | -0.81 | 1.40E-19 | ENSG00000237651.6 | C2orf74 | Brain - Cerebellum |
| rs11125871 | 2p15 | chr2_61242991_C_T_b38 | -0.66 | 1.20E-18 | ENSG00000237651.6 | C2orf74 | Pituitary |
| rs11125871 | 2p15 | chr2_61242991_C_T_b38 | -0.75 | 2.50E-18 | ENSG00000237651.6 | C2orf74 | Brain - Cortex |
| rs11125871 | 2p15 | chr2_61242991_C_T_b38 | 0.35 | 4.40E-18 | ENSG00000173209.22 | AHSA2 | Nerve - Tibial |
| rs11125871 | 2p15 | chr2_61242991_C_T_b38 | -0.48 | 4.70E-18 | ENSG00000271889.1 | RP11-493E12.1 | Cells - Cultured fibroblasts |
| rs11125871 | 2p15 | chr2_61242991_C_T_b38 | -0.36 | 9.80E-18 | ENSG00000237651.6 | C2orf74 | Whole Blood |
| rs11125871 | 2p15 | chr2_61242991_C_T_b38 | 0.29 | 1.40E-17 | ENSG00000173209.22 | AHSA2 | Adipose - Subcutaneous |
| rs11125871 | 2p15 | chr2_61242991_C_T_b38 | -0.26 | 1.50E-16 | ENSG00000271889.1 | RP11-493E12.1 | Testis |
| rs11125871 | 2p15 | chr2_61242991_C_T_b38 | 0.18 | 7.10E-16 | ENSG00000173209.22 | AHSA2 | Muscle - Skeletal |
| rs11125871 | 2p15 | chr2_61242991_C_T_b38 | 0.26 | 1.40E-15 | ENSG00000212978.6 | AC016747.3 | Whole Blood |
| rs11125871 | 2p15 | chr2_61242991_C_T_b38 | 0.18 | 1.60E-15 | ENSG00000173209.22 | AHSA2 | Skin - Not Sun Exposed (Suprapubic) |
| rs11125871 | 2p15 | chr2_61242991_C_T_b38 | -0.59 | 1.90E-15 | ENSG00000237651.6 | C2orf74 | Prostate |
| rs11125871 | 2p15 | chr2_61242991_C_T_b38 | -0.22 | 2.10E-15 | ENSG00000270820.5 | RP11-355B11.2 | Thyroid |
| rs11125871 | 2p15 | chr2_61242991_C_T_b38 | -0.6 | 3.90E-15 | ENSG00000237651.6 | C2orf74 | Brain - Frontal Cortex (BA9) |
| rs11125871 | 2p15 | chr2_61242991_C_T_b38 | -0.38 | 1.20E-14 | ENSG00000273302.1 | RP11-493E12.2 | Nerve - Tibial |
| rs11125871 | 2p15 | chr2_61242991_C_T_b38 | -0.54 | 1.40E-14 | ENSG00000237651.6 | C2orf74 | Ovary |
| rs11125871 | 2p15 | chr2_61242991_C_T_b38 | -0.64 | 4.30E-14 | ENSG00000237651.6 | C2orf74 | Brain - Cerebellar Hemisphere |
| rs11125871 | 2p15 | chr2_61242991_C_T_b38 | 0.32 | 4.50E-14 | ENSG00000212978.6 | AC016747.3 | Heart - Atrial Appendage |
| rs11125871 | 2p15 | chr2_61242991_C_T_b38 | -0.7 | 7.00E-14 | ENSG00000237651.6 | C2orf74 | Minor Salivary Gland |
| rs11125871 | 2p15 | chr2_61242991_C_T_b38 | -0.58 | 4.20E-13 | ENSG00000237651.6 | C2orf74 | Brain - Anterior cingulate cortex (BA24) |

| | | | | | | | |
|---|---|---|---|---|---|---|---|
| rs11125871 | 2p15 | chr2_61242991_C_T_b38 | -0.2 | 5.20E-13 | ENSG00000270820.5 | RP11-355B11.2 | Whole Blood |
| rs11125871 | 2p15 | chr2_61242991_C_T_b38 | -0.26 | 5.90E-13 | ENSG00000270820.5 | RP11-355B11.2 | Skin - Sun Exposed (Lower leg) |
| rs11125871 | 2p15 | chr2_61242991_C_T_b38 | -0.37 | 6.00E-13 | ENSG00000271889.1 | RP11-493E12.1 | Thyroid |
| rs11125871 | 2p15 | chr2_61242991_C_T_b38 | 0.14 | 7.30E-13 | ENSG00000173209.22 | AHSA2 | Skin - Sun Exposed (Lower leg) |
| rs11125871 | 2p15 | chr2_61242991_C_T_b38 | -0.55 | 1.30E-12 | ENSG00000237651.6 | C2orf74 | Vagina |
| rs11125871 | 2p15 | chr2_61242991_C_T_b38 | -0.63 | 1.60E-12 | ENSG00000237651.6 | C2orf74 | Artery - Coronary |
| rs11125871 | 2p15 | chr2_61242991_C_T_b38 | 0.38 | 2.00E-12 | ENSG00000173209.22 | AHSA2 | Testis |
| rs11125871 | 2p15 | chr2_61242991_C_T_b38 | -0.52 | 2.60E-12 | ENSG00000237651.6 | C2orf74 | Liver |
| rs11125871 | 2p15 | chr2_61242991_C_T_b38 | 0.44 | 3.10E-12 | ENSG00000212978.6 | AC016747.3 | Spleen |
| rs11125871 | 2p15 | chr2_61242991_C_T_b38 | 0.49 | 1.00E-11 | ENSG00000173209.22 | AHSA2 | Brain - Hippocampus |
| rs11125871 | 2p15 | chr2_61242991_C_T_b38 | 0.34 | 1.60E-11 | ENSG00000212978.6 | AC016747.3 | Heart - Left Ventricle |
| rs11125871 | 2p15 | chr2_61242991_C_T_b38 | 0.22 | 2.00E-11 | ENSG00000212978.6 | AC016747.3 | Adipose - Visceral (Omentum) |
| rs11125871 | 2p15 | chr2_61242991_C_T_b38 | -0.39 | 2.10E-11 | ENSG00000237651.6 | C2orf74 | Brain - Putamen (basal ganglia) |
| rs11125871 | 2p15 | chr2_61242991_C_T_b38 | -0.33 | 2.20E-11 | ENSG00000270820.5 | RP11-355B11.2 | Testis |
| rs11125871 | 2p15 | chr2_61242991_C_T_b38 | 0.19 | 2.30E-11 | ENSG00000173209.22 | AHSA2 | Thyroid |
| rs11125871 | 2p15 | chr2_61242991_C_T_b38 | -0.45 | 3.10E-11 | ENSG00000237651.6 | C2orf74 | Brain - Hippocampus |
| rs11125871 | 2p15 | chr2_61242991_C_T_b38 | 0.51 | 3.40E-11 | ENSG00000173209.22 | AHSA2 | Pituitary |
| rs11125871 | 2p15 | chr2_61242991_C_T_b38 | 0.58 | 5.70E-11 | ENSG00000173209.22 | AHSA2 | Brain - Cerebellum |
| rs11125871 | 2p15 | chr2_61242991_C_T_b38 | -0.33 | 6.20E-11 | ENSG00000237651.6 | C2orf74 | Brain - Caudate (basal ganglia) |
| rs11125871 | 2p15 | chr2_61242991_C_T_b38 | 0.15 | 6.70E-11 | ENSG00000173209.22 | AHSA2 | Esophagus - Mucosa |
| rs11125871 | 2p15 | chr2_61242991_C_T_b38 | -0.78 | 8.70E-11 | ENSG00000237651.6 | C2orf74 | Cells - EBV-transformed lymphocytes |
| rs11125871 | 2p15 | chr2_61242991_C_T_b38 | 0.44 | 1.20E-10 | ENSG00000173209.22 | AHSA2 | Brain - Nucleus accumbens (basal ganglia) |
| rs11125871 | 2p15 | chr2_61242991_C_T_b38 | 0.18 | 1.40E-10 | ENSG00000173209.22 | AHSA2 | Lung |
| rs11125871 | 2p15 | chr2_61242991_C_T_b38 | 0.21 | 2.20E-10 | ENSG00000173209.22 | AHSA2 | Adipose - Visceral (Omentum) |
| rs11125871 | 2p15 | chr2_61242991_C_T_b38 | -0.3 | 2.70E-10 | ENSG00000237651.6 | C2orf74 | Brain - Nucleus accumbens (basal ganglia) |
| rs11125871 | 2p15 | chr2_61242991_C_T_b38 | -0.3 | 6.50E-10 | ENSG00000237651.6 | C2orf74 | Small Intestine - Terminal Ileum |
| rs11125871 | 2p15 | chr2_61242991_C_T_b38 | 0.22 | 6.60E-10 | ENSG00000212978.6 | AC016747.3 | Muscle - Skeletal |
| rs11125871 | 2p15 | chr2_61242991_C_T_b38 | 0.38 | 7.20E-10 | ENSG00000173209.22 | AHSA2 | Brain - Cortex |
| rs11125871 | 2p15 | chr2_61242991_C_T_b38 | 0.19 | 8.50E-10 | ENSG00000173209.22 | AHSA2 | Esophagus - Muscularis |
| rs11125871 | 2p15 | chr2_61242991_C_T_b38 | -0.31 | 1.50E-09 | ENSG00000273302.1 | RP11-493E12.2 | Adipose - Subcutaneous |
| rs11125871 | 2p15 | chr2_61242991_C_T_b38 | -0.31 | 2.00E-09 | ENSG00000273302.1 | RP11-493E12.2 | Esophagus - Muscularis |
| rs11125871 | 2p15 | chr2_61242991_C_T_b38 | 0.46 | 2.10E-09 | ENSG00000173209.22 | AHSA2 | Brain - Anterior cingulate cortex (BA24) |
| rs11125871 | 2p15 | chr2_61242991_C_T_b38 | 0.15 | 2.50E-09 | ENSG00000173209.22 | AHSA2 | Cells - Cultured fibroblasts |
| rs11125871 | 2p15 | chr2_61242991_C_T_b38 | 0.22 | 2.70E-09 | ENSG00000173209.22 | AHSA2 | Breast - Mammary Tissue |
| rs11125871 | 2p15 | chr2_61242991_C_T_b38 | 0.26 | 2.90E-09 | ENSG00000173209.22 | AHSA2 | Colon - Sigmoid |
| rs11125871 | 2p15 | chr2_61242991_C_T_b38 | -0.14 | 5.10E-09 | ENSG00000237651.6 | C2orf74 | Testis |
| rs11125871 | 2p15 | chr2_61242991_C_T_b38 | 0.49 | 7.20E-09 | ENSG00000173209.22 | AHSA2 | Brain - Putamen (basal ganglia) |
| rs11125871 | 2p15 | chr2_61242991_C_T_b38 | -0.57 | 1.70E-08 | ENSG00000237651.6 | C2orf74 | Uterus |
| rs11125871 | 2p15 | chr2_61242991_C_T_b38 | 0.36 | 2.40E-08 | ENSG00000173209.22 | AHSA2 | Brain - Frontal Cortex (BA9) |
| rs11125871 | 2p15 | chr2_61242991_C_T_b38 | 0.47 | 3.20E-08 | ENSG00000173209.22 | AHSA2 | Brain - Cerebellar Hemisphere |
| rs11125871 | 2p15 | chr2_61242991_C_T_b38 | 0.38 | 3.30E-08 | ENSG00000173209.22 | AHSA2 | Brain - Hypothalamus |
| rs11125871 | 2p15 | chr2_61242991_C_T_b38 | 0.18 | 3.40E-08 | ENSG00000270820.5 | RP11-355B11.2 | Muscle - Skeletal |
| rs11125871 | 2p15 | chr2_61242991_C_T_b38 | -0.25 | 5.70E-08 | ENSG00000270820.5 | RP11-355B11.2 | Pituitary |
| rs11125871 | 2p15 | chr2_61242991_C_T_b38 | -0.16 | 8.80E-08 | ENSG00000273302.1 | RP11-493E12.2 | Whole Blood |
| rs11125871 | 2p15 | chr2_61242991_C_T_b38 | -0.46 | 9.50E-08 | ENSG00000237651.6 | C2orf74 | Brain - Substantia nigra |
| rs11125871 | 2p15 | chr2_61242991_C_T_b38 | -0.42 | 9.70E-08 | ENSG00000237651.6 | C2orf74 | Brain - Spinal cord (cervical c-1) |
| rs11125871 | 2p15 | chr2_61242991_C_T_b38 | 0.22 | 1.10E-07 | ENSG00000173209.22 | AHSA2 | Liver |
| rs11125871 | 2p15 | chr2_61242991_C_T_b38 | 0.19 | 1.20E-07 | ENSG00000173209.22 | AHSA2 | Heart - Atrial Appendage |
| rs11125871 | 2p15 | chr2_61242991_C_T_b38 | -0.37 | 1.20E-07 | ENSG00000237651.6 | C2orf74 | Brain - Hypothalamus |
| rs11125871 | 2p15 | chr2_61242991_C_T_b38 | 0.52 | 1.30E-07 | ENSG00000173209.22 | AHSA2 | Brain - Spinal cord (cervical c-1) |
| rs11125871 | 2p15 | chr2_61242991_C_T_b38 | -0.14 | 1.40E-07 | ENSG00000270820.5 | RP11-355B11.2 | Nerve - Tibial |

| | | | | | | | |
|---|---|---|---|---|---|---|---|
| rs11125871 | 2p15 | chr2_61242991_C_T_b38 | -0.33 | 2.10E-07 | ENSG00000271889.1 | RP11-493E12.1 | Esophagus - Muscularis |
| rs11125871 | 2p15 | chr2_61242991_C_T_b38 | -0.48 | 3.00E-07 | ENSG00000237651.6 | C2orf74 | Kidney - Cortex |
| rs11125871 | 2p15 | chr2_61242991_C_T_b38 | -0.18 | 3.10E-07 | ENSG00000270820.5 | RP11-355B11.2 | Skin - Not Sun Exposed (Suprapubic) |
| rs11125871 | 2p15 | chr2_61242991_C_T_b38 | -0.22 | 4.60E-07 | ENSG00000273302.1 | RP11-493E12.2 | Thyroid |
| rs11125871 | 2p15 | chr2_61242991_C_T_b38 | -0.44 | 4.70E-07 | ENSG00000273302.1 | RP11-493E12.2 | Brain - Hippocampus |
| rs11125871 | 2p15 | chr2_61242991_C_T_b38 | 0.34 | 6.40E-07 | ENSG00000173209.22 | AHSA2 | Brain - Caudate (basal ganglia) |
| rs11125871 | 2p15 | chr2_61242991_C_T_b38 | -0.38 | 6.80E-07 | ENSG00000273302.1 | RP11-493E12.2 | Ovary |
| rs11125871 | 2p15 | chr2_61242991_C_T_b38 | -0.29 | 7.10E-07 | ENSG00000271889.1 | RP11-493E12.1 | Esophagus - Mucosa |
| rs11125871 | 2p15 | chr2_61242991_C_T_b38 | 0.29 | 1.50E-06 | ENSG00000173209.22 | AHSA2 | Spleen |
| rs11125871 | 2p15 | chr2_61242991_C_T_b38 | 0.27 | 2.10E-06 | ENSG00000212978.6 | AC016747.3 | Pituitary |
| rs11125871 | 2p15 | chr2_61242991_C_T_b38 | 0.18 | 2.10E-06 | ENSG00000212978.6 | AC016747.3 | Breast - Mammary Tissue |
| rs11125871 | 2p15 | chr2_61242991_C_T_b38 | 0.2 | 2.20E-06 | ENSG00000173209.22 | AHSA2 | Pancreas |
| rs11125871 | 2p15 | chr2_61242991_C_T_b38 | -0.33 | 2.30E-06 | ENSG00000271889.1 | RP11-493E12.1 | Breast - Mammary Tissue |
| rs11125871 | 2p15 | chr2_61242991_C_T_b38 | 0.43 | 2.50E-06 | ENSG00000173209.22 | AHSA2 | Brain - Amygdala |
| rs11125871 | 2p15 | chr2_61242991_C_T_b38 | -0.43 | 3.00E-06 | ENSG00000273302.1 | RP11-493E12.2 | Prostate |
| rs11125871 | 2p15 | chr2_61242991_C_T_b38 | 0.29 | 3.10E-06 | ENSG00000173209.22 | AHSA2 | Prostate |
| rs11125871 | 2p15 | chr2_61242991_C_T_b38 | 0.14 | 3.50E-06 | ENSG00000173209.22 | AHSA2 | Artery - Tibial |
| rs11125871 | 2p15 | chr2_61242991_C_T_b38 | 0.1 | 3.60E-06 | ENSG00000115464.14 | USP34 | Cells - Cultured fibroblasts |
| rs11125871 | 2p15 | chr2_61242991_C_T_b38 | -0.34 | 3.80E-06 | ENSG00000237651.6 | C2orf74 | Brain - Amygdala |
| rs11125871 | 2p15 | chr2_61242991_C_T_b38 | -0.35 | 3.90E-06 | ENSG00000273302.1 | RP11-493E12.2 | Adrenal Gland |
| rs11125871 | 2p15 | chr2_61242991_C_T_b38 | 0.25 | 4.40E-06 | ENSG00000162927.13 | PUS10 | Colon - Sigmoid |
| rs11125871 | 2p15 | chr2_61242991_C_T_b38 | 0.19 | 5.40E-06 | ENSG00000173209.22 | AHSA2 | Colon - Transverse |
| rs11125871 | 2p15 | chr2_61242991_C_T_b38 | 0.17 | 6.70E-06 | ENSG00000212978.6 | AC016747.3 | Adipose - Subcutaneous |
| rs11125871 | 2p15 | chr2_61242991_C_T_b38 | 0.12 | 6.70E-06 | ENSG00000173209.22 | AHSA2 | Heart - Left Ventricle |
| rs11125871 | 2p15 | chr2_61242991_C_T_b38 | -0.31 | 7.80E-06 | ENSG00000271889.1 | RP11-493E12.1 | Pituitary |
| rs11125871 | 2p15 | chr2_61242991_C_T_b38 | -0.38 | 8.70E-06 | ENSG00000271889.1 | RP11-493E12.1 | Pancreas |
| rs11125871 | 2p15 | chr2_61242991_C_T_b38 | -0.22 | 9.40E-06 | ENSG00000273302.1 | RP11-493E12.2 | Muscle - Skeletal |
| rs11125871 | 2p15 | chr2_61242991_C_T_b38 | 0.099 | 1.30E-05 | ENSG00000115464.14 | USP34 | Lung |
| rs11125871 | 2p15 | chr2_61242991_C_T_b38 | -0.32 | 1.40E-05 | ENSG00000273302.1 | RP11-493E12.2 | Colon - Transverse |
| rs11125871 | 2p15 | chr2_61242991_C_T_b38 | 0.26 | 1.50E-05 | ENSG00000162927.13 | PUS10 | Adrenal Gland |
| rs11125871 | 2p15 | chr2_61242991_C_T_b38 | 0.17 | 1.50E-05 | ENSG00000162927.13 | PUS10 | Nerve - Tibial |
| rs11125871 | 2p15 | chr2_61242991_C_T_b38 | -0.26 | 1.70E-05 | ENSG00000273302.1 | RP11-493E12.2 | Colon - Sigmoid |
| rs11125871 | 2p15 | chr2_61242991_C_T_b38 | -0.25 | 2.00E-05 | ENSG00000273302.1 | RP11-493E12.2 | Artery - Aorta |
| rs11125871 | 2p15 | chr2_61242991_C_T_b38 | -0.29 | 2.60E-05 | ENSG00000271889.1 | RP11-493E12.1 | Stomach |
| rs11125871 | 2p15 | chr2_61242991_C_T_b38 | 0.17 | 2.70E-05 | ENSG00000212978.6 | AC016747.3 | Nerve - Tibial |
| rs11125871 | 2p15 | chr2_61242991_C_T_b38 | -0.28 | 2.70E-05 | ENSG00000270820.5 | RP11-355B11.2 | Brain - Cerebellum |
| rs11125871 | 2p15 | chr2_61242991_C_T_b38 | 0.17 | 2.70E-05 | ENSG00000162927.13 | PUS10 | Esophagus - Mucosa |
| rs11125871 | 2p15 | chr2_61242991_C_T_b38 | -0.33 | 3.40E-05 | ENSG00000271889.1 | RP11-493E12.1 | Adrenal Gland |
| rs11125871 | 2p15 | chr2_61242991_C_T_b38 | -0.39 | 4.90E-05 | ENSG00000271889.1 | RP11-493E12.1 | Brain - Cerebellum |
| rs11125871 | 2p15 | chr2_61242991_C_T_b38 | 0.15 | 5.00E-05 | ENSG00000173209.22 | AHSA2 | Stomach |
| rs11125871 | 2p15 | chr2_61242991_C_T_b38 | 0.23 | 5.30E-05 | ENSG00000212978.6 | AC016747.3 | Artery - Coronary |
| rs11125871 | 2p15 | chr2_61242991_C_T_b38 | -0.29 | 7.00E-05 | ENSG00000271889.1 | RP11-493E12.1 | Colon - Transverse |
| rs11125871 | 2p15 | chr2_61242991_C_T_b38 | -0.19 | 8.70E-05 | ENSG00000273302.1 | RP11-493E12.2 | Artery - Tibial |
| rs11125871 | 2p15 | chr2_61242991_C_T_b38 | 0.16 | 0.00011 | ENSG00000173209.22 | AHSA2 | Esophagus - Gastroesophageal Junction |
| rs11125871 | 2p15 | chr2_61242991_C_T_b38 | 0.061 | 0.00013 | ENSG00000115464.14 | USP34 | Skin - Sun Exposed (Lower leg) |
| rs11125871 | 2p15 | chr2_61242991_C_T_b38 | 0.13 | 0.00014 | ENSG00000162927.13 | PUS10 | Cells - Cultured fibroblasts |
| rs11125871 | 2p15 | chr2_61242991_C_T_b38 | -0.21 | 0.00018 | ENSG00000273302.1 | RP11-493E12.2 | Lung |
| rs11125871 | 2p15 | chr2_61242991_C_T_b38 | -0.15 | 0.00022 | ENSG00000212978.6 | AC016747.3 | Cells - Cultured fibroblasts |
| rs11125871 | 2p15 | chr2_61242991_C_T_b38 | 0.17 | 0.00023 | ENSG00000212978.6 | AC016747.3 | Esophagus - Muscularis |
| rs11264280 | 1q21.3 | chr1_154890476_C_T_b38 | 0.16 | 9.60E-08 | ENSG00000163354.14 | DCST2 | Thyroid |
| rs11264280 | 1q21.3 | chr1_154890476_C_T_b38 | 0.33 | 1.30E-07 | ENSG00000232093.1 | DCST1-AS1 | Testis |

| | | | | | | | |
|---|---|---|---|---|---|---|---|
| rs11264280 | 1q21.3 | chr1_154890476_C_T_b38 | 0.12 | 2.70E-06 | ENSG00000185499.16 | MUC1 | Cells - Cultured fibroblasts |
| rs11264280 | 1q21.3 | chr1_154890476_C_T_b38 | 0.22 | 4.30E-06 | ENSG00000143603.18 | KCNN3 | Brain - Hypothalamus |
| rs11264280 | 1q21.3 | chr1_154890476_C_T_b38 | 0.17 | 8.50E-06 | ENSG00000143603.18 | KCNN3 | Lung |
| rs11264280 | 1q21.3 | chr1_154890476_C_T_b38 | 0.28 | 1.30E-05 | ENSG00000163354.14 | DCST2 | Pituitary |
| rs11264280 | 1q21.3 | chr1_154890476_C_T_b38 | 0.14 | 1.30E-05 | ENSG00000143603.18 | KCNN3 | Artery - Tibial |
| rs11264280 | 1q21.3 | chr1_154890476_C_T_b38 | 0.12 | 2.20E-05 | ENSG00000163346.16 | PBXIP1 | Skin - Not Sun Exposed (Suprapubic) |
| rs11264280 | 1q21.3 | chr1_154890476_C_T_b38 | 0.17 | 2.50E-05 | ENSG00000143603.18 | KCNN3 | Brain - Cerebellum |
| rs11264280 | 1q21.3 | chr1_154890476_C_T_b38 | 0.19 | 2.60E-05 | ENSG00000143603.18 | KCNN3 | Nerve - Tibial |
| rs11264280 | 1q21.3 | chr1_154890476_C_T_b38 | -0.12 | 3.50E-05 | ENSG00000143537.13 | ADAM15 | Whole Blood |
| rs11264280 | 1q21.3 | chr1_154890476_C_T_b38 | 0.089 | 3.60E-05 | ENSG00000163346.16 | PBXIP1 | Nerve - Tibial |
| rs11264280 | 1q21.3 | chr1_154890476_C_T_b38 | 0.31 | 4.80E-05 | ENSG00000163354.14 | DCST2 | Brain - Cortex |
| rs11264280 | 1q21.3 | chr1_154890476_C_T_b38 | -0.11 | 5.30E-05 | ENSG00000160741.16 | CRTC2 | Testis |
| rs11264280 | 1q21.3 | chr1_154890476_C_T_b38 | 0.14 | 6.60E-05 | ENSG00000185499.16 | MUC1 | Muscle - Skeletal |
| rs11264280 | 1q21.3 | chr1_154890476_C_T_b38 | 0.099 | 8.00E-05 | ENSG00000163346.16 | PBXIP1 | Esophagus - Mucosa |
| rs11264280 | 1q21.3 | chr1_154890476_C_T_b38 | -0.15 | 8.90E-05 | ENSG00000177628.15 | GBA | Esophagus - Mucosa |
| rs11264280 | 1q21.3 | chr1_154890476_C_T_b38 | 0.097 | 9.00E-05 | ENSG00000163346.16 | PBXIP1 | Adipose - Subcutaneous |
| rs11264280 | 1q21.3 | chr1_154890476_C_T_b38 | 0.12 | 9.40E-05 | ENSG00000232093.1 | DCST1-AS1 | Skin - Not Sun Exposed (Suprapubic) |
| rs11264280 | 1q21.3 | chr1_154890476_C_T_b38 | -0.13 | 0.00011 | ENSG00000203761.5 | MSTO2P | Muscle - Skeletal |
| rs11264280 | 1q21.3 | chr1_154890476_C_T_b38 | 0.12 | 0.00017 | ENSG00000163354.14 | DCST2 | Nerve - Tibial |
| rs11264280 | 1q21.3 | chr1_154890476_C_T_b38 | 0.15 | 2.00E-04 | ENSG00000143603.18 | KCNN3 | Thyroid |
| rs11264280 | 1q21.3 | chr1_154890476_C_T_b38 | -0.097 | 0.00034 | ENSG00000143537.13 | ADAM15 | Cells - Cultured fibroblasts |
| rs11264280 | 1q21.3 | chr1_154890476_C_T_b38 | 0.067 | 0.00034 | ENSG00000163346.16 | PBXIP1 | Cells - Cultured fibroblasts |
| rs11598047 | 10q24.33 | chr10_103582915_A_G_b38 | -0.46 | 1.40E-11 | ENSG00000107954.10 | NEURL1 | Cells - Cultured fibroblasts |
| rs11598047 | 10q24.33 | chr10_103582915_A_G_b38 | -0.23 | 0.00021 | ENSG00000107954.10 | NEURL1 | Skin - Sun Exposed (Lower leg) |
| rs116600817 | 17p13.1 | chr17_7555478_A_G_b38 | 0.36 | 3.20E-45 | ENSG00000239697.10 | TNFSF12 | Cells - Cultured fibroblasts |
| rs116600817 | 17p13.1 | chr17_7555478_A_G_b38 | -0.45 | 9.90E-41 | ENSG00000170175.10 | CHRNB1 | Muscle - Skeletal |
| rs116600817 | 17p13.1 | chr17_7555478_A_G_b38 | 0.33 | 3.60E-31 | ENSG00000239697.10 | TNFSF12 | Whole Blood |
| rs116600817 | 17p13.1 | chr17_7555478_A_G_b38 | 0.29 | 3.50E-24 | ENSG00000161955.16 | TNFSF13 | Cells - Cultured fibroblasts |
| rs116600817 | 17p13.1 | chr17_7555478_A_G_b38 | -0.22 | 3.20E-14 | ENSG00000141504.11 | SAT2 | Muscle - Skeletal |
| rs116600817 | 17p13.1 | chr17_7555478_A_G_b38 | 0.25 | 5.40E-08 | ENSG00000239697.10 | TNFSF12 | Spleen |
| rs116600817 | 17p13.1 | chr17_7555478_A_G_b38 | 0.34 | 8.00E-08 | ENSG00000239697.10 | TNFSF12 | Brain - Spinal cord (cervical c-1) |
| rs116600817 | 17p13.1 | chr17_7555478_A_G_b38 | -0.25 | 1.60E-07 | ENSG00000170175.10 | CHRNB1 | Esophagus - Mucosa |
| rs116600817 | 17p13.1 | chr17_7555478_A_G_b38 | 0.2 | 1.70E-07 | ENSG00000239697.10 | TNFSF12 | Brain - Cerebellar Hemisphere |
| rs116600817 | 17p13.1 | chr17_7555478_A_G_b38 | 0.12 | 3.20E-07 | ENSG00000239697.10 | TNFSF12 | Adipose - Visceral (Omentum) |
| rs116600817 | 17p13.1 | chr17_7555478_A_G_b38 | 0.13 | 1.20E-06 | ENSG00000161955.16 | TNFSF13 | Adipose - Subcutaneous |
| rs116600817 | 17p13.1 | chr17_7555478_A_G_b38 | -0.15 | 1.50E-06 | ENSG00000141504.11 | SAT2 | Esophagus - Mucosa |
| rs116600817 | 17p13.1 | chr17_7555478_A_G_b38 | 0.099 | 2.60E-06 | ENSG00000239697.10 | TNFSF12 | Nerve - Tibial |
| rs116600817 | 17p13.1 | chr17_7555478_A_G_b38 | 0.12 | 4.00E-06 | ENSG00000239697.10 | TNFSF12 | Esophagus - Muscularis |
| rs116600817 | 17p13.1 | chr17_7555478_A_G_b38 | 0.21 | 7.80E-06 | ENSG00000174282.11 | ZBTB4 | Testis |
| rs116600817 | 17p13.1 | chr17_7555478_A_G_b38 | 0.21 | 8.10E-06 | ENSG00000239697.10 | TNFSF12 | Brain - Hypothalamus |
| rs116600817 | 17p13.1 | chr17_7555478_A_G_b38 | -0.25 | 1.60E-05 | ENSG00000170175.10 | CHRNB1 | Brain - Cerebellar Hemisphere |
| rs116600817 | 17p13.1 | chr17_7555478_A_G_b38 | -0.082 | 1.60E-05 | ENSG00000141504.11 | SAT2 | Cells - Cultured fibroblasts |
| rs116600817 | 17p13.1 | chr17_7555478_A_G_b38 | 0.2 | 1.90E-05 | ENSG00000239697.10 | TNFSF12 | Brain - Cerebellum |
| rs116600817 | 17p13.1 | chr17_7555478_A_G_b38 | -0.08 | 2.10E-05 | ENSG00000141504.11 | SAT2 | Whole Blood |
| rs116600817 | 17p13.1 | chr17_7555478_A_G_b38 | -0.11 | 2.40E-05 | ENSG00000129226.13 | CD68 | Esophagus - Mucosa |
| rs116600817 | 17p13.1 | chr17_7555478_A_G_b38 | -0.16 | 2.90E-05 | ENSG00000141504.11 | SAT2 | Stomach |
| rs116600817 | 17p13.1 | chr17_7555478_A_G_b38 | -0.18 | 2.90E-05 | ENSG00000170175.10 | CHRNB1 | Heart - Atrial Appendage |
| rs116600817 | 17p13.1 | chr17_7555478_A_G_b38 | -0.16 | 3.10E-05 | ENSG00000141504.11 | SAT2 | Heart - Atrial Appendage |
| rs116600817 | 17p13.1 | chr17_7555478_A_G_b38 | -0.13 | 4.50E-05 | ENSG00000141504.11 | SAT2 | Esophagus - Muscularis |
| rs116600817 | 17p13.1 | chr17_7555478_A_G_b38 | -0.12 | 6.00E-05 | ENSG00000161956.12 | SENP3 | Nerve - Tibial |
| rs116600817 | 17p13.1 | chr17_7555478_A_G_b38 | 0.19 | 8.00E-05 | ENSG00000181885.18 | CLDN7 | Cells - Cultured fibroblasts |

| | | | | | | | |
|---|---|---|---|---|---|---|---|
| rs116600817 | 17p13.1 | chr17_7555478_A_G_b38 | -0.095 | 9.00E-05 | ENSG00000161956.12 | SENP3 | Skin - Not Sun Exposed (Suprapubic) |
| rs11782313 | 8q24.3 | chr8_140954084_C_T_b38 | 0.38 | 3.80E-13 | ENSG00000169398.19 | PTK2 | Brain - Cerebellum |
| rs11782313 | 8q24.3 | chr8_140954084_C_T_b38 | 0.24 | 4.50E-13 | ENSG00000169398.19 | PTK2 | Brain - Caudate (basal ganglia) |
| rs11782313 | 8q24.3 | chr8_140954084_C_T_b38 | 0.25 | 2.90E-09 | ENSG00000169398.19 | PTK2 | Brain - Nucleus accumbens (basal ganglia) |
| rs11782313 | 8q24.3 | chr8_140954084_C_T_b38 | -0.29 | 4.10E-09 | ENSG00000123908.11 | Aug-02 | Testis |
| rs11782313 | 8q24.3 | chr8_140954084_C_T_b38 | -0.26 | 7.60E-09 | ENSG00000280303.2 | ERICD | Artery - Tibial |
| rs11782313 | 8q24.3 | chr8_140954084_C_T_b38 | 0.32 | 1.70E-08 | ENSG00000169398.19 | PTK2 | Brain - Anterior cingulate cortex (BA24) |
| rs11782313 | 8q24.3 | chr8_140954084_C_T_b38 | 0.24 | 8.90E-08 | ENSG00000169398.19 | PTK2 | Brain - Putamen (basal ganglia) |
| rs11782313 | 8q24.3 | chr8_140954084_C_T_b38 | -0.34 | 3.50E-07 | ENSG00000280303.2 | ERICD | Testis |
| rs11782313 | 8q24.3 | chr8_140954084_C_T_b38 | 0.2 | 3.60E-07 | ENSG00000169398.19 | PTK2 | Colon - Sigmoid |
| rs11782313 | 8q24.3 | chr8_140954084_C_T_b38 | 0.26 | 4.60E-07 | ENSG00000169398.19 | PTK2 | Brain - Amygdala |
| rs11782313 | 8q24.3 | chr8_140954084_C_T_b38 | -0.24 | 2.20E-06 | ENSG00000280303.2 | ERICD | Cells - Cultured fibroblasts |
| rs11782313 | 8q24.3 | chr8_140954084_C_T_b38 | -0.18 | 3.00E-06 | ENSG00000279766.1 | RP11-642A1.2 | Nerve - Tibial |
| rs11782313 | 8q24.3 | chr8_140954084_C_T_b38 | -0.15 | 3.70E-06 | ENSG00000169398.19 | PTK2 | Skin - Not Sun Exposed (Suprapubic) |
| rs11782313 | 8q24.3 | chr8_140954084_C_T_b38 | 0.19 | 7.30E-06 | ENSG00000169398.19 | PTK2 | Brain - Cortex |
| rs11782313 | 8q24.3 | chr8_140954084_C_T_b38 | -0.13 | 1.40E-05 | ENSG00000279766.1 | RP11-642A1.2 | Adipose - Visceral (Omentum) |
| rs11782313 | 8q24.3 | chr8_140954084_C_T_b38 | 0.3 | 1.60E-05 | ENSG00000169398.19 | PTK2 | Brain - Cerebellar Hemisphere |
| rs11782313 | 8q24.3 | chr8_140954084_C_T_b38 | 0.11 | 6.70E-05 | ENSG00000169398.19 | PTK2 | Nerve - Tibial |
| rs12211255 | 6q14.1 | chr6_75478614_C_A_b38 | 0.15 | 7.50E-08 | ENSG00000112701.17 | SENP6 | Whole Blood |
| rs12211255 | 6q14.1 | chr6_75478614_C_A_b38 | -0.22 | 5.50E-07 | ENSG00000118407.14 | FILIP1 | Muscle - Skeletal |
| rs12211255 | 6q14.1 | chr6_75478614_C_A_b38 | 0.11 | 7.50E-05 | ENSG00000112701.17 | SENP6 | Artery - Tibial |
| rs12211255 | 6q14.1 | chr6_75478614_C_A_b38 | -0.41 | 8.00E-05 | ENSG00000196586.13 | MYO6 | Brain - Cerebellar Hemisphere |
| rs12211255 | 6q14.1 | chr6_75478614_C_A_b38 | 0.16 | 1.00E-04 | ENSG00000112701.17 | SENP6 | Cells - Cultured fibroblasts |
| rs12260801 | 10q25.2 | chr10_110814644_C_T_b38 | 0.17 | 2.50E-06 | ENSG00000214413.7 | BBIP1 | Thyroid |
| rs12260801 | 10q25.2 | chr10_110814644_C_T_b38 | -0.15 | 9.20E-06 | ENSG00000203867.7 | RBM20 | Esophagus - Mucosa |
| rs12908004 | 15q25.1 | chr15_80384583_A_G_b38 | 0.86 | 1.80E-54 | ENSG00000172379.20 | ARNT2 | Heart - Atrial Appendage |
| rs12908004 | 15q25.1 | chr15_80384583_A_G_b38 | 0.87 | 4.80E-43 | ENSG00000259495.2 | RP11-210M15.2 | Heart - Atrial Appendage |
| rs12908004 | 15q25.1 | chr15_80384583_A_G_b38 | 0.93 | 6.40E-29 | ENSG00000259417.2 | LINC01314 | Heart - Atrial Appendage |
| rs12908004 | 15q25.1 | chr15_80384583_A_G_b38 | 0.31 | 9.70E-13 | ENSG00000259495.2 | RP11-210M15.2 | Muscle - Skeletal |
| rs12908004 | 15q25.1 | chr15_80384583_A_G_b38 | 0.31 | 1.70E-06 | ENSG00000172379.20 | ARNT2 | Heart - Left Ventricle |
| rs12908004 | 15q25.1 | chr15_80384583_A_G_b38 | 0.33 | 1.50E-05 | ENSG00000259495.2 | RP11-210M15.2 | Heart - Left Ventricle |
| rs13191450 | 6q22.31 | chr6_122070990_A_C_b38 | -0.13 | 1.00E-04 | ENSG00000279453.1 | RP3-425C14.4 | Thyroid |
| rs17171711 | 5q31.2 | chr5_138029106_C_T_b38 | -0.66 | 7.30E-15 | ENSG00000246323.2 | RP11-325L7.1 | Heart - Left Ventricle |
| rs17171711 | 5q31.2 | chr5_138029106_C_T_b38 | -0.3 | 3.00E-11 | ENSG00000031003.10 | FAM13B | Heart - Atrial Appendage |
| rs17171711 | 5q31.2 | chr5_138029106_C_T_b38 | -0.44 | 4.30E-09 | ENSG00000250260.1 | RP11-325L7.2 | Heart - Atrial Appendage |
| rs17171711 | 5q31.2 | chr5_138029106_C_T_b38 | -0.24 | 1.60E-08 | ENSG00000146021.14 | KLHL3 | Skin - Not Sun Exposed (Suprapubic) |
| rs17171711 | 5q31.2 | chr5_138029106_C_T_b38 | -0.56 | 4.30E-08 | ENSG00000146013.10 | GFRA3 | Pituitary |
| rs17171711 | 5q31.2 | chr5_138029106_C_T_b38 | -0.41 | 1.40E-07 | ENSG00000246323.2 | RP11-325L7.1 | Heart - Atrial Appendage |
| rs17171711 | 5q31.2 | chr5_138029106_C_T_b38 | -0.23 | 2.10E-07 | ENSG00000146021.14 | KLHL3 | Esophagus - Mucosa |
| rs17171711 | 5q31.2 | chr5_138029106_C_T_b38 | -0.2 | 5.00E-07 | ENSG00000031003.10 | FAM13B | Heart - Left Ventricle |
| rs17171711 | 5q31.2 | chr5_138029106_C_T_b38 | -0.16 | 2.00E-06 | ENSG00000146021.14 | KLHL3 | Skin - Sun Exposed (Lower leg) |
| rs17171711 | 5q31.2 | chr5_138029106_C_T_b38 | -0.3 | 3.00E-06 | ENSG00000132563.15 | REEP2 | Heart - Atrial Appendage |
| rs17171711 | 5q31.2 | chr5_138029106_C_T_b38 | 0.27 | 5.00E-06 | ENSG00000146021.14 | KLHL3 | Brain - Cerebellum |
| rs17171711 | 5q31.2 | chr5_138029106_C_T_b38 | -0.37 | 1.00E-04 | ENSG00000253404.1 | AC034243.1 | Esophagus - Muscularis |
| rs17171711 | 5q31.2 | chr5_138029106_C_T_b38 | 0.14 | 0.00017 | ENSG00000094880.10 | CDC23 | Testis |
| rs17171711 | 5q31.2 | chr5_138029106_C_T_b38 | 0.22 | 0.00019 | ENSG00000112984.11 | KIF20A | Testis |
| rs2269001 | 7q36.1 | chr7_150954728_G_A_b38 | -0.29 | 9.90E-06 | ENSG00000055118.14 | KCNH2 | Adrenal Gland |
| rs2269001 | 7q36.1 | chr7_150954728_G_A_b38 | -0.15 | 9.40E-05 | ENSG00000055118.14 | KCNH2 | Artery - Tibial |
| rs2738413 | 14q23.2 | chr14_64213242_A_G_b38 | -0.6 | 5.00E-48 | ENSG00000140009.18 | ESR2 | Skin - Sun Exposed (Lower leg) |
| rs2738413 | 14q23.2 | chr14_64213242_A_G_b38 | -0.25 | 1.00E-22 | ENSG00000100714.15 | MTHFD1 | Nerve - Tibial |
| rs2738413 | 14q23.2 | chr14_64213242_A_G_b38 | -0.25 | 8.20E-19 | ENSG00000100714.15 | MTHFD1 | Thyroid |

| | | | | | | | |
|---|---|---|---|---|---|---|---|
| rs2738413 | 14q23.2 | chr14_64213242_A_G_b38 | -0.28 | 2.30E-16 | ENSG00000100714.15 | MTHFD1 | Skin - Not Sun Exposed (Suprapubic) |
| rs2738413 | 14q23.2 | chr14_64213242_A_G_b38 | -0.26 | 3.60E-16 | ENSG00000100714.15 | MTHFD1 | Skin - Sun Exposed (Lower leg) |
| rs2738413 | 14q23.2 | chr14_64213242_A_G_b38 | -0.22 | 2.30E-15 | ENSG00000100714.15 | MTHFD1 | Artery - Tibial |
| rs2738413 | 14q23.2 | chr14_64213242_A_G_b38 | 0.21 | 7.10E-15 | ENSG00000054654.16 | SYNE2 | Heart - Left Ventricle |
| rs2738413 | 14q23.2 | chr14_64213242_A_G_b38 | -0.28 | 1.20E-14 | ENSG00000140009.18 | ESR2 | Skin - Not Sun Exposed (Suprapubic) |
| rs2738413 | 14q23.2 | chr14_64213242_A_G_b38 | -0.16 | 3.40E-14 | ENSG00000100714.15 | MTHFD1 | Lung |
| rs2738413 | 14q23.2 | chr14_64213242_A_G_b38 | -0.12 | 3.00E-11 | ENSG00000100714.15 | MTHFD1 | Adipose - Subcutaneous |
| rs2738413 | 14q23.2 | chr14_64213242_A_G_b38 | -0.23 | 1.40E-10 | ENSG00000140009.18 | ESR2 | Whole Blood |
| rs2738413 | 14q23.2 | chr14_64213242_A_G_b38 | -0.26 | 1.20E-09 | ENSG00000100714.15 | MTHFD1 | Pancreas |
| rs2738413 | 14q23.2 | chr14_64213242_A_G_b38 | -0.15 | 2.10E-08 | ENSG00000100714.15 | MTHFD1 | Breast - Mammary Tissue |
| rs2738413 | 14q23.2 | chr14_64213242_A_G_b38 | -0.25 | 2.10E-08 | ENSG00000140009.18 | ESR2 | Lung |
| rs2738413 | 14q23.2 | chr14_64213242_A_G_b38 | -0.22 | 1.80E-07 | ENSG00000100714.15 | MTHFD1 | Artery - Aorta |
| rs2738413 | 14q23.2 | chr14_64213242_A_G_b38 | -0.096 | 2.30E-07 | ENSG00000100714.15 | MTHFD1 | Adipose - Visceral (Omentum) |
| rs2738413 | 14q23.2 | chr14_64213242_A_G_b38 | -0.21 | 3.00E-07 | ENSG00000140009.18 | ESR2 | Thyroid |
| rs2738413 | 14q23.2 | chr14_64213242_A_G_b38 | -0.16 | 5.10E-07 | ENSG00000100714.15 | MTHFD1 | Esophagus - Gastroesophageal Junction |
| rs2738413 | 14q23.2 | chr14_64213242_A_G_b38 | -0.073 | 5.40E-07 | ENSG00000100714.15 | MTHFD1 | Esophagus - Mucosa |
| rs2738413 | 14q23.2 | chr14_64213242_A_G_b38 | 0.13 | 6.30E-07 | ENSG00000054654.16 | SYNE2 | Heart - Atrial Appendage |
| rs2738413 | 14q23.2 | chr14_64213242_A_G_b38 | -0.24 | 7.10E-07 | ENSG00000100714.15 | MTHFD1 | Prostate |
| rs2738413 | 14q23.2 | chr14_64213242_A_G_b38 | -0.27 | 1.60E-06 | ENSG00000100714.15 | MTHFD1 | Ovary |
| rs2738413 | 14q23.2 | chr14_64213242_A_G_b38 | -0.3 | 2.40E-06 | ENSG00000140009.18 | ESR2 | Pancreas |
| rs2738413 | 14q23.2 | chr14_64213242_A_G_b38 | -0.31 | 2.70E-06 | ENSG00000140009.18 | ESR2 | Spleen |
| rs2738413 | 14q23.2 | chr14_64213242_A_G_b38 | -0.16 | 3.30E-06 | ENSG00000100714.15 | MTHFD1 | Stomach |
| rs2738413 | 14q23.2 | chr14_64213242_A_G_b38 | -0.12 | 3.70E-06 | ENSG00000100714.15 | MTHFD1 | Testis |
| rs2738413 | 14q23.2 | chr14_64213242_A_G_b38 | -0.24 | 5.00E-06 | ENSG00000140009.18 | ESR2 | Nerve - Tibial |
| rs2738413 | 14q23.2 | chr14_64213242_A_G_b38 | -0.11 | 6.20E-06 | ENSG00000100714.15 | MTHFD1 | Colon - Transverse |
| rs2738413 | 14q23.2 | chr14_64213242_A_G_b38 | -0.14 | 1.50E-05 | ENSG00000140009.18 | ESR2 | Muscle - Skeletal |
| rs2738413 | 14q23.2 | chr14_64213242_A_G_b38 | -0.076 | 4.00E-05 | ENSG00000126822.16 | PLEKHG3 | Artery - Tibial |
| rs2738413 | 14q23.2 | chr14_64213242_A_G_b38 | -0.13 | 0.00017 | ENSG00000272909.1 | CTD-2555O16.4 | Thyroid |
| rs2857265 | 2q31.2 | chr2_178534096_G_A_b38 | 0.21 | 1.90E-11 | ENSG00000079150.17 | FKBP7 | Cells - Cultured fibroblasts |
| rs2857265 | 2q31.2 | chr2_178534096_G_A_b38 | 0.37 | 7.50E-11 | ENSG00000079150.17 | FKBP7 | Artery - Aorta |
| rs2857265 | 2q31.2 | chr2_178534096_G_A_b38 | 0.21 | 4.40E-10 | ENSG00000079150.17 | FKBP7 | Adipose - Subcutaneous |
| rs2857265 | 2q31.2 | chr2_178534096_G_A_b38 | 0.24 | 3.80E-09 | ENSG00000079150.17 | FKBP7 | Nerve - Tibial |
| rs2857265 | 2q31.2 | chr2_178534096_G_A_b38 | 0.23 | 6.90E-09 | ENSG00000079150.17 | FKBP7 | Thyroid |
| rs2857265 | 2q31.2 | chr2_178534096_G_A_b38 | 0.26 | 1.30E-08 | ENSG00000079150.17 | FKBP7 | Artery - Tibial |
| rs2857265 | 2q31.2 | chr2_178534096_G_A_b38 | -0.23 | 4.10E-08 | ENSG00000116095.10 | PLEKHA3 | Cells - Cultured fibroblasts |
| rs2857265 | 2q31.2 | chr2_178534096_G_A_b38 | 0.24 | 3.10E-06 | ENSG00000079150.17 | FKBP7 | Esophagus - Gastroesophageal Junction |
| rs2857265 | 2q31.2 | chr2_178534096_G_A_b38 | -0.66 | 1.20E-05 | ENSG00000237298.9 | TTN-AS1 | Brain - Cerebellar Hemisphere |
| rs2857265 | 2q31.2 | chr2_178534096_G_A_b38 | -0.19 | 2.20E-05 | ENSG00000237298.9 | TTN-AS1 | Whole Blood |
| rs2857265 | 2q31.2 | chr2_178534096_G_A_b38 | -0.33 | 3.00E-05 | ENSG00000267784.1 | RP11-171I2.1 | Thyroid |
| rs2857265 | 2q31.2 | chr2_178534096_G_A_b38 | -0.59 | 3.00E-05 | ENSG00000237298.9 | TTN-AS1 | Brain - Cerebellum |
| rs2857265 | 2q31.2 | chr2_178534096_G_A_b38 | 0.24 | 5.40E-05 | ENSG00000079150.17 | FKBP7 | Colon - Sigmoid |
| rs2857265 | 2q31.2 | chr2_178534096_G_A_b38 | 0.12 | 9.60E-05 | ENSG00000079150.17 | FKBP7 | Skin - Sun Exposed (Lower leg) |
| rs2857265 | 2q31.2 | chr2_178534096_G_A_b38 | 0.17 | 9.90E-05 | ENSG00000079150.17 | FKBP7 | Heart - Atrial Appendage |
| rs2857265 | 2q31.2 | chr2_178534096_G_A_b38 | -0.33 | 0.00016 | ENSG00000271401.1 | RP11-171I2.3 | Lung |
| rs2857265 | 2q31.2 | chr2_178534096_G_A_b38 | 0.17 | 0.00022 | ENSG00000079150.17 | FKBP7 | Esophagus - Muscularis |
| rs2885697 | 1p34.2 | chr1_41078607_G_T_b38 | 0.33 | 2.20E-23 | ENSG00000010803.16 | SCMH1 | Nerve - Tibial |
| rs2885697 | 1p34.2 | chr1_41078607_G_T_b38 | 0.24 | 1.50E-14 | ENSG00000010803.16 | SCMH1 | Artery - Aorta |
| rs2885697 | 1p34.2 | chr1_41078607_G_T_b38 | -0.31 | 4.10E-11 | ENSG00000281207.1 | SLFNL1-AS1 | Testis |
| rs2885697 | 1p34.2 | chr1_41078607_G_T_b38 | 0.14 | 4.50E-10 | ENSG00000010803.16 | SCMH1 | Esophagus - Muscularis |
| rs2885697 | 1p34.2 | chr1_41078607_G_T_b38 | 0.14 | 4.50E-10 | ENSG00000010803.16 | SCMH1 | Artery - Tibial |
| rs2885697 | 1p34.2 | chr1_41078607_G_T_b38 | -0.21 | 9.60E-10 | ENSG00000010803.16 | SCMH1 | Testis |

| | | | | | | | |
|---|---|---|---|---|---|---|---|
| rs2885697 | 1p34.2 | chr1_41078607_G_T_b38 | -0.4 | 8.00E-09 | ENSG00000204060.6 | FOXO6 | Adrenal Gland |
| rs2885697 | 1p34.2 | chr1_41078607_G_T_b38 | -0.47 | 8.70E-09 | ENSG00000204060.6 | FOXO6 | Brain - Cerebellum |
| rs2885697 | 1p34.2 | chr1_41078607_G_T_b38 | 0.18 | 1.40E-08 | ENSG00000010803.16 | SCMH1 | Heart - Atrial Appendage |
| rs2885697 | 1p34.2 | chr1_41078607_G_T_b38 | -0.21 | 5.10E-08 | ENSG00000281207.1 | SLFNL1-AS1 | Thyroid |
| rs2885697 | 1p34.2 | chr1_41078607_G_T_b38 | -0.42 | 1.80E-06 | ENSG00000204060.6 | FOXO6 | Liver |
| rs2885697 | 1p34.2 | chr1_41078607_G_T_b38 | -0.28 | 2.40E-06 | ENSG00000204060.6 | FOXO6 | Cells - Cultured fibroblasts |
| rs2885697 | 1p34.2 | chr1_41078607_G_T_b38 | 0.14 | 5.70E-06 | ENSG00000010803.16 | SCMH1 | Colon - Sigmoid |
| rs2885697 | 1p34.2 | chr1_41078607_G_T_b38 | 0.093 | 1.10E-05 | ENSG00000010803.16 | SCMH1 | Colon - Transverse |
| rs2885697 | 1p34.2 | chr1_41078607_G_T_b38 | -0.18 | 1.40E-05 | ENSG00000281207.1 | SLFNL1-AS1 | Adipose - Visceral (Omentum) |
| rs2885697 | 1p34.2 | chr1_41078607_G_T_b38 | -0.14 | 3.80E-05 | ENSG00000171793.13 | CTPS1 | Heart - Atrial Appendage |
| rs2885697 | 1p34.2 | chr1_41078607_G_T_b38 | -0.18 | 4.40E-05 | ENSG00000281207.1 | SLFNL1-AS1 | Adipose - Subcutaneous |
| rs2885697 | 1p34.2 | chr1_41078607_G_T_b38 | 0.13 | 4.40E-05 | ENSG00000010803.16 | SCMH1 | Adipose - Visceral (Omentum) |
| rs2885697 | 1p34.2 | chr1_41078607_G_T_b38 | 0.21 | 5.10E-05 | ENSG00000235358.1 | RP11-399E6.1 | Testis |
| rs2885697 | 1p34.2 | chr1_41078607_G_T_b38 | 0.11 | 7.40E-05 | ENSG00000010803.16 | SCMH1 | Adipose - Subcutaneous |
| rs2885697 | 1p34.2 | chr1_41078607_G_T_b38 | -0.11 | 9.80E-05 | ENSG00000171790.15 | SLFNL1 | Testis |
| rs3176326 | 6p21.2 | chr6_36679512_G_A_b38 | -0.85 | 5.60E-08 | ENSG00000255587.7 | RAB44 | Cells - EBV-transformed lymphocytes |
| rs3176326 | 6p21.2 | chr6_36679512_G_A_b38 | -0.12 | 1.40E-05 | ENSG00000124762.13 | CDKN1A | Muscle - Skeletal |
| rs3746471 | 20q11.23 | chr20_38213512_G_A_b38 | -0.14 | 9.60E-06 | ENSG00000149633.11 | KIAA1755 | Skin - Sun Exposed (Lower leg) |
| rs3807989 | 7q31.2 | chr7_116546187_A_G_b38 | 0.2 | 5.30E-11 | ENSG00000243243.5 | AC073130.3 | Nerve - Tibial |
| rs3807989 | 7q31.2 | chr7_116546187_A_G_b38 | 0.21 | 3.00E-10 | ENSG00000237813.3 | AC002066.1 | Nerve - Tibial |
| rs3807989 | 7q31.2 | chr7_116546187_A_G_b38 | -0.24 | 8.00E-07 | ENSG00000105971.14 | CAV2 | Testis |
| rs3807989 | 7q31.2 | chr7_116546187_A_G_b38 | 0.18 | 1.10E-06 | ENSG00000237813.3 | AC002066.1 | Artery - Tibial |
| rs3807989 | 7q31.2 | chr7_116546187_A_G_b38 | -0.096 | 3.20E-06 | ENSG00000105971.14 | CAV2 | Skin - Sun Exposed (Lower leg) |
| rs3807989 | 7q31.2 | chr7_116546187_A_G_b38 | -0.13 | 4.20E-06 | ENSG00000105974.11 | CAV1 | Heart - Atrial Appendage |
| rs3807989 | 7q31.2 | chr7_116546187_A_G_b38 | -0.21 | 1.50E-05 | ENSG00000279086.1 | RP11-667F14.1 | Adipose - Subcutaneous |
| rs3807989 | 7q31.2 | chr7_116546187_A_G_b38 | -0.11 | 1.50E-05 | ENSG00000105971.14 | CAV2 | Heart - Atrial Appendage |
| rs3807989 | 7q31.2 | chr7_116546187_A_G_b38 | -0.21 | 2.10E-05 | ENSG00000279086.1 | RP11-667F14.1 | Thyroid |
| rs3807989 | 7q31.2 | chr7_116546187_A_G_b38 | -0.094 | 3.20E-05 | ENSG00000105971.14 | CAV2 | Skin - Not Sun Exposed (Suprapubic) |
| rs3807989 | 7q31.2 | chr7_116546187_A_G_b38 | 0.08 | 5.20E-05 | ENSG00000105974.11 | CAV1 | Nerve - Tibial |
| rs3807989 | 7q31.2 | chr7_116546187_A_G_b38 | 0.24 | 5.80E-05 | ENSG00000105971.14 | CAV2 | Pancreas |
| rs41292535 | 1p32.2 | chr1_51472967_G_A_b38 | -0.38 | 1.60E-08 | ENSG00000117862.11 | TXNDC12 | Whole Blood |
| rs41292535 | 1p32.2 | chr1_51472967_G_A_b38 | 0.91 | 1.20E-05 | ENSG00000203356.2 | LINC01562 | Adipose - Subcutaneous |
| rs41292535 | 1p32.2 | chr1_51472967_G_A_b38 | -1.7 | 1.80E-05 | ENSG00000233406.5 | RP11-296A18.5 | Brain - Hypothalamus |
| rs41292535 | 1p32.2 | chr1_51472967_G_A_b38 | -0.37 | 6.50E-05 | ENSG00000117862.11 | TXNDC12 | Skin - Not Sun Exposed (Suprapubic) |
| rs41292535 | 1p32.2 | chr1_51472967_G_A_b38 | 0.87 | 9.00E-05 | ENSG00000203356.2 | LINC01562 | Artery - Tibial |
| rs41292535 | 1p32.2 | chr1_51472967_G_A_b38 | -0.45 | 0.00016 | ENSG00000085832.16 | EPS15 | Artery - Tibial |
| rs41292535 | 1p32.2 | chr1_51472967_G_A_b38 | -0.28 | 0.00032 | ENSG00000117862.11 | TXNDC12 | Thyroid |
| rs4385527 | 9q22.32 | chr9_94886305_G_A_b38 | -0.13 | 2.30E-06 | ENSG00000148120.16 | C9orf3 | Heart - Atrial Appendage |
| rs4385527 | 9q22.32 | chr9_94886305_G_A_b38 | -0.16 | 2.20E-05 | ENSG00000148120.16 | C9orf3 | Skin - Sun Exposed (Lower leg) |
| rs4385527 | 9q22.32 | chr9_94886305_G_A_b38 | -0.21 | 2.50E-05 | ENSG00000158169.12 | FANCC | Brain - Cerebellum |
| rs55754224 | 4q26 | chr4_113507558_C_T_b38 | 0.12 | 4.60E-05 | ENSG00000145349.16 | CAMK2D | Heart - Atrial Appendage |
| rs55985730 | 7q32.1 | chr7_128776990_T_G_b38 | -0.34 | 6.50E-15 | ENSG00000128591.15 | FLNC | Cells - Cultured fibroblasts |
| rs55985730 | 7q32.1 | chr7_128776990_T_G_b38 | -0.47 | 7.60E-12 | ENSG00000128591.15 | FLNC | Nerve - Tibial |
| rs55985730 | 7q32.1 | chr7_128776990_T_G_b38 | -0.51 | 1.50E-09 | ENSG00000128591.15 | FLNC | Testis |
| rs55985730 | 7q32.1 | chr7_128776990_T_G_b38 | -0.31 | 9.40E-09 | ENSG00000128591.15 | FLNC | Thyroid |
| rs55985730 | 7q32.1 | chr7_128776990_T_G_b38 | -0.26 | 2.80E-08 | ENSG00000128591.15 | FLNC | Adipose - Subcutaneous |
| rs55985730 | 7q32.1 | chr7_128776990_T_G_b38 | -0.23 | 1.50E-07 | ENSG00000128591.15 | FLNC | Artery - Tibial |
| rs55985730 | 7q32.1 | chr7_128776990_T_G_b38 | -0.56 | 2.90E-06 | ENSG00000128591.15 | FLNC | Ovary |
| rs55985730 | 7q32.1 | chr7_128776990_T_G_b38 | -0.37 | 1.00E-05 | ENSG00000128591.15 | FLNC | Breast - Mammary Tissue |
| rs55985730 | 7q32.1 | chr7_128776990_T_G_b38 | -0.3 | 9.50E-05 | ENSG00000229413.2 | RP11-274B21.12 | Thyroid |
| rs55985730 | 7q32.1 | chr7_128776990_T_G_b38 | 0.38 | 0.00021 | ENSG00000224940.8 | PRRT4 | Skin - Not Sun Exposed (Suprapubic) |

| | | | | | | | |
|---|---|---|---|---|---|---|---|
| rs56181519 | 2q31.1 | chr2_174690986_C_T_b38 | 0.25 | 3.90E-17 | ENSG00000115935.17 | WIPF1 | Heart - Left Ventricle |
| rs56181519 | 2q31.1 | chr2_174690986_C_T_b38 | 0.17 | 4.50E-13 | ENSG00000115935.17 | WIPF1 | Cells - Cultured fibroblasts |
| rs56181519 | 2q31.1 | chr2_174690986_C_T_b38 | 0.25 | 2.80E-10 | ENSG00000163328.13 | GPR155 | Thyroid |
| rs56181519 | 2q31.1 | chr2_174690986_C_T_b38 | 0.13 | 2.60E-09 | ENSG00000115935.17 | WIPF1 | Adipose - Visceral (Omentum) |
| rs56181519 | 2q31.1 | chr2_174690986_C_T_b38 | 0.17 | 5.90E-07 | ENSG00000115935.17 | WIPF1 | Esophagus - Muscularis |
| rs56181519 | 2q31.1 | chr2_174690986_C_T_b38 | 0.14 | 6.50E-07 | ENSG00000115935.17 | WIPF1 | Heart - Atrial Appendage |
| rs56181519 | 2q31.1 | chr2_174690986_C_T_b38 | 0.19 | 1.80E-06 | ENSG00000115935.17 | WIPF1 | Esophagus - Gastroesophageal Junction |
| rs56181519 | 2q31.1 | chr2_174690986_C_T_b38 | 0.097 | 1.30E-05 | ENSG00000115935.17 | WIPF1 | Adipose - Subcutaneous |
| rs56181519 | 2q31.1 | chr2_174690986_C_T_b38 | 0.23 | 1.50E-05 | ENSG00000279884.1 | RP11-483E17.1 | Spleen |
| rs56181519 | 2q31.1 | chr2_174690986_C_T_b38 | 0.11 | 3.50E-05 | ENSG00000115935.17 | WIPF1 | Stomach |
| rs635634 | 9q34.2 | chr9_133279427_T_C_b38 | 0.76 | 3.60E-34 | ENSG00000175164.13 | ABO | Whole Blood |
| rs635634 | 9q34.2 | chr9_133279427_T_C_b38 | -0.55 | 3.40E-24 | ENSG00000175164.13 | ABO | Adipose - Visceral (Omentum) |
| rs635634 | 9q34.2 | chr9_133279427_T_C_b38 | -0.52 | 8.20E-24 | ENSG00000175164.13 | ABO | Adipose - Subcutaneous |
| rs635634 | 9q34.2 | chr9_133279427_T_C_b38 | -0.46 | 9.20E-18 | ENSG00000175164.13 | ABO | Muscle - Skeletal |
| rs635634 | 9q34.2 | chr9_133279427_T_C_b38 | -0.29 | 7.70E-16 | ENSG00000175164.13 | ABO | Artery - Tibial |
| rs635634 | 9q34.2 | chr9_133279427_T_C_b38 | -0.38 | 1.10E-13 | ENSG00000175164.13 | ABO | Heart - Left Ventricle |
| rs635634 | 9q34.2 | chr9_133279427_T_C_b38 | -0.4 | 1.80E-12 | ENSG00000175164.13 | ABO | Esophagus - Muscularis |
| rs635634 | 9q34.2 | chr9_133279427_T_C_b38 | 0.37 | 2.20E-12 | ENSG00000148290.9 | SURF1 | Heart - Atrial Appendage |
| rs635634 | 9q34.2 | chr9_133279427_T_C_b38 | -0.36 | 1.20E-11 | ENSG00000175164.13 | ABO | Esophagus - Mucosa |
| rs635634 | 9q34.2 | chr9_133279427_T_C_b38 | -0.58 | 9.40E-11 | ENSG00000175164.13 | ABO | Adrenal Gland |
| rs635634 | 9q34.2 | chr9_133279427_T_C_b38 | 0.35 | 1.70E-10 | ENSG00000148290.9 | SURF1 | Esophagus - Gastroesophageal Junction |
| rs635634 | 9q34.2 | chr9_133279427_T_C_b38 | 0.27 | 2.20E-10 | ENSG00000148290.9 | SURF1 | Adipose - Subcutaneous |
| rs635634 | 9q34.2 | chr9_133279427_T_C_b38 | -0.4 | 4.30E-10 | ENSG00000175164.13 | ABO | Pituitary |
| rs635634 | 9q34.2 | chr9_133279427_T_C_b38 | -0.44 | 6.60E-10 | ENSG00000175164.13 | ABO | Breast - Mammary Tissue |
| rs635634 | 9q34.2 | chr9_133279427_T_C_b38 | 0.34 | 1.20E-09 | ENSG00000148290.9 | SURF1 | Colon - Transverse |
| rs635634 | 9q34.2 | chr9_133279427_T_C_b38 | 0.24 | 1.70E-09 | ENSG00000148290.9 | SURF1 | Adipose - Visceral (Omentum) |
| rs635634 | 9q34.2 | chr9_133279427_T_C_b38 | -0.68 | 3.60E-09 | ENSG00000175164.13 | ABO | Liver |
| rs635634 | 9q34.2 | chr9_133279427_T_C_b38 | 0.31 | 4.50E-09 | ENSG00000148290.9 | SURF1 | Heart - Left Ventricle |
| rs635634 | 9q34.2 | chr9_133279427_T_C_b38 | 0.31 | 7.20E-09 | ENSG00000148290.9 | SURF1 | Esophagus - Muscularis |
| rs635634 | 9q34.2 | chr9_133279427_T_C_b38 | -0.29 | 7.60E-09 | ENSG00000175164.13 | ABO | Colon - Sigmoid |
| rs635634 | 9q34.2 | chr9_133279427_T_C_b38 | 0.24 | 1.60E-08 | ENSG00000148290.9 | SURF1 | Muscle - Skeletal |
| rs635634 | 9q34.2 | chr9_133279427_T_C_b38 | -0.37 | 2.50E-08 | ENSG00000175164.13 | ABO | Heart - Atrial Appendage |
| rs635634 | 9q34.2 | chr9_133279427_T_C_b38 | -0.56 | 6.80E-08 | ENSG00000175164.13 | ABO | Spleen |
| rs635634 | 9q34.2 | chr9_133279427_T_C_b38 | -0.29 | 7.50E-08 | ENSG00000175164.13 | ABO | Nerve - Tibial |
| rs635634 | 9q34.2 | chr9_133279427_T_C_b38 | -0.29 | 1.20E-07 | ENSG00000175164.13 | ABO | Artery - Coronary |
| rs635634 | 9q34.2 | chr9_133279427_T_C_b38 | 0.2 | 1.50E-07 | ENSG00000148290.9 | SURF1 | Lung |
| rs635634 | 9q34.2 | chr9_133279427_T_C_b38 | 0.46 | 1.80E-07 | ENSG00000148290.9 | SURF1 | Brain - Cerebellum |
| rs635634 | 9q34.2 | chr9_133279427_T_C_b38 | 0.23 | 2.50E-07 | ENSG00000148290.9 | SURF1 | Breast - Mammary Tissue |
| rs635634 | 9q34.2 | chr9_133279427_T_C_b38 | 0.23 | 3.90E-07 | ENSG00000148290.9 | SURF1 | Skin - Not Sun Exposed (Suprapubic) |
| rs635634 | 9q34.2 | chr9_133279427_T_C_b38 | 0.21 | 6.70E-07 | ENSG00000148290.9 | SURF1 | Nerve - Tibial |
| rs635634 | 9q34.2 | chr9_133279427_T_C_b38 | 0.22 | 9.40E-07 | ENSG00000148290.9 | SURF1 | Skin - Sun Exposed (Lower leg) |
| rs635634 | 9q34.2 | chr9_133279427_T_C_b38 | 0.18 | 1.10E-06 | ENSG00000148290.9 | SURF1 | Artery - Tibial |
| rs635634 | 9q34.2 | chr9_133279427_T_C_b38 | 0.26 | 1.10E-06 | ENSG00000148290.9 | SURF1 | Esophagus - Mucosa |
| rs635634 | 9q34.2 | chr9_133279427_T_C_b38 | -0.3 | 1.70E-06 | ENSG00000175164.13 | ABO | Esophagus - Gastroesophageal Junction |
| rs635634 | 9q34.2 | chr9_133279427_T_C_b38 | 0.19 | 2.00E-06 | ENSG00000148290.9 | SURF1 | Thyroid |
| rs635634 | 9q34.2 | chr9_133279427_T_C_b38 | -0.2 | 3.70E-06 | ENSG00000175164.13 | ABO | Lung |
| rs635634 | 9q34.2 | chr9_133279427_T_C_b38 | 0.45 | 1.20E-05 | ENSG00000148290.9 | SURF1 | Brain - Putamen (basal ganglia) |
| rs635634 | 9q34.2 | chr9_133279427_T_C_b38 | 0.087 | 2.10E-05 | ENSG00000148290.9 | SURF1 | Whole Blood |
| rs635634 | 9q34.2 | chr9_133279427_T_C_b38 | -0.14 | 2.40E-05 | ENSG00000175164.13 | ABO | Colon - Transverse |
| rs635634 | 9q34.2 | chr9_133279427_T_C_b38 | 0.15 | 4.40E-05 | ENSG00000148300.11 | REXO4 | Cells - Cultured fibroblasts |
| rs635634 | 9q34.2 | chr9_133279427_T_C_b38 | -0.21 | 5.80E-05 | ENSG00000175164.13 | ABO | Testis |

| | | | | | | | |
|---|---|---|---|---|---|---|---|
| rs635634 | 9q34.2 | chr9_133279427_T_C_b38 | 0.22 | 7.70E-05 | ENSG00000148290.9 | SURF1 | Cells - Cultured fibroblasts |
| rs6598541 | 15q26.3 | chr15_98727906_A_G_b38 | 0.22 | 7.10E-10 | ENSG00000140443.13 | IGF1R | Heart - Left Ventricle |
| rs6747542 | 2p13.3 | chr2_69879700_T_C_b38 | -0.36 | 3.00E-36 | ENSG00000124380.10 | SNRNP27 | Muscle - Skeletal |
| rs6747542 | 2p13.3 | chr2_69879700_T_C_b38 | -0.23 | 9.60E-16 | ENSG00000087338.4 | GMCL1 | Muscle - Skeletal |
| rs6747542 | 2p13.3 | chr2_69879700_T_C_b38 | 0.11 | 1.10E-14 | ENSG00000087338.4 | GMCL1 | Whole Blood |
| rs6747542 | 2p13.3 | chr2_69879700_T_C_b38 | 0.22 | 1.10E-13 | ENSG00000087338.4 | GMCL1 | Cells - Cultured fibroblasts |
| rs6747542 | 2p13.3 | chr2_69879700_T_C_b38 | -0.31 | 1.40E-13 | ENSG00000124380.10 | SNRNP27 | Testis |
| rs6747542 | 2p13.3 | chr2_69879700_T_C_b38 | -0.33 | 5.70E-13 | ENSG00000124380.10 | SNRNP27 | Brain - Cortex |
| rs6747542 | 2p13.3 | chr2_69879700_T_C_b38 | -0.29 | 1.80E-12 | ENSG00000087338.4 | GMCL1 | Brain - Cerebellum |
| rs6747542 | 2p13.3 | chr2_69879700_T_C_b38 | -0.28 | 1.20E-11 | ENSG00000087338.4 | GMCL1 | Pituitary |
| rs6747542 | 2p13.3 | chr2_69879700_T_C_b38 | -0.22 | 7.80E-11 | ENSG00000124380.10 | SNRNP27 | Heart - Atrial Appendage |
| rs6747542 | 2p13.3 | chr2_69879700_T_C_b38 | -0.35 | 2.30E-10 | ENSG00000124380.10 | SNRNP27 | Brain - Frontal Cortex (BA9) |
| rs6747542 | 2p13.3 | chr2_69879700_T_C_b38 | -0.37 | 3.60E-10 | ENSG00000124380.10 | SNRNP27 | Brain - Hippocampus |
| rs6747542 | 2p13.3 | chr2_69879700_T_C_b38 | -0.11 | 5.70E-10 | ENSG00000196975.15 | ANXA4 | Cells - Cultured fibroblasts |
| rs6747542 | 2p13.3 | chr2_69879700_T_C_b38 | -0.14 | 1.00E-09 | ENSG00000196975.15 | ANXA4 | Whole Blood |
| rs6747542 | 2p13.3 | chr2_69879700_T_C_b38 | -0.21 | 1.80E-09 | ENSG00000087338.4 | GMCL1 | Heart - Left Ventricle |
| rs6747542 | 2p13.3 | chr2_69879700_T_C_b38 | -0.28 | 6.80E-08 | ENSG00000087338.4 | GMCL1 | Brain - Cortex |
| rs6747542 | 2p13.3 | chr2_69879700_T_C_b38 | -0.24 | 1.10E-07 | ENSG00000124380.10 | SNRNP27 | Brain - Cerebellum |
| rs6747542 | 2p13.3 | chr2_69879700_T_C_b38 | -0.28 | 5.60E-07 | ENSG00000124380.10 | SNRNP27 | Brain - Nucleus accumbens (basal ganglia) |
| rs6747542 | 2p13.3 | chr2_69879700_T_C_b38 | -0.28 | 6.80E-07 | ENSG00000087338.4 | GMCL1 | Brain - Cerebellar Hemisphere |
| rs6747542 | 2p13.3 | chr2_69879700_T_C_b38 | -0.19 | 9.90E-07 | ENSG00000124380.10 | SNRNP27 | Colon - Sigmoid |
| rs6747542 | 2p13.3 | chr2_69879700_T_C_b38 | -0.26 | 1.40E-06 | ENSG00000124380.10 | SNRNP27 | Brain - Caudate (basal ganglia) |
| rs6747542 | 2p13.3 | chr2_69879700_T_C_b38 | -0.12 | 2.50E-06 | ENSG00000124380.10 | SNRNP27 | Heart - Left Ventricle |
| rs6747542 | 2p13.3 | chr2_69879700_T_C_b38 | 0.12 | 4.00E-06 | ENSG00000087338.4 | GMCL1 | Adipose - Subcutaneous |
| rs6747542 | 2p13.3 | chr2_69879700_T_C_b38 | -0.17 | 6.40E-06 | ENSG00000087338.4 | GMCL1 | Heart - Atrial Appendage |
| rs6747542 | 2p13.3 | chr2_69879700_T_C_b38 | -0.15 | 8.10E-06 | ENSG00000124380.10 | SNRNP27 | Esophagus - Muscularis |
| rs6747542 | 2p13.3 | chr2_69879700_T_C_b38 | -0.26 | 1.60E-05 | ENSG00000087338.4 | GMCL1 | Brain - Hypothalamus |
| rs6747542 | 2p13.3 | chr2_69879700_T_C_b38 | -0.27 | 2.10E-05 | ENSG00000124380.10 | SNRNP27 | Brain - Putamen (basal ganglia) |
| rs6747542 | 2p13.3 | chr2_69879700_T_C_b38 | -0.26 | 2.40E-05 | ENSG00000124380.10 | SNRNP27 | Brain - Hypothalamus |
| rs6747542 | 2p13.3 | chr2_69879700_T_C_b38 | -0.19 | 4.60E-05 | ENSG00000087338.4 | GMCL1 | Prostate |
| rs6747542 | 2p13.3 | chr2_69879700_T_C_b38 | -0.28 | 4.70E-05 | ENSG00000124380.10 | SNRNP27 | Brain - Anterior cingulate cortex (BA24) |
| rs680084 | 1q24.2 | chr1_170659114_G_A_b38 | 0.42 | 1.10E-23 | ENSG00000271811.1 | RP1-79C4.4 | Adipose - Subcutaneous |
| rs680084 | 1q24.2 | chr1_170659114_G_A_b38 | 0.37 | 8.50E-21 | ENSG00000271811.1 | RP1-79C4.4 | Skin - Sun Exposed (Lower leg) |
| rs680084 | 1q24.2 | chr1_170659114_G_A_b38 | 0.37 | 2.90E-17 | ENSG00000271811.1 | RP1-79C4.4 | Skin - Not Sun Exposed (Suprapubic) |
| rs680084 | 1q24.2 | chr1_170659114_G_A_b38 | 0.35 | 1.10E-15 | ENSG00000271811.1 | RP1-79C4.4 | Nerve - Tibial |
| rs680084 | 1q24.2 | chr1_170659114_G_A_b38 | 0.51 | 5.60E-15 | ENSG00000271811.1 | RP1-79C4.4 | Testis |
| rs680084 | 1q24.2 | chr1_170659114_G_A_b38 | 0.28 | 1.30E-14 | ENSG00000271811.1 | RP1-79C4.4 | Thyroid |
| rs680084 | 1q24.2 | chr1_170659114_G_A_b38 | 0.35 | 1.40E-13 | ENSG00000271811.1 | RP1-79C4.4 | Cells - Cultured fibroblasts |
| rs680084 | 1q24.2 | chr1_170659114_G_A_b38 | 0.34 | 2.80E-13 | ENSG00000271811.1 | RP1-79C4.4 | Adipose - Visceral (Omentum) |
| rs680084 | 1q24.2 | chr1_170659114_G_A_b38 | 0.29 | 5.30E-13 | ENSG00000271811.1 | RP1-79C4.4 | Artery - Tibial |
| rs680084 | 1q24.2 | chr1_170659114_G_A_b38 | 0.32 | 2.00E-11 | ENSG00000271811.1 | RP1-79C4.4 | Heart - Left Ventricle |
| rs680084 | 1q24.2 | chr1_170659114_G_A_b38 | 0.22 | 3.00E-11 | ENSG00000271811.1 | RP1-79C4.4 | Lung |
| rs680084 | 1q24.2 | chr1_170659114_G_A_b38 | 0.26 | 3.20E-11 | ENSG00000271811.1 | RP1-79C4.4 | Muscle - Skeletal |
| rs680084 | 1q24.2 | chr1_170659114_G_A_b38 | 0.31 | 3.60E-11 | ENSG00000271811.1 | RP1-79C4.4 | Esophagus - Gastroesophageal Junction |
| rs680084 | 1q24.2 | chr1_170659114_G_A_b38 | 0.45 | 4.10E-10 | ENSG00000271811.1 | RP1-79C4.4 | Prostate |
| rs680084 | 1q24.2 | chr1_170659114_G_A_b38 | 0.28 | 1.60E-09 | ENSG00000271811.1 | RP1-79C4.4 | Heart - Atrial Appendage |
| rs680084 | 1q24.2 | chr1_170659114_G_A_b38 | 0.29 | 6.30E-09 | ENSG00000271811.1 | RP1-79C4.4 | Breast - Mammary Tissue |
| rs680084 | 1q24.2 | chr1_170659114_G_A_b38 | 0.45 | 1.50E-08 | ENSG00000271811.1 | RP1-79C4.4 | Brain - Cortex |
| rs680084 | 1q24.2 | chr1_170659114_G_A_b38 | 0.21 | 4.10E-08 | ENSG00000271811.1 | RP1-79C4.4 | Esophagus - Muscularis |
| rs680084 | 1q24.2 | chr1_170659114_G_A_b38 | 0.25 | 5.20E-08 | ENSG00000116132.11 | PRRX1 | Heart - Left Ventricle |
| rs680084 | 1q24.2 | chr1_170659114_G_A_b38 | 0.44 | 8.10E-08 | ENSG00000271811.1 | RP1-79C4.4 | Brain - Anterior cingulate cortex (BA24) |

| | | | | | | | |
|---|---|---|---|---|---|---|---|
| rs680084 | 1q24.2 | chr1_170659114_G_A_b38 | 0.45 | 6.80E-07 | ENSG00000271811.1 | RP1-79C4.4 | Brain - Frontal Cortex (BA9) |
| rs680084 | 1q24.2 | chr1_170659114_G_A_b38 | 0.24 | 1.70E-06 | ENSG00000116132.11 | PRRX1 | Heart - Atrial Appendage |
| rs680084 | 1q24.2 | chr1_170659114_G_A_b38 | 0.34 | 1.80E-06 | ENSG00000271811.1 | RP1-79C4.4 | Brain - Nucleus accumbens (basal ganglia) |
| rs680084 | 1q24.2 | chr1_170659114_G_A_b38 | 0.39 | 2.20E-06 | ENSG00000271811.1 | RP1-79C4.4 | Brain - Cerebellum |
| rs680084 | 1q24.2 | chr1_170659114_G_A_b38 | 0.42 | 2.30E-06 | ENSG00000271811.1 | RP1-79C4.4 | Brain - Cerebellar Hemisphere |
| rs680084 | 1q24.2 | chr1_170659114_G_A_b38 | 0.31 | 2.70E-06 | ENSG00000271811.1 | RP1-79C4.4 | Artery - Coronary |
| rs680084 | 1q24.2 | chr1_170659114_G_A_b38 | 0.46 | 3.60E-06 | ENSG00000271811.1 | RP1-79C4.4 | Brain - Hippocampus |
| rs680084 | 1q24.2 | chr1_170659114_G_A_b38 | 0.19 | 4.60E-06 | ENSG00000271811.1 | RP1-79C4.4 | Esophagus - Mucosa |
| rs680084 | 1q24.2 | chr1_170659114_G_A_b38 | 0.36 | 6.00E-06 | ENSG00000271811.1 | RP1-79C4.4 | Ovary |
| rs680084 | 1q24.2 | chr1_170659114_G_A_b38 | 0.32 | 9.10E-06 | ENSG00000271811.1 | RP1-79C4.4 | Brain - Caudate (basal ganglia) |
| rs680084 | 1q24.2 | chr1_170659114_G_A_b38 | 0.34 | 3.10E-05 | ENSG00000271811.1 | RP1-79C4.4 | Brain - Putamen (basal ganglia) |
| rs680084 | 1q24.2 | chr1_170659114_G_A_b38 | 0.18 | 7.20E-05 | ENSG00000271811.1 | RP1-79C4.4 | Artery - Aorta |
| rs6891790 | 5q35.1 | chr5_173243742_G_T_b38 | 0.83 | 3.40E-24 | ENSG00000183072.9 | NKX2-5 | Pituitary |
| rs6891790 | 5q35.1 | chr5_173243742_G_T_b38 | 0.74 | 1.60E-13 | ENSG00000183072.9 | NKX2-5 | Brain - Cerebellum |
| rs6891790 | 5q35.1 | chr5_173243742_G_T_b38 | 0.61 | 1.70E-07 | ENSG00000183072.9 | NKX2-5 | Brain - Cerebellar Hemisphere |
| rs6891790 | 5q35.1 | chr5_173243742_G_T_b38 | 0.18 | 3.70E-06 | ENSG00000113734.17 | BNIP1 | Adipose - Subcutaneous |
| rs6891790 | 5q35.1 | chr5_173243742_G_T_b38 | 0.18 | 1.10E-05 | ENSG00000113734.17 | BNIP1 | Skin - Not Sun Exposed (Suprapubic) |
| rs6891790 | 5q35.1 | chr5_173243742_G_T_b38 | 0.14 | 6.80E-05 | ENSG00000113734.17 | BNIP1 | Artery - Tibial |
| rs6891790 | 5q35.1 | chr5_173243742_G_T_b38 | 0.11 | 1.00E-04 | ENSG00000113734.17 | BNIP1 | Whole Blood |
| rs74399915 | 11q24.3 | chr11_128899115_C_T_b38 | 0.49 | 1.70E-11 | ENSG00000174370.9 | C11orf45 | Artery - Tibial |
| rs74399915 | 11q24.3 | chr11_128899115_C_T_b38 | 0.35 | 8.10E-08 | ENSG00000174370.9 | C11orf45 | Artery - Aorta |
| rs74399915 | 11q24.3 | chr11_128899115_C_T_b38 | 0.35 | 4.40E-07 | ENSG00000120457.11 | KCNJ5 | Heart - Atrial Appendage |
| rs74399915 | 11q24.3 | chr11_128899115_C_T_b38 | 0.68 | 5.60E-07 | ENSG00000174370.9 | C11orf45 | Adrenal Gland |
| rs74399915 | 11q24.3 | chr11_128899115_C_T_b38 | -0.28 | 1.10E-06 | ENSG00000120457.11 | KCNJ5 | Lung |
| rs74399915 | 11q24.3 | chr11_128899115_C_T_b38 | 0.31 | 1.30E-05 | ENSG00000174370.9 | C11orf45 | Heart - Left Ventricle |
| rs74399915 | 11q24.3 | chr11_128899115_C_T_b38 | 0.31 | 2.10E-05 | ENSG00000174370.9 | C11orf45 | Heart - Atrial Appendage |
| rs74399915 | 11q24.3 | chr11_128899115_C_T_b38 | 0.29 | 0.00018 | ENSG00000174370.9 | C11orf45 | Cells - Cultured fibroblasts |
| rs76774446 | 17q21.32 | chr17_46969002_C_A_b38 | -0.46 | 4.80E-12 | ENSG00000179673.4 | RPRML | Muscle - Skeletal |
| rs76774446 | 17q21.32 | chr17_46969002_C_A_b38 | -0.34 | 2.30E-07 | ENSG00000262031.1 | LINC01974 | Testis |
| rs76774446 | 17q21.32 | chr17_46969002_C_A_b38 | 0.22 | 5.50E-07 | ENSG00000108433.16 | GOSR2 | Cells - Cultured fibroblasts |
| rs76774446 | 17q21.32 | chr17_46969002_C_A_b38 | -0.37 | 3.90E-06 | ENSG00000179673.4 | RPRML | Adipose - Subcutaneous |
| rs76774446 | 17q21.32 | chr17_46969002_C_A_b38 | 0.36 | 1.40E-05 | ENSG00000260075.1 | NSFP1 | Whole Blood |
| rs76774446 | 17q21.32 | chr17_46969002_C_A_b38 | 0.31 | 1.80E-05 | ENSG00000108379.9 | WNT3 | Adipose - Subcutaneous |
| rs76774446 | 17q21.32 | chr17_46969002_C_A_b38 | 0.22 | 1.90E-05 | ENSG00000262879.5 | RP11-156P1.3 | Testis |
| rs76774446 | 17q21.32 | chr17_46969002_C_A_b38 | -0.28 | 2.30E-05 | ENSG00000262879.5 | RP11-156P1.3 | Muscle - Skeletal |
| rs76774446 | 17q21.32 | chr17_46969002_C_A_b38 | 0.66 | 5.70E-05 | ENSG00000260075.1 | NSFP1 | Artery - Coronary |
| rs76774446 | 17q21.32 | chr17_46969002_C_A_b38 | 0.61 | 6.60E-05 | ENSG00000260075.1 | NSFP1 | Pituitary |
| rs76774446 | 17q21.32 | chr17_46969002_C_A_b38 | 0.16 | 8.30E-05 | ENSG00000006025.11 | OSBPL7 | Thyroid |
| rs76774446 | 17q21.32 | chr17_46969002_C_A_b38 | 0.21 | 0.00024 | ENSG00000108433.16 | GOSR2 | Skin - Sun Exposed (Lower leg) |
| rs76774446 | 17q21.32 | chr17_46969002_C_A_b38 | 0.34 | 0.00035 | ENSG00000260075.1 | NSFP1 | Skin - Sun Exposed (Lower leg) |
| rs78249997 | 10q22.2 | chr10_73662556_C_T_b38 | 1 | 4.00E-42 | ENSG00000177791.11 | MYOZ1 | Heart - Atrial Appendage |
| rs78249997 | 10q22.2 | chr10_73662556_C_T_b38 | -0.45 | 9.60E-16 | ENSG00000196968.10 | FUT11 | Nerve - Tibial |
| rs78249997 | 10q22.2 | chr10_73662556_C_T_b38 | -0.36 | 9.10E-13 | ENSG00000196968.10 | FUT11 | Adipose - Subcutaneous |
| rs78249997 | 10q22.2 | chr10_73662556_C_T_b38 | -0.24 | 1.80E-09 | ENSG00000196968.10 | FUT11 | Lung |
| rs78249997 | 10q22.2 | chr10_73662556_C_T_b38 | -0.18 | 1.60E-08 | ENSG00000166317.11 | SYNPO2L | Heart - Atrial Appendage |
| rs78249997 | 10q22.2 | chr10_73662556_C_T_b38 | -0.28 | 6.20E-08 | ENSG00000196968.10 | FUT11 | Adipose - Visceral (Omentum) |
| rs78249997 | 10q22.2 | chr10_73662556_C_T_b38 | -0.27 | 8.90E-08 | ENSG00000196968.10 | FUT11 | Esophagus - Mucosa |
| rs78249997 | 10q22.2 | chr10_73662556_C_T_b38 | 0.16 | 8.90E-08 | ENSG00000279088.1 | RP11-574K11.24 | Skin - Sun Exposed (Lower leg) |
| rs78249997 | 10q22.2 | chr10_73662556_C_T_b38 | -0.23 | 6.20E-07 | ENSG00000242338.6 | BMS1P4 | Esophagus - Mucosa |
| rs78249997 | 10q22.2 | chr10_73662556_C_T_b38 | -0.18 | 9.20E-07 | ENSG00000242338.6 | BMS1P4 | Cells - Cultured fibroblasts |
| rs78249997 | 10q22.2 | chr10_73662556_C_T_b38 | 0.17 | 1.10E-06 | ENSG00000279088.1 | RP11-574K11.24 | Artery - Tibial |

| | | | | | | | |
|---|---|---|---|---|---|---|---|
| rs78249997 | 10q22.2 | chr10_73662556_C_T_b38 | 0.18 | 1.90E-06 | ENSG00000279088.1 | RP11-574K11.24 | Adipose - Subcutaneous |
| rs78249997 | 10q22.2 | chr10_73662556_C_T_b38 | 0.11 | 2.00E-06 | ENSG00000166317.11 | SYNPO2L | Muscle - Skeletal |
| rs78249997 | 10q22.2 | chr10_73662556_C_T_b38 | -0.12 | 2.30E-06 | ENSG00000214655.10 | ZSWIM8 | Esophagus - Mucosa |
| rs78249997 | 10q22.2 | chr10_73662556_C_T_b38 | -0.17 | 2.70E-06 | ENSG00000196968.10 | FUT11 | Muscle - Skeletal |
| rs78249997 | 10q22.2 | chr10_73662556_C_T_b38 | -0.28 | 3.10E-06 | ENSG00000242338.6 | BMS1P4 | Testis |
| rs78249997 | 10q22.2 | chr10_73662556_C_T_b38 | 0.34 | 3.40E-06 | ENSG00000166507.17 | NDST2 | Spleen |
| rs78249997 | 10q22.2 | chr10_73662556_C_T_b38 | -0.13 | 4.30E-06 | ENSG00000196968.10 | FUT11 | Whole Blood |
| rs78249997 | 10q22.2 | chr10_73662556_C_T_b38 | 0.16 | 4.50E-06 | ENSG00000279088.1 | RP11-574K11.24 | Skin - Not Sun Exposed (Suprapubic) |
| rs78249997 | 10q22.2 | chr10_73662556_C_T_b38 | -0.26 | 6.20E-06 | ENSG00000166321.13 | NUDT13 | Esophagus - Mucosa |
| rs78249997 | 10q22.2 | chr10_73662556_C_T_b38 | -0.21 | 6.30E-06 | ENSG00000196968.10 | FUT11 | Breast - Mammary Tissue |
| rs78249997 | 10q22.2 | chr10_73662556_C_T_b38 | -0.27 | 6.50E-06 | ENSG00000196968.10 | FUT11 | Esophagus - Gastroesophageal Junction |
| rs78249997 | 10q22.2 | chr10_73662556_C_T_b38 | -0.19 | 7.80E-06 | ENSG00000242338.6 | BMS1P4 | Nerve - Tibial |
| rs78249997 | 10q22.2 | chr10_73662556_C_T_b38 | -0.23 | 7.80E-06 | ENSG00000196968.10 | FUT11 | Esophagus - Muscularis |
| rs78249997 | 10q22.2 | chr10_73662556_C_T_b38 | -0.29 | 8.00E-06 | ENSG00000166321.13 | NUDT13 | Adipose - Visceral (Omentum) |
| rs78249997 | 10q22.2 | chr10_73662556_C_T_b38 | -0.18 | 8.40E-06 | ENSG00000166321.13 | NUDT13 | Lung |
| rs78249997 | 10q22.2 | chr10_73662556_C_T_b38 | 0.27 | 9.30E-06 | ENSG00000224195.1 | RP11-574K11.5 | Artery - Aorta |
| rs78249997 | 10q22.2 | chr10_73662556_C_T_b38 | -0.45 | 1.00E-05 | ENSG00000196968.10 | FUT11 | Brain - Cerebellum |
| rs78249997 | 10q22.2 | chr10_73662556_C_T_b38 | -0.22 | 1.10E-05 | ENSG00000166321.13 | NUDT13 | Skin - Sun Exposed (Lower leg) |
| rs78249997 | 10q22.2 | chr10_73662556_C_T_b38 | -0.26 | 1.40E-05 | ENSG00000214655.10 | ZSWIM8 | Brain - Cerebellar Hemisphere |
| rs78249997 | 10q22.2 | chr10_73662556_C_T_b38 | -0.18 | 1.50E-05 | ENSG00000196968.10 | FUT11 | Cells - Cultured fibroblasts |
| rs78249997 | 10q22.2 | chr10_73662556_C_T_b38 | -0.23 | 1.70E-05 | ENSG00000196968.10 | FUT11 | Heart - Atrial Appendage |
| rs78249997 | 10q22.2 | chr10_73662556_C_T_b38 | 0.23 | 2.40E-05 | ENSG00000172650.13 | AGAP5 | Heart - Atrial Appendage |
| rs78249997 | 10q22.2 | chr10_73662556_C_T_b38 | 0.2 | 2.50E-05 | ENSG00000172650.13 | AGAP5 | Thyroid |
| rs78249997 | 10q22.2 | chr10_73662556_C_T_b38 | -0.16 | 3.10E-05 | ENSG00000166348.18 | USP54 | Nerve - Tibial |
| rs78249997 | 10q22.2 | chr10_73662556_C_T_b38 | -0.43 | 3.80E-05 | ENSG00000196968.10 | FUT11 | Pituitary |
| rs78249997 | 10q22.2 | chr10_73662556_C_T_b38 | 0.16 | 4.30E-05 | ENSG00000235316.1 | DUSP8P5 | Skin - Not Sun Exposed (Suprapubic) |
| rs78249997 | 10q22.2 | chr10_73662556_C_T_b38 | -0.17 | 4.40E-05 | ENSG00000196968.10 | FUT11 | Skin - Sun Exposed (Lower leg) |
| rs78249997 | 10q22.2 | chr10_73662556_C_T_b38 | -0.21 | 4.70E-05 | ENSG00000196968.10 | FUT11 | Testis |
| rs78249997 | 10q22.2 | chr10_73662556_C_T_b38 | 0.085 | 4.80E-05 | ENSG00000166507.17 | NDST2 | Whole Blood |
| rs78249997 | 10q22.2 | chr10_73662556_C_T_b38 | -0.34 | 5.00E-05 | ENSG00000166321.13 | NUDT13 | Colon - Sigmoid |
| rs78249997 | 10q22.2 | chr10_73662556_C_T_b38 | -0.42 | 5.20E-05 | ENSG00000242338.6 | BMS1P4 | Brain - Cortex |
| rs78249997 | 10q22.2 | chr10_73662556_C_T_b38 | 0.12 | 5.20E-05 | ENSG00000279088.1 | RP11-574K11.24 | Nerve - Tibial |
| rs78249997 | 10q22.2 | chr10_73662556_C_T_b38 | 0.14 | 5.70E-05 | ENSG00000138286.14 | FAM149B1 | Whole Blood |
| rs78249997 | 10q22.2 | chr10_73662556_C_T_b38 | 0.17 | 5.80E-05 | ENSG00000138286.14 | FAM149B1 | Adipose - Visceral (Omentum) |
| rs78249997 | 10q22.2 | chr10_73662556_C_T_b38 | 0.18 | 7.40E-05 | ENSG00000138286.14 | FAM149B1 | Adipose - Subcutaneous |
| rs78249997 | 10q22.2 | chr10_73662556_C_T_b38 | -0.54 | 9.90E-05 | ENSG00000196968.10 | FUT11 | Brain - Anterior cingulate cortex (BA24) |
| rs78249997 | 10q22.2 | chr10_73662556_C_T_b38 | -0.14 | 1.00E-04 | ENSG00000196968.10 | FUT11 | Artery - Tibial |
| rs78249997 | 10q22.2 | chr10_73662556_C_T_b38 | -0.27 | 0.00011 | ENSG00000166321.13 | NUDT13 | Esophagus - Muscularis |
| rs78249997 | 10q22.2 | chr10_73662556_C_T_b38 | -0.31 | 0.00013 | ENSG00000196968.10 | FUT11 | Colon - Sigmoid |
| rs78249997 | 10q22.2 | chr10_73662556_C_T_b38 | 0.22 | 0.00015 | ENSG00000166317.11 | SYNPO2L | Artery - Tibial |
| rs78249997 | 10q22.2 | chr10_73662556_C_T_b38 | -0.18 | 0.00016 | ENSG00000196968.10 | FUT11 | Thyroid |
| rs78249997 | 10q22.2 | chr10_73662556_C_T_b38 | -0.24 | 0.00017 | ENSG00000242338.6 | BMS1P4 | Adipose - Visceral (Omentum) |
| rs78249997 | 10q22.2 | chr10_73662556_C_T_b38 | -0.21 | 0.00019 | ENSG00000196968.10 | FUT11 | Heart - Left Ventricle |
| rs79187193 | 1q21.2 | chr1_147783720_G_A_b38 | -0.39 | 1.20E-07 | ENSG00000263956.6 | NBPF11 | Skin - Sun Exposed (Lower leg) |
| rs79187193 | 1q21.2 | chr1_147783720_G_A_b38 | 0.63 | 2.50E-05 | ENSG00000280649.2 | CH17-172B3.1 | Brain - Putamen (basal ganglia) |
| rs79187193 | 1q21.2 | chr1_147783720_G_A_b38 | -0.3 | 0.00012 | ENSG00000263956.6 | NBPF11 | Thyroid |
| rs79187193 | 1q21.2 | chr1_147783720_G_A_b38 | -0.29 | 0.00025 | ENSG00000263956.6 | NBPF11 | Nerve - Tibial |
| rs883079 | 12q24.21 | chr12_114355435_C_T_b38 | 0.14 | 4.10E-06 | ENSG00000089225.19 | TBX5 | Esophagus - Muscularis |

#### Online Table VIII. Significant eQTLGen eQTLs of the index variants from MTAG-CES

| SNP | Locus | EA | OA | Zscore | Pvalue | Gene | GeneSymbol | NrSamples |
|---|---|---|---|---|---|---|---|---|
| rs10272350 | 7q11.23 | A | G | -48.30 | 0.00E+00 | ENSG00000135211 | TMEM60 | 20990 |
| rs10272350 | 7q11.23 | A | G | -24.50 | 1.40E-132 | ENSG00000127947 | PTPN12 | 25851 |
| rs10272350 | 7q11.23 | A | G | 15.63 | 4.58E-55 | ENSG00000187257 | RSBN1L | 26065 |
| rs10272350 | 7q11.23 | A | G | -7.04 | 1.86E-12 | ENSG00000135205 | CCDC146 | 26065 |
| rs10272350 | 7q11.23 | A | G | -6.86 | 7.06E-12 | ENSG00000186088 | PION | 16000 |
| rs10272350 | 7q11.23 | A | G | -4.98 | 6.37E-07 | ENSG00000006576 | PHTF2 | 25851 |
| rs10753933 | 1q32.1 | T | G | -83.81 | 0.00E+00 | ENSG00000143847 | PPFIA4 | 26278 |
| rs10753933 | 1q32.1 | T | G | -7.73 | 1.05E-14 | ENSG00000163444 | TMEM183A | 21004 |
| rs10753933 | 1q32.1 | T | G | -5.23 | 1.67E-07 | ENSG00000163485 | ADORA1 | 31313 |
| rs10753933 | 1q32.1 | T | G | 4.52 | 6.23E-06 | ENSG00000117153 | KLHL12 | 31353 |
| rs10753933 | 1q32.1 | T | G | -4.29 | 1.75E-05 | ENSG00000133048 | CHI3L1 | 25695 |
| rs11057583 | 12q24.31 | A | G | -6.54 | 6.33E-11 | ENSG00000196498 | NCOR2 | 31354 |
| rs11125871 | 2p15 | T | C | -46.35 | 0.00E+00 | ENSG00000162929 | KIAA1841 | 26180 |
| rs11125871 | 2p15 | T | C | 45.46 | 0.00E+00 | ENSG00000173209 | AHSA2 | 14262 |
| rs11125871 | 2p15 | T | C | 21.96 | 6.48E-107 | ENSG00000212978 | AC016747.3 | 8268 |
| rs11125871 | 2p15 | T | C | 11.72 | 1.04E-31 | ENSG00000162927 | PUS10 | 14262 |
| rs11125871 | 2p15 | T | C | 4.77 | 1.82E-06 | ENSG00000162924 | REL | 31528 |
| rs11125871 | 2p15 | T | C | -4.33 | 1.52E-05 | ENSG00000162928 | PEX13 | 31528 |
| rs11264280 | 1q21.3 | T | C | -19.27 | 9.41E-83 | ENSG00000143537 | ADAM15 | 31599 |
| rs11264280 | 1q21.3 | T | C | 13.03 | 8.44E-39 | ENSG00000163346 | PBXIP1 | 31599 |
| rs11264280 | 1q21.3 | T | C | 10.09 | 6.44E-24 | ENSG00000163344 | PMVK | 31599 |
| rs11264280 | 1q21.3 | T | C | 8.02 | 1.10E-15 | ENSG00000169231 | THBS3 | 31599 |
| rs11264280 | 1q21.3 | T | C | 7.04 | 1.87E-12 | ENSG00000185499 | MUC1 | 31599 |
| rs11264280 | 1q21.3 | T | C | 5.63 | 1.78E-08 | ENSG00000232093 | RP11-307C12.11 | 4783 |
| rs11264280 | 1q21.3 | T | C | -5.34 | 9.43E-08 | ENSG00000132718 | SYT11 | 31385 |
| rs11264280 | 1q21.3 | T | C | -5.23 | 1.65E-07 | ENSG00000132676 | DAP3 | 31599 |
| rs11264280 | 1q21.3 | T | C | 5.22 | 1.83E-07 | ENSG00000160685 | ZBTB7B | 31599 |
| rs11264280 | 1q21.3 | T | C | -4.40 | 1.08E-05 | ENSG00000163374 | YY1AP1 | 31599 |
| rs11598047 | 10q24.33 | G | A | 6.38 | 1.82E-10 | ENSG00000173915 | USMG5 | 31568 |
| rs116600817 | 17p13.1 | G | A | 65.02 | 0.00E+00 | ENSG00000239697 | TNFSF12 | 31569 |
| rs116600817 | 17p13.1 | G | A | -18.83 | 4.61E-79 | ENSG00000141504 | SAT2 | 31569 |
| rs116600817 | 17p13.1 | G | A | -17.49 | 1.66E-68 | ENSG00000129226 | CD68 | 31355 |
| rs116600817 | 17p13.1 | G | A | -17.43 | 5.17E-68 | ENSG00000233223 | AC113189.5 | 5502 |
| rs116600817 | 17p13.1 | G | A | -16.17 | 7.90E-59 | ENSG00000170175 | CHRNB1 | 31569 |
| rs116600817 | 17p13.1 | G | A | 11.69 | 1.52E-31 | ENSG00000161955 | TNFSF13 | 23727 |
| rs116600817 | 17p13.1 | G | A | -8.13 | 4.31E-16 | ENSG00000072818 | ACAP1 | 14263 |
| rs116600817 | 17p13.1 | G | A | -6.73 | 1.64E-11 | ENSG00000129255 | MPDU1 | 31315 |
| rs116600817 | 17p13.1 | G | A | 5.98 | 2.27E-09 | ENSG00000187838 | PLSCR3 | 31355 |
| rs116600817 | 17p13.1 | G | A | 5.25 | 1.55E-07 | ENSG00000213859 | KCTD11 | 31569 |
| rs116600817 | 17p13.1 | G | A | 5.18 | 2.25E-07 | ENSG00000264772 | RP11-186B7.4 | 16474 |
| rs116600817 | 17p13.1 | G | A | -4.36 | 1.31E-05 | ENSG00000170004 | CHD3 | 31355 |
| rs116600817 | 17p13.1 | G | A | 4.35 | 1.38E-05 | ENSG00000174282 | ZBTB4 | 31355 |
| rs11782313 | 8q24.3 | C | T | -19.80 | 3.09E-87 | ENSG00000169398 | PTK2 | 31564 |
| rs11782313 | 8q24.3 | C | T | -10.35 | 4.38E-25 | ENSG00000104472 | CHRAC1 | 31564 |
| rs11782313 | 8q24.3 | C | T | -4.93 | 8.36E-07 | ENSG00000253210 | RP11-809O17.1 | 5497 |
| rs12211255 | 6q14.1 | A | C | 18.80 | 8.05E-79 | ENSG00000112701 | SENP6 | 31354 |

| | | | | | | | | |
|---|---|---|---|---|---|---|---|---|
| rs12211255 | 6q14.1 | A | C | -10.27 | 9.72E-25 | ENSG00000196586 | MYO6 | 31568 |
| rs12211255 | 6q14.1 | A | C | 8.00 | 1.24E-15 | ENSG00000112697 | TMEM30A | 31354 |
| rs12260801 | 10q25.2 | T | C | 4.34 | 1.42E-05 | ENSG00000214413 | BBIP1 | 6408 |
| rs13191450 | 6q22.31 | C | A | 7.99 | 1.40E-15 | ENSG00000025156 | HSF2 | 31354 |
| rs140185678 | 16p13.3 | A | G | 8.53 | 1.43E-17 | ENSG00000255198 | SNHG9 | 7374 |
| rs17171711 | 5q31.2 | T | C | 18.49 | 2.32E-76 | ENSG00000146021 | KLHL3 | 31568 |
| rs17171711 | 5q31.2 | T | C | -14.01 | 1.31E-44 | ENSG00000120733 | KDM3B | 25744 |
| rs17171711 | 5q31.2 | T | C | 11.94 | 7.45E-33 | ENSG00000044115 | CTNNA1 | 31528 |
| rs1898096 | 10q22.3 | G | A | 4.40 | 1.07E-05 | ENSG00000165655 | ZNF503 | 26492 |
| rs2106261 | 16q22.3 | T | C | 6.16 | 7.24E-10 | ENSG00000257017 | HP | 30802 |
| rs2269001 | 7q36.1 | A | G | -11.22 | 3.22E-29 | ENSG00000055118 | KCNH2 | 31561 |
| rs2269001 | 7q36.1 | A | G | -7.90 | 2.74E-15 | ENSG00000002726 | ABP1 | 26486 |
| rs2269001 | 7q36.1 | A | G | -5.48 | 4.17E-08 | ENSG00000133612 | AGAP3 | 31347 |
| rs2738413 | 14q23.2 | A | G | 20.32 | 8.52E-92 | ENSG00000140009 | ESR2 | 29950 |
| rs2738413 | 14q23.2 | A | G | 17.12 | 9.96E-66 | ENSG00000089775 | ZBTB25 | 29950 |
| rs2738413 | 14q23.2 | A | G | 16.92 | 3.43E-64 | ENSG00000054654 | SYNE2 | 29950 |
| rs2738413 | 14q23.2 | A | G | 11.98 | 4.37E-33 | ENSG00000100714 | MTHFD1 | 14262 |
| rs2738413 | 14q23.2 | A | G | 4.44 | 9.00E-06 | ENSG00000126822 | PLEKHG3 | 29415 |
| rs2857265 | 2q31.2 | A | G | 7.90 | 2.89E-15 | ENSG00000187231 | SESTD1 | 30721 |
| rs2857265 | 2q31.2 | A | G | -5.98 | 2.21E-09 | ENSG00000155657 | TTN | 31470 |
| rs2857265 | 2q31.2 | A | G | -4.63 | 3.64E-06 | ENSG00000237298 | TTN-AS1 | 16923 |
| rs2885697 | 1p34.2 | G | T | 18.65 | 1.29E-77 | ENSG00000010803 | SCMH1 | 31684 |
| rs2885697 | 1p34.2 | G | T | 9.71 | 2.67E-22 | ENSG00000171793 | CTPS1 | 25690 |
| rs2885697 | 1p34.2 | G | T | -7.72 | 1.18E-14 | ENSG00000179862 | CITED4 | 31684 |
| rs2885697 | 1p34.2 | G | T | -5.12 | 2.99E-07 | ENSG00000117016 | RIMS3 | 31684 |
| rs2885697 | 1p34.2 | G | T | 5.01 | 5.40E-07 | ENSG00000204060 | FOXO6 | 17261 |
| rs2885697 | 1p34.2 | G | T | 4.81 | 1.49E-06 | ENSG00000171790 | SLFNL1 | 31684 |
| rs3176326 | 6p21.2 | A | G | -22.33 | 2.06E-110 | ENSG00000124762 | CDKN1A | 31344 |
| rs3176326 | 6p21.2 | A | G | 7.77 | 7.63E-15 | ENSG00000255587 | RAB44 | 5491 |
| rs3176326 | 6p21.2 | A | G | 4.81 | 1.50E-06 | ENSG00000112078 | KCTD20 | 31344 |
| rs3176326 | 6p21.2 | A | G | -4.58 | 4.55E-06 | ENSG00000010030 | ETV7 | 31090 |
| rs3807989 | 7q31.2 | A | G | -10.41 | 2.27E-25 | ENSG00000135269 | TES | 31086 |
| rs41292535 | 1p32.2 | A | G | -16.61 | 5.61E-62 | ENSG00000117862 | TXNDC12 | 26556 |
| rs41292535 | 1p32.2 | A | G | -5.96 | 2.53E-09 | ENSG00000123091 | RNF11 | 26516 |
| rs4385527 | 9q22.32 | A | G | -11.13 | 9.23E-29 | ENSG00000148120 | C9orf3 | 26196 |
| rs4385527 | 9q22.32 | A | G | 5.20 | 2.01E-07 | ENSG00000148110 | HIATL1 | 31684 |
| rs4385527 | 9q22.32 | A | G | -4.57 | 4.80E-06 | ENSG00000175787 | ZNF169 | 31684 |
| rs55985730 | 7q32.1 | G | T | 11.90 | 1.18E-32 | ENSG00000128595 | CALU | 30537 |
| rs55985730 | 7q32.1 | G | T | -9.54 | 1.48E-21 | ENSG00000230715 | RP11-274B21.4 | 5492 |
| rs55985730 | 7q32.1 | G | T | -8.77 | 1.85E-18 | ENSG00000243679 | RP11-274B21.3 | 5492 |
| rs55985730 | 7q32.1 | G | T | -6.42 | 1.36E-10 | ENSG00000243302 | RP11-274B21.2 | 5492 |
| rs55985730 | 7q32.1 | G | T | -5.44 | 5.31E-08 | ENSG00000165055 | METTL2B | 25462 |
| rs55985730 | 7q32.1 | G | T | -4.92 | 8.81E-07 | ENSG00000242588 | RP11-274B21.1 | 21463 |
| rs55985730 | 7q32.1 | G | T | -4.78 | 1.72E-06 | ENSG00000128604 | IRF5 | 29748 |
| rs55985730 | 7q32.1 | G | T | -4.71 | 2.44E-06 | ENSG00000106344 | RBM28 | 30537 |
| rs56181519 | 2q31.1 | T | C | 8.25 | 1.57E-16 | ENSG00000138433 | CIR1 | 31684 |

| | | | | | | | | |
|---|---|---|---|---|---|---|---|---|
| rs56181519 | 2q31.1 | T | C | 6.20 | 5.50E-10 | ENSG00000163328 | GPR155 | 30935 |
| rs56181519 | 2q31.1 | T | C | 4.67 | 3.04E-06 | ENSG00000115935 | WIPF1 | 31470 |
| rs635634 | 9q34.2 | T | C | -35.69 | 5.22E-279 | ENSG00000175164 | ABO | 9188 |
| rs635634 | 9q34.2 | T | C | -11.45 | 2.40E-30 | ENSG00000148290 | SURF1 | 9188 |
| rs635634 | 9q34.2 | T | C | -10.28 | 8.93E-25 | ENSG00000148288 | GBGT1 | 31185 |
| rs635634 | 9q34.2 | T | C | -6.09 | 1.12E-09 | ENSG00000160325 | CACFD1 | 25191 |
| rs635634 | 9q34.2 | T | C | 5.12 | 3.03E-07 | ENSG00000148297 | MED22 | 26110 |
| rs6598541 | 15q26.3 | A | G | 4.43 | 9.27E-06 | ENSG00000140443 | IGF1R | 26278 |
| rs6747542 | 2p13.3 | C | T | -25.89 | 9.57E-148 | ENSG00000196975 | ANXA4 | 30935 |
| rs6747542 | 2p13.3 | C | T | 19.95 | 1.63E-88 | ENSG00000087338 | GMCL1 | 31470 |
| rs6747542 | 2p13.3 | C | T | 19.01 | 1.30E-80 | ENSG00000124380 | SNRNP27 | 31470 |
| rs6747542 | 2p13.3 | C | T | -7.64 | 2.23E-14 | ENSG00000244617 | ASPRV1 | 26609 |
| rs6747542 | 2p13.3 | C | T | -6.85 | 7.15E-12 | ENSG00000179818 | PCBP1-AS1 | 10577 |
| rs6747542 | 2p13.3 | C | T | 6.85 | 7.40E-12 | ENSG00000059728 | MXD1 | 31470 |
| rs6747542 | 2p13.3 | C | T | -6.34 | 2.32E-10 | ENSG00000115998 | C2orf42 | 31684 |
| rs6747542 | 2p13.3 | C | T | -5.57 | 2.59E-08 | ENSG00000035141 | FAM136A | 31684 |
| rs680084 | 1q24.2 | G | A | 9.67 | 4.12E-22 | ENSG00000120370 | GORAB | 31526 |
| rs680084 | 1q24.2 | G | A | -7.76 | 8.47E-15 | ENSG00000188404 | SELL | 31566 |
| rs6891790 | 5q35.1 | T | G | 12.20 | 3.00E-34 | ENSG00000113734 | BNIP1 | 31470 |
| rs6891790 | 5q35.1 | T | G | 7.67 | 1.71E-14 | ENSG00000164463 | CREBRF | 25650 |
| rs7172038 | 15q24.1 | G | T | 17.33 | 3.04E-67 | ENSG00000067141 | NEO1 | 31564 |
| rs74399915 | 11q24.3 | T | C | 5.14 | 2.80E-07 | ENSG00000174370 | C11orf45 | 28204 |
| rs76774446 | 17q21.32 | A | C | 14.85 | 7.30E-50 | ENSG00000176681 | LRRC37A | 6418 |
| rs76774446 | 17q21.32 | A | C | 13.08 | 4.32E-39 | ENSG00000260075 | NSFP1 | 5499 |
| rs76774446 | 17q21.32 | A | C | 10.22 | 1.60E-24 | ENSG00000228696 | ARL17B | 8266 |
| rs76774446 | 17q21.32 | A | C | -7.35 | 1.97E-13 | ENSG00000232300 | FAM215B | 4022 |
| rs76774446 | 17q21.32 | A | C | 5.28 | 1.29E-07 | ENSG00000073969 | NSF | 31566 |
| rs76774446 | 17q21.32 | A | C | 5.02 | 5.10E-07 | ENSG00000198933 | TBKBP1 | 30817 |
| rs78249997 | 10q22.2 | T | C | 15.97 | 1.95E-57 | ENSG00000166507 | NDST2 | 28580 |
| rs78249997 | 10q22.2 | T | C | -14.69 | 7.23E-49 | ENSG00000122882 | ECD | 28481 |
| rs78249997 | 10q22.2 | T | C | -14.11 | 3.21E-45 | ENSG00000166321 | NUDT13 | 28794 |
| rs78249997 | 10q22.2 | T | C | 11.28 | 1.64E-29 | ENSG00000182180 | MRPS16 | 28794 |
| rs78249997 | 10q22.2 | T | C | -10.37 | 3.37E-25 | ENSG00000156042 | TTC18 | 11488 |
| rs78249997 | 10q22.2 | T | C | 9.71 | 2.85E-22 | ENSG00000138286 | FAM149B1 | 23306 |
| rs78249997 | 10q22.2 | T | C | -6.21 | 5.30E-10 | ENSG00000244365 | RP11-152N13.12 | 8906 |
| rs78249997 | 10q22.2 | T | C | 5.62 | 1.91E-08 | ENSG00000122884 | P4HA1 | 28794 |
| rs78249997 | 10q22.2 | T | C | 5.02 | 5.14E-07 | ENSG00000224195 | RP11-574K11.5 | 4986 |
| rs78249997 | 10q22.2 | T | C | 4.82 | 1.42E-06 | ENSG00000156110 | ADK | 6413 |
| rs78249997 | 10q22.2 | T | C | -4.82 | 1.46E-06 | ENSG00000213551 | DNAJC9 | 6413 |
| rs78249997 | 10q22.2 | T | C | -4.59 | 4.52E-06 | ENSG00000251582 | RP11-464F9.9 | 4339 |
| rs78249997 | 10q22.2 | T | C | -4.52 | 6.33E-06 | ENSG00000242288 | RP11-464F9.1 | 4339 |
| rs78249997 | 10q22.2 | T | C | 4.40 | 1.09E-05 | ENSG00000148660 | CAMK2G | 28580 |

**Online Table IX. Results of CES analysis on the European independent cohort.** AA: Assesed allele, OA: Other allele, CHR: chromosome, BP: base pairs, SE: Standard Error, B: beta, Z: z-score; P: p-value. Genomic location is in Hg19

| | | | | | | | | | MTAG CES | | | | CES analysis (Spanish cohort) | | | |
|---|---|---|---|---|---|---|---|---|---|---|---|---|---|---|---|---|
| **Index Variant** | **Gene** | **SNP** | **r2** | **D'** | **CHR** | **BP** | **AA** | **OA** | **B** | **SE** | **Z** | **P** | **B** | **SE** | **Z** | **P** |
| rs10753933 | PPFIA4 | rs4245712 | 0.98 | 0.99 | 1 | 203022023 | T | G | 0.012 | 0.002 | 7.789 | 6.76E-15 | -0.004 | 0.005 | -0.786 | 4.32E-01 |
| rs10753933 | PPFIA4 | rs10753933 | 1 | 1 | 1 | 203026214 | T | G | 0.013 | 0.002 | 8.241 | 1.70E-16 | -0.004 | 0.005 | -0.812 | 4.17E-01 |
| rs10753933 | PPFIA4 | rs2270543 | 0.98 | 1 | 1 | 203030685 | T | C | 0.013 | 0.002 | 8.047 | 8.51E-16 | -0.004 | 0.005 | -0.752 | 4.52E-01 |
| rs10753933 | PPFIA4 | rs11579055 | 1 | 1 | 1 | 203031315 | T | G | -0.013 | 0.002 | -8.076 | 6.70E-16 | 0.004 | 0.005 | 0.742 | 4.58E-01 |
| rs10753933 | PPFIA4 | rs11579558 | 1 | 1 | 1 | 203032038 | T | G | -0.013 | 0.002 | -8.144 | 3.81E-16 | 0.003 | 0.005 | 0.726 | 4.68E-01 |
| rs10753933 | PPFIA4 | rs4950913 | 1 | 1 | 1 | 203032153 | A | G | 0.013 | 0.002 | 8.186 | 2.70E-16 | -0.003 | 0.005 | -0.696 | 4.87E-01 |
| rs10753933 | PPFIA4 | rs7538696 | 0.99 | 1 | 1 | 203034309 | A | G | -0.013 | 0.002 | -8.015 | 1.10E-15 | 0.003 | 0.005 | 0.681 | 4.96E-01 |
| rs10753933 | PPFIA4 | rs3737883 | 0.99 | 1 | 1 | 203034906 | A | G | 0.013 | 0.002 | 8.009 | 1.16E-15 | -0.003 | 0.005 | -0.647 | 5.18E-01 |
| rs10753933 | PPFIA4 | rs2275354 | 0.97 | 1 | 1 | 203036757 | T | C | 0.013 | 0.002 | 8.058 | 7.75E-16 | -0.003 | 0.005 | -0.709 | 4.79E-01 |
| rs11057583 | NCOR2 | rs12306068 | 0.96 | 0.99 | 12 | 124806981 | T | G | -0.014 | 0.002 | -5.619 | 1.92E-08 | 0.000 | 0.009 | -0.048 | 9.62E-01 |
| rs11057583 | NCOR2 | rs11057583 | 1 | 1 | 12 | 124810329 | A | G | 0.014 | 0.002 | 5.666 | 1.46E-08 | 0.002 | 0.009 | 0.249 | 8.03E-01 |
| rs11099098 | FGF5, PRDM8 | rs11099097 | 0.95 | 1 | 4 | 81167309 | T | C | 0.009 | 0.002 | 5.500 | 3.81E-08 | 0.000 | 0.005 | 0.063 | 9.50E-01 |
| rs11099098 | FGF5, PRDM8 | rs11099098 | 1 | 1 | 4 | 81169912 | T | G | 0.010 | 0.002 | 5.582 | 2.38E-08 | -0.001 | 0.005 | -0.113 | 9.10E-01 |
| rs11099098 | FGF5, PRDM8 | rs13125101 | 0.98 | 0.99 | 4 | 81174592 | A | G | 0.010 | 0.002 | 5.567 | 2.59E-08 | 0.000 | 0.005 | -0.065 | 9.48E-01 |
| rs11099098 | FGF5, PRDM8 | rs12509595 | 0.95 | 0.99 | 4 | 81182554 | T | C | -0.009 | 0.002 | -5.496 | 3.88E-08 | 0.000 | 0.005 | 0.066 | 9.47E-01 |
| rs11125871 | USP34 | rs12998475 | 0.9 | 0.99 | 2 | 61437487 | T | C | 0.009 | 0.002 | 5.474 | 4.41E-08 | -0.005 | 0.005 | -0.977 | 3.29E-01 |
| rs11125871 | USP34 | rs4672426 | 0.81 | 0.98 | 2 | 61457148 | A | G | -0.009 | 0.002 | -5.454 | 4.91E-08 | 0.005 | 0.005 | 1.060 | 2.89E-01 |
| rs11125871 | USP34 | rs4671399 | 0.82 | 0.98 | 2 | 61458518 | T | C | -0.009 | 0.002 | -5.474 | 4.40E-08 | 0.005 | 0.005 | 1.018 | 3.09E-01 |
| rs11125871 | USP34 | rs6545848 | 0.83 | 0.99 | 2 | 61461183 | T | G | -0.009 | 0.002 | -5.529 | 3.22E-08 | 0.005 | 0.005 | 1.040 | 2.98E-01 |
| rs11125871 | USP34 | rs12621879 | 0.82 | 0.98 | 2 | 61461829 | A | G | -0.009 | 0.002 | -5.488 | 4.07E-08 | 0.005 | 0.005 | 1.071 | 2.84E-01 |
| rs11125871 | USP34 | rs11125871 | 1 | 1 | 2 | 61470126 | T | C | -0.009 | 0.002 | -5.745 | 9.19E-09 | 0.005 | 0.005 | 0.974 | 3.30E-01 |
| rs11264280 | KCNN3 | rs4578216 | 0.84 | 0.94 | 1 | 154900721 | T | C | -0.023 | 0.002 | -13.811 | 2.19E-43 | -0.009 | 0.005 | -1.790 | 7.35E-02 |
| rs11264280 | KCNN3 | rs11264280 | 1 | 1 | 1 | 154862952 | T | C | 0.024 | 0.002 | 14.217 | 7.17E-46 | 0.009 | 0.005 | 1.779 | 7.53E-02 |
| rs11598047 | NEURL1 | rs60469668 | 0.8 | 0.98 | 10 | 105331866 | A | G | -0.026 | 0.002 | -13.048 | 6.51E-39 | -0.016 | 0.006 | -2.660 | **7.82E-03** |
| rs11598047 | NEURL1 | rs12411463 | 0.8 | 0.98 | 10 | 105333613 | T | C | 0.026 | 0.002 | 13.081 | 4.21E-39 | 0.016 | 0.006 | 2.644 | **8.18E-03** |
| rs11598047 | NEURL1 | rs11815048 | 0.92 | 0.99 | 10 | 105338822 | A | G | 0.028 | 0.002 | 13.271 | 3.39E-40 | 0.019 | 0.006 | 3.104 | **1.91E-03** |
| rs11598047 | NEURL1 | rs11598047 | 1 | 1 | 10 | 105342672 | A | G | -0.028 | 0.002 | -13.384 | 7.50E-41 | -0.020 | 0.006 | -3.201 | **1.37E-03** |
| rs116600817 | TNFSF12 | rs62059793 | 0.82 | 0.93 | 17 | 7438687 | A | G | 0.010 | 0.002 | 5.513 | 3.53E-08 | 0.003 | 0.005 | 0.629 | 5.30E-01 |
| rs116600817 | TNFSF12 | rs35049113 | 0.86 | 0.95 | 17 | 7439726 | T | C | 0.010 | 0.002 | 5.555 | 2.78E-08 | 0.005 | 0.005 | 0.847 | 3.97E-01 |

| | | | | | | | | | | | | | | | | |
|---|---|---|---|---|---|---|---|---|---|---|---|---|---|---|---|---|
| rs116600817 | TNFSF12 | rs62059797 | 0.92 | 0.97 | 17 | 7443957 | A | G | 0.010 | 0.002 | 5.551 | 2.85E-08 | 0.003 | 0.005 | 0.610 | 5.42E-01 |
| rs116600817 | TNFSF12 | rs9905587 | 0.94 | 0.97 | 17 | 7444945 | T | C | -0.010 | 0.002 | -5.523 | 3.33E-08 | -0.004 | 0.005 | -0.750 | 4.53E-01 |
| rs116600817 | TNFSF12 | rs12948869 | 0.94 | 0.98 | 17 | 7449048 | A | G | -0.010 | 0.002 | -5.514 | 3.50E-08 | -0.004 | 0.005 | -0.702 | 4.82E-01 |
| rs116600817 | TNFSF12 | rs9899183 | 0.96 | -0.99 | 17 | 7452977 | T | C | 0.010 | 0.002 | 5.608 | 2.05E-08 | 0.003 | 0.005 | 0.553 | 5.80E-01 |
| rs116600817 | TNFSF12 | rs62059804 | 0.96 | 0.99 | 17 | 7453505 | A | C | -0.010 | 0.002 | -5.558 | 2.73E-08 | -0.004 | 0.005 | -0.738 | 4.60E-01 |
| rs116600817 | TNFSF12 | rs62059805 | 0.99 | 1 | 17 | 7455391 | T | C | -0.010 | 0.002 | -5.561 | 2.69E-08 | -0.003 | 0.005 | -0.524 | 6.00E-01 |
| rs116600817 | TNFSF12 | rs116600817 | 1 | 1 | 17 | 7458795 | A | G | 0.010 | 0.002 | 5.682 | 1.33E-08 | 0.003 | 0.005 | 0.575 | 5.65E-01 |
| rs116600817 | TNFSF12 | rs78744936 | 0.92 | 0.99 | 17 | 7461343 | A | G | -0.010 | 0.002 | -5.490 | 4.02E-08 | -0.002 | 0.005 | -0.356 | 7.22E-01 |
| rs11782313 | PTK2 | rs13276301 | 0.82 | 0.91 | 8 | 141730727 | T | G | 0.009 | 0.002 | 5.472 | 4.44E-08 | 0.006 | 0.005 | 1.177 | 2.39E-01 |
| rs11782313 | PTK2 | rs13271554 | 0.94 | 0.97 | 8 | 141873688 | A | G | 0.009 | 0.002 | 5.512 | 3.55E-08 | 0.005 | 0.005 | 0.969 | 3.33E-01 |
| rs11782313 | PTK2 | rs1868280 | 0.94 | 0.97 | 8 | 141896254 | A | G | -0.009 | 0.002 | -5.473 | 4.41E-08 | -0.005 | 0.005 | -1.081 | 2.80E-01 |
| rs11782313 | PTK2 | rs11991796 | 0.95 | 0.98 | 8 | 141898518 | T | C | 0.009 | 0.002 | 5.465 | 4.63E-08 | 0.004 | 0.005 | 0.808 | 4.19E-01 |
| rs11782313 | PTK2 | rs13250888 | 0.94 | 0.97 | 8 | 141899030 | A | G | 0.009 | 0.002 | 5.480 | 4.26E-08 | 0.005 | 0.005 | 1.012 | 3.11E-01 |
| rs11782313 | PTK2 | rs11783743 | 0.95 | 0.98 | 8 | 141907217 | A | G | 0.009 | 0.002 | 5.521 | 3.37E-08 | 0.006 | 0.005 | 1.178 | 2.39E-01 |
| rs11782313 | PTK2 | rs13277099 | 0.93 | 0.97 | 8 | 141915071 | T | G | 0.009 | 0.002 | 5.479 | 4.27E-08 | 0.004 | 0.005 | 0.758 | 4.49E-01 |
| rs11782313 | PTK2 | rs13270279 | 0.94 | 0.97 | 8 | 141915664 | T | C | -0.009 | 0.002 | -5.495 | 3.90E-08 | -0.003 | 0.005 | -0.664 | 5.07E-01 |
| rs11782313 | PTK2 | rs13250787 | 0.95 | 0.98 | 8 | 141916932 | T | G | -0.009 | 0.002 | -5.581 | 2.39E-08 | -0.003 | 0.005 | -0.669 | 5.03E-01 |
| rs11782313 | PTK2 | rs11777839 | 0.95 | 0.98 | 8 | 141917894 | T | G | -0.009 | 0.002 | -5.543 | 2.97E-08 | -0.003 | 0.005 | -0.682 | 4.95E-01 |
| rs11782313 | PTK2 | rs13264974 | 0.95 | 0.98 | 8 | 141918720 | A | C | -0.009 | 0.002 | -5.485 | 4.14E-08 | -0.003 | 0.005 | -0.669 | 5.03E-01 |
| rs11782313 | PTK2 | rs11778798 | 0.95 | 0.98 | 8 | 141918880 | A | G | -0.009 | 0.002 | -5.564 | 2.63E-08 | -0.003 | 0.005 | -0.669 | 5.03E-01 |
| rs11782313 | PTK2 | rs10089610 | 0.95 | 0.98 | 8 | 141921613 | T | G | -0.009 | 0.002 | -5.488 | 4.06E-08 | -0.003 | 0.005 | -0.676 | 4.99E-01 |
| rs11782313 | PTK2 | rs4357336 | 0.95 | 0.98 | 8 | 141922847 | T | C | -0.009 | 0.002 | -5.488 | 4.06E-08 | -0.003 | 0.005 | -0.676 | 4.99E-01 |
| rs11782313 | PTK2 | rs13254151 | 0.95 | 0.98 | 8 | 141925310 | T | C | 0.009 | 0.002 | 5.497 | 3.86E-08 | 0.003 | 0.005 | 0.676 | 4.99E-01 |
| rs11782313 | PTK2 | rs10111363 | 0.95 | 0.98 | 8 | 141928350 | A | C | 0.009 | 0.002 | 5.464 | 4.66E-08 | 0.003 | 0.005 | 0.686 | 4.92E-01 |
| rs11782313 | PTK2 | rs10103265 | 0.95 | 0.98 | 8 | 141934938 | T | C | -0.009 | 0.002 | -5.478 | 4.30E-08 | -0.003 | 0.005 | -0.676 | 4.99E-01 |
| rs11782313 | PTK2 | rs13250232 | 0.95 | 0.98 | 8 | 141937206 | T | C | 0.009 | 0.002 | 5.474 | 4.40E-08 | 0.003 | 0.005 | 0.684 | 4.94E-01 |
| rs11782313 | PTK2 | rs11781072 | 0.96 | 0.99 | 8 | 141938139 | T | C | -0.009 | 0.002 | -5.528 | 3.23E-08 | -0.003 | 0.005 | -0.684 | 4.94E-01 |
| rs11782313 | PTK2 | rs11786423 | 0.96 | 0.99 | 8 | 141939570 | T | C | 0.009 | 0.002 | 5.478 | 4.30E-08 | 0.003 | 0.005 | 0.676 | 4.99E-01 |
| rs11782313 | PTK2 | rs11784127 | 0.96 | 0.99 | 8 | 141941507 | T | C | -0.009 | 0.002 | -5.542 | 2.98E-08 | -0.003 | 0.005 | -0.676 | 4.99E-01 |
| rs11782313 | PTK2 | rs1349081 | 0.96 | 0.99 | 8 | 141944082 | T | C | 0.009 | 0.002 | 5.459 | 4.79E-08 | 0.003 | 0.005 | 0.676 | 4.99E-01 |

| | | | | | | | | | | | | | | | | |
|---|---|---|---|---|---|---|---|---|---|---|---|---|---|---|---|---|
| rs11782313 | PTK2 | rs13279233 | 0.96 | 0.99 | 8 | 141946474 | T | C | -0.009 | 0.002 | -5.473 | 4.42E-08 | -0.003 | 0.005 | -0.658 | 5.11E-01 |
| rs11782313 | PTK2 | rs9644449 | 0.96 | 0.99 | 8 | 141947285 | T | C | 0.009 | 0.002 | 5.456 | 4.87E-08 | 0.003 | 0.005 | 0.658 | 5.11E-01 |
| rs11782313 | PTK2 | rs11775547 | 0.96 | 0.99 | 8 | 141948448 | A | G | 0.009 | 0.002 | 5.474 | 4.40E-08 | 0.003 | 0.005 | 0.658 | 5.11E-01 |
| rs11782313 | PTK2 | rs11777476 | 0.97 | 0.99 | 8 | 141950375 | A | G | 0.009 | 0.002 | 5.454 | 4.92E-08 | 0.003 | 0.005 | 0.658 | 5.11E-01 |
| rs11782313 | PTK2 | rs11774024 | 0.97 | 0.99 | 8 | 141950534 | T | C | -0.009 | 0.002 | -5.454 | 4.92E-08 | -0.003 | 0.005 | -0.658 | 5.11E-01 |
| rs11782313 | PTK2 | rs11167010 | 0.97 | 0.99 | 8 | 141954210 | A | G | -0.009 | 0.002 | -5.512 | 3.55E-08 | -0.003 | 0.005 | -0.647 | 5.17E-01 |
| rs11782313 | PTK2 | rs2166869 | 0.98 | 1 | 8 | 141960578 | T | C | 0.009 | 0.002 | 5.464 | 4.67E-08 | 0.003 | 0.005 | 0.658 | 5.11E-01 |
| rs11782313 | PTK2 | rs7843337 | 0.98 | 1 | 8 | 141962642 | T | C | 0.009 | 0.002 | 5.584 | 2.35E-08 | 0.003 | 0.005 | 0.658 | 5.11E-01 |
| rs11782313 | PTK2 | rs11782313 | 1 | 1 | 8 | 141964183 | T | C | -0.009 | 0.002 | -5.645 | 1.65E-08 | -0.003 | 0.005 | -0.658 | 5.11E-01 |
| rs11782313 | PTK2 | rs11992645 | 0.98 | 1 | 8 | 141965397 | A | G | 0.009 | 0.002 | 5.481 | 4.22E-08 | 0.003 | 0.005 | 0.658 | 5.11E-01 |
| rs11782313 | PTK2 | rs1375062 | 0.98 | 1 | 8 | 141965781 | T | G | -0.009 | 0.002 | -5.502 | 3.76E-08 | -0.003 | 0.005 | -0.658 | 5.11E-01 |
| rs11782313 | PTK2 | rs13259836 | 0.98 | 1 | 8 | 141965888 | A | G | 0.009 | 0.002 | 5.514 | 3.52E-08 | 0.003 | 0.005 | 0.658 | 5.11E-01 |
| rs11782313 | PTK2 | rs11997937 | 0.97 | 0.99 | 8 | 141967351 | A | G | -0.009 | 0.002 | -5.480 | 4.26E-08 | -0.003 | 0.005 | -0.682 | 4.95E-01 |
| rs11782313 | PTK2 | rs11775885 | 0.98 | 1 | 8 | 141969994 | A | G | -0.009 | 0.002 | -5.511 | 3.58E-08 | -0.003 | 0.005 | -0.631 | 5.28E-01 |
| rs11782313 | PTK2 | rs13280609 | 0.98 | 1 | 8 | 141971740 | A | G | -0.009 | 0.002 | -5.514 | 3.52E-08 | -0.003 | 0.005 | -0.613 | 5.40E-01 |
| rs11782313 | PTK2 | rs10103124 | 0.97 | 0.98 | 8 | 141972190 | T | C | -0.009 | 0.002 | -5.487 | 4.08E-08 | -0.003 | 0.005 | -0.631 | 5.28E-01 |
| rs11782313 | PTK2 | rs10092613 | 0.97 | 0.99 | 8 | 141972242 | T | G | 0.009 | 0.002 | 5.515 | 3.48E-08 | 0.003 | 0.005 | 0.613 | 5.40E-01 |
| rs11782313 | PTK2 | rs10089966 | 0.97 | 0.99 | 8 | 141972542 | A | C | 0.009 | 0.002 | 5.492 | 3.98E-08 | 0.003 | 0.005 | 0.594 | 5.53E-01 |
| rs11782313 | PTK2 | rs13265704 | 0.97 | 0.99 | 8 | 141973152 | T | C | -0.009 | 0.002 | -5.505 | 3.70E-08 | -0.003 | 0.005 | -0.622 | 5.34E-01 |
| rs11782313 | PTK2 | rs9324538 | 0.97 | 0.99 | 8 | 141976738 | A | G | 0.009 | 0.002 | 5.489 | 4.05E-08 | 0.003 | 0.005 | 0.640 | 5.22E-01 |
| rs11782313 | PTK2 | rs9324539 | 0.97 | 0.99 | 8 | 141976997 | T | C | -0.009 | 0.002 | -5.492 | 3.98E-08 | -0.003 | 0.005 | -0.640 | 5.22E-01 |
| rs11782313 | PTK2 | rs11990900 | 0.97 | 0.99 | 8 | 141977920 | T | C | -0.009 | 0.002 | -5.492 | 3.98E-08 | -0.003 | 0.005 | -0.640 | 5.22E-01 |
| rs11782313 | PTK2 | rs10087115 | 0.97 | 0.99 | 8 | 141978830 | T | C | -0.009 | 0.002 | -5.495 | 3.92E-08 | -0.003 | 0.005 | -0.640 | 5.22E-01 |
| rs11782313 | PTK2 | rs13278775 | 0.97 | 0.99 | 8 | 141981929 | A | G | -0.009 | 0.002 | -5.484 | 4.15E-08 | -0.003 | 0.005 | -0.686 | 4.93E-01 |
| rs11782313 | PTK2 | rs8180999 | 0.97 | 0.99 | 8 | 141982481 | A | G | -0.009 | 0.002 | -5.489 | 4.05E-08 | -0.003 | 0.005 | -0.686 | 4.93E-01 |
| rs11782313 | PTK2 | rs11786859 | 0.97 | 0.99 | 8 | 141984881 | T | C | -0.009 | 0.002 | -5.554 | 2.80E-08 | -0.003 | 0.005 | -0.681 | 4.96E-01 |
| rs11782313 | PTK2 | rs9324540 | 0.95 | 0.98 | 8 | 141990421 | T | G | 0.009 | 0.002 | 5.507 | 3.66E-08 | 0.003 | 0.005 | 0.583 | 5.60E-01 |
| rs11782313 | PTK2 | rs10099696 | 0.93 | 0.97 | 8 | 141990510 | A | G | 0.009 | 0.002 | 5.569 | 2.56E-08 | 0.003 | 0.005 | 0.583 | 5.60E-01 |
| rs11782313 | PTK2 | rs11780777 | 0.95 | 0.98 | 8 | 141993216 | T | G | 0.009 | 0.002 | 5.453 | 4.95E-08 | 0.005 | 0.005 | 1.101 | 2.71E-01 |
| rs11782313 | PTK2 | rs13276704 | 0.95 | 0.98 | 8 | 141993696 | T | C | -0.009 | 0.002 | -5.457 | 4.83E-08 | -0.005 | 0.005 | -1.073 | 2.83E-01 |

| | | | | | | | | | | | | | | | | |
|---|---|---|---|---|---|---|---|---|---|---|---|---|---|---|---|---|
| rs11782313 | PTK2 | rs13258775 | 0.95 | 0.98 | 8 | 141994240 | T | C | 0.009 | 0.002 | 5.463 | 4.67E-08 | 0.005 | 0.005 | 1.098 | 2.72E-01 |
| rs11782313 | PTK2 | rs11781807 | 0.95 | 0.98 | 8 | 141994393 | T | G | 0.009 | 0.002 | 5.498 | 3.84E-08 | 0.005 | 0.005 | 1.112 | 2.66E-01 |
| rs11782313 | PTK2 | rs11167017 | 0.95 | 0.98 | 8 | 142000071 | T | C | -0.009 | 0.002 | -5.485 | 4.13E-08 | -0.005 | 0.005 | -1.080 | 2.80E-01 |
| rs11782313 | PTK2 | rs11786709 | 0.95 | 0.98 | 8 | 142001948 | A | G | -0.009 | 0.002 | -5.451 | 5.00E-08 | -0.005 | 0.005 | -1.098 | 2.72E-01 |
| rs11782313 | PTK2 | rs11780104 | 0.94 | 0.97 | 8 | 142002107 | T | C | -0.009 | 0.002 | -5.470 | 4.50E-08 | -0.005 | 0.005 | -1.108 | 2.68E-01 |
| rs11782313 | PTK2 | rs10106750 | 0.95 | 0.98 | 8 | 142004909 | A | G | 0.009 | 0.002 | 5.497 | 3.86E-08 | 0.005 | 0.005 | 1.098 | 2.72E-01 |
| rs11782313 | PTK2 | rs11784642 | 0.93 | 0.97 | 8 | 142005392 | A | G | -0.009 | 0.002 | -5.492 | 3.98E-08 | -0.005 | 0.005 | -1.098 | 2.72E-01 |
| rs11782313 | PTK2 | rs11782305 | 0.95 | 0.98 | 8 | 142005671 | T | C | 0.009 | 0.002 | 5.453 | 4.95E-08 | 0.006 | 0.005 | 1.141 | 2.54E-01 |
| rs11782313 | PTK2 | rs11785430 | 0.95 | 0.98 | 8 | 142009527 | A | G | 0.009 | 0.002 | 5.486 | 4.11E-08 | 0.005 | 0.005 | 1.116 | 2.64E-01 |
| rs12211255 | FILIP1 | rs12211255 | 1 | 1 | 6 | 76188330 | A | C | 0.014 | 0.003 | 5.607 | 2.06E-08 | 0.014 | 0.008 | 1.693 | 9.04E-02 |
| rs12908004 | ARNT2 | rs12908004 | 1 | 1 | 15 | 80676925 | A | G | -0.015 | 0.002 | -7.347 | 2.03E-13 | -0.004 | 0.006 | -0.658 | 5.10E-01 |
| rs13010313 | TEX41 | rs1438897 | 0.92 | 0.96 | 2 | 145674958 | A | G | 0.014 | 0.003 | 5.505 | 3.69E-08 | 0.018 | 0.008 | 2.150 | **3.15E-02** |
| rs13010313 | TEX41 | rs11679718 | 0.93 | 0.97 | 2 | 145681935 | A | G | 0.015 | 0.003 | 5.613 | 1.99E-08 | 0.018 | 0.009 | 2.142 | **3.22E-02** |
| rs13010313 | TEX41 | rs56795645 | 0.89 | 0.97 | 2 | 145703760 | T | G | -0.014 | 0.003 | -5.499 | 3.81E-08 | -0.014 | 0.008 | -1.725 | 8.46E-02 |
| rs13010313 | TEX41 | rs11689317 | 0.99 | 1 | 2 | 145710234 | A | G | -0.014 | 0.003 | -5.481 | 4.22E-08 | -0.014 | 0.008 | -1.726 | 8.44E-02 |
| rs13010313 | TEX41 | rs13010313 | 1 | 1 | 2 | 145721524 | T | C | 0.015 | 0.003 | 5.673 | 1.41E-08 | 0.014 | 0.008 | 1.717 | 8.60E-02 |
| rs13010313 | TEX41 | rs34948552 | 1 | 1 | 2 | 145722371 | T | C | 0.014 | 0.003 | 5.578 | 2.43E-08 | 0.014 | 0.008 | 1.717 | 8.60E-02 |
| rs13010313 | TEX41 | rs12621647 | 0.93 | 0.99 | 2 | 145734946 | A | G | 0.014 | 0.003 | 5.577 | 2.45E-08 | 0.014 | 0.008 | 1.709 | 8.75E-02 |
| rs13191450 | HSF2 | rs1402538 | 0.84 | 0.99 | 6 | 122388851 | A | G | -0.011 | 0.002 | -7.094 | 1.31E-12 | 0.000 | 0.005 | -0.090 | 9.28E-01 |
| rs13191450 | HSF2 | rs939601 | 0.99 | 1 | 6 | 122389643 | T | C | 0.012 | 0.002 | 7.640 | 2.17E-14 | -0.002 | 0.005 | -0.423 | 6.72E-01 |
| rs13191450 | HSF2 | rs868155 | 0.99 | 1 | 6 | 122389906 | T | C | -0.012 | 0.002 | -7.570 | 3.74E-14 | 0.002 | 0.005 | 0.435 | 6.63E-01 |
| rs13191450 | HSF2 | rs868154 | 0.99 | 1 | 6 | 122389940 | A | G | -0.012 | 0.002 | -7.453 | 9.13E-14 | 0.002 | 0.005 | 0.435 | 6.63E-01 |
| rs13191450 | HSF2 | rs868153 | 0.99 | 1 | 6 | 122389955 | T | G | 0.012 | 0.002 | 7.535 | 4.87E-14 | -0.002 | 0.005 | -0.435 | 6.63E-01 |
| rs13191450 | HSF2 | rs1357051 | 0.85 | 0.99 | 6 | 122391084 | T | C | -0.011 | 0.002 | -6.864 | 6.70E-12 | 0.000 | 0.005 | -0.042 | 9.66E-01 |
| rs13191450 | HSF2 | rs1357050 | 0.99 | 1 | 6 | 122391135 | T | G | 0.012 | 0.002 | 7.537 | 4.82E-14 | -0.002 | 0.005 | -0.423 | 6.72E-01 |
| rs13191450 | HSF2 | rs2259327 | 0.99 | 1 | 6 | 122391745 | T | C | -0.012 | 0.002 | -7.534 | 4.93E-14 | 0.002 | 0.005 | 0.405 | 6.85E-01 |
| rs13191450 | HSF2 | rs13191450 | 1 | 1 | 6 | 122392136 | A | C | 0.013 | 0.002 | 7.654 | 1.95E-14 | -0.002 | 0.005 | -0.371 | 7.11E-01 |
| rs13191450 | HSF2 | rs17084154 | 1 | 1 | 6 | 122393444 | T | C | -0.012 | 0.002 | -7.594 | 3.10E-14 | 0.003 | 0.005 | 0.529 | 5.97E-01 |
| rs13191450 | HSF2 | rs17084158 | 0.99 | 1 | 6 | 122393642 | A | C | -0.010 | 0.002 | -6.052 | 1.43E-09 | 0.003 | 0.005 | 0.588 | 5.57E-01 |
| rs13191450 | HSF2 | rs2176990 | 0.98 | 0.99 | 6 | 122393718 | T | C | -0.012 | 0.002 | -7.450 | 9.35E-14 | 0.003 | 0.005 | 0.549 | 5.83E-01 |

| | | | | | | | | | | | | | | | | |
|---|---|---|---|---|---|---|---|---|---|---|---|---|---|---|---|---|
| rs13191450 | HSF2 | rs6938223 | 0.99 | 1 | 6 | 122394272 | T | C | -0.012 | 0.002 | -7.504 | 6.17E-14 | 0.003 | 0.005 | 0.526 | 5.99E-01 |
| rs13191450 | HSF2 | rs59064689 | 0.99 | 1 | 6 | 122395180 | T | G | -0.012 | 0.002 | -7.493 | 6.73E-14 | 0.003 | 0.005 | 0.526 | 5.99E-01 |
| rs13191450 | HSF2 | rs6901617 | 0.99 | 0.99 | 6 | 122395469 | T | C | -0.012 | 0.002 | -7.490 | 6.88E-14 | 0.003 | 0.005 | 0.526 | 5.99E-01 |
| rs13191450 | HSF2 | rs2816097 | 0.96 | 0.99 | 6 | 122395647 | T | C | 0.012 | 0.002 | 7.171 | 7.47E-13 | -0.002 | 0.005 | -0.434 | 6.64E-01 |
| rs13191450 | HSF2 | rs6930575 | 0.98 | 0.99 | 6 | 122396000 | T | C | -0.012 | 0.002 | -7.460 | 8.68E-14 | 0.003 | 0.005 | 0.526 | 5.99E-01 |
| rs13191450 | HSF2 | rs72966339 | 0.98 | 0.99 | 6 | 122398241 | T | C | -0.012 | 0.002 | -7.550 | 4.34E-14 | 0.002 | 0.005 | 0.393 | 6.95E-01 |
| rs13191450 | HSF2 | rs34283499 | 0.98 | 0.99 | 6 | 122402574 | T | C | -0.012 | 0.002 | -7.501 | 6.36E-14 | 0.003 | 0.005 | 0.539 | 5.90E-01 |
| rs13191450 | HSF2 | rs13195459 | 0.98 | 0.99 | 6 | 122403559 | A | G | -0.012 | 0.002 | -7.502 | 6.30E-14 | 0.003 | 0.005 | 0.537 | 5.91E-01 |
| rs13191450 | HSF2 | rs6907870 | 0.97 | 0.99 | 6 | 122407844 | T | C | 0.012 | 0.002 | 7.403 | 1.33E-13 | -0.003 | 0.005 | -0.601 | 5.48E-01 |
| rs13191450 | HSF2 | rs6928224 | 0.96 | 0.99 | 6 | 122407982 | T | C | -0.012 | 0.002 | -7.386 | 1.51E-13 | 0.004 | 0.005 | 0.734 | 4.63E-01 |
| rs13191450 | HSF2 | rs13219206 | 0.95 | 0.98 | 6 | 122414157 | T | C | -0.012 | 0.002 | -7.350 | 1.98E-13 | 0.004 | 0.005 | 0.759 | 4.48E-01 |
| rs13191450 | HSF2 | rs17084191 | 0.94 | 0.98 | 6 | 122419081 | T | G | 0.012 | 0.002 | 7.383 | 1.55E-13 | -0.005 | 0.005 | -0.921 | 3.57E-01 |
| rs13191450 | HSF2 | rs13196347 | 0.92 | 0.97 | 6 | 122421142 | A | G | -0.012 | 0.002 | -7.564 | 3.90E-14 | 0.005 | 0.005 | 0.920 | 3.57E-01 |
| rs17042098 | PITX2 | rs17042098 | 1 | 1 | 4 | 111664158 | A | G | 0.078 | 0.002 | 32.862 | 7.73E-237 | 0.013 | 0.007 | 1.965 | **4.94E-02** |
| rs17171711 | FAM13B, KLHL3, | rs11957898 | 0.8 | 0.91 | 5 | 137217270 | T | C | -0.021 | 0.002 | -10.166 | 2.82E-24 | 0.000 | 0.006 | -0.076 | 9.40E-01 |
| rs17171711 | FAM13B, KLHL3, | rs76734697 | 0.84 | 0.92 | 5 | 137233469 | T | G | 0.021 | 0.002 | 10.413 | 2.16E-25 | 0.002 | 0.007 | 0.339 | 7.34E-01 |
| rs17171711 | FAM13B, KLHL3, | rs10076361 | 0.94 | 0.97 | 5 | 137252940 | A | G | 0.022 | 0.002 | 10.541 | 5.60E-26 | 0.007 | 0.006 | 1.033 | 3.01E-01 |
| rs17171711 | FAM13B, KLHL3, | rs73300168 | 0.98 | 0.99 | 5 | 137270655 | T | C | -0.022 | 0.002 | -10.486 | 1.01E-25 | -0.007 | 0.006 | -1.084 | 2.78E-01 |
| rs17171711 | FAM13B, KLHL3, | rs73299210 | 0.98 | 0.99 | 5 | 137282806 | A | G | 0.022 | 0.002 | 10.471 | 1.17E-25 | 0.006 | 0.006 | 0.984 | 3.25E-01 |
| rs17171711 | FAM13B, KLHL3, | rs73299219 | 0.98 | 0.99 | 5 | 137285940 | A | G | 0.022 | 0.002 | 10.478 | 1.09E-25 | 0.006 | 0.006 | 0.984 | 3.25E-01 |
| rs17171711 | FAM13B, KLHL3, | rs13362264 | 0.98 | 0.99 | 5 | 137299438 | A | G | 0.021 | 0.002 | 10.365 | 3.59E-25 | 0.006 | 0.006 | 0.974 | 3.30E-01 |
| rs17171711 | FAM13B, KLHL3, | rs78081438 | 0.91 | 0.99 | 5 | 137299621 | T | C | -0.022 | 0.002 | -10.504 | 8.32E-26 | -0.006 | 0.006 | -0.974 | 3.30E-01 |
| rs17171711 | FAM13B, KLHL3, | rs73299268 | 0.98 | 0.99 | 5 | 137328323 | A | G | -0.022 | 0.002 | -10.458 | 1.34E-25 | -0.006 | 0.006 | -0.976 | 3.29E-01 |
| rs17171711 | FAM13B, KLHL3, | rs11959181 | 1 | 1 | 5 | 137340815 | A | G | -0.022 | 0.002 | -10.470 | 1.19E-25 | -0.007 | 0.006 | -1.025 | 3.06E-01 |
| rs17171711 | FAM13B, KLHL3, | rs9327807 | 1 | 1 | 5 | 137350478 | T | C | -0.022 | 0.002 | -10.526 | 6.57E-26 | -0.007 | 0.006 | -1.012 | 3.11E-01 |
| rs17171711 | FAM13B, KLHL3, | rs113331339 | 1 | 1 | 5 | 137351159 | A | C | -0.022 | 0.002 | -10.470 | 1.19E-25 | -0.007 | 0.006 | -1.012 | 3.11E-01 |
| rs17171711 | FAM13B, KLHL3, | rs17171711 | 1 | 1 | 5 | 137364795 | T | C | 0.022 | 0.002 | 10.571 | 4.04E-26 | 0.006 | 0.006 | 0.916 | 3.60E-01 |
| rs17171711 | FAM13B, KLHL3, | rs77915370 | 0.99 | 1 | 5 | 137370990 | A | G | 0.022 | 0.002 | 10.527 | 6.50E-26 | 0.006 | 0.006 | 0.914 | 3.61E-01 |
| rs17171711 | FAM13B, KLHL3, | rs13355516 | 0.99 | 1 | 5 | 137380603 | A | G | -0.022 | 0.002 | -10.549 | 5.11E-26 | -0.006 | 0.006 | -0.964 | 3.35E-01 |
| rs17171711 | FAM13B, KLHL3, | rs148378888 | 0.99 | 1 | 5 | 137383078 | A | C | 0.022 | 0.002 | 10.530 | 6.25E-26 | 0.006 | 0.006 | 0.914 | 3.61E-01 |

| | | | | | | | | | | | | | | | | |
|---|---|---|---|---|---|---|---|---|---|---|---|---|---|---|---|---|
| rs17171711 | FAM13B, KLHL3, | rs2040862 | 0.93 | 0.97 | 5 | 137419989 | T | C | 0.022 | 0.002 | 10.499 | 8.74E-26 | 0.006 | 0.006 | 0.928 | 3.54E-01 |
| rs1898096 | LRMDA | rs1898097 | 1 | 1 | 10 | 77925497 | A | G | 0.011 | 0.002 | 5.541 | 3.01E-08 | 0.003 | 0.006 | 0.481 | 6.31E-01 |
| rs1898096 | LRMDA | rs11001652 | 1 | 1 | 10 | 77926280 | A | G | -0.011 | 0.002 | -5.512 | 3.56E-08 | -0.004 | 0.006 | -0.638 | 5.24E-01 |
| rs1898096 | LRMDA | rs4360646 | 1 | 1 | 10 | 77927296 | A | G | -0.011 | 0.002 | -5.498 | 3.84E-08 | -0.004 | 0.006 | -0.638 | 5.24E-01 |
| rs1898096 | LRMDA | rs2395343 | 1 | 1 | 10 | 77927404 | A | G | -0.011 | 0.002 | -5.455 | 4.90E-08 | -0.004 | 0.006 | -0.574 | 5.66E-01 |
| rs1898096 | LRMDA | rs4362088 | 1 | 1 | 10 | 77927525 | A | G | -0.011 | 0.002 | -5.460 | 4.75E-08 | -0.004 | 0.006 | -0.575 | 5.65E-01 |
| rs1898096 | LRMDA | rs11001655 | 1 | 1 | 10 | 77927556 | A | G | -0.011 | 0.002 | -5.454 | 4.93E-08 | -0.004 | 0.006 | -0.575 | 5.65E-01 |
| rs1898096 | LRMDA | rs33997838 | 1 | 1 | 10 | 77928304 | A | G | -0.011 | 0.002 | -5.463 | 4.69E-08 | -0.004 | 0.006 | -0.638 | 5.24E-01 |
| rs1898096 | LRMDA | rs1898096 | 1 | 1 | 10 | 77928689 | A | G | -0.011 | 0.002 | -5.601 | 2.13E-08 | -0.003 | 0.006 | -0.479 | 6.32E-01 |
| rs1898096 | LRMDA | rs11001660 | 1 | 1 | 10 | 77929725 | T | C | -0.011 | 0.002 | -5.500 | 3.79E-08 | -0.002 | 0.006 | -0.356 | 7.21E-01 |
| rs1898096 | LRMDA | rs12569600 | 1 | 1 | 10 | 77930390 | T | C | -0.011 | 0.002 | -5.495 | 3.91E-08 | -0.003 | 0.006 | -0.421 | 6.74E-01 |
| rs1898096 | LRMDA | rs1898093 | 1 | 1 | 10 | 77930464 | A | G | -0.011 | 0.002 | -5.516 | 3.47E-08 | -0.003 | 0.006 | -0.520 | 6.03E-01 |
| rs1898096 | LRMDA | rs1898092 | 1 | 1 | 10 | 77930600 | A | G | -0.011 | 0.002 | -5.582 | 2.37E-08 | -0.003 | 0.006 | -0.421 | 6.74E-01 |
| rs1898096 | LRMDA | rs12245788 | 1 | 1 | 10 | 77932402 | T | C | -0.011 | 0.002 | -5.473 | 4.44E-08 | -0.003 | 0.006 | -0.423 | 6.72E-01 |
| rs1898096 | LRMDA | rs11001666 | 1 | 1 | 10 | 77933210 | A | G | -0.011 | 0.002 | -5.503 | 3.73E-08 | -0.002 | 0.006 | -0.324 | 7.46E-01 |
| rs1898096 | LRMDA | rs4746371 | 0.99 | 1 | 10 | 77934487 | T | C | 0.011 | 0.002 | 5.474 | 4.39E-08 | 0.003 | 0.006 | 0.438 | 6.61E-01 |
| rs1898096 | LRMDA | rs4433517 | 0.99 | 1 | 10 | 77934866 | T | C | 0.011 | 0.002 | 5.466 | 4.61E-08 | 0.003 | 0.006 | 0.503 | 6.15E-01 |
| rs1898096 | LRMDA | rs11001667 | 0.99 | 1 | 10 | 77935345 | A | G | -0.011 | 0.002 | -5.550 | 2.85E-08 | -0.002 | 0.006 | -0.386 | 7.00E-01 |
| rs2106261 | ZFHX3 | rs4404097 | 0.81 | 1 | 16 | 73032032 | A | G | 0.036 | 0.002 | 16.319 | 7.21E-60 | 0.013 | 0.007 | 1.897 | 5.78E-02 |
| rs2106261 | ZFHX3 | rs79891915 | 0.81 | 1 | 16 | 73032033 | T | C | 0.036 | 0.002 | 16.354 | 4.10E-60 | 0.013 | 0.007 | 1.897 | 5.78E-02 |
| rs2106261 | ZFHX3 | rs57828240 | 0.86 | 0.99 | 16 | 73041940 | T | G | -0.035 | 0.002 | -16.424 | 1.28E-60 | -0.013 | 0.007 | -1.911 | 5.61E-02 |
| rs2106261 | ZFHX3 | rs12051512 | 0.86 | 0.99 | 16 | 73043777 | A | G | 0.035 | 0.002 | 16.496 | 3.91E-61 | 0.014 | 0.007 | 2.008 | **4.47E-02** |
| rs2106261 | ZFHX3 | rs67402452 | 0.82 | 0.94 | 16 | 73046957 | T | C | 0.033 | 0.002 | 15.878 | 8.97E-57 | 0.009 | 0.007 | 1.305 | 1.92E-01 |
| rs2106261 | ZFHX3 | rs67329386 | 0.99 | 1 | 16 | 73048367 | T | C | 0.036 | 0.002 | 17.335 | 2.57E-67 | 0.014 | 0.007 | 2.164 | **3.05E-02** |
| rs2106261 | ZFHX3 | rs2106261 | 1 | 1 | 16 | 73051620 | T | C | 0.036 | 0.002 | 17.480 | 2.02E-68 | 0.014 | 0.007 | 2.148 | **3.17E-02** |
| rs2106261 | ZFHX3 | rs4499262 | 0.97 | 0.99 | 16 | 73059159 | A | C | 0.034 | 0.002 | 16.490 | 4.36E-61 | 0.014 | 0.007 | 2.131 | **3.31E-02** |
| rs2106261 | ZFHX3 | rs879324 | 0.91 | 0.97 | 16 | 73068678 | A | G | 0.035 | 0.002 | 17.059 | 2.98E-65 | 0.012 | 0.007 | 1.792 | 7.31E-02 |
| rs2106261 | ZFHX3 | rs12932445 | 0.91 | 0.97 | 16 | 73069888 | T | C | -0.035 | 0.002 | -17.259 | 9.58E-67 | -0.013 | 0.007 | -1.966 | **4.93E-02** |
| rs2269001 | KCNH2 | rs758887 | 0.86 | 0.93 | 7 | 150634448 | T | C | -0.010 | 0.002 | -5.508 | 3.64E-08 | -0.001 | 0.006 | -0.228 | 8.19E-01 |
| rs2269001 | KCNH2 | rs13228494 | 0.84 | 0.93 | 7 | 150637094 | A | G | 0.011 | 0.002 | 5.866 | 4.48E-09 | 0.002 | 0.006 | 0.280 | 7.79E-01 |

| | | | | | | | | | | | | | | | | |
|---|---|---|---|---|---|---|---|---|---|---|---|---|---|---|---|---|
| rs2269001 | KCNH2 | rs4725982 | 0.86 | 0.93 | 7 | 150637863 | T | C | -0.010 | 0.002 | -5.527 | 3.26E-08 | -0.002 | 0.006 | -0.272 | 7.86E-01 |
| rs2269001 | KCNH2 | rs3815459 | 0.85 | 0.93 | 7 | 150644394 | T | C | -0.011 | 0.002 | -5.944 | 2.77E-09 | -0.001 | 0.006 | -0.171 | 8.64E-01 |
| rs2269001 | KCNH2 | rs1805120 | 0.93 | 0.98 | 7 | 150649531 | A | G | -0.011 | 0.002 | -5.966 | 2.43E-09 | -0.001 | 0.006 | -0.136 | 8.92E-01 |
| rs2269001 | KCNH2 | rs740952 | 0.91 | 0.97 | 7 | 150649603 | A | G | -0.011 | 0.002 | -5.684 | 1.31E-08 | -0.003 | 0.006 | -0.537 | 5.91E-01 |
| rs2269001 | KCNH2 | rs2269000 | 0.96 | 1 | 7 | 150650792 | A | G | -0.011 | 0.002 | -6.027 | 1.67E-09 | -0.003 | 0.006 | -0.534 | 5.93E-01 |
| rs2269001 | KCNH2 | rs2269001 | 1 | 1 | 7 | 150651816 | A | G | -0.012 | 0.002 | -6.109 | 1.00E-09 | -0.003 | 0.006 | -0.592 | 5.54E-01 |
| rs2269001 | KCNH2 | rs34657537 | 0.95 | 0.98 | 7 | 150657808 | A | G | -0.011 | 0.002 | -5.942 | 2.82E-09 | -0.003 | 0.006 | -0.565 | 5.72E-01 |
| rs2269001 | KCNH2 | rs3823587 | 0.96 | 0.99 | 7 | 150657890 | A | C | 0.011 | 0.002 | 5.937 | 2.90E-09 | 0.003 | 0.006 | 0.578 | 5.63E-01 |
| rs2269001 | KCNH2 | rs3807376 | 0.95 | 0.98 | 7 | 150658199 | T | C | 0.011 | 0.002 | 5.991 | 2.08E-09 | 0.003 | 0.006 | 0.544 | 5.86E-01 |
| rs2269001 | KCNH2 | rs35760656 | 0.96 | 0.99 | 7 | 150658678 | A | G | -0.011 | 0.002 | -5.855 | 4.76E-09 | -0.002 | 0.006 | -0.297 | 7.67E-01 |
| rs2269001 | KCNH2 | rs3778874 | 0.95 | 0.98 | 7 | 150659051 | A | G | -0.011 | 0.002 | -5.843 | 5.12E-09 | -0.002 | 0.006 | -0.273 | 7.84E-01 |
| rs2738413 | SYNE2 | rs1255984 | 0.82 | 0.92 | 14 | 64654431 | A | G | 0.014 | 0.002 | 9.010 | 2.05E-19 | 0.011 | 0.005 | 2.275 | **2.29E-02** |
| rs2738413 | SYNE2 | rs1255986 | 0.82 | 0.92 | 14 | 64655161 | T | C | -0.015 | 0.002 | -9.300 | 1.41E-20 | -0.011 | 0.005 | -2.223 | **2.62E-02** |
| rs2738413 | SYNE2 | rs1255988 | 0.82 | 0.92 | 14 | 64657684 | A | C | -0.015 | 0.002 | -9.339 | 9.68E-21 | -0.010 | 0.005 | -2.071 | **3.84E-02** |
| rs2738413 | SYNE2 | rs2738413 | 1 | 1 | 14 | 64679960 | A | G | 0.015 | 0.002 | 9.524 | 1.67E-21 | 0.012 | 0.005 | 2.553 | **1.07E-02** |
| rs2738413 | SYNE2 | rs1152591 | 0.99 | 1 | 14 | 64680848 | A | G | 0.015 | 0.002 | 9.279 | 1.71E-20 | 0.013 | 0.005 | 2.668 | **7.64E-03** |
| rs2834618 | RUNX1 | rs2834618 | 1 | 1 | 21 | 36119111 | T | G | 0.019 | 0.002 | 7.562 | 3.96E-14 | 0.011 | 0.008 | 1.407 | 1.59E-01 |
| rs2857265 | TTN | rs16866358 | 0.93 | 0.97 | 2 | 179380096 | A | G | -0.018 | 0.002 | -7.772 | 7.72E-15 | -0.001 | 0.007 | -0.169 | 8.66E-01 |
| rs2857265 | TTN | rs13029629 | 0.94 | 0.98 | 2 | 179380165 | A | G | -0.018 | 0.002 | -7.770 | 7.86E-15 | 0.000 | 0.007 | -0.070 | 9.44E-01 |
| rs2857265 | TTN | rs12618265 | 0.94 | 0.98 | 2 | 179382809 | T | G | -0.018 | 0.002 | -7.761 | 8.41E-15 | -0.001 | 0.007 | -0.101 | 9.20E-01 |
| rs2857265 | TTN | rs13034990 | 0.94 | 0.98 | 2 | 179386240 | A | G | -0.018 | 0.002 | -7.709 | 1.26E-14 | 0.000 | 0.007 | -0.036 | 9.71E-01 |
| rs2857265 | TTN | rs7597154 | 1 | 1 | 2 | 179388490 | A | G | 0.018 | 0.002 | 8.236 | 1.78E-16 | 0.000 | 0.007 | -0.023 | 9.82E-01 |
| rs2857265 | TTN | rs13005601 | 1 | 1 | 2 | 179388657 | A | G | 0.018 | 0.002 | 8.148 | 3.70E-16 | 0.000 | 0.007 | -0.023 | 9.82E-01 |
| rs2857265 | TTN | rs16866369 | 1 | 1 | 2 | 179388994 | A | G | -0.018 | 0.002 | -8.100 | 5.48E-16 | 0.000 | 0.007 | 0.023 | 9.82E-01 |
| rs2857265 | TTN | rs3829748 | 1 | 1 | 2 | 179396354 | A | G | 0.018 | 0.002 | 8.255 | 1.51E-16 | 0.001 | 0.007 | 0.092 | 9.27E-01 |
| rs2857265 | TTN | rs3829747 | 1 | 1 | 2 | 179397561 | T | C | 0.019 | 0.002 | 8.272 | 1.31E-16 | 0.001 | 0.007 | 0.124 | 9.01E-01 |
| rs2857265 | TTN | rs2857265 | 1 | 1 | 2 | 179398823 | A | G | 0.019 | 0.002 | 8.424 | 3.65E-17 | 0.001 | 0.007 | 0.105 | 9.17E-01 |
| rs2857265 | TTN | rs2303539 | 1 | 1 | 2 | 179403593 | A | G | 0.018 | 0.002 | 8.157 | 3.42E-16 | 0.001 | 0.007 | 0.163 | 8.70E-01 |
| rs2857265 | TTN | rs3731751 | 1 | 1 | 2 | 179405885 | T | C | -0.018 | 0.002 | -8.134 | 4.16E-16 | -0.001 | 0.007 | -0.107 | 9.15E-01 |
| rs2857265 | TTN | rs3731749 | 1 | 1 | 2 | 179406191 | T | C | 0.018 | 0.002 | 8.152 | 3.57E-16 | 0.001 | 0.007 | 0.107 | 9.15E-01 |

| | | | | | | | | | | | | | | | | |
|---|---|---|---|---|---|---|---|---|---|---|---|---|---|---|---|---|
| rs2857265 | TTN | rs3731748 | 1 | 1 | 2 | 179413110 | A | G | 0.018 | 0.002 | 8.227 | 1.92E-16 | 0.000 | 0.007 | 0.027 | 9.78E-01 |
| rs2857265 | TTN | rs16866397 | 0.96 | 0.98 | 2 | 179421470 | T | G | -0.018 | 0.002 | -8.155 | 3.49E-16 | 0.000 | 0.007 | 0.001 | 9.99E-01 |
| rs2857265 | TTN | rs744426 | 0.96 | 0.98 | 2 | 179436020 | A | G | 0.018 | 0.002 | 8.184 | 2.74E-16 | 0.001 | 0.007 | 0.081 | 9.36E-01 |
| rs2857265 | TTN | rs1007025 | 0.97 | 0.99 | 2 | 179451660 | A | G | -0.018 | 0.002 | -8.230 | 1.87E-16 | -0.001 | 0.007 | -0.127 | 8.99E-01 |
| rs2857265 | TTN | rs16866406 | 0.97 | 0.99 | 2 | 179457147 | A | G | 0.018 | 0.002 | 8.050 | 8.28E-16 | 0.001 | 0.007 | 0.136 | 8.92E-01 |
| rs2857265 | TTN | rs2288569 | 0.97 | 0.99 | 2 | 179458591 | T | C | 0.018 | 0.002 | 8.014 | 1.11E-15 | 0.001 | 0.007 | 0.135 | 8.92E-01 |
| rs2885697 | SCMH1 | rs2885697 | 1 | 1 | 1 | 41544279 | T | G | -0.009 | 0.002 | -5.672 | 1.41E-08 | -0.004 | 0.005 | -0.749 | 4.54E-01 |
| rs3176326 | CDKN1A | rs3176326 | 1 | 1 | 6 | 36647289 | A | G | -0.014 | 0.002 | -7.225 | 5.01E-13 | -0.007 | 0.006 | -1.219 | 2.23E-01 |
| rs337705 | KCNN2 | rs337705 | 1 | 1 | 5 | 113737062 | T | G | -0.011 | 0.002 | -7.051 | 1.77E-12 | 0.003 | 0.005 | 0.544 | 5.86E-01 |
| rs337705 | KCNN2 | rs163210 | 0.99 | 0.99 | 5 | 113745738 | T | C | -0.011 | 0.002 | -6.761 | 1.37E-11 | 0.003 | 0.005 | 0.584 | 5.59E-01 |
| rs337705 | KCNN2 | rs337708 | 0.99 | 0.99 | 5 | 113746180 | A | G | -0.011 | 0.002 | -6.951 | 3.62E-12 | 0.003 | 0.005 | 0.602 | 5.47E-01 |
| rs337705 | KCNN2 | rs337711 | 0.98 | 0.99 | 5 | 113748571 | T | C | 0.011 | 0.002 | 6.891 | 5.54E-12 | -0.003 | 0.005 | -0.581 | 5.61E-01 |
| rs337705 | KCNN2 | rs168090 | 0.98 | 0.99 | 5 | 113749946 | T | C | -0.011 | 0.002 | -6.815 | 9.45E-12 | 0.003 | 0.005 | 0.602 | 5.47E-01 |
| rs337705 | KCNN2 | rs6859284 | 0.96 | 0.98 | 5 | 113757965 | A | G | -0.011 | 0.002 | -6.805 | 1.01E-11 | 0.003 | 0.005 | 0.533 | 5.94E-01 |
| rs337705 | KCNN2 | rs337683 | 0.95 | 0.98 | 5 | 113759435 | A | G | 0.011 | 0.002 | 6.858 | 6.97E-12 | -0.002 | 0.005 | -0.390 | 6.96E-01 |
| rs337705 | KCNN2 | rs338623 | 0.96 | -0.98 | 5 | 113764982 | A | G | 0.011 | 0.002 | 6.901 | 5.16E-12 | -0.002 | 0.005 | -0.349 | 7.27E-01 |
| rs3746471 | KIAA1755 | rs3746471 | 1 | 1 | 20 | 36841914 | A | G | 0.009 | 0.002 | 5.514 | 3.50E-08 | 0.011 | 0.005 | 2.236 | **2.53E-02** |
| rs3746471 | KIAA1755 | rs6127466 | 0.99 | 1 | 20 | 36842883 | A | G | -0.009 | 0.002 | -5.480 | 4.25E-08 | -0.011 | 0.005 | -2.236 | **2.53E-02** |
| rs3807989 | CAV1 | rs3807989 | 1 | 1 | 7 | 116186241 | A | G | -0.021 | 0.002 | -13.097 | 3.43E-39 | -0.015 | 0.005 | -3.187 | **1.44E-03** |
| rs3807989 | CAV1 | rs11773845 | 0.99 | 1 | 7 | 116191301 | A | C | 0.021 | 0.002 | 12.953 | 2.25E-38 | 0.015 | 0.005 | 3.166 | **1.55E-03** |
| rs3807989 | CAV1 | rs1997571 | 0.93 | 0.97 | 7 | 116198621 | A | G | 0.020 | 0.002 | 12.803 | 1.57E-37 | 0.015 | 0.005 | 3.147 | **1.65E-03** |
| rs3807989 | CAV1 | rs1997572 | 0.93 | 0.97 | 7 | 116198828 | A | G | -0.019 | 0.002 | -12.004 | 3.39E-33 | -0.015 | 0.005 | -3.139 | **1.70E-03** |
| rs41292535 | EPS15 | rs56284497 | 0.92 | 0.96 | 1 | 51808467 | T | C | 0.030 | 0.005 | 6.632 | 3.31E-11 | 0.004 | 0.018 | 0.217 | 8.28E-01 |
| rs41292535 | EPS15 | rs6673415 | 0.92 | 0.96 | 1 | 51809479 | A | G | -0.030 | 0.005 | -6.623 | 3.52E-11 | -0.004 | 0.018 | -0.217 | 8.28E-01 |
| rs41292535 | EPS15 | rs57519873 | 0.92 | 0.96 | 1 | 51811185 | T | C | 0.030 | 0.005 | 6.664 | 2.66E-11 | 0.004 | 0.018 | 0.217 | 8.28E-01 |
| rs41292535 | EPS15 | rs56176004 | 0.92 | 0.96 | 1 | 51814797 | T | C | 0.031 | 0.005 | 6.817 | 9.29E-12 | 0.002 | 0.018 | 0.109 | 9.13E-01 |
| rs41292535 | EPS15 | rs72694121 | 0.92 | 0.96 | 1 | 51815091 | T | C | 0.030 | 0.005 | 6.616 | 3.70E-11 | 0.002 | 0.018 | 0.109 | 9.13E-01 |
| rs41292535 | EPS15 | rs72694122 | 0.92 | 0.96 | 1 | 51815124 | A | C | 0.030 | 0.005 | 6.638 | 3.17E-11 | 0.001 | 0.018 | 0.070 | 9.44E-01 |
| rs41292535 | EPS15 | rs55915918 | 0.92 | 0.96 | 1 | 51815588 | T | C | -0.030 | 0.005 | -6.663 | 2.68E-11 | -0.001 | 0.018 | -0.070 | 9.44E-01 |
| rs41292535 | EPS15 | rs142608110 | 0.92 | 0.96 | 1 | 51816759 | T | C | -0.031 | 0.005 | -6.814 | 9.52E-12 | -0.001 | 0.018 | -0.070 | 9.44E-01 |

| | | | | | | | | | | | | | | | | |
|---|---|---|---|---|---|---|---|---|---|---|---|---|---|---|---|---|
| rs41292535 | EPS15 | rs7538988 | 0.92 | 0.96 | 1 | 51817541 | T | C | -0.031 | 0.005 | -6.816 | 9.39E-12 | -0.001 | 0.018 | -0.070 | 9.44E-01 |
| rs41292535 | EPS15 | rs55643430 | 0.92 | 0.96 | 1 | 51817581 | T | G | 0.031 | 0.005 | 6.812 | 9.64E-12 | 0.001 | 0.018 | 0.070 | 9.44E-01 |
| rs41292535 | EPS15 | rs55880270 | 0.92 | 0.96 | 1 | 51818481 | A | G | -0.030 | 0.005 | -6.670 | 2.57E-11 | -0.001 | 0.018 | -0.070 | 9.44E-01 |
| rs41292535 | EPS15 | rs76782211 | 0.92 | 0.96 | 1 | 51820415 | A | G | 0.031 | 0.005 | 6.810 | 9.78E-12 | 0.001 | 0.018 | 0.070 | 9.44E-01 |
| rs41292535 | EPS15 | rs72694128 | 1 | 1 | 1 | 51826210 | T | C | 0.031 | 0.005 | 6.801 | 1.04E-11 | 0.001 | 0.018 | 0.070 | 9.44E-01 |
| rs41292535 | EPS15 | rs72694130 | 1 | 1 | 1 | 51828128 | A | G | -0.030 | 0.005 | -6.647 | 2.99E-11 | -0.001 | 0.018 | -0.070 | 9.44E-01 |
| rs41292535 | EPS15 | rs55671617 | 1 | 1 | 1 | 51828324 | A | C | -0.030 | 0.005 | -6.624 | 3.49E-11 | -0.001 | 0.018 | -0.070 | 9.44E-01 |
| rs41292535 | EPS15 | rs6664884 | 1 | 1 | 1 | 51828498 | T | G | 0.030 | 0.005 | 6.572 | 4.97E-11 | 0.001 | 0.018 | 0.070 | 9.44E-01 |
| rs41292535 | EPS15 | rs6656341 | 1 | 1 | 1 | 51828499 | T | C | -0.029 | 0.005 | -6.398 | 1.57E-10 | -0.001 | 0.018 | -0.070 | 9.44E-01 |
| rs41292535 | EPS15 | rs6702800 | 1 | 1 | 1 | 51828925 | A | G | -0.030 | 0.005 | -6.686 | 2.29E-11 | -0.001 | 0.018 | -0.070 | 9.44E-01 |
| rs41292535 | EPS15 | rs72694131 | 1 | 1 | 1 | 51829281 | T | G | -0.031 | 0.005 | -6.791 | 1.12E-11 | -0.001 | 0.018 | -0.070 | 9.44E-01 |
| rs41292535 | EPS15 | rs77742485 | 1 | 1 | 1 | 51831570 | T | C | -0.030 | 0.005 | -6.645 | 3.02E-11 | -0.001 | 0.018 | -0.070 | 9.44E-01 |
| rs41292535 | EPS15 | rs72694138 | 1 | 1 | 1 | 51832943 | A | G | 0.030 | 0.005 | 6.648 | 2.97E-11 | 0.001 | 0.018 | 0.070 | 9.44E-01 |
| rs41292535 | EPS15 | rs56064204 | 1 | 1 | 1 | 51833554 | A | G | 0.030 | 0.005 | 6.654 | 2.85E-11 | 0.003 | 0.018 | 0.141 | 8.88E-01 |
| rs41292535 | EPS15 | rs7522189 | 1 | 1 | 1 | 51834521 | A | G | 0.030 | 0.005 | 6.591 | 4.36E-11 | 0.003 | 0.018 | 0.141 | 8.88E-01 |
| rs41292535 | EPS15 | rs72694142 | 1 | 1 | 1 | 51835307 | T | G | 0.031 | 0.005 | 6.794 | 1.09E-11 | 0.003 | 0.018 | 0.141 | 8.88E-01 |
| rs41292535 | EPS15 | rs113539819 | 1 | 1 | 1 | 51836510 | T | C | 0.030 | 0.005 | 6.655 | 2.84E-11 | 0.003 | 0.018 | 0.141 | 8.88E-01 |
| rs41292535 | EPS15 | rs72694143 | 1 | 1 | 1 | 51837697 | A | C | -0.030 | 0.005 | -6.647 | 2.99E-11 | -0.003 | 0.018 | -0.141 | 8.88E-01 |
| rs41292535 | EPS15 | rs7535155 | 1 | 1 | 1 | 51838677 | A | G | 0.029 | 0.005 | 6.443 | 1.17E-10 | 0.003 | 0.018 | 0.141 | 8.88E-01 |
| rs41292535 | EPS15 | rs72694144 | 1 | 1 | 1 | 51840447 | A | G | 0.030 | 0.005 | 6.646 | 3.00E-11 | 0.003 | 0.018 | 0.141 | 8.88E-01 |
| rs41292535 | EPS15 | rs139600178 | 1 | 1 | 1 | 51840953 | A | G | 0.031 | 0.005 | 6.796 | 1.07E-11 | 0.003 | 0.018 | 0.141 | 8.88E-01 |
| rs41292535 | EPS15 | rs6664504 | 1 | 1 | 1 | 51841328 | T | C | -0.031 | 0.005 | -6.791 | 1.12E-11 | -0.003 | 0.018 | -0.141 | 8.88E-01 |
| rs41292535 | EPS15 | rs112444602 | 1 | 1 | 1 | 51844060 | A | G | 0.031 | 0.005 | 6.786 | 1.15E-11 | 0.005 | 0.018 | 0.264 | 7.92E-01 |
| rs41292535 | EPS15 | rs1002721 | 1 | 1 | 1 | 51844346 | T | C | 0.029 | 0.005 | 6.466 | 1.00E-10 | 0.005 | 0.018 | 0.264 | 7.92E-01 |
| rs41292535 | EPS15 | rs1002720 | 1 | 1 | 1 | 51844358 | A | C | -0.029 | 0.005 | -6.458 | 1.06E-10 | -0.005 | 0.018 | -0.264 | 7.92E-01 |
| rs41292535 | EPS15 | rs72694150 | 1 | 1 | 1 | 51845562 | T | C | 0.031 | 0.005 | 6.782 | 1.19E-11 | 0.005 | 0.018 | 0.264 | 7.92E-01 |
| rs41292535 | EPS15 | rs72694151 | 1 | 1 | 1 | 51846983 | A | G | -0.030 | 0.005 | -6.623 | 3.52E-11 | -0.005 | 0.018 | -0.264 | 7.92E-01 |
| rs41292535 | EPS15 | rs111550988 | 1 | 1 | 1 | 51847213 | T | C | -0.030 | 0.005 | -6.646 | 3.02E-11 | -0.005 | 0.018 | -0.264 | 7.92E-01 |
| rs41292535 | EPS15 | rs72694153 | 1 | 1 | 1 | 51847351 | T | C | -0.031 | 0.005 | -6.781 | 1.19E-11 | -0.005 | 0.018 | -0.264 | 7.92E-01 |
| rs41292535 | EPS15 | rs56120161 | 1 | 1 | 1 | 51848151 | T | C | -0.030 | 0.005 | -6.581 | 4.66E-11 | -0.005 | 0.018 | -0.264 | 7.92E-01 |

| | | | | | | | | | | | | | | | | |
|---|---|---|---|---|---|---|---|---|---|---|---|---|---|---|---|---|
| rs41292535 | EPS15 | rs72694159 | 1 | 1 | 1 | 51848949 | T | C | -0.030 | 0.005 | -6.646 | 3.02E-11 | -0.005 | 0.018 | -0.264 | 7.92E-01 |
| rs41292535 | EPS15 | rs72694161 | 1 | 1 | 1 | 51850525 | A | G | 0.031 | 0.005 | 6.795 | 1.09E-11 | 0.004 | 0.018 | 0.238 | 8.12E-01 |
| rs41292535 | EPS15 | rs6694227 | 1 | 1 | 1 | 51850801 | T | C | 0.030 | 0.005 | 6.644 | 3.05E-11 | 0.005 | 0.018 | 0.264 | 7.92E-01 |
| rs41292535 | EPS15 | rs72694164 | 1 | 1 | 1 | 51851417 | T | C | -0.030 | 0.005 | -6.650 | 2.93E-11 | -0.005 | 0.018 | -0.264 | 7.92E-01 |
| rs41292535 | EPS15 | rs72694166 | 1 | 1 | 1 | 51851785 | T | C | 0.031 | 0.005 | 6.784 | 1.17E-11 | 0.005 | 0.018 | 0.264 | 7.92E-01 |
| rs41292535 | EPS15 | rs72694167 | 1 | 1 | 1 | 51852096 | T | C | -0.030 | 0.005 | -6.625 | 3.47E-11 | -0.005 | 0.018 | -0.264 | 7.92E-01 |
| rs41292535 | EPS15 | rs72694171 | 1 | 1 | 1 | 51857917 | T | C | -0.030 | 0.005 | -6.653 | 2.87E-11 | -0.005 | 0.018 | -0.264 | 7.92E-01 |
| rs41292535 | EPS15 | rs72694173 | 1 | 1 | 1 | 51858450 | T | C | 0.030 | 0.005 | 6.638 | 3.17E-11 | 0.005 | 0.018 | 0.264 | 7.92E-01 |
| rs41292535 | EPS15 | rs114032030 | 1 | 1 | 1 | 51860679 | T | C | 0.030 | 0.005 | 6.640 | 3.14E-11 | 0.005 | 0.018 | 0.253 | 8.00E-01 |
| rs41292535 | EPS15 | rs72694177 | 1 | 1 | 1 | 51860790 | T | C | -0.030 | 0.005 | -6.642 | 3.08E-11 | -0.005 | 0.018 | -0.253 | 8.00E-01 |
| rs41292535 | EPS15 | rs72694183 | 1 | 1 | 1 | 51865403 | T | C | 0.030 | 0.005 | 6.724 | 1.77E-11 | 0.005 | 0.018 | 0.253 | 8.00E-01 |
| rs41292535 | EPS15 | rs74898383 | 1 | 1 | 1 | 51869004 | T | C | -0.031 | 0.005 | -6.789 | 1.13E-11 | -0.005 | 0.018 | -0.253 | 8.00E-01 |
| rs41292535 | EPS15 | rs140074095 | 1 | 1 | 1 | 51869828 | A | C | 0.031 | 0.005 | 6.786 | 1.15E-11 | 0.005 | 0.018 | 0.253 | 8.00E-01 |
| rs41292535 | EPS15 | rs149891466 | 1 | 1 | 1 | 51869846 | A | G | 0.031 | 0.005 | 6.790 | 1.12E-11 | 0.005 | 0.018 | 0.253 | 8.00E-01 |
| rs41292535 | EPS15 | rs72694189 | 1 | 1 | 1 | 51870525 | A | G | -0.030 | 0.005 | -6.647 | 2.99E-11 | -0.005 | 0.018 | -0.253 | 8.00E-01 |
| rs41292535 | EPS15 | rs72694190 | 1 | 1 | 1 | 51870729 | A | C | -0.030 | 0.005 | -6.650 | 2.94E-11 | -0.005 | 0.018 | -0.253 | 8.00E-01 |
| rs41292535 | EPS15 | rs72694192 | 1 | 1 | 1 | 51873381 | T | C | 0.030 | 0.005 | 6.651 | 2.92E-11 | 0.005 | 0.018 | 0.253 | 8.00E-01 |
| rs41292535 | EPS15 | rs72694193 | 1 | 1 | 1 | 51873395 | T | G | 0.030 | 0.005 | 6.650 | 2.93E-11 | 0.005 | 0.018 | 0.253 | 8.00E-01 |
| rs41292535 | EPS15 | rs143637015 | 1 | 1 | 1 | 51874860 | T | C | -0.031 | 0.005 | -6.785 | 1.16E-11 | -0.005 | 0.018 | -0.253 | 8.00E-01 |
| rs41292535 | EPS15 | rs139036273 | 1 | 1 | 1 | 51875805 | A | G | 0.031 | 0.005 | 6.790 | 1.12E-11 | 0.005 | 0.018 | 0.253 | 8.00E-01 |
| rs41292535 | EPS15 | rs59715648 | 1 | 1 | 1 | 51876499 | T | C | -0.030 | 0.005 | -6.654 | 2.85E-11 | -0.005 | 0.018 | -0.253 | 8.00E-01 |
| rs41292535 | EPS15 | rs60782272 | 1 | 1 | 1 | 51876749 | T | C | 0.030 | 0.005 | 6.652 | 2.88E-11 | 0.005 | 0.018 | 0.253 | 8.00E-01 |
| rs41292535 | EPS15 | rs72694198 | 1 | 1 | 1 | 51876782 | T | C | -0.031 | 0.005 | -6.783 | 1.18E-11 | -0.005 | 0.018 | -0.253 | 8.00E-01 |
| rs41292535 | EPS15 | rs55972866 | 1 | 1 | 1 | 51877590 | T | C | -0.030 | 0.005 | -6.653 | 2.87E-11 | -0.005 | 0.018 | -0.253 | 8.00E-01 |
| rs41292535 | EPS15 | rs7526469 | 1 | 1 | 1 | 51878785 | T | C | -0.031 | 0.005 | -6.790 | 1.12E-11 | -0.005 | 0.018 | -0.253 | 8.00E-01 |
| rs41292535 | EPS15 | rs72694200 | 1 | 1 | 1 | 51878994 | A | G | 0.030 | 0.005 | 6.654 | 2.85E-11 | 0.005 | 0.018 | 0.253 | 8.00E-01 |
| rs41292535 | EPS15 | rs77910451 | 1 | 1 | 1 | 51879033 | A | G | -0.031 | 0.005 | -6.790 | 1.12E-11 | -0.005 | 0.018 | -0.253 | 8.00E-01 |
| rs41292535 | EPS15 | rs111876348 | 1 | 1 | 1 | 51879565 | A | C | 0.030 | 0.005 | 6.654 | 2.85E-11 | 0.005 | 0.018 | 0.253 | 8.00E-01 |
| rs41292535 | EPS15 | rs111307188 | 0.92 | 1 | 1 | 51881357 | A | G | -0.030 | 0.005 | -6.660 | 2.74E-11 | -0.005 | 0.018 | -0.264 | 7.92E-01 |
| rs41292535 | EPS15 | rs72696103 | 1 | 1 | 1 | 51887120 | A | G | -0.030 | 0.005 | -6.658 | 2.78E-11 | -0.005 | 0.018 | -0.264 | 7.92E-01 |

| | | | | | | | | | | | | | | | | |
|---|---|---|---|---|---|---|---|---|---|---|---|---|---|---|---|---|
| rs41292535 | EPS15 | rs72696107 | 1 | 1 | 1 | 51889202 | A | C | 0.030 | 0.005 | 6.660 | 2.74E-11 | 0.005 | 0.018 | 0.264 | 7.92E-01 |
| rs41292535 | EPS15 | rs72696108 | 1 | 1 | 1 | 51889585 | A | G | -0.031 | 0.005 | -6.800 | 1.04E-11 | -0.005 | 0.018 | -0.264 | 7.92E-01 |
| rs41292535 | EPS15 | rs72696110 | 1 | 1 | 1 | 51889891 | T | C | -0.030 | 0.005 | -6.661 | 2.72E-11 | -0.005 | 0.018 | -0.264 | 7.92E-01 |
| rs41292535 | EPS15 | rs58867548 | 1 | 1 | 1 | 51890625 | A | C | 0.030 | 0.005 | 6.664 | 2.67E-11 | 0.005 | 0.018 | 0.264 | 7.92E-01 |
| rs41292535 | EPS15 | rs72696112 | 1 | 1 | 1 | 51890793 | A | C | -0.031 | 0.005 | -6.797 | 1.07E-11 | -0.005 | 0.018 | -0.264 | 7.92E-01 |
| rs41292535 | EPS15 | rs7553078 | 1 | 1 | 1 | 51897733 | T | C | -0.030 | 0.005 | -6.618 | 3.63E-11 | -0.003 | 0.018 | -0.163 | 8.70E-01 |
| rs41292535 | EPS15 | rs6588409 | 1 | 1 | 1 | 51901022 | T | C | 0.031 | 0.005 | 6.794 | 1.09E-11 | 0.003 | 0.018 | 0.163 | 8.70E-01 |
| rs41292535 | EPS15 | rs72696122 | 1 | 1 | 1 | 51902791 | T | G | 0.030 | 0.005 | 6.669 | 2.58E-11 | 0.003 | 0.018 | 0.163 | 8.70E-01 |
| rs41292535 | EPS15 | rs56251532 | 1 | 1 | 1 | 51903430 | T | C | 0.030 | 0.005 | 6.672 | 2.52E-11 | 0.003 | 0.018 | 0.163 | 8.70E-01 |
| rs41292535 | EPS15 | rs72696126 | 1 | 1 | 1 | 51905540 | A | G | 0.030 | 0.005 | 6.673 | 2.50E-11 | 0.003 | 0.018 | 0.163 | 8.70E-01 |
| rs41292535 | EPS15 | rs72696128 | 1 | 1 | 1 | 51907312 | A | C | -0.031 | 0.005 | -6.812 | 9.60E-12 | -0.003 | 0.018 | -0.163 | 8.70E-01 |
| rs41292535 | EPS15 | rs72696129 | 1 | 1 | 1 | 51908076 | T | C | -0.030 | 0.005 | -6.673 | 2.50E-11 | -0.003 | 0.018 | -0.163 | 8.70E-01 |
| rs41292535 | EPS15 | rs55654180 | 1 | 1 | 1 | 51911785 | A | G | 0.030 | 0.005 | 6.690 | 2.23E-11 | 0.003 | 0.018 | 0.163 | 8.70E-01 |
| rs41292535 | EPS15 | rs187495104 | 0.96 | 1 | 1 | 51912157 | T | C | -0.030 | 0.005 | -6.677 | 2.43E-11 | -0.003 | 0.018 | -0.163 | 8.70E-01 |
| rs41292535 | EPS15 | rs7526429 | 1 | 1 | 1 | 51915027 | A | G | -0.029 | 0.005 | -6.483 | 8.97E-11 | -0.003 | 0.018 | -0.163 | 8.70E-01 |
| rs41292535 | EPS15 | rs78108947 | 1 | 1 | 1 | 51915697 | T | C | -0.030 | 0.005 | -6.690 | 2.24E-11 | -0.003 | 0.018 | -0.163 | 8.70E-01 |
| rs41292535 | EPS15 | rs72696134 | 1 | 1 | 1 | 51915699 | A | C | 0.031 | 0.005 | 6.821 | 9.06E-12 | 0.003 | 0.018 | 0.163 | 8.70E-01 |
| rs41292535 | EPS15 | rs72696135 | 1 | 1 | 1 | 51916365 | A | G | 0.030 | 0.005 | 6.678 | 2.42E-11 | 0.003 | 0.018 | 0.163 | 8.70E-01 |
| rs41292535 | EPS15 | rs56255751 | 1 | 1 | 1 | 51916782 | A | G | -0.031 | 0.005 | -6.823 | 8.93E-12 | -0.003 | 0.018 | -0.163 | 8.70E-01 |
| rs41292535 | EPS15 | rs72696137 | 0.84 | 0.96 | 1 | 51917244 | T | G | 0.030 | 0.005 | 6.641 | 3.12E-11 | 0.003 | 0.018 | 0.163 | 8.70E-01 |
| rs41292535 | EPS15 | rs72696138 | 1 | 1 | 1 | 51917538 | A | G | 0.031 | 0.005 | 6.779 | 1.21E-11 | 0.003 | 0.018 | 0.163 | 8.70E-01 |
| rs41292535 | EPS15 | rs7549556 | 1 | 1 | 1 | 51919252 | A | G | 0.029 | 0.005 | 6.490 | 8.58E-11 | 0.003 | 0.018 | 0.163 | 8.70E-01 |
| rs41292535 | EPS15 | rs7530091 | 1 | 1 | 1 | 51919263 | T | C | 0.029 | 0.005 | 6.490 | 8.58E-11 | 0.003 | 0.018 | 0.163 | 8.70E-01 |
| rs41292535 | EPS15 | rs72696145 | 1 | 1 | 1 | 51919816 | T | C | 0.030 | 0.005 | 6.679 | 2.40E-11 | 0.003 | 0.018 | 0.163 | 8.70E-01 |
| rs41292535 | EPS15 | rs111625476 | 1 | 1 | 1 | 51921203 | A | G | 0.030 | 0.005 | 6.641 | 3.11E-11 | 0.003 | 0.018 | 0.163 | 8.70E-01 |
| rs41292535 | EPS15 | rs72696148 | 1 | 1 | 1 | 51921439 | T | C | 0.030 | 0.005 | 6.615 | 3.71E-11 | 0.003 | 0.018 | 0.163 | 8.70E-01 |
| rs41292535 | EPS15 | rs72696149 | 1 | 1 | 1 | 51922078 | A | G | 0.030 | 0.005 | 6.687 | 2.28E-11 | 0.003 | 0.018 | 0.163 | 8.70E-01 |
| rs41292535 | EPS15 | rs183345174 | 0.96 | 1 | 1 | 51922774 | A | G | -0.030 | 0.005 | -6.692 | 2.21E-11 | -0.003 | 0.018 | -0.163 | 8.70E-01 |
| rs41292535 | EPS15 | rs72696151 | 1 | 1 | 1 | 51923360 | T | C | -0.029 | 0.005 | -6.489 | 8.67E-11 | -0.003 | 0.018 | -0.163 | 8.70E-01 |
| rs41292535 | EPS15 | rs56167325 | 0.92 | 1 | 1 | 51924159 | A | G | 0.030 | 0.005 | 6.677 | 2.44E-11 | 0.003 | 0.018 | 0.163 | 8.70E-01 |

| | | | | | | | | | | | | | | | | |
|---|---|---|---|---|---|---|---|---|---|---|---|---|---|---|---|---|
| rs41292535 | EPS15 | rs56261634 | 1 | 1 | 1 | 51924519 | A | G | 0.031 | 0.005 | 6.821 | 9.02E-12 | 0.003 | 0.018 | 0.163 | 8.70E-01 |
| rs41292535 | EPS15 | rs150449315 | 1 | 1 | 1 | 51925019 | A | G | 0.030 | 0.005 | 6.665 | 2.64E-11 | 0.003 | 0.018 | 0.163 | 8.70E-01 |
| rs41292535 | EPS15 | rs72696161 | 1 | 1 | 1 | 51927117 | T | C | -0.030 | 0.005 | -6.673 | 2.51E-11 | -0.003 | 0.018 | -0.163 | 8.70E-01 |
| rs41292535 | EPS15 | rs113436173 | 1 | 1 | 1 | 51927967 | A | G | -0.030 | 0.005 | -6.685 | 2.31E-11 | -0.003 | 0.018 | -0.163 | 8.70E-01 |
| rs41292535 | EPS15 | rs150288978 | 1 | 1 | 1 | 51929985 | T | C | 0.031 | 0.005 | 6.818 | 9.21E-12 | 0.003 | 0.018 | 0.163 | 8.70E-01 |
| rs41292535 | EPS15 | rs7512458 | 1 | 1 | 1 | 51930232 | A | C | -0.029 | 0.005 | -6.496 | 8.24E-11 | -0.003 | 0.018 | -0.163 | 8.70E-01 |
| rs41292535 | EPS15 | rs56326822 | 1 | 1 | 1 | 51931089 | T | C | 0.030 | 0.005 | 6.690 | 2.23E-11 | 0.003 | 0.018 | 0.163 | 8.70E-01 |
| rs41292535 | EPS15 | rs72679103 | 1 | 1 | 1 | 51933247 | T | C | 0.030 | 0.005 | 6.687 | 2.27E-11 | 0.003 | 0.018 | 0.163 | 8.70E-01 |
| rs41292535 | EPS15 | rs7553017 | 1 | 1 | 1 | 51933918 | A | C | -0.029 | 0.005 | -6.494 | 8.37E-11 | -0.003 | 0.018 | -0.163 | 8.70E-01 |
| rs41292535 | EPS15 | rs72679106 | 1 | 1 | 1 | 51935344 | A | C | -0.031 | 0.005 | -6.806 | 1.00E-11 | -0.003 | 0.018 | -0.163 | 8.70E-01 |
| rs41292535 | EPS15 | rs41292535 | 1 | 1 | 1 | 51938639 | A | G | 0.034 | 0.005 | 6.991 | 2.73E-12 | 0.003 | 0.018 | 0.163 | 8.70E-01 |
| rs41292535 | EPS15 | rs55672422 | 1 | 1 | 1 | 51942474 | A | G | -0.030 | 0.005 | -6.688 | 2.26E-11 | -0.003 | 0.018 | -0.163 | 8.70E-01 |
| rs41292535 | EPS15 | rs72679112 | 1 | 1 | 1 | 51945365 | A | G | 0.031 | 0.005 | 6.814 | 9.52E-12 | 0.003 | 0.018 | 0.163 | 8.70E-01 |
| rs41292535 | EPS15 | rs6665465 | 1 | 1 | 1 | 51946276 | T | C | 0.031 | 0.005 | 6.816 | 9.34E-12 | 0.003 | 0.018 | 0.163 | 8.70E-01 |
| rs41292535 | EPS15 | rs72679113 | 1 | 1 | 1 | 51946549 | T | C | -0.031 | 0.005 | -6.815 | 9.40E-12 | -0.003 | 0.018 | -0.163 | 8.70E-01 |
| rs41292535 | EPS15 | rs72679114 | 1 | 1 | 1 | 51948107 | A | C | -0.031 | 0.005 | -6.828 | 8.61E-12 | -0.003 | 0.018 | -0.163 | 8.70E-01 |
| rs41292535 | EPS15 | rs72691868 | 1 | 1 | 1 | 51949190 | A | G | 0.030 | 0.005 | 6.650 | 2.92E-11 | 0.003 | 0.018 | 0.163 | 8.70E-01 |
| rs41292535 | EPS15 | rs113039001 | 1 | 1 | 1 | 51949307 | A | G | 0.030 | 0.005 | 6.689 | 2.25E-11 | 0.003 | 0.018 | 0.163 | 8.70E-01 |
| rs41292535 | EPS15 | rs55868733 | 1 | 1 | 1 | 51949550 | T | C | -0.030 | 0.005 | -6.697 | 2.12E-11 | -0.003 | 0.018 | -0.163 | 8.70E-01 |
| rs41292535 | EPS15 | rs55667545 | 1 | 1 | 1 | 51949832 | A | G | 0.030 | 0.005 | 6.687 | 2.28E-11 | 0.003 | 0.018 | 0.163 | 8.70E-01 |
| rs41292535 | EPS15 | rs72691871 | 1 | 1 | 1 | 51950251 | A | G | 0.030 | 0.005 | 6.694 | 2.17E-11 | 0.003 | 0.018 | 0.163 | 8.70E-01 |
| rs41292535 | EPS15 | rs72691874 | 1 | 1 | 1 | 51950703 | A | C | 0.030 | 0.005 | 6.686 | 2.29E-11 | 0.003 | 0.018 | 0.163 | 8.70E-01 |
| rs41292535 | EPS15 | rs78715030 | 1 | 1 | 1 | 51952783 | T | C | -0.031 | 0.005 | -6.789 | 1.13E-11 | -0.003 | 0.018 | -0.163 | 8.70E-01 |
| rs41292535 | EPS15 | rs72691881 | 0.88 | 0.96 | 1 | 51954664 | A | G | 0.029 | 0.004 | 6.581 | 4.69E-11 | 0.003 | 0.018 | 0.163 | 8.70E-01 |
| rs41292535 | EPS15 | rs113544672 | 1 | 1 | 1 | 51955020 | A | G | 0.030 | 0.005 | 6.620 | 3.58E-11 | 0.003 | 0.018 | 0.163 | 8.70E-01 |
| rs41292535 | EPS15 | rs60698990 | 1 | 1 | 1 | 51955827 | A | G | 0.030 | 0.005 | 6.694 | 2.16E-11 | 0.001 | 0.018 | 0.049 | 9.61E-01 |
| rs41292535 | EPS15 | rs79339614 | 1 | 1 | 1 | 51956304 | A | C | -0.031 | 0.005 | -6.826 | 8.74E-12 | -0.001 | 0.018 | -0.049 | 9.61E-01 |
| rs41292535 | EPS15 | rs56360606 | 1 | 1 | 1 | 51957136 | T | C | -0.030 | 0.005 | -6.660 | 2.73E-11 | -0.001 | 0.018 | -0.049 | 9.61E-01 |
| rs41292535 | EPS15 | rs55691002 | 1 | 1 | 1 | 51957320 | T | C | -0.030 | 0.005 | -6.690 | 2.23E-11 | -0.001 | 0.018 | -0.049 | 9.61E-01 |
| rs41292535 | EPS15 | rs112576034 | 1 | 1 | 1 | 51958809 | A | G | 0.030 | 0.005 | 6.687 | 2.28E-11 | 0.001 | 0.018 | 0.049 | 9.61E-01 |

| | | | | | | | | | | | | | | | | |
|---|---|---|---|---|---|---|---|---|---|---|---|---|---|---|---|---|
| rs41292535 | EPS15 | rs111280073 | 1 | 1 | 1 | 51959476 | A | G | 0.030 | 0.005 | 6.683 | 2.34E-11 | 0.001 | 0.018 | 0.049 | 9.61E-01 |
| rs41292535 | EPS15 | rs6685946 | 1 | 1 | 1 | 51963530 | T | C | -0.029 | 0.005 | -6.493 | 8.43E-11 | -0.001 | 0.018 | -0.049 | 9.61E-01 |
| rs41292535 | EPS15 | rs72691891 | 1 | 1 | 1 | 51965500 | A | G | -0.031 | 0.005 | -6.817 | 9.30E-12 | 0.000 | 0.018 | -0.007 | 9.94E-01 |
| rs41292535 | EPS15 | rs59302433 | 1 | 1 | 1 | 51966611 | A | G | 0.030 | 0.005 | 6.679 | 2.40E-11 | 0.000 | 0.018 | 0.007 | 9.94E-01 |
| rs41292535 | EPS15 | rs112167117 | 1 | 1 | 1 | 51966943 | A | G | 0.030 | 0.005 | 6.679 | 2.40E-11 | 0.000 | 0.018 | 0.007 | 9.94E-01 |
| rs41292535 | EPS15 | rs72691894 | 1 | 1 | 1 | 51967137 | T | G | 0.030 | 0.005 | 6.672 | 2.53E-11 | 0.000 | 0.018 | 0.007 | 9.94E-01 |
| rs41292535 | EPS15 | rs113518234 | 1 | 1 | 1 | 51967299 | T | C | -0.030 | 0.005 | -6.657 | 2.79E-11 | 0.000 | 0.018 | -0.007 | 9.94E-01 |
| rs41292535 | EPS15 | rs72674557 | 1 | 1 | 1 | 51969336 | A | C | -0.030 | 0.005 | -6.680 | 2.39E-11 | 0.000 | 0.018 | -0.007 | 9.94E-01 |
| rs41292535 | EPS15 | rs6657654 | 1 | 1 | 1 | 51969417 | T | C | -0.031 | 0.005 | -6.814 | 9.50E-12 | 0.000 | 0.018 | -0.007 | 9.94E-01 |
| rs41292535 | EPS15 | rs72674558 | 1 | 1 | 1 | 51970450 | A | C | 0.030 | 0.005 | 6.667 | 2.60E-11 | 0.000 | 0.018 | 0.007 | 9.94E-01 |
| rs41292535 | EPS15 | rs72674559 | 1 | 1 | 1 | 51970511 | A | C | -0.030 | 0.005 | -6.663 | 2.69E-11 | 0.000 | 0.018 | -0.007 | 9.94E-01 |
| rs41292535 | EPS15 | rs61007089 | 1 | 1 | 1 | 51971600 | T | C | 0.030 | 0.005 | 6.632 | 3.31E-11 | 0.000 | 0.018 | 0.007 | 9.94E-01 |
| rs41292535 | EPS15 | rs60375184 | 1 | 1 | 1 | 51974344 | T | C | -0.030 | 0.005 | -6.672 | 2.51E-11 | 0.000 | 0.018 | -0.007 | 9.94E-01 |
| rs41292535 | EPS15 | rs72674566 | 1 | 1 | 1 | 51975948 | T | C | 0.030 | 0.005 | 6.678 | 2.43E-11 | 0.000 | 0.018 | 0.007 | 9.94E-01 |
| rs41292535 | EPS15 | rs55831531 | 1 | 1 | 1 | 51976520 | A | G | 0.030 | 0.005 | 6.673 | 2.51E-11 | 0.000 | 0.018 | 0.007 | 9.94E-01 |
| rs41292535 | EPS15 | rs7514142 | 1 | 1 | 1 | 51979294 | A | C | 0.030 | 0.005 | 6.683 | 2.34E-11 | 0.000 | 0.018 | 0.007 | 9.94E-01 |
| rs41292535 | EPS15 | rs7526085 | 1 | 1 | 1 | 51979625 | A | G | -0.030 | 0.005 | -6.670 | 2.56E-11 | 0.000 | 0.018 | -0.007 | 9.94E-01 |
| rs41292535 | EPS15 | rs72674575 | 1 | 1 | 1 | 51980230 | T | C | -0.030 | 0.005 | -6.667 | 2.61E-11 | 0.000 | 0.018 | -0.007 | 9.94E-01 |
| rs41292535 | EPS15 | rs72674578 | 1 | 1 | 1 | 51981761 | T | C | 0.030 | 0.005 | 6.644 | 3.05E-11 | 0.000 | 0.018 | 0.007 | 9.94E-01 |
| rs41292535 | EPS15 | rs12039831 | 1 | 1 | 1 | 51983466 | T | C | -0.030 | 0.004 | -6.725 | 1.76E-11 | -0.005 | 0.017 | -0.286 | 7.75E-01 |
| rs41292535 | EPS15 | rs78094029 | 1 | 1 | 1 | 51987313 | A | G | 0.031 | 0.005 | 6.795 | 1.09E-11 | 0.000 | 0.018 | 0.007 | 9.94E-01 |
| rs41292535 | EPS15 | rs72674582 | 0.89 | 1 | 1 | 51988100 | A | G | 0.030 | 0.004 | 6.687 | 2.27E-11 | 0.000 | 0.018 | 0.007 | 9.94E-01 |
| rs41292535 | EPS15 | rs56084508 | 1 | 1 | 1 | 51989293 | A | G | 0.030 | 0.005 | 6.660 | 2.74E-11 | 0.000 | 0.018 | 0.007 | 9.94E-01 |
| rs41292535 | EPS15 | rs56157924 | 0.96 | 1 | 1 | 51989746 | T | C | 0.030 | 0.004 | 6.812 | 9.60E-12 | -0.001 | 0.017 | -0.048 | 9.61E-01 |
| rs41292535 | EPS15 | rs72674586 | 1 | 1 | 1 | 51990321 | T | G | -0.031 | 0.005 | -6.792 | 1.10E-11 | 0.000 | 0.018 | -0.007 | 9.94E-01 |
| rs41292535 | EPS15 | rs72674590 | 1 | 1 | 1 | 51991008 | T | C | -0.031 | 0.005 | -6.791 | 1.12E-11 | 0.000 | 0.018 | -0.007 | 9.94E-01 |
| rs41292535 | EPS15 | rs7533508 | 1 | 1 | 1 | 51992305 | A | C | -0.029 | 0.005 | -6.435 | 1.23E-10 | 0.000 | 0.018 | -0.007 | 9.94E-01 |
| rs41292535 | EPS15 | rs12027854 | 1 | 1 | 1 | 51994888 | A | C | 0.030 | 0.004 | 6.817 | 9.31E-12 | 0.005 | 0.017 | 0.286 | 7.75E-01 |
| rs41292535 | EPS15 | rs12045600 | 1 | 1 | 1 | 51994894 | T | G | -0.030 | 0.004 | -6.818 | 9.25E-12 | -0.005 | 0.017 | -0.286 | 7.75E-01 |
| rs41292535 | EPS15 | rs7543230 | 1 | 1 | 1 | 51995495 | A | C | -0.029 | 0.004 | -6.521 | 6.98E-11 | -0.004 | 0.017 | -0.249 | 8.03E-01 |

| | | | | | | | | | | | | | | | | |
|---|---|---|---|---|---|---|---|---|---|---|---|---|---|---|---|---|
| rs41292535 | EPS15 | rs12026316 | 1 | 1 | 1 | 51998263 | T | C | 0.030 | 0.004 | 6.829 | 8.56E-12 | 0.005 | 0.017 | 0.286 | 7.75E-01 |
| rs41292535 | EPS15 | rs56280508 | 1 | 1 | 1 | 52000331 | T | C | 0.030 | 0.004 | 6.671 | 2.55E-11 | 0.008 | 0.017 | 0.445 | 6.56E-01 |
| rs41292535 | EPS15 | rs72674597 | 1 | 1 | 1 | 52000390 | A | G | 0.030 | 0.004 | 6.682 | 2.36E-11 | 0.008 | 0.017 | 0.445 | 6.56E-01 |
| rs41292535 | EPS15 | rs72661386 | 1 | 1 | 1 | 52002010 | A | G | 0.031 | 0.004 | 6.852 | 7.27E-12 | 0.008 | 0.017 | 0.445 | 6.56E-01 |
| rs41292535 | EPS15 | rs12022046 | 1 | 1 | 1 | 52004497 | T | C | -0.030 | 0.004 | -6.692 | 2.21E-11 | -0.008 | 0.017 | -0.445 | 6.56E-01 |
| rs41292535 | EPS15 | rs72661388 | 1 | 1 | 1 | 52004550 | T | C | -0.030 | 0.004 | -6.705 | 2.01E-11 | -0.008 | 0.017 | -0.445 | 6.56E-01 |
| rs41292535 | EPS15 | rs12562916 | 0.89 | 1 | 1 | 52010097 | A | G | -0.029 | 0.004 | -6.683 | 2.34E-11 | -0.018 | 0.015 | -1.239 | 2.15E-01 |
| rs41292535 | EPS15 | rs148643004 | 0.89 | 1 | 1 | 52010557 | T | C | 0.029 | 0.004 | 6.632 | 3.30E-11 | 0.018 | 0.015 | 1.227 | 2.20E-01 |
| rs4385527 | AOPEP | rs7851726 | 0.82 | -0.91 | 9 | 97503985 | T | C | 0.016 | 0.002 | 10.202 | 1.95E-24 | 0.000 | 0.005 | 0.050 | 9.60E-01 |
| rs4385527 | AOPEP | rs7866848 | 0.81 | -0.92 | 9 | 97505608 | A | G | -0.016 | 0.002 | -9.790 | 1.24E-22 | 0.000 | 0.005 | 0.026 | 9.80E-01 |
| rs4385527 | AOPEP | rs446540 | 0.86 | -0.93 | 9 | 97509199 | A | G | 0.016 | 0.002 | 9.940 | 2.80E-23 | 0.000 | 0.005 | 0.066 | 9.47E-01 |
| rs4385527 | AOPEP | rs183066 | 0.87 | -0.94 | 9 | 97517432 | T | C | -0.016 | 0.002 | -9.926 | 3.20E-23 | 0.000 | 0.005 | -0.032 | 9.75E-01 |
| rs4385527 | AOPEP | rs368138 | 0.86 | -0.93 | 9 | 97523583 | A | G | 0.016 | 0.002 | 9.968 | 2.10E-23 | 0.000 | 0.005 | 0.028 | 9.78E-01 |
| rs4385527 | AOPEP | rs28540393 | 0.85 | -0.92 | 9 | 97527520 | T | C | 0.016 | 0.002 | 9.758 | 1.70E-22 | 0.000 | 0.005 | 0.032 | 9.75E-01 |
| rs4385527 | AOPEP | rs2584806 | 0.87 | -0.94 | 9 | 97529278 | A | C | -0.016 | 0.002 | -9.925 | 3.23E-23 | 0.000 | 0.005 | -0.096 | 9.24E-01 |
| rs4385527 | AOPEP | rs7019720 | 0.8 | -0.92 | 9 | 97539488 | A | G | 0.015 | 0.002 | 9.612 | 7.08E-22 | 0.000 | 0.005 | -0.054 | 9.57E-01 |
| rs4385527 | AOPEP | rs356141 | 0.8 | -0.92 | 9 | 97547297 | A | C | 0.015 | 0.002 | 9.605 | 7.64E-22 | 0.000 | 0.005 | -0.071 | 9.43E-01 |
| rs4385527 | AOPEP | rs356131 | 0.8 | -0.92 | 9 | 97567327 | A | G | 0.015 | 0.002 | 9.599 | 8.06E-22 | -0.001 | 0.005 | -0.138 | 8.90E-01 |
| rs4385527 | AOPEP | rs4744378 | 0.85 | 0.92 | 9 | 97579146 | A | G | 0.016 | 0.002 | 9.916 | 3.54E-23 | 0.000 | 0.005 | -0.056 | 9.55E-01 |
| rs4385527 | AOPEP | rs35487382 | 0.84 | 0.93 | 9 | 97585788 | A | G | -0.015 | 0.002 | -9.635 | 5.69E-22 | 0.000 | 0.005 | 0.088 | 9.30E-01 |
| rs4385527 | AOPEP | rs10821401 | 0.89 | 0.96 | 9 | 97592688 | A | G | 0.016 | 0.002 | 9.956 | 2.38E-23 | 0.000 | 0.005 | -0.062 | 9.51E-01 |
| rs4385527 | AOPEP | rs10993365 | 0.93 | 0.98 | 9 | 97600392 | A | G | 0.016 | 0.002 | 9.901 | 4.12E-23 | 0.000 | 0.005 | -0.062 | 9.51E-01 |
| rs4385527 | AOPEP | rs10993377 | 0.96 | 0.99 | 9 | 97630827 | T | C | 0.016 | 0.002 | 9.751 | 1.82E-22 | -0.001 | 0.005 | -0.153 | 8.78E-01 |
| rs4385527 | AOPEP | rs4385527 | 1 | 1 | 9 | 97648587 | A | G | 0.016 | 0.002 | 10.339 | 4.69E-25 | 0.000 | 0.005 | -0.080 | 9.36E-01 |
| rs4385527 | AOPEP | rs11787821 | 0.95 | 0.98 | 9 | 97660961 | T | C | 0.016 | 0.002 | 10.034 | 1.08E-23 | -0.001 | 0.005 | -0.110 | 9.12E-01 |
| rs4385527 | AOPEP | rs2183958 | 0.89 | 0.97 | 9 | 97669673 | A | G | -0.015 | 0.002 | -9.417 | 4.64E-21 | 0.001 | 0.005 | 0.130 | 8.96E-01 |
| rs4385527 | AOPEP | rs12235316 | 0.89 | 0.96 | 9 | 97677556 | A | G | -0.016 | 0.002 | -9.848 | 6.98E-23 | 0.001 | 0.005 | 0.263 | 7.93E-01 |
| rs4385527 | AOPEP | rs10993394 | 0.89 | 0.96 | 9 | 97678911 | A | G | 0.016 | 0.002 | 9.997 | 1.57E-23 | -0.001 | 0.005 | -0.227 | 8.21E-01 |
| rs4385527 | AOPEP | rs10993397 | 0.89 | 0.96 | 9 | 97679771 | T | C | 0.015 | 0.002 | 9.709 | 2.77E-22 | -0.001 | 0.005 | -0.264 | 7.91E-01 |
| rs4385527 | AOPEP | rs4744411 | 0.88 | 0.96 | 9 | 97689045 | A | G | 0.016 | 0.002 | 9.942 | 2.73E-23 | -0.001 | 0.005 | -0.248 | 8.04E-01 |

| | | | | | | | | | | | | | | | | |
|---|---|---|---|---|---|---|---|---|---|---|---|---|---|---|---|---|
| rs4385527 | AOPEP | rs1123534 | 0.88 | 0.95 | 9 | 97709725 | T | C | -0.015 | 0.002 | -9.721 | 2.45E-22 | 0.001 | 0.005 | 0.249 | 8.04E-01 |
| rs4385527 | AOPEP | rs10821415 | 0.87 | 0.94 | 9 | 97713459 | A | C | 0.016 | 0.002 | 10.060 | 8.28E-24 | -0.001 | 0.005 | -0.237 | 8.12E-01 |
| rs4385527 | AOPEP | rs7864171 | 0.86 | 0.93 | 9 | 97723266 | A | G | 0.016 | 0.002 | 9.928 | 3.14E-23 | -0.001 | 0.005 | -0.145 | 8.85E-01 |
| rs4385527 | AOPEP | rs7865239 | 0.89 | 0.94 | 9 | 97723449 | T | C | 0.016 | 0.002 | 9.792 | 1.22E-22 | 0.000 | 0.005 | -0.097 | 9.22E-01 |
| rs4385527 | AOPEP | rs10993418 | 0.9 | 0.96 | 9 | 97732256 | A | G | 0.016 | 0.002 | 9.792 | 1.22E-22 | 0.000 | 0.005 | 0.005 | 9.96E-01 |
| rs55754224 | CAMK2D | rs17620390 | 0.92 | 0.97 | 4 | 114384328 | A | C | -0.011 | 0.002 | -6.249 | 4.12E-10 | 0.000 | 0.005 | -0.073 | 9.42E-01 |
| rs55754224 | CAMK2D | rs35430511 | 0.94 | 0.97 | 4 | 114387138 | T | C | -0.011 | 0.002 | -6.366 | 1.93E-10 | -0.001 | 0.005 | -0.101 | 9.20E-01 |
| rs55754224 | CAMK2D | rs2285703 | 0.94 | 0.97 | 4 | 114388820 | A | G | -0.011 | 0.002 | -6.357 | 2.06E-10 | -0.001 | 0.005 | -0.179 | 8.58E-01 |
| rs55754224 | CAMK2D | rs35317913 | 0.95 | 0.98 | 4 | 114398814 | A | G | -0.011 | 0.002 | -6.368 | 1.91E-10 | 0.000 | 0.005 | -0.082 | 9.35E-01 |
| rs55754224 | CAMK2D | rs35370464 | 0.95 | 0.98 | 4 | 114401370 | T | C | 0.011 | 0.002 | 6.390 | 1.66E-10 | 0.001 | 0.005 | 0.104 | 9.17E-01 |
| rs55754224 | CAMK2D | rs13102179 | 0.95 | 0.98 | 4 | 114407519 | A | G | 0.011 | 0.002 | 6.289 | 3.20E-10 | 0.000 | 0.005 | 0.018 | 9.85E-01 |
| rs55754224 | CAMK2D | rs17530747 | 0.95 | 0.98 | 4 | 114408111 | T | C | 0.011 | 0.002 | 6.346 | 2.21E-10 | 0.002 | 0.005 | 0.293 | 7.70E-01 |
| rs55754224 | CAMK2D | rs35165228 | 0.95 | 0.98 | 4 | 114408680 | T | C | -0.011 | 0.002 | -6.302 | 2.93E-10 | -0.001 | 0.005 | -0.280 | 7.80E-01 |
| rs55754224 | CAMK2D | rs17446418 | 0.98 | 0.99 | 4 | 114418006 | T | G | -0.011 | 0.002 | -6.392 | 1.64E-10 | -0.002 | 0.005 | -0.420 | 6.74E-01 |
| rs55754224 | CAMK2D | rs17446453 | 0.97 | 0.99 | 4 | 114421712 | T | C | 0.012 | 0.002 | 6.484 | 8.92E-11 | 0.002 | 0.005 | 0.373 | 7.09E-01 |
| rs55754224 | CAMK2D | rs4834347 | 0.98 | 1 | 4 | 114425934 | A | C | 0.011 | 0.002 | 6.300 | 2.98E-10 | 0.002 | 0.005 | 0.372 | 7.10E-01 |
| rs55754224 | CAMK2D | rs55754224 | 1 | 1 | 4 | 114428714 | T | C | 0.012 | 0.002 | 6.573 | 4.92E-11 | 0.003 | 0.005 | 0.504 | 6.14E-01 |
| rs55754224 | CAMK2D | rs11098193 | 0.99 | 1 | 4 | 114429407 | T | C | 0.011 | 0.002 | 6.411 | 1.45E-10 | 0.001 | 0.005 | 0.263 | 7.92E-01 |
| rs55754224 | CAMK2D | rs11946291 | 0.92 | 0.97 | 4 | 114439476 | T | C | -0.011 | 0.002 | -6.196 | 5.80E-10 | -0.001 | 0.005 | -0.198 | 8.43E-01 |
| rs55754224 | CAMK2D | rs7700110 | 0.91 | 0.97 | 4 | 114439894 | A | G | 0.011 | 0.002 | 6.264 | 3.74E-10 | 0.001 | 0.005 | 0.155 | 8.77E-01 |
| rs55754224 | CAMK2D | rs56087422 | 0.89 | 0.97 | 4 | 114444394 | A | G | -0.011 | 0.002 | -6.353 | 2.11E-10 | -0.001 | 0.005 | -0.241 | 8.09E-01 |
| rs55754224 | CAMK2D | rs6533694 | 0.89 | 0.95 | 4 | 114444761 | T | G | -0.011 | 0.002 | -6.193 | 5.91E-10 | -0.001 | 0.005 | -0.221 | 8.25E-01 |
| rs55754224 | CAMK2D | rs6829664 | 0.9 | 0.97 | 4 | 114448656 | A | G | -0.012 | 0.002 | -6.478 | 9.27E-11 | -0.001 | 0.005 | -0.243 | 8.08E-01 |
| rs55754224 | CAMK2D | rs11938486 | 0.9 | 0.97 | 4 | 114449719 | A | C | -0.011 | 0.002 | -6.412 | 1.44E-10 | -0.001 | 0.005 | -0.284 | 7.76E-01 |
| rs55985730 | CALU, FLNC | rs55985730 | 1 | 1 | 7 | 128417044 | T | G | -0.018 | 0.003 | -5.538 | 3.06E-08 | -0.004 | 0.013 | -0.285 | 7.76E-01 |
| rs56181519 | WIPF1 | rs35215597 | 0.94 | 0.98 | 2 | 175543782 | A | G | 0.013 | 0.002 | 7.306 | 2.75E-13 | 0.011 | 0.005 | 2.071 | **3.84E-02** |
| rs56181519 | WIPF1 | rs56181519 | 1 | 1 | 2 | 175555714 | T | C | -0.013 | 0.002 | -7.350 | 1.98E-13 | -0.011 | 0.005 | -2.160 | **3.08E-02** |
| rs56181519 | WIPF1 | rs2358891 | 0.83 | 0.95 | 2 | 175558804 | A | G | -0.011 | 0.002 | -6.229 | 4.69E-10 | -0.011 | 0.005 | -2.003 | **4.51E-02** |
| rs635634 | ABO | rs507666 | 0.99 | 1 | 9 | 136149399 | A | G | 0.011 | 0.002 | 5.467 | 4.58E-08 | 0.010 | 0.006 | 1.690 | 9.10E-02 |
| rs635634 | ABO | rs532436 | 0.99 | 1 | 9 | 136149830 | A | G | 0.011 | 0.002 | 5.520 | 3.38E-08 | 0.010 | 0.006 | 1.707 | 8.79E-02 |

| | | | | | | | | | | | | | | | | |
|---|---|---|---|---|---|---|---|---|---|---|---|---|---|---|---|---|
| rs635634 | ABO | rs635634 | 1 | 1 | 9 | 136155000 | T | C | 0.011 | 0.002 | 5.725 | 1.04E-08 | 0.009 | 0.006 | 1.619 | 1.05E-01 |
| rs6598541 | IGF1R | rs6598541 | 1 | 1 | 15 | 99271135 | A | G | 0.009 | 0.002 | 5.811 | 6.21E-09 | 0.009 | 0.005 | 1.753 | 7.96E-02 |
| rs6598541 | IGF1R | rs7166287 | 0.99 | 1 | 15 | 99273075 | T | C | -0.009 | 0.002 | -5.593 | 2.23E-08 | -0.009 | 0.005 | -1.784 | 7.44E-02 |
| rs6598541 | IGF1R | rs4966019 | 0.96 | 0.99 | 15 | 99274326 | T | C | -0.009 | 0.002 | -5.635 | 1.75E-08 | -0.009 | 0.005 | -1.889 | 5.89E-02 |
| rs6598541 | IGF1R | rs2871974 | 0.95 | 0.98 | 15 | 99284074 | T | C | -0.009 | 0.002 | -5.522 | 3.35E-08 | -0.008 | 0.005 | -1.722 | 8.51E-02 |
| rs6598541 | IGF1R | rs4966021 | 0.95 | 0.99 | 15 | 99285056 | T | C | -0.009 | 0.002 | -5.460 | 4.77E-08 | -0.009 | 0.005 | -1.905 | 5.67E-02 |
| rs6598541 | IGF1R | rs12908437 | 0.93 | 0.98 | 15 | 99287375 | T | C | 0.009 | 0.002 | 5.503 | 3.74E-08 | 0.009 | 0.005 | 1.890 | 5.87E-02 |
| rs6598541 | IGF1R | rs12898337 | 0.87 | 0.94 | 15 | 99294355 | T | C | 0.009 | 0.002 | 5.469 | 4.53E-08 | 0.010 | 0.005 | 2.051 | **4.02E-02** |
| rs6747542 | GMCL1, ANXA4, | rs3771540 | 0.84 | 0.92 | 2 | 70031941 | A | G | 0.011 | 0.002 | 6.822 | 8.97E-12 | -0.002 | 0.005 | -0.407 | 6.84E-01 |
| rs6747542 | GMCL1, ANXA4, | rs2168115 | 0.87 | 0.94 | 2 | 70034793 | A | G | 0.011 | 0.002 | 6.791 | 1.11E-11 | -0.002 | 0.005 | -0.344 | 7.31E-01 |
| rs6747542 | GMCL1, ANXA4, | rs13392884 | 0.89 | 0.95 | 2 | 70036711 | A | G | 0.011 | 0.002 | 6.677 | 2.43E-11 | -0.001 | 0.005 | -0.116 | 9.08E-01 |
| rs6747542 | GMCL1, ANXA4, | rs3771537 | 0.89 | 0.95 | 2 | 70038792 | A | C | 0.011 | 0.002 | 6.865 | 6.63E-12 | -0.001 | 0.005 | -0.166 | 8.69E-01 |
| rs6747542 | GMCL1, ANXA4, | rs4853027 | 0.89 | 0.95 | 2 | 70039224 | T | C | -0.010 | 0.002 | -6.545 | 5.93E-11 | 0.000 | 0.005 | -0.024 | 9.81E-01 |
| rs6747542 | GMCL1, ANXA4, | rs2305523 | 0.91 | 0.96 | 2 | 70039677 | A | C | 0.010 | 0.002 | 6.445 | 1.16E-10 | -0.001 | 0.005 | -0.132 | 8.95E-01 |
| rs6747542 | GMCL1, ANXA4, | rs2312555 | 0.91 | 0.96 | 2 | 70040542 | A | G | -0.010 | 0.002 | -6.563 | 5.28E-11 | 0.000 | 0.005 | 0.086 | 9.31E-01 |
| rs6747542 | GMCL1, ANXA4, | rs11673826 | 0.91 | 0.96 | 2 | 70041033 | A | G | -0.010 | 0.002 | -6.519 | 7.05E-11 | 0.000 | 0.005 | 0.035 | 9.72E-01 |
| rs6747542 | GMCL1, ANXA4, | rs7563062 | 0.93 | 0.97 | 2 | 70050725 | A | G | -0.010 | 0.002 | -6.498 | 8.11E-11 | 0.000 | 0.005 | -0.053 | 9.58E-01 |
| rs6747542 | GMCL1, ANXA4, | rs1531025 | 0.92 | 0.96 | 2 | 70055505 | A | C | -0.010 | 0.002 | -6.538 | 6.23E-11 | -0.001 | 0.005 | -0.165 | 8.69E-01 |
| rs6747542 | GMCL1, ANXA4, | rs3732266 | 0.93 | 0.97 | 2 | 70057448 | T | G | 0.010 | 0.002 | 6.673 | 2.51E-11 | 0.001 | 0.005 | 0.191 | 8.48E-01 |
| rs6747542 | GMCL1, ANXA4, | rs10167142 | 0.95 | 0.97 | 2 | 70062360 | T | C | 0.010 | 0.002 | 6.668 | 2.60E-11 | 0.001 | 0.005 | 0.202 | 8.40E-01 |
| rs6747542 | GMCL1, ANXA4, | rs10202159 | 0.95 | 0.97 | 2 | 70062454 | T | G | -0.011 | 0.002 | -6.680 | 2.39E-11 | 0.000 | 0.005 | -0.095 | 9.24E-01 |
| rs6747542 | GMCL1, ANXA4, | rs4241261 | 0.95 | 0.97 | 2 | 70063660 | A | G | 0.011 | 0.002 | 6.709 | 1.95E-11 | 0.001 | 0.005 | 0.232 | 8.17E-01 |
| rs6747542 | GMCL1, ANXA4, | rs2872075 | 0.96 | 0.98 | 2 | 70068635 | T | G | -0.010 | 0.002 | -6.599 | 4.13E-11 | -0.001 | 0.005 | -0.282 | 7.78E-01 |
| rs6747542 | GMCL1, ANXA4, | rs6546551 | 0.96 | 0.98 | 2 | 70073703 | T | C | -0.011 | 0.002 | -6.681 | 2.37E-11 | -0.001 | 0.005 | -0.244 | 8.07E-01 |
| rs6747542 | GMCL1, ANXA4, | rs1551374 | 0.95 | 0.98 | 2 | 70079633 | T | C | -0.011 | 0.002 | -6.735 | 1.64E-11 | -0.001 | 0.005 | -0.185 | 8.53E-01 |
| rs6747542 | GMCL1, ANXA4, | rs10865380 | 0.98 | 0.99 | 2 | 70091246 | T | C | 0.011 | 0.002 | 6.953 | 3.59E-12 | 0.001 | 0.005 | 0.160 | 8.73E-01 |
| rs6747542 | GMCL1, ANXA4, | rs57278310 | 0.98 | 0.99 | 2 | 70093647 | T | C | 0.011 | 0.002 | 7.000 | 2.57E-12 | 0.001 | 0.005 | 0.167 | 8.67E-01 |
| rs6747542 | GMCL1, ANXA4, | rs60207187 | 0.98 | 0.99 | 2 | 70093910 | A | G | -0.011 | 0.002 | -7.050 | 1.79E-12 | -0.001 | 0.005 | -0.209 | 8.34E-01 |
| rs6747542 | GMCL1, ANXA4, | rs3214007 | 0.99 | 1 | 2 | 70098808 | A | G | 0.011 | 0.002 | 6.808 | 9.89E-12 | 0.001 | 0.005 | 0.140 | 8.89E-01 |
| rs6747542 | GMCL1, ANXA4, | rs6747542 | 1 | 1 | 2 | 70106832 | T | C | 0.011 | 0.002 | 7.063 | 1.63E-12 | 0.001 | 0.005 | 0.206 | 8.37E-01 |

| | | | | | | | | | | | | | | | | |
|---|---|---|---|---|---|---|---|---|---|---|---|---|---|---|---|---|
| rs6747542 | GMCL1, ANXA4, | rs6546553 | 0.93 | 1 | 2 | 70111711 | A | C | -0.010 | 0.002 | -6.568 | 5.10E-11 | 0.001 | 0.005 | 0.307 | 7.59E-01 |
| rs6747542 | GMCL1, ANXA4, | rs10165883 | 0.9 | 0.97 | 2 | 70117015 | T | C | -0.010 | 0.002 | -6.391 | 1.65E-10 | 0.001 | 0.005 | 0.203 | 8.39E-01 |
| rs6747542 | GMCL1, ANXA4, | rs12232914 | 0.92 | 0.99 | 2 | 70117176 | T | C | -0.010 | 0.002 | -6.420 | 1.36E-10 | 0.001 | 0.005 | 0.275 | 7.83E-01 |
| rs6747542 | GMCL1, ANXA4, | rs12233079 | 0.92 | 0.99 | 2 | 70117372 | A | G | 0.010 | 0.002 | 6.440 | 1.19E-10 | -0.001 | 0.005 | -0.234 | 8.15E-01 |
| rs6747542 | GMCL1, ANXA4, | rs2054047 | 0.92 | 0.99 | 2 | 70117421 | A | C | 0.010 | 0.002 | 6.434 | 1.25E-10 | -0.001 | 0.005 | -0.206 | 8.37E-01 |
| rs6747542 | GMCL1, ANXA4, | rs13002091 | 0.92 | 0.99 | 2 | 70118388 | T | C | -0.010 | 0.002 | -6.407 | 1.48E-10 | 0.001 | 0.005 | 0.193 | 8.47E-01 |
| rs6747542 | GMCL1, ANXA4, | rs3755389 | 0.92 | 0.99 | 2 | 70119469 | T | C | 0.010 | 0.002 | 6.422 | 1.34E-10 | -0.001 | 0.005 | -0.201 | 8.40E-01 |
| rs6747542 | GMCL1, ANXA4, | rs6720498 | 0.88 | 0.97 | 2 | 70124805 | T | C | 0.010 | 0.002 | 6.610 | 3.83E-11 | 0.000 | 0.005 | 0.075 | 9.40E-01 |
| rs6747542 | GMCL1, ANXA4, | rs13015457 | 0.91 | 0.99 | 2 | 70125763 | T | G | -0.010 | 0.002 | -6.514 | 7.33E-11 | 0.001 | 0.005 | 0.238 | 8.12E-01 |
| rs6747542 | GMCL1, ANXA4, | rs6716937 | 0.9 | 0.98 | 2 | 70127229 | A | G | -0.010 | 0.002 | -6.470 | 9.82E-11 | 0.001 | 0.005 | 0.252 | 8.01E-01 |
| rs6747542 | GMCL1, ANXA4, | rs7598283 | 0.89 | 0.98 | 2 | 70128717 | A | G | 0.010 | 0.002 | 6.474 | 9.57E-11 | -0.002 | 0.005 | -0.398 | 6.90E-01 |
| rs6747542 | GMCL1, ANXA4, | rs1048266 | 0.89 | 0.98 | 2 | 70132676 | A | G | 0.010 | 0.002 | 6.463 | 1.02E-10 | -0.002 | 0.005 | -0.430 | 6.67E-01 |
| rs6747542 | GMCL1, ANXA4, | rs10166011 | 0.89 | 0.98 | 2 | 70134107 | A | G | 0.010 | 0.002 | 6.504 | 7.84E-11 | -0.003 | 0.005 | -0.549 | 5.83E-01 |
| rs6747542 | GMCL1, ANXA4, | rs6546554 | 0.89 | 0.98 | 2 | 70135706 | A | G | -0.010 | 0.002 | -6.491 | 8.55E-11 | 0.003 | 0.005 | 0.549 | 5.83E-01 |
| rs6747542 | GMCL1, ANXA4, | rs6546555 | 0.89 | 0.98 | 2 | 70135760 | A | G | 0.010 | 0.002 | 6.514 | 7.33E-11 | -0.002 | 0.005 | -0.517 | 6.05E-01 |
| rs6747542 | GMCL1, ANXA4, | rs6736408 | 0.89 | 0.98 | 2 | 70137774 | T | C | -0.010 | 0.002 | -6.510 | 7.53E-11 | 0.003 | 0.005 | 0.573 | 5.67E-01 |
| rs6747542 | GMCL1, ANXA4, | rs6739780 | 0.89 | 0.98 | 2 | 70138111 | A | G | -0.010 | 0.002 | -6.520 | 7.04E-11 | 0.003 | 0.005 | 0.573 | 5.67E-01 |
| rs6747542 | GMCL1, ANXA4, | rs6754896 | 0.89 | 0.98 | 2 | 70138170 | T | C | 0.010 | 0.002 | 6.513 | 7.38E-11 | -0.003 | 0.005 | -0.573 | 5.67E-01 |
| rs6747542 | GMCL1, ANXA4, | rs6546556 | 0.89 | 0.98 | 2 | 70139001 | A | G | 0.010 | 0.002 | 6.523 | 6.90E-11 | -0.003 | 0.005 | -0.596 | 5.51E-01 |
| rs6747542 | GMCL1, ANXA4, | rs7592647 | 0.89 | 0.97 | 2 | 70139134 | T | C | -0.010 | 0.002 | -6.520 | 7.03E-11 | 0.003 | 0.005 | 0.553 | 5.80E-01 |
| rs6747542 | GMCL1, ANXA4, | rs897122 | 0.89 | 0.98 | 2 | 70140271 | T | C | -0.010 | 0.002 | -6.510 | 7.53E-11 | 0.003 | 0.005 | 0.573 | 5.67E-01 |
| rs6747542 | GMCL1, ANXA4, | rs11902198 | 0.87 | 0.96 | 2 | 70144596 | A | C | -0.010 | 0.002 | -6.362 | 1.99E-10 | 0.003 | 0.005 | 0.552 | 5.81E-01 |
| rs6747542 | GMCL1, ANXA4, | rs6546558 | 0.89 | 0.98 | 2 | 70145717 | T | C | -0.010 | 0.002 | -6.481 | 9.12E-11 | 0.002 | 0.005 | 0.454 | 6.50E-01 |
| rs6747542 | GMCL1, ANXA4, | rs6546559 | 0.89 | 0.98 | 2 | 70145833 | A | G | -0.010 | 0.002 | -6.418 | 1.38E-10 | 0.003 | 0.005 | 0.532 | 5.95E-01 |
| rs6747542 | GMCL1, ANXA4, | rs6721891 | 0.89 | 0.98 | 2 | 70146121 | A | G | 0.010 | 0.002 | 6.394 | 1.61E-10 | -0.003 | 0.005 | -0.532 | 5.95E-01 |
| rs6747542 | GMCL1, ANXA4, | rs6750488 | 0.89 | 0.98 | 2 | 70146125 | T | C | -0.010 | 0.002 | -6.329 | 2.47E-10 | 0.003 | 0.005 | 0.587 | 5.57E-01 |
| rs6747542 | GMCL1, ANXA4, | rs7419837 | 0.89 | 0.98 | 2 | 70146625 | T | C | 0.010 | 0.002 | 6.518 | 7.15E-11 | -0.003 | 0.005 | -0.586 | 5.58E-01 |
| rs6747542 | GMCL1, ANXA4, | rs1454498 | 0.88 | 0.97 | 2 | 70149225 | A | C | -0.010 | 0.002 | -6.353 | 2.11E-10 | 0.003 | 0.005 | 0.557 | 5.77E-01 |
| rs6747542 | GMCL1, ANXA4, | rs6738174 | 0.8 | 0.94 | 2 | 70150215 | A | C | 0.010 | 0.002 | 6.577 | 4.78E-11 | -0.003 | 0.005 | -0.625 | 5.32E-01 |
| rs6747542 | GMCL1, ANXA4, | rs6725425 | 0.88 | 0.97 | 2 | 70150674 | T | C | 0.010 | 0.002 | 6.527 | 6.72E-11 | -0.003 | 0.005 | -0.625 | 5.32E-01 |

| | | | | | | | | | | | | | | | | |
|---|---|---|---|---|---|---|---|---|---|---|---|---|---|---|---|---|
| rs6747542 | GMCL1, ANXA4, | rs10205487 | 0.88 | 0.97 | 2 | 70151471 | A | G | 0.010 | 0.002 | 6.486 | 8.83E-11 | -0.003 | 0.005 | -0.605 | 5.45E-01 |
| rs6747542 | GMCL1, ANXA4, | rs10496174 | 0.88 | 0.97 | 2 | 70153436 | T | G | 0.010 | 0.002 | 6.034 | 1.60E-09 | -0.003 | 0.005 | -0.605 | 5.45E-01 |
| rs6747542 | GMCL1, ANXA4, | rs7573442 | 0.88 | 0.97 | 2 | 70154037 | A | G | -0.010 | 0.002 | -6.474 | 9.54E-11 | 0.003 | 0.005 | 0.605 | 5.45E-01 |
| rs6747542 | GMCL1, ANXA4, | rs12713682 | 0.88 | 0.97 | 2 | 70154134 | A | G | -0.010 | 0.002 | -6.513 | 7.38E-11 | 0.003 | 0.005 | 0.605 | 5.45E-01 |
| rs6747542 | GMCL1, ANXA4, | rs11893500 | 0.88 | 0.97 | 2 | 70154187 | T | C | 0.010 | 0.002 | 6.543 | 6.03E-11 | -0.003 | 0.005 | -0.605 | 5.45E-01 |
| rs6747542 | GMCL1, ANXA4, | rs4144081 | 0.88 | 0.97 | 2 | 70155482 | A | G | -0.010 | 0.002 | -6.411 | 1.44E-10 | 0.003 | 0.005 | 0.625 | 5.32E-01 |
| rs6747542 | GMCL1, ANXA4, | rs6741449 | 0.88 | 0.97 | 2 | 70156589 | A | G | 0.010 | 0.002 | 6.343 | 2.25E-10 | -0.003 | 0.005 | -0.625 | 5.32E-01 |
| rs6747542 | GMCL1, ANXA4, | rs897119 | 0.85 | 0.96 | 2 | 70160658 | T | C | 0.010 | 0.002 | 6.455 | 1.08E-10 | -0.003 | 0.005 | -0.646 | 5.18E-01 |
| rs6747542 | GMCL1, ANXA4, | rs897120 | 0.86 | 0.96 | 2 | 70160982 | A | G | 0.010 | 0.002 | 6.525 | 6.78E-11 | -0.003 | 0.005 | -0.609 | 5.42E-01 |
| rs6747542 | GMCL1, ANXA4, | rs6729760 | 0.85 | 0.96 | 2 | 70161032 | T | C | -0.010 | 0.002 | -6.465 | 1.01E-10 | 0.002 | 0.005 | 0.505 | 6.13E-01 |
| rs6747542 | GMCL1, ANXA4, | rs12613947 | 0.86 | 0.96 | 2 | 70161342 | A | G | 0.010 | 0.002 | 6.489 | 8.63E-11 | -0.003 | 0.005 | -0.646 | 5.18E-01 |
| rs6747542 | GMCL1, ANXA4, | rs34586537 | 0.86 | 0.96 | 2 | 70163316 | T | G | 0.010 | 0.002 | 6.464 | 1.02E-10 | -0.002 | 0.005 | -0.500 | 6.17E-01 |
| rs6747542 | GMCL1, ANXA4, | rs11126251 | 0.86 | 0.96 | 2 | 70163434 | T | G | 0.010 | 0.002 | 6.502 | 7.92E-11 | -0.002 | 0.005 | -0.369 | 7.12E-01 |
| rs6747542 | GMCL1, ANXA4, | rs3771531 | 0.86 | 0.96 | 2 | 70164034 | T | C | 0.010 | 0.002 | 6.415 | 1.41E-10 | -0.003 | 0.005 | -0.530 | 5.96E-01 |
| rs6747542 | GMCL1, ANXA4, | rs12475412 | 0.85 | 0.96 | 2 | 70167197 | T | C | -0.010 | 0.002 | -6.606 | 3.95E-11 | 0.002 | 0.005 | 0.383 | 7.01E-01 |
| rs680084 | PRRX1 | rs12755237 | 0.86 | -0.96 | 1 | 170568714 | A | G | 0.017 | 0.002 | 10.973 | 5.13E-28 | 0.010 | 0.005 | 2.093 | **3.63E-02** |
| rs680084 | PRRX1 | rs6658866 | 0.84 | -0.94 | 1 | 170560604 | A | G | -0.017 | 0.002 | -10.779 | 4.33E-27 | -0.010 | 0.005 | -2.071 | **3.84E-02** |
| rs680084 | PRRX1 | rs12760630 | 0.86 | -0.96 | 1 | 170569689 | A | G | 0.017 | 0.002 | 10.992 | 4.16E-28 | 0.010 | 0.005 | 2.182 | **2.91E-02** |
| rs680084 | PRRX1 | rs7412231 | 0.86 | -0.96 | 1 | 170570499 | T | C | -0.018 | 0.002 | -11.086 | 1.47E-28 | -0.010 | 0.005 | -2.141 | **3.23E-02** |
| rs680084 | PRRX1 | rs12742164 | 0.82 | -0.94 | 1 | 170568896 | T | G | -0.017 | 0.002 | -10.917 | 9.60E-28 | -0.010 | 0.005 | -2.076 | **3.79E-02** |
| rs680084 | PRRX1 | rs2206062 | 0.86 | -0.96 | 1 | 170576107 | T | C | -0.017 | 0.002 | -11.013 | 3.32E-28 | -0.010 | 0.005 | -2.159 | **3.09E-02** |
| rs680084 | PRRX1 | rs10919444 | 0.86 | -0.96 | 1 | 170581634 | A | G | 0.017 | 0.002 | 11.014 | 3.28E-28 | 0.010 | 0.005 | 2.070 | **3.84E-02** |
| rs680084 | PRRX1 | rs1952675 | 0.86 | -0.96 | 1 | 170582575 | A | G | -0.018 | 0.002 | -11.132 | 8.75E-29 | -0.010 | 0.005 | -2.053 | **4.00E-02** |
| rs680084 | PRRX1 | rs61217505 | 0.86 | -0.96 | 1 | 170573258 | T | C | 0.018 | 0.002 | 11.111 | 1.11E-28 | 0.010 | 0.005 | 2.053 | **4.00E-02** |
| rs680084 | PRRX1 | rs6690642 | 0.86 | -0.96 | 1 | 170583944 | T | C | -0.017 | 0.002 | -11.000 | 3.84E-28 | -0.010 | 0.005 | -2.169 | **3.01E-02** |
| rs680084 | PRRX1 | rs763567 | 0.86 | -0.96 | 1 | 170566005 | A | G | -0.017 | 0.002 | -10.852 | 1.95E-27 | -0.010 | 0.005 | -2.025 | **4.28E-02** |
| rs680084 | PRRX1 | rs1928715 | 0.86 | -0.96 | 1 | 170567437 | T | C | 0.017 | 0.002 | 10.863 | 1.72E-27 | 0.010 | 0.005 | 2.076 | **3.79E-02** |
| rs680084 | PRRX1 | rs1234275 | 0.86 | 0.96 | 1 | 170596258 | A | G | 0.018 | 0.002 | 11.169 | 5.77E-29 | 0.009 | 0.005 | 1.961 | **4.99E-02** |
| rs680084 | PRRX1 | rs1234274 | 0.86 | 0.96 | 1 | 170596373 | A | G | 0.017 | 0.002 | 11.053 | 2.12E-28 | 0.009 | 0.005 | 1.961 | **4.99E-02** |
| rs680084 | PRRX1 | rs3903239 | 0.86 | -0.96 | 1 | 170569317 | A | G | -0.017 | 0.002 | -10.991 | 4.21E-28 | -0.010 | 0.005 | -2.182 | **2.91E-02** |

| | | | | | | | | | | | | | | | |
|---|---|---|---|---|---|---|---|---|---|---|---|---|---|---|---|
| rs680084 | PRRX1 | rs598993 | 0.92 | 0.97 | 1 | 170604122 | T | C | 0.018 | 0.002 | 11.228 | 2.96E-29 | 0.009 | 0.005 | 1.992 | **4.64E-02** |
| rs680084 | PRRX1 | rs541557 | 0.92 | 0.97 | 1 | 170604938 | A | G | 0.018 | 0.002 | 11.323 | 1.01E-29 | 0.009 | 0.005 | 1.992 | **4.64E-02** |
| rs680084 | PRRX1 | rs539045 | 0.92 | 0.97 | 1 | 170608081 | T | C | -0.018 | 0.002 | -11.294 | 1.41E-29 | -0.009 | 0.005 | -1.984 | **4.72E-02** |
| rs680084 | PRRX1 | rs473832 | 0.89 | 0.97 | 1 | 170609190 | A | C | 0.017 | 0.002 | 11.020 | 3.06E-28 | 0.010 | 0.005 | 2.090 | **3.66E-02** |
| rs680084 | PRRX1 | rs580487 | 0.92 | 0.97 | 1 | 170609625 | T | C | 0.018 | 0.002 | 11.178 | 5.20E-29 | 0.009 | 0.005 | 1.923 | 5.44E-02 |
| rs680084 | PRRX1 | rs492804 | 0.88 | 0.97 | 1 | 170612262 | T | C | -0.018 | 0.002 | -11.088 | 1.44E-28 | -0.010 | 0.005 | -2.118 | **3.42E-02** |
| rs680084 | PRRX1 | rs6677540 | 0.86 | -0.96 | 1 | 170583511 | A | G | 0.017 | 0.002 | 10.852 | 1.95E-27 | 0.010 | 0.005 | 2.159 | **3.09E-02** |
| rs680084 | PRRX1 | rs544478 | 0.89 | 0.97 | 1 | 170614042 | T | C | -0.018 | 0.002 | -11.148 | 7.31E-29 | -0.010 | 0.005 | -2.031 | **4.22E-02** |
| rs680084 | PRRX1 | rs593560 | 0.88 | 0.97 | 1 | 170615917 | A | G | -0.017 | 0.002 | -11.073 | 1.70E-28 | -0.010 | 0.005 | -2.011 | **4.44E-02** |
| rs680084 | PRRX1 | rs494521 | 0.88 | 0.94 | 1 | 170596072 | T | C | -0.017 | 0.002 | -10.963 | 5.76E-28 | -0.009 | 0.005 | -1.884 | 5.96E-02 |
| rs680084 | PRRX1 | rs623752 | 0.88 | 0.98 | 1 | 170618291 | A | G | 0.018 | 0.002 | 11.261 | 2.04E-29 | 0.009 | 0.005 | 1.966 | **4.93E-02** |
| rs680084 | PRRX1 | rs531007 | 0.88 | 0.98 | 1 | 170618382 | T | G | 0.018 | 0.002 | 11.414 | 3.54E-30 | 0.009 | 0.005 | 1.966 | **4.93E-02** |
| rs680084 | PRRX1 | rs638704 | 0.86 | 0.96 | 1 | 170598187 | T | C | -0.018 | 0.002 | -11.242 | 2.55E-29 | -0.010 | 0.005 | -2.049 | **4.04E-02** |
| rs680084 | PRRX1 | rs680084 | 1 | 1 | 1 | 170628255 | A | G | -0.018 | 0.002 | -11.460 | 2.10E-30 | -0.008 | 0.005 | -1.720 | 8.55E-02 |
| rs680084 | PRRX1 | rs629234 | 0.99 | 1 | 1 | 170631763 | T | C | 0.018 | 0.002 | 11.403 | 4.03E-30 | 0.008 | 0.005 | 1.640 | 1.01E-01 |
| rs680084 | PRRX1 | rs525489 | 0.9 | 0.99 | 1 | 170635002 | T | G | 0.018 | 0.002 | 11.318 | 1.07E-29 | 0.010 | 0.005 | 2.027 | **4.27E-02** |
| rs680084 | PRRX1 | rs503706 | 0.9 | 0.99 | 1 | 170635084 | T | C | 0.018 | 0.002 | 11.273 | 1.79E-29 | 0.010 | 0.005 | 2.056 | **3.98E-02** |
| rs6891790 | NKX2-5 | rs2277923 | 0.88 | 0.97 | 5 | 172662024 | T | C | 0.016 | 0.002 | 9.100 | 9.03E-20 | 0.010 | 0.005 | 2.026 | **4.28E-02** |
| rs6891790 | NKX2-5 | rs6882776 | 0.91 | 0.96 | 5 | 172664163 | A | G | -0.016 | 0.002 | -9.239 | 2.49E-20 | -0.008 | 0.005 | -1.576 | 1.15E-01 |
| rs6891790 | NKX2-5 | rs10071514 | 0.98 | 0.99 | 5 | 172669771 | A | G | 0.016 | 0.002 | 9.169 | 4.78E-20 | 0.009 | 0.005 | 1.667 | 9.55E-02 |
| rs6891790 | NKX2-5 | rs6891790 | 1 | 1 | 5 | 172670745 | T | G | -0.016 | 0.002 | -9.288 | 1.57E-20 | -0.009 | 0.005 | -1.707 | 8.78E-02 |
| rs7172038 | HCN4 | rs7164883 | 0.89 | 0.94 | 15 | 73652174 | A | G | -0.020 | 0.002 | -9.548 | 1.32E-21 | -0.003 | 0.007 | -0.386 | 6.99E-01 |
| rs7172038 | HCN4 | rs60512978 | 0.85 | 0.93 | 15 | 73653502 | T | C | -0.020 | 0.002 | -9.472 | 2.74E-21 | -0.002 | 0.006 | -0.346 | 7.30E-01 |
| rs7172038 | HCN4 | rs7172796 | 0.9 | 0.95 | 15 | 73661687 | T | G | -0.021 | 0.002 | -9.778 | 1.40E-22 | -0.004 | 0.006 | -0.649 | 5.16E-01 |
| rs7172038 | HCN4 | rs11630367 | 0.91 | 0.95 | 15 | 73662855 | A | G | -0.021 | 0.002 | -9.674 | 3.87E-22 | -0.004 | 0.006 | -0.665 | 5.06E-01 |
| rs7172038 | HCN4 | rs8038766 | 0.91 | 0.95 | 15 | 73663314 | A | G | -0.021 | 0.002 | -9.745 | 1.94E-22 | -0.004 | 0.006 | -0.638 | 5.23E-01 |
| rs7172038 | HCN4 | rs7172808 | 0.91 | 0.95 | 15 | 73663632 | A | G | -0.021 | 0.002 | -9.731 | 2.23E-22 | -0.004 | 0.006 | -0.638 | 5.23E-01 |
| rs7172038 | HCN4 | rs7174098 | 0.9 | 0.95 | 15 | 73664041 | A | G | 0.020 | 0.002 | 9.616 | 6.87E-22 | 0.004 | 0.007 | 0.638 | 5.23E-01 |
| rs7172038 | HCN4 | rs8035896 | 0.91 | 0.95 | 15 | 73664310 | T | G | -0.021 | 0.002 | -9.711 | 2.71E-22 | -0.004 | 0.006 | -0.638 | 5.23E-01 |
| rs7172038 | HCN4 | rs8040533 | 0.91 | 0.95 | 15 | 73664741 | T | G | -0.021 | 0.002 | -9.863 | 6.02E-23 | -0.004 | 0.006 | -0.657 | 5.11E-01 |

| | | | | | | | | | | | | | | | | |
|---|---|---|---|---|---|---|---|---|---|---|---|---|---|---|---|---|
| rs7172038 | HCN4 | rs4489968 | 0.91 | 0.95 | 15 | 73665506 | T | G | -0.021 | 0.002 | -9.816 | 9.60E-23 | -0.004 | 0.006 | -0.681 | 4.96E-01 |
| rs7172038 | HCN4 | rs7172038 | 1 | 1 | 15 | 73667255 | T | G | -0.021 | 0.002 | -10.040 | 1.02E-23 | -0.003 | 0.006 | -0.426 | 6.70E-01 |
| rs7172038 | HCN4 | rs74022964 | 0.86 | 0.93 | 15 | 73677264 | T | C | 0.021 | 0.002 | 9.795 | 1.18E-22 | 0.004 | 0.006 | 0.591 | 5.55E-01 |
| rs74399915 | KCNJ5 | rs75557443 | 0.98 | 1 | 11 | 128766366 | T | C | 0.022 | 0.003 | 7.873 | 3.45E-15 | -0.002 | 0.010 | -0.178 | 8.59E-01 |
| rs74399915 | KCNJ5 | rs78907918 | 0.87 | 1 | 11 | 128767957 | T | C | -0.021 | 0.003 | -7.028 | 2.10E-12 | 0.004 | 0.010 | 0.401 | 6.89E-01 |
| rs74399915 | KCNJ5 | rs113223494 | 1 | 1 | 11 | 128768852 | A | G | 0.022 | 0.003 | 7.970 | 1.59E-15 | -0.002 | 0.010 | -0.178 | 8.59E-01 |
| rs74399915 | KCNJ5 | rs74399915 | 1 | 1 | 11 | 128769010 | T | C | 0.022 | 0.003 | 8.026 | 1.01E-15 | -0.001 | 0.010 | -0.070 | 9.44E-01 |
| rs74399915 | KCNJ5 | rs76403033 | 1 | 1 | 11 | 128769387 | A | C | 0.022 | 0.003 | 8.004 | 1.21E-15 | -0.001 | 0.010 | -0.070 | 9.44E-01 |
| rs76774446 | GOSR2 | rs17608766 | 0.92 | 1 | 17 | 45013271 | T | C | -0.015 | 0.002 | -6.413 | 1.42E-10 | -0.009 | 0.007 | -1.226 | 2.20E-01 |
| rs76774446 | GOSR2 | rs11874 | 0.99 | 1 | 17 | 45017193 | A | G | 0.015 | 0.002 | 6.474 | 9.55E-11 | 0.005 | 0.007 | 0.732 | 4.64E-01 |
| rs76774446 | GOSR2 | rs76774446 | 1 | 1 | 17 | 45046368 | A | C | 0.015 | 0.002 | 6.632 | 3.32E-11 | 0.004 | 0.007 | 0.577 | 5.64E-01 |
| rs76774446 | GOSR2 | rs80335285 | 0.94 | 0.98 | 17 | 45055107 | A | G | -0.015 | 0.002 | -6.505 | 7.78E-11 | -0.004 | 0.007 | -0.488 | 6.25E-01 |
| rs78249997 | SYNPO2L | rs41280404 | 0.9 | 0.99 | 10 | 75405635 | T | G | 0.023 | 0.002 | 10.132 | 3.97E-24 | 0.011 | 0.007 | 1.512 | 1.31E-01 |
| rs78249997 | SYNPO2L | rs3740293 | 0.92 | 0.99 | 10 | 75406141 | A | C | 0.022 | 0.002 | 9.934 | 2.95E-23 | 0.012 | 0.007 | 1.681 | 9.28E-02 |
| rs78249997 | SYNPO2L | rs34163229 | 0.92 | 0.99 | 10 | 75406912 | T | G | -0.022 | 0.002 | -9.911 | 3.74E-23 | -0.011 | 0.007 | -1.538 | 1.24E-01 |
| rs78249997 | SYNPO2L | rs3812629 | 0.92 | 0.99 | 10 | 75407290 | A | G | -0.022 | 0.002 | -9.916 | 3.53E-23 | -0.012 | 0.007 | -1.642 | 1.01E-01 |
| rs78249997 | SYNPO2L | rs4746139 | 0.92 | 0.99 | 10 | 75407649 | A | C | 0.022 | 0.002 | 9.926 | 3.19E-23 | 0.012 | 0.007 | 1.550 | 1.21E-01 |
| rs78249997 | SYNPO2L | rs2177843 | 0.83 | 0.95 | 10 | 75409877 | T | C | -0.022 | 0.002 | -9.860 | 6.18E-23 | -0.012 | 0.007 | -1.602 | 1.09E-01 |
| rs78249997 | SYNPO2L | rs60632610 | 0.93 | 0.99 | 10 | 75415677 | T | C | -0.023 | 0.002 | -10.138 | 3.75E-24 | -0.013 | 0.007 | -1.768 | 7.71E-02 |
| rs78249997 | SYNPO2L | rs12570126 | 0.83 | 0.98 | 10 | 75416789 | A | G | 0.021 | 0.002 | 9.572 | 1.04E-21 | 0.012 | 0.007 | 1.672 | 9.45E-02 |
| rs78249997 | SYNPO2L | rs11000734 | 0.83 | 0.98 | 10 | 75419663 | T | G | 0.021 | 0.002 | 9.632 | 5.84E-22 | 0.012 | 0.007 | 1.676 | 9.37E-02 |
| rs78249997 | SYNPO2L | rs6480708 | 0.83 | 0.98 | 10 | 75420114 | A | C | -0.021 | 0.002 | -9.836 | 7.91E-23 | -0.011 | 0.007 | -1.611 | 1.07E-01 |
| rs78249997 | SYNPO2L | rs7915134 | 0.98 | 1 | 10 | 75420180 | T | C | -0.023 | 0.002 | -10.298 | 7.18E-25 | -0.013 | 0.007 | -1.749 | 8.03E-02 |
| rs78249997 | SYNPO2L | rs7900932 | 0.97 | 1 | 10 | 75420570 | T | C | 0.023 | 0.002 | 10.256 | 1.11E-24 | 0.013 | 0.007 | 1.749 | 8.02E-02 |
| rs78249997 | SYNPO2L | rs10824026 | 0.83 | -0.99 | 10 | 75421208 | A | G | 0.021 | 0.002 | 9.510 | 1.90E-21 | 0.011 | 0.007 | 1.623 | 1.05E-01 |
| rs78249997 | SYNPO2L | rs7394152 | 0.99 | 1 | 10 | 75421446 | T | C | -0.023 | 0.002 | -10.313 | 6.14E-25 | -0.013 | 0.007 | -1.753 | 7.97E-02 |
| rs78249997 | SYNPO2L | rs7394178 | 0.99 | 1 | 10 | 75421580 | A | G | -0.023 | 0.002 | -10.315 | 6.03E-25 | -0.013 | 0.007 | -1.724 | 8.47E-02 |
| rs78249997 | SYNPO2L | rs78249997 | 1 | 1 | 10 | 75422314 | T | C | -0.023 | 0.002 | -10.319 | 5.80E-25 | -0.013 | 0.007 | -1.734 | 8.29E-02 |
| rs78249997 | SYNPO2L | rs147790633 | 0.82 | 0.91 | 10 | 75447582 | T | C | 0.022 | 0.002 | 10.000 | 1.53E-23 | 0.014 | 0.008 | 1.905 | 5.67E-02 |
| rs79187193 | GJA5 | rs116546158 | 0.98 | 1 | 1 | 147250414 | A | G | -0.021 | 0.003 | -6.258 | 3.91E-10 | -0.009 | 0.014 | -0.629 | 5.29E-01 |

| | | | | | | | | | | | | | | | | |
|---|---|---|---|---|---|---|---|---|---|---|---|---|---|---|---|---|
| rs79187193 | GJA5 | rs74511541 | 0.98 | 1 | 1 | 147251948 | T | C | 0.021 | 0.003 | 6.266 | 3.71E-10 | 0.009 | 0.014 | 0.658 | 5.11E-01 |
| rs79187193 | GJA5 | rs116228788 | 0.98 | 1 | 1 | 147251361 | T | C | 0.021 | 0.003 | 6.274 | 3.53E-10 | 0.009 | 0.014 | 0.658 | 5.11E-01 |
| rs79187193 | GJA5 | rs79730440 | 1 | 1 | 1 | 147254644 | A | G | -0.021 | 0.003 | -6.296 | 3.05E-10 | -0.004 | 0.014 | -0.306 | 7.60E-01 |
| rs79187193 | GJA5 | rs17160660 | 1 | 1 | 1 | 147255323 | A | G | 0.021 | 0.003 | 6.302 | 2.94E-10 | 0.004 | 0.014 | 0.306 | 7.60E-01 |
| rs79187193 | GJA5 | rs142546567 | 0.98 | 1 | 1 | 147250526 | T | G | -0.021 | 0.003 | -6.264 | 3.75E-10 | -0.009 | 0.014 | -0.629 | 5.29E-01 |
| rs79187193 | GJA5 | rs76966341 | 1 | 1 | 1 | 147255905 | T | C | -0.021 | 0.003 | -6.387 | 1.69E-10 | -0.008 | 0.014 | -0.592 | 5.54E-01 |
| rs79187193 | GJA5 | rs76480392 | 1 | 1 | 1 | 147256108 | A | G | 0.021 | 0.003 | 6.343 | 2.25E-10 | 0.008 | 0.014 | 0.592 | 5.54E-01 |
| rs79187193 | GJA5 | rs74791680 | 1 | 1 | 1 | 147259321 | A | G | 0.021 | 0.003 | 6.355 | 2.09E-10 | 0.008 | 0.014 | 0.592 | 5.54E-01 |
| rs79187193 | GJA5 | rs76816335 | 1 | 1 | 1 | 147259336 | T | G | -0.021 | 0.003 | -6.364 | 1.97E-10 | -0.008 | 0.014 | -0.592 | 5.54E-01 |
| rs79187193 | GJA5 | rs75375598 | 1 | 1 | 1 | 147259640 | A | G | -0.021 | 0.003 | -6.351 | 2.14E-10 | -0.008 | 0.014 | -0.592 | 5.54E-01 |
| rs79187193 | GJA5 | rs1908628 | 1 | 1 | 1 | 147260568 | A | G | -0.021 | 0.003 | -6.378 | 1.80E-10 | -0.004 | 0.014 | -0.288 | 7.73E-01 |
| rs79187193 | GJA5 | rs76814955 | 0.98 | 1 | 1 | 147251688 | T | G | 0.021 | 0.003 | 6.265 | 3.73E-10 | 0.009 | 0.014 | 0.658 | 5.11E-01 |
| rs79187193 | GJA5 | rs74847853 | 1 | 1 | 1 | 147260812 | A | G | -0.021 | 0.003 | -6.347 | 2.20E-10 | -0.008 | 0.014 | -0.574 | 5.66E-01 |
| rs79187193 | GJA5 | rs75909378 | 1 | 1 | 1 | 147254227 | A | G | -0.021 | 0.003 | -6.334 | 2.39E-10 | -0.008 | 0.014 | -0.592 | 5.54E-01 |
| rs79187193 | GJA5 | rs79280699 | 1 | 1 | 1 | 147254365 | T | C | 0.021 | 0.003 | 6.331 | 2.44E-10 | 0.008 | 0.014 | 0.592 | 5.54E-01 |
| rs79187193 | GJA5 | rs78849618 | 1 | 1 | 1 | 147265956 | T | C | -0.021 | 0.003 | -6.242 | 4.31E-10 | -0.005 | 0.014 | -0.346 | 7.29E-01 |
| rs79187193 | GJA5 | rs75523969 | 1 | 1 | 1 | 147266890 | A | G | 0.021 | 0.003 | 6.224 | 4.83E-10 | 0.009 | 0.014 | 0.613 | 5.40E-01 |
| rs79187193 | GJA5 | rs79187193 | 1 | 1 | 1 | 147255831 | A | G | -0.021 | 0.003 | -6.470 | 9.79E-11 | -0.006 | 0.014 | -0.398 | 6.91E-01 |
| rs79187193 | GJA5 | rs77715544 | 1 | 1 | 1 | 147267616 | T | G | 0.021 | 0.003 | 6.260 | 3.86E-10 | 0.009 | 0.014 | 0.625 | 5.32E-01 |
| rs79187193 | GJA5 | rs140622021 | 1 | 1 | 1 | 147268529 | T | G | 0.021 | 0.003 | 6.263 | 3.77E-10 | 0.009 | 0.014 | 0.625 | 5.32E-01 |
| rs79187193 | GJA5 | rs79954696 | 1 | 1 | 1 | 147269771 | A | G | -0.021 | 0.003 | -6.252 | 4.04E-10 | -0.009 | 0.014 | -0.625 | 5.32E-01 |
| rs79187193 | GJA5 | rs76815884 | 1 | 1 | 1 | 147271396 | A | G | 0.021 | 0.003 | 6.285 | 3.28E-10 | 0.009 | 0.014 | 0.625 | 5.32E-01 |
| rs79187193 | GJA5 | rs80343037 | 0.98 | 1 | 1 | 147273120 | T | G | 0.021 | 0.003 | 6.246 | 4.22E-10 | 0.009 | 0.014 | 0.625 | 5.32E-01 |
| rs79187193 | GJA5 | rs17160683 | 1 | 1 | 1 | 147273306 | T | C | -0.021 | 0.003 | -6.177 | 6.51E-10 | -0.008 | 0.014 | -0.605 | 5.45E-01 |
| rs79187193 | GJA5 | rs1908627 | 1 | 1 | 1 | 147260765 | A | G | -0.021 | 0.003 | -6.347 | 2.19E-10 | -0.004 | 0.014 | -0.288 | 7.73E-01 |
| rs79187193 | GJA5 | rs76968567 | 1 | 1 | 1 | 147273885 | T | C | -0.021 | 0.003 | -6.342 | 2.28E-10 | -0.009 | 0.014 | -0.643 | 5.20E-01 |
| rs79187193 | GJA5 | rs75031333 | 1 | 1 | 1 | 147274016 | T | G | -0.021 | 0.003 | -6.169 | 6.89E-10 | -0.009 | 0.014 | -0.625 | 5.32E-01 |
| rs79187193 | GJA5 | rs76018518 | 1 | 1 | 1 | 147264779 | A | G | -0.021 | 0.003 | -6.373 | 1.85E-10 | -0.009 | 0.014 | -0.613 | 5.40E-01 |
| rs79187193 | GJA5 | rs74336706 | 0.84 | 1 | 1 | 147274359 | T | C | 0.017 | 0.003 | 5.510 | 3.59E-08 | 0.010 | 0.012 | 0.839 | 4.01E-01 |
| rs79187193 | GJA5 | rs79295811 | 0.84 | 1 | 1 | 147274365 | T | C | 0.017 | 0.003 | 5.504 | 3.72E-08 | 0.010 | 0.012 | 0.839 | 4.01E-01 |

| | | | | | | | | | | | | | | | | |
|---|---|---|---|---|---|---|---|---|---|---|---|---|---|---|---|---|
| rs79187193 | GJA5 | rs11240123 | 0.84 | 1 | 1 | 147267110 | T | C | 0.017 | 0.003 | 5.464 | 4.66E-08 | 0.006 | 0.012 | 0.469 | 6.39E-01 |
| rs79187193 | GJA5 | rs74919642 | 0.84 | 1 | 1 | 147276898 | A | G | -0.017 | 0.003 | -5.494 | 3.93E-08 | -0.010 | 0.012 | -0.823 | 4.10E-01 |
| rs79187193 | GJA5 | rs80109414 | 0.84 | 1 | 1 | 147276943 | T | G | -0.017 | 0.003 | -5.473 | 4.42E-08 | -0.010 | 0.012 | -0.823 | 4.10E-01 |
| rs79187193 | GJA5 | rs79208541 | 0.84 | 1 | 1 | 147276997 | A | G | -0.017 | 0.003 | -5.543 | 2.97E-08 | -0.011 | 0.012 | -0.845 | 3.98E-01 |
| rs79187193 | GJA5 | rs79103823 | 1 | 1 | 1 | 147277190 | T | C | -0.021 | 0.003 | -6.204 | 5.50E-10 | -0.006 | 0.014 | -0.412 | 6.80E-01 |
| rs79187193 | GJA5 | rs79259189 | 1 | 1 | 1 | 147277359 | T | C | -0.020 | 0.003 | -6.168 | 6.94E-10 | -0.006 | 0.014 | -0.412 | 6.80E-01 |
| rs79187193 | GJA5 | rs75715473 | 1 | 1 | 1 | 147277857 | T | C | -0.021 | 0.003 | -6.192 | 5.93E-10 | -0.006 | 0.014 | -0.412 | 6.80E-01 |
| rs79187193 | GJA5 | rs76634001 | 1 | 1 | 1 | 147278229 | A | G | 0.021 | 0.003 | 6.224 | 4.85E-10 | 0.006 | 0.014 | 0.457 | 6.48E-01 |
| rs79187193 | GJA5 | rs78291659 | 1 | 1 | 1 | 147278614 | A | C | 0.021 | 0.003 | 6.290 | 3.18E-10 | 0.009 | 0.014 | 0.607 | 5.44E-01 |
| rs79187193 | GJA5 | rs116430440 | 1 | 1 | 1 | 147279434 | A | G | -0.021 | 0.003 | -6.317 | 2.67E-10 | -0.008 | 0.014 | -0.585 | 5.58E-01 |
| rs79187193 | GJA5 | rs75391436 | 0.98 | 1 | 1 | 147279613 | T | C | 0.021 | 0.003 | 6.390 | 1.66E-10 | 0.008 | 0.014 | 0.585 | 5.58E-01 |
| rs79187193 | GJA5 | rs77459137 | 1 | 1 | 1 | 147280333 | A | G | -0.021 | 0.003 | -6.364 | 1.96E-10 | -0.007 | 0.014 | -0.503 | 6.15E-01 |
| rs79187193 | GJA5 | rs78034278 | 1 | 1 | 1 | 147280870 | A | G | 0.021 | 0.003 | 6.375 | 1.84E-10 | 0.003 | 0.014 | 0.195 | 8.45E-01 |
| rs79187193 | GJA5 | rs78757957 | 1 | 1 | 1 | 147280875 | A | C | 0.021 | 0.003 | 6.375 | 1.84E-10 | 0.003 | 0.014 | 0.195 | 8.45E-01 |
| rs79187193 | GJA5 | rs78586200 | 1 | 1 | 1 | 147281403 | T | C | 0.021 | 0.003 | 6.364 | 1.96E-10 | 0.004 | 0.014 | 0.272 | 7.86E-01 |
| rs79187193 | GJA5 | rs76574563 | 1 | 1 | 1 | 147281532 | A | G | 0.021 | 0.003 | 6.349 | 2.16E-10 | 0.004 | 0.014 | 0.272 | 7.86E-01 |
| rs79187193 | GJA5 | rs76421454 | 1 | 1 | 1 | 147283038 | A | G | 0.021 | 0.003 | 6.348 | 2.18E-10 | 0.003 | 0.014 | 0.250 | 8.03E-01 |
| rs79187193 | GJA5 | rs78059190 | 1 | 1 | 1 | 147286791 | A | G | 0.021 | 0.003 | 6.397 | 1.58E-10 | 0.002 | 0.014 | 0.130 | 8.96E-01 |
| rs79187193 | GJA5 | rs79624963 | 1 | 1 | 1 | 147288498 | T | C | -0.021 | 0.003 | -6.387 | 1.69E-10 | -0.003 | 0.014 | -0.237 | 8.12E-01 |
| rs79187193 | GJA5 | rs78658109 | 1 | 1 | 1 | 147290244 | T | C | -0.021 | 0.003 | -6.392 | 1.64E-10 | -0.003 | 0.014 | -0.237 | 8.12E-01 |
| rs79187193 | GJA5 | rs74535507 | 1 | 1 | 1 | 147291689 | T | C | -0.021 | 0.003 | -6.368 | 1.91E-10 | -0.002 | 0.014 | -0.170 | 8.65E-01 |
| rs79187193 | GJA5 | rs79204928 | 1 | 1 | 1 | 147291724 | T | C | -0.021 | 0.003 | -6.360 | 2.01E-10 | -0.002 | 0.014 | -0.170 | 8.65E-01 |
| rs79187193 | GJA5 | rs74968867 | 1 | 1 | 1 | 147292064 | T | C | -0.021 | 0.003 | -6.362 | 1.99E-10 | -0.004 | 0.014 | -0.277 | 7.82E-01 |
| rs79187193 | GJA5 | rs76035254 | 1 | 1 | 1 | 147293251 | T | C | -0.021 | 0.003 | -6.324 | 2.55E-10 | -0.005 | 0.013 | -0.361 | 7.18E-01 |
| rs79187193 | GJA5 | rs79157808 | 1 | 1 | 1 | 147293292 | T | C | 0.021 | 0.003 | 6.332 | 2.42E-10 | 0.005 | 0.013 | 0.361 | 7.18E-01 |
| rs79187193 | GJA5 | rs77131757 | 1 | 1 | 1 | 147293319 | A | G | -0.021 | 0.003 | -6.321 | 2.60E-10 | -0.005 | 0.013 | -0.361 | 7.18E-01 |
| rs79187193 | GJA5 | rs75663428 | 0.8 | 1 | 1 | 147294870 | T | C | -0.019 | 0.003 | -5.769 | 7.96E-09 | -0.005 | 0.013 | -0.361 | 7.18E-01 |
| rs79187193 | GJA5 | rs79487014 | 0.82 | 1 | 1 | 147294871 | T | C | -0.019 | 0.003 | -5.759 | 8.48E-09 | -0.005 | 0.013 | -0.361 | 7.18E-01 |
| rs79187193 | GJA5 | rs77532222 | 1 | 1 | 1 | 147295667 | T | C | -0.021 | 0.003 | -6.314 | 2.72E-10 | -0.005 | 0.013 | -0.361 | 7.18E-01 |
| rs79187193 | GJA5 | rs76522614 | 1 | 1 | 1 | 147295782 | T | C | 0.021 | 0.003 | 6.357 | 2.06E-10 | 0.005 | 0.013 | 0.361 | 7.18E-01 |

| | | | | | | | | | | | | | | | | |
|---|---|---|---|---|---|---|---|---|---|---|---|---|---|---|---|---|
| rs79187193 | GJA5 | rs1874394 | 0.84 | -1 | 1 | 147296308 | T | C | 0.017 | 0.003 | 5.509 | 3.60E-08 | 0.001 | 0.012 | 0.122 | 9.03E-01 |
| rs79187193 | GJA5 | rs79624916 | 1 | 1 | 1 | 147296366 | T | G | -0.021 | 0.003 | -6.317 | 2.67E-10 | -0.003 | 0.013 | -0.257 | 7.97E-01 |
| rs79187193 | GJA5 | rs77474276 | 1 | 1 | 1 | 147296409 | A | G | 0.021 | 0.003 | 6.317 | 2.67E-10 | 0.005 | 0.013 | 0.361 | 7.18E-01 |
| rs79187193 | GJA5 | rs77381999 | 1 | 1 | 1 | 147296453 | T | C | 0.021 | 0.003 | 6.331 | 2.43E-10 | 0.005 | 0.013 | 0.361 | 7.18E-01 |
| rs79187193 | GJA5 | rs74509621 | 1 | 1 | 1 | 147296489 | T | G | 0.021 | 0.003 | 6.326 | 2.51E-10 | 0.005 | 0.013 | 0.361 | 7.18E-01 |
| rs79187193 | GJA5 | rs77863448 | 1 | 1 | 1 | 147296599 | A | G | -0.021 | 0.003 | -6.318 | 2.65E-10 | -0.005 | 0.013 | -0.361 | 7.18E-01 |
| rs79187193 | GJA5 | rs75544532 | 1 | 1 | 1 | 147296603 | A | G | 0.021 | 0.003 | 6.318 | 2.65E-10 | 0.005 | 0.013 | 0.361 | 7.18E-01 |
| rs79187193 | GJA5 | rs75979898 | 1 | 1 | 1 | 147297243 | A | G | 0.021 | 0.003 | 6.320 | 2.62E-10 | 0.005 | 0.013 | 0.361 | 7.18E-01 |
| rs79187193 | GJA5 | rs115240411 | 1 | 1 | 1 | 147297850 | T | G | 0.021 | 0.003 | 6.328 | 2.48E-10 | 0.005 | 0.013 | 0.361 | 7.18E-01 |
| rs79187193 | GJA5 | rs80245413 | 1 | 1 | 1 | 147297958 | A | C | 0.021 | 0.003 | 6.370 | 1.88E-10 | 0.005 | 0.013 | 0.361 | 7.18E-01 |
| rs79187193 | GJA5 | rs115480103 | 1 | 1 | 1 | 147298566 | T | C | -0.021 | 0.003 | -6.266 | 3.71E-10 | -0.003 | 0.013 | -0.223 | 8.24E-01 |
| rs79187193 | GJA5 | rs116321716 | 1 | 1 | 1 | 147299286 | A | G | 0.021 | 0.003 | 6.316 | 2.69E-10 | 0.005 | 0.013 | 0.361 | 7.18E-01 |
| rs79187193 | GJA5 | rs144970042 | 1 | 1 | 1 | 147299407 | T | C | -0.021 | 0.003 | -6.323 | 2.57E-10 | -0.005 | 0.013 | -0.361 | 7.18E-01 |
| rs79187193 | GJA5 | rs149029102 | 1 | 1 | 1 | 147299447 | T | C | 0.021 | 0.003 | 6.321 | 2.59E-10 | 0.001 | 0.013 | 0.079 | 9.37E-01 |
| rs79187193 | GJA5 | rs115008394 | 1 | 1 | 1 | 147300184 | T | C | 0.021 | 0.003 | 6.308 | 2.83E-10 | 0.001 | 0.013 | 0.079 | 9.37E-01 |
| rs79187193 | GJA5 | rs114032655 | 1 | 1 | 1 | 147300260 | A | C | 0.021 | 0.003 | 6.303 | 2.91E-10 | 0.001 | 0.013 | 0.079 | 9.37E-01 |
| rs79187193 | GJA5 | rs146526746 | 1 | 1 | 1 | 147300470 | T | C | 0.021 | 0.003 | 6.317 | 2.67E-10 | 0.000 | 0.013 | -0.025 | 9.80E-01 |
| rs79187193 | GJA5 | rs149344021 | 1 | 1 | 1 | 147300771 | T | C | 0.021 | 0.003 | 6.315 | 2.69E-10 | 0.000 | 0.013 | -0.025 | 9.80E-01 |
| rs79187193 | GJA5 | rs115129368 | 1 | 1 | 1 | 147301055 | T | G | -0.021 | 0.003 | -6.317 | 2.67E-10 | -0.001 | 0.013 | -0.062 | 9.51E-01 |
| rs79187193 | GJA5 | rs148263084 | 1 | 1 | 1 | 147301165 | T | C | -0.021 | 0.003 | -6.440 | 1.20E-10 | 0.000 | 0.013 | -0.005 | 9.96E-01 |
| rs79187193 | GJA5 | rs146287012 | 0.83 | 1 | 1 | 147301177 | A | G | -0.020 | 0.003 | -6.133 | 8.63E-10 | -0.001 | 0.013 | -0.062 | 9.51E-01 |
| rs79187193 | GJA5 | rs116403883 | 1 | 1 | 1 | 147301911 | T | C | 0.021 | 0.003 | 6.347 | 2.20E-10 | -0.001 | 0.013 | -0.043 | 9.66E-01 |
| rs79187193 | GJA5 | rs140139006 | 1 | 1 | 1 | 147302177 | A | G | 0.021 | 0.003 | 6.331 | 2.43E-10 | 0.002 | 0.013 | 0.173 | 8.63E-01 |
| rs79187193 | GJA5 | rs116175751 | 1 | 1 | 1 | 147302435 | A | G | 0.021 | 0.003 | 6.321 | 2.59E-10 | 0.002 | 0.013 | 0.173 | 8.63E-01 |
| rs79187193 | GJA5 | rs115140387 | 1 | 1 | 1 | 147303174 | T | C | -0.021 | 0.003 | -6.301 | 2.95E-10 | -0.002 | 0.013 | -0.173 | 8.63E-01 |
| rs79187193 | GJA5 | rs115948478 | 1 | 1 | 1 | 147303573 | A | C | -0.021 | 0.003 | -6.316 | 2.69E-10 | -0.002 | 0.013 | -0.173 | 8.63E-01 |
| rs79187193 | GJA5 | rs140505020 | 1 | 1 | 1 | 147303727 | A | G | 0.021 | 0.003 | 6.321 | 2.60E-10 | 0.002 | 0.013 | 0.173 | 8.63E-01 |
| rs79187193 | GJA5 | rs115381114 | 1 | 1 | 1 | 147303837 | A | G | -0.021 | 0.003 | -6.282 | 3.35E-10 | -0.002 | 0.013 | -0.173 | 8.63E-01 |
| rs79187193 | GJA5 | rs138152129 | 1 | 1 | 1 | 147304668 | T | C | 0.021 | 0.003 | 6.319 | 2.64E-10 | 0.002 | 0.013 | 0.173 | 8.63E-01 |
| rs79187193 | GJA5 | rs116713931 | 1 | 1 | 1 | 147305144 | A | G | -0.021 | 0.003 | -6.317 | 2.66E-10 | -0.002 | 0.013 | -0.173 | 8.63E-01 |

| | | | | | | | | | | | | | | | | |
|---|---|---|---|---|---|---|---|---|---|---|---|---|---|---|---|---|
| rs79187193 | GJA5 | rs116258566 | 0.98 | 1 | 1 | 147305707 | T | C | -0.020 | 0.003 | -6.224 | 4.86E-10 | -0.002 | 0.013 | -0.173 | 8.63E-01 |
| rs79187193 | GJA5 | rs75255644 | 1 | 1 | 1 | 147305877 | T | C | 0.021 | 0.003 | 6.312 | 2.75E-10 | 0.002 | 0.013 | 0.173 | 8.63E-01 |
| rs79187193 | GJA5 | rs115583060 | 1 | 1 | 1 | 147305948 | A | C | -0.021 | 0.003 | -6.322 | 2.58E-10 | -0.002 | 0.013 | -0.173 | 8.63E-01 |
| rs79187193 | GJA5 | rs115063397 | 1 | 1 | 1 | 147307304 | A | G | -0.021 | 0.003 | -6.263 | 3.78E-10 | -0.002 | 0.014 | -0.131 | 8.96E-01 |
| rs79187193 | GJA5 | rs75702874 | 1 | 1 | 1 | 147308051 | A | G | -0.021 | 0.003 | -6.233 | 4.58E-10 | -0.002 | 0.014 | -0.131 | 8.96E-01 |
| rs79187193 | GJA5 | rs79561825 | 1 | 1 | 1 | 147309279 | T | C | 0.021 | 0.003 | 6.236 | 4.49E-10 | 0.002 | 0.014 | 0.131 | 8.96E-01 |
| rs79187193 | GJA5 | rs76492793 | 1 | 1 | 1 | 147309363 | T | G | -0.021 | 0.003 | -6.254 | 4.01E-10 | -0.002 | 0.014 | -0.131 | 8.96E-01 |
| rs79187193 | GJA5 | rs76570849 | 1 | 1 | 1 | 147310073 | T | G | -0.021 | 0.003 | -6.239 | 4.41E-10 | -0.002 | 0.014 | -0.131 | 8.96E-01 |
| rs79187193 | GJA5 | rs76467474 | 1 | 1 | 1 | 147310327 | T | C | 0.021 | 0.003 | 6.221 | 4.94E-10 | 0.001 | 0.014 | 0.103 | 9.18E-01 |
| rs79187193 | GJA5 | rs75931967 | 1 | 1 | 1 | 147310476 | T | G | -0.021 | 0.003 | -6.222 | 4.92E-10 | -0.001 | 0.014 | -0.103 | 9.18E-01 |
| rs79187193 | GJA5 | rs79572172 | 0.96 | 0.98 | 1 | 147311726 | T | C | -0.021 | 0.003 | -6.229 | 4.71E-10 | -0.001 | 0.014 | -0.103 | 9.18E-01 |
| rs79187193 | GJA5 | rs76706962 | 0.96 | 0.98 | 1 | 147312128 | T | C | 0.021 | 0.003 | 6.224 | 4.86E-10 | 0.001 | 0.014 | 0.103 | 9.18E-01 |
| rs79187193 | GJA5 | rs190745908 | 0.93 | 0.98 | 1 | 147312355 | T | C | -0.021 | 0.003 | -6.304 | 2.89E-10 | -0.001 | 0.014 | -0.068 | 9.46E-01 |
| rs79187193 | GJA5 | rs114158239 | 0.96 | 0.98 | 1 | 147312959 | T | C | 0.021 | 0.003 | 6.224 | 4.85E-10 | 0.001 | 0.014 | 0.066 | 9.47E-01 |
| rs79187193 | GJA5 | rs78581286 | 0.91 | 0.98 | 1 | 147316831 | A | G | -0.020 | 0.003 | -6.246 | 4.21E-10 | 0.000 | 0.013 | 0.012 | 9.91E-01 |
| rs79187193 | GJA5 | rs75185665 | 0.91 | 0.98 | 1 | 147317675 | T | C | -0.020 | 0.003 | -6.241 | 4.35E-10 | 0.000 | 0.013 | -0.013 | 9.90E-01 |
| rs79187193 | GJA5 | rs79618733 | 0.93 | 0.98 | 1 | 147323754 | A | G | 0.020 | 0.003 | 6.108 | 1.01E-09 | -0.003 | 0.013 | -0.199 | 8.42E-01 |
| rs79187193 | GJA5 | rs142757930 | 0.93 | 0.98 | 1 | 147324603 | T | C | -0.020 | 0.003 | -6.121 | 9.30E-10 | 0.003 | 0.013 | 0.195 | 8.45E-01 |
| rs79187193 | GJA5 | rs78945808 | 0.93 | 0.98 | 1 | 147324902 | A | G | -0.020 | 0.003 | -6.083 | 1.18E-09 | 0.003 | 0.013 | 0.255 | 7.99E-01 |
| rs79187193 | GJA5 | rs79957673 | 0.93 | 0.98 | 1 | 147326949 | T | C | -0.020 | 0.003 | -6.086 | 1.16E-09 | 0.002 | 0.013 | 0.123 | 9.02E-01 |
| rs79187193 | GJA5 | rs115932902 | 0.93 | 0.98 | 1 | 147327286 | T | C | 0.020 | 0.003 | 6.094 | 1.10E-09 | -0.002 | 0.013 | -0.123 | 9.02E-01 |
| rs79187193 | GJA5 | rs142111766 | 0.93 | 0.98 | 1 | 147328647 | T | C | -0.020 | 0.003 | -6.088 | 1.14E-09 | 0.000 | 0.013 | 0.023 | 9.82E-01 |
| rs79187193 | GJA5 | rs143350464 | 0.89 | 0.98 | 1 | 147334370 | T | C | -0.020 | 0.003 | -6.089 | 1.14E-09 | 0.001 | 0.013 | 0.091 | 9.27E-01 |
| rs79187193 | GJA5 | rs144918358 | 0.93 | 0.98 | 1 | 147339115 | A | C | -0.020 | 0.003 | -6.057 | 1.39E-09 | 0.001 | 0.013 | 0.091 | 9.27E-01 |
| rs79187193 | GJA5 | rs75945956 | 0.93 | 0.98 | 1 | 147344998 | T | G | -0.020 | 0.003 | -6.062 | 1.35E-09 | -0.002 | 0.013 | -0.185 | 8.53E-01 |
| rs79187193 | GJA5 | rs75456552 | 0.84 | 1 | 1 | 147274281 | T | C | -0.017 | 0.003 | -5.503 | 3.73E-08 | -0.010 | 0.012 | -0.839 | 4.01E-01 |
| rs79187193 | GJA5 | rs74568222 | 1 | 1 | 1 | 147273651 | T | C | -0.021 | 0.003 | -6.315 | 2.71E-10 | -0.009 | 0.014 | -0.625 | 5.32E-01 |
| rs79187193 | GJA5 | rs75626343 | 0.84 | 1 | 1 | 147263958 | T | C | -0.017 | 0.003 | -5.510 | 3.58E-08 | -0.006 | 0.012 | -0.498 | 6.18E-01 |
| rs79187193 | GJA5 | rs78515471 | 1 | 1 | 1 | 147274703 | T | C | 0.021 | 0.003 | 6.288 | 3.21E-10 | 0.009 | 0.014 | 0.625 | 5.32E-01 |
| rs883079 | TBX5 | rs2384407 | 0.88 | 0.95 | 12 | 114789226 | A | G | 0.019 | 0.002 | 10.653 | 1.69E-26 | 0.004 | 0.005 | 0.683 | 4.94E-01 |

| | | | | | | | | | | | | | | | | |
|---|---|---|---|---|---|---|---|---|---|---|---|---|---|---|---|---|
| rs883079 | TBX5 | rs7977083 | 0.81 | 1 | 12 | 114790500 | A | G | -0.019 | 0.002 | -10.417 | 2.07E-25 | -0.006 | 0.005 | -1.084 | 2.78E-01 |
| rs883079 | TBX5 | rs883079 | 1 | 1 | 12 | 114793240 | T | C | 0.019 | 0.002 | 10.960 | 5.97E-28 | 0.004 | 0.005 | 0.838 | 4.02E-01 |
| rs883079 | TBX5 | rs10507248 | 0.94 | 0.99 | 12 | 114797093 | T | G | 0.019 | 0.002 | 10.871 | 1.59E-27 | 0.005 | 0.005 | 1.028 | 3.04E-01 |
| rs883079 | TBX5 | rs7955405 | 0.94 | 0.99 | 12 | 114797306 | A | G | -0.019 | 0.002 | -10.762 | 5.21E-27 | -0.005 | 0.005 | -0.952 | 3.41E-01 |
| rs883079 | TBX5 | rs7312625 | 0.9 | 0.99 | 12 | 114799974 | A | G | 0.019 | 0.002 | 10.894 | 1.23E-27 | 0.007 | 0.005 | 1.267 | 2.05E-01 |
| rs883079 | TBX5 | rs4767237 | 0.85 | 0.99 | 12 | 114800813 | A | G | -0.019 | 0.002 | -10.653 | 1.69E-26 | -0.006 | 0.005 | -1.184 | 2.36E-01 |
| rs883079 | TBX5 | rs7135659 | 0.88 | 0.99 | 12 | 114801772 | A | G | 0.019 | 0.002 | 10.811 | 3.04E-27 | 0.006 | 0.005 | 1.171 | 2.41E-01 |
| rs883079 | TBX5 | rs1895585 | 0.83 | 0.99 | 12 | 114802138 | A | G | -0.019 | 0.002 | -10.683 | 1.22E-26 | -0.007 | 0.005 | -1.257 | 2.09E-01 |
| rs883079 | TBX5 | rs1946295 | 0.85 | 1 | 12 | 114802361 | A | G | -0.019 | 0.002 | -10.556 | 4.75E-26 | -0.006 | 0.005 | -1.205 | 2.28E-01 |
| rs883079 | TBX5 | rs1946293 | 0.89 | 0.99 | 12 | 114802760 | A | G | 0.019 | 0.002 | 10.739 | 6.71E-27 | 0.006 | 0.005 | 1.082 | 2.79E-01 |
| rs883079 | TBX5 | rs1895582 | 0.8 | 0.94 | 12 | 114807035 | A | G | 0.019 | 0.002 | 10.558 | 4.66E-26 | 0.007 | 0.005 | 1.285 | 1.99E-01 |

#### Online Table X. Gene Ontology of biological systems results for the gene sets of prioritized genes associated with a risk of cardioembolic stroke.

| geneSet | description | link | size | overlap | expect | enrichmentRatio | pValue | FDR | overlapId | userId |
|---|---|---|---|---|---|---|---|---|---|---|
| GO:0060048 | cardiac muscle contraction | http://amig | 126 | 9 | 0.36 | 25.33 | 6.47E-11 | 4.48E-07 | 817;857;1482;27 | KCNH2;GJA5;CAMK2D;KCNN2;KCNJ5;TTN;NKX2-5;HCN4;CAV1 |
| GO:0030048 | actin filament-based movement | http://amig | 132 | 9 | 0.37 | 24.17 | 9.85E-11 | 4.48E-07 | 817;857;2702;37 | KCNH2;GJA5;CAMK2D;KCNN2;WIPF1;KCNJ5;TTN;SYNE2;CAV1 |
| GO:0061337 | cardiac conduction | http://amig | 105 | 8 | 0.30 | 27.01 | 4.86E-10 | 1.25E-06 | 817;857;1482;27 | KCNH2;GJA5;CAMK2D;KCNN2;KCNJ5;NKX2-5;HCN4;CAV1 |
| GO:0006941 | striated muscle contraction | http://amig | 160 | 9 | 0.45 | 19.94 | 5.52E-10 | 1.25E-06 | 817;857;1482;27 | KCNH2;GJA5;CAMK2D;KCNN2;KCNJ5;TTN;NKX2-5;HCN4;CAV1 |
| GO:0086001 | cardiac muscle cell action potential | http://amig | 68 | 7 | 0.19 | 36.50 | 7.58E-10 | 1.38E-06 | 817;857;2702;37 | KCNH2;GJA5;CAMK2D;KCNN2;KCNJ5;HCN4;CAV1 |
| GO:0086091 | regulation of heart rate by cardiac conduction | http://amig | 38 | 6 | 0.11 | 55.98 | 9.32E-10 | 1.41E-06 | 817;857;2702;37 | KCNH2;GJA5;CAMK2D;KCNJ5;HCN4;CAV1 |
| GO:0086065 | cell communication involved in cardiac conduction | http://amig | 53 | 6 | 0.15 | 40.14 | 7.51E-09 | 9.76E-06 | 817;857;2702;37 | GJA5;CAMK2D;KCNN2;KCNJ5;HCN4;CAV1 |
| GO:0060047 | heart contraction | http://amig | 231 | 9 | 0.65 | 13.81 | 1.39E-08 | 1.58E-05 | 817;857;1482;27 | KCNH2;GJA5;CAMK2D;KCNN2;KCNJ5;TTN;NKX2-5;HCN4;CAV1 |
| GO:0003015 | heart process | http://amig | 240 | 9 | 0.68 | 13.30 | 1.94E-08 | 1.73E-05 | 817;857;1482;27 | KCNH2;GJA5;CAMK2D;KCNN2;KCNJ5;TTN;NKX2-5;HCN4;CAV1 |
| GO:0086003 | cardiac muscle cell contraction | http://amig | 63 | 6 | 0.18 | 33.77 | 2.18E-08 | 1.73E-05 | 817;857;2702;37 | KCNH2;GJA5;CAMK2D;KCNN2;KCNJ5;CAV1 |
| GO:0035637 | multicellular organismal signaling | http://amig | 170 | 8 | 0.48 | 16.68 | 2.23E-08 | 1.73E-05 | 817;857;1482;27 | KCNH2;GJA5;CAMK2D;KCNN2;KCNJ5;NKX2-5;HCN4;CAV1 |
| GO:0070252 | actin-mediated cell contraction | http://amig | 110 | 7 | 0.31 | 22.56 | 2.28E-08 | 1.73E-05 | 817;857;2702;37 | KCNH2;GJA5;CAMK2D;KCNN2;KCNJ5;TTN;CAV1 |
| GO:0001508 | action potential | http://amig | 127 | 7 | 0.36 | 19.54 | 6.17E-08 | 4.32E-05 | 817;857;2702;37 | KCNH2;GJA5;CAMK2D;KCNN2;KCNJ5;HCN4;CAV1 |
| GO:0008016 | regulation of heart contraction | http://amig | 205 | 8 | 0.58 | 13.84 | 9.53E-08 | 6.19E-05 | 817;857;1482;27 | KCNH2;GJA5;CAMK2D;KCNN2;KCNJ5;NKX2-5;HCN4;CAV1 |
| GO:0086009 | membrane repolarization | http://amig | 44 | 5 | 0.12 | 40.29 | 1.43E-07 | 8.69E-05 | 857;2702;3757;3 | KCNH2;GJA5;KCNN2;KCNJ5;CAV1 |
| GO:0003012 | muscle system process | http://amig | 423 | 10 | 1.19 | 8.38 | 2.23E-07 | 1.23E-04 | 817;857;1482;27 | PTK2;KCNH2;GJA5;CAMK2D;KCNN2;KCNJ5;TTN;NKX2-5;HCN4;CAV1 |
| GO:0002027 | regulation of heart rate | http://amig | 93 | 6 | 0.26 | 22.87 | 2.29E-07 | 1.23E-04 | 817;857;2702;37 | KCNH2;GJA5;CAMK2D;KCNJ5;HCN4;CAV1 |
| GO:0086002 | cardiac muscle cell action potential involved in cont | http://amig | 51 | 5 | 0.14 | 34.76 | 3.06E-07 | 1.54E-04 | 857;2702;3757;3 | KCNH2;GJA5;KCNN2;KCNJ5;CAV1 |
| GO:0006936 | muscle contraction | http://amig | 339 | 9 | 0.96 | 9.41 | 3.70E-07 | 1.77E-04 | 817;857;1482;27 | KCNH2;GJA5;CAMK2D;KCNN2;KCNJ5;TTN;NKX2-5;HCN4;CAV1 |
| GO:1903522 | regulation of blood circulation | http://amig | 248 | 8 | 0.70 | 11.44 | 4.10E-07 | 1.86E-04 | 817;857;1482;27 | KCNH2;GJA5;CAMK2D;KCNN2;KCNJ5;NKX2-5;HCN4;CAV1 |
| GO:0008015 | blood circulation | http://amig | 471 | 10 | 1.33 | 7.53 | 5.99E-07 | 2.59E-04 | 817;857;1482;27 | KCNH2;GJA5;CAMK2D;KCNN2;KCNJ5;TTN;NKX2-5;HCN4;AOPEP;CAV1 |
| GO:0003013 | circulatory system process | http://amig | 480 | 10 | 1.35 | 7.39 | 7.12E-07 | 2.94E-04 | 817;857;1482;27 | KCNH2;GJA5;CAMK2D;KCNN2;KCNJ5;TTN;NKX2-5;HCN4;AOPEP;CAV1 |
| GO:0086011 | membrane repolarization during action potential | http://amig | 26 | 4 | 0.07 | 54.55 | 7.93E-07 | 3.14E-04 | 857;3757;3762;3 | KCNH2;KCNN2;KCNJ5;CAV1 |
| GO:0098901 | regulation of cardiac muscle cell action potential | http://amig | 28 | 4 | 0.08 | 50.65 | 1.08E-06 | 4.10E-04 | 817;857;2702;10 | GJA5;CAMK2D;HCN4;CAV1 |
| GO:0098909 | regulation of cardiac muscle cell action potential inv | http://amig | 8 | 3 | 0.02 | 132.96 | 1.17E-06 | 4.24E-04 | 817;857;10021 | CAMK2D;HCN4;CAV1 |
| GO:0015672 | monovalent inorganic cation transport | http://amig | 513 | 10 | 1.45 | 6.91 | 1.30E-06 | 4.55E-04 | 817;857;1482;27 | KCNH2;GJA5;CAMK2D;KCNN2;KCNJ5;NKX2-5;HCN4;KLHL3;CAV1;KCNN3 |
| GO:0086019 | cell-cell signaling involved in cardiac conduction | http://amig | 30 | 4 | 0.08 | 47.27 | 1.44E-06 | 4.86E-04 | 2702;3762;3781; | GJA5;KCNN2;KCNJ5;HCN4 |
| GO:0055117 | regulation of cardiac muscle contraction | http://amig | 71 | 5 | 0.20 | 24.97 | 1.62E-06 | 5.27E-04 | 817;857;1482;27 | GJA5;CAMK2D;NKX2-5;HCN4;CAV1 |
| GO:0086005 | ventricular cardiac muscle cell action potential | http://amig | 34 | 4 | 0.10 | 41.71 | 2.42E-06 | 7.59E-04 | 857;2702;3757;3 | KCNH2;GJA5;KCNJ5;CAV1 |
| GO:0099622 | cardiac muscle cell membrane repolarization | http://amig | 36 | 4 | 0.10 | 39.39 | 3.06E-06 | 9.28E-04 | 2702;3757;3762; | KCNH2;GJA5;KCNN2;KCNJ5 |
| GO:0006813 | potassium ion transport | http://amig | 229 | 7 | 0.65 | 10.84 | 3.33E-06 | 9.73E-04 | 857;2702;3757;3 | KCNH2;GJA5;KCNN2;KCNJ5;HCN4;CAV1;KCNN3 |
| GO:0086004 | regulation of cardiac muscle cell contraction | http://amig | 37 | 4 | 0.10 | 38.33 | 3.43E-06 | 9.73E-04 | 817;857;2702;10 | GJA5;CAMK2D;HCN4;CAV1 |
| GO:0030029 | actin filament-based process | http://amig | 715 | 11 | 2.02 | 5.45 | 3.56E-06 | 9.81E-04 | 817;857;1482;27 | KCNH2;GJA5;CAMK2D;KCNN2;WIPF1;KCNJ5;TTN;NKX2-5;SYNE2;SYNPO2L;CAV1 |
| GO:0051899 | membrane depolarization | http://amig | 85 | 5 | 0.24 | 20.86 | 3.97E-06 | 1.03E-03 | 817;857;2702;37 | KCNH2;GJA5;CAMK2D;HCN4;CAV1 |
| GO:0006942 | regulation of striated muscle contraction | http://amig | 85 | 5 | 0.24 | 20.86 | 3.97E-06 | 1.03E-03 | 817;857;1482;27 | GJA5;CAMK2D;NKX2-5;HCN4;CAV1 |
| GO:0003161 | cardiac conduction system development | http://amig | 12 | 3 | 0.03 | 88.64 | 4.54E-06 | 1.15E-03 | 1482;2702;6910 | GJA5;NKX2-5;TBX5 |
| GO:1903115 | regulation of actin filament-based movement | http://amig | 42 | 4 | 0.12 | 33.77 | 5.75E-06 | 1.41E-03 | 817;857;2702;10 | GJA5;CAMK2D;HCN4;CAV1 |
| GO:0003007 | heart morphogenesis | http://amig | 250 | 7 | 0.71 | 9.93 | 5.93E-06 | 1.42E-03 | 1482;2702;5308; | PTK2;GJA5;TTN;NKX2-5;SYNPO2L;TBX5;PITX2 |
| GO:0098900 | regulation of action potential | http://amig | 49 | 4 | 0.14 | 28.94 | 1.07E-05 | 2.50E-03 | 817;857;2702;10 | GJA5;CAMK2D;HCN4;CAV1 |
| GO:0060413 | atrial septum morphogenesis | http://amig | 16 | 3 | 0.05 | 66.48 | 1.15E-05 | 2.61E-03 | 1482;2702;6910 | GJA5;NKX2-5;TBX5 |
| GO:1903779 | regulation of cardiac conduction | http://amig | 51 | 4 | 0.14 | 27.81 | 1.26E-05 | 2.79E-03 | 817;857;1482;27 | GJA5;CAMK2D;NKX2-5;CAV1 |
| GO:0044057 | regulation of system process | http://amig | 535 | 9 | 1.51 | 5.96 | 1.55E-05 | 3.34E-03 | 817;857;1482;27 | PTK2;KCNH2;GJA5;CAMK2D;KCNN2;KCNJ5;NKX2-5;HCN4;CAV1 |
| GO:0007507 | heart development | http://amig | 536 | 9 | 1.51 | 5.95 | 1.58E-05 | 3.34E-03 | 1026;1482;2702; | PTK2;RBM20;GJA5;CDKN1A;TTN;NKX2-5;SYNPO2L;TBX5;PITX2 |
| GO:0072359 | circulatory system development | http://amig | 1026 | 12 | 2.89 | 4.15 | 1.95E-05 | 3.82E-03 | 857;861;1026;14 | PTK2;RBM20;GJA5;CDKN1A;RUNX1;TTN;NKX2-5;SYNPO2L;TBX5;PRRX1;CAV1;PITX2 |
| GO:0086014 | atrial cardiac muscle cell action potential | http://amig | 19 | 3 | 0.05 | 55.98 | 1.97E-05 | 3.82E-03 | 2702;3762;3781 | GJA5;KCNN2;KCNJ5 |
| GO:0086026 | atrial cardiac muscle cell to AV node cell signaling | http://amig | 19 | 3 | 0.05 | 55.98 | 1.97E-05 | 3.82E-03 | 2702;3762;3781 | GJA5;KCNN2;KCNJ5 |
| GO:0086066 | atrial cardiac muscle cell to AV node cell communic | http://amig | 19 | 3 | 0.05 | 55.98 | 1.97E-05 | 3.82E-03 | 2702;3762;3781 | GJA5;KCNN2;KCNJ5 |

| | | | | | | | | | | |
|---|---|---|---|---|---|---|---|---|---|---|
| GO:0071804 | cellular potassium ion transport | http://amig | 202 | 6 | 0.57 | 10.53 | 2.09E-05 | 3.88E-03 | 857;3757;3762;3 | KCNH2;KCNN2;KCNJ5;HCN4;CAV1;KCNN3 |
| GO:0071805 | potassium ion transmembrane transport | http://amig | 202 | 6 | 0.57 | 10.53 | 2.09E-05 | 3.88E-03 | 857;3757;3762;3 | KCNH2;KCNN2;KCNJ5;HCN4;CAV1;KCNN3 |
| GO:0003283 | atrial septum development | http://amig | 20 | 3 | 0.06 | 53.18 | 2.32E-05 | 4.21E-03 | 1482;2702;6910 | GJA5;NKX2-5;TBX5 |
| GO:0090257 | regulation of muscle system process | http://amig | 222 | 6 | 0.63 | 9.58 | 3.56E-05 | 6.14E-03 | 817;857;1482;27 | PTK2;GJA5;CAMK2D;NKX2-5;HCN4;CAV1 |
| GO:0086013 | membrane repolarization during cardiac muscle cell | http://amig | 23 | 3 | 0.06 | 46.25 | 3.58E-05 | 6.14E-03 | 3757;3762;3781 | KCNH2;KCNN2;KCNJ5 |
| GO:0086064 | cell communication by electrical coupling involved ir | http://amig | 23 | 3 | 0.06 | 46.25 | 3.58E-05 | 6.14E-03 | 817;857;2702 | GJA5;CAMK2D;CAV1 |
| GO:0099625 | ventricular cardiac muscle cell membrane repolariz | http://amig | 26 | 3 | 0.07 | 40.91 | 5.22E-05 | 8.79E-03 | 2702;3757;3762 | KCNH2;GJA5;KCNJ5 |
| GO:0006928 | movement of cell or subcellular component | http://amig | 1967 | 16 | 5.55 | 2.88 | 5.54E-05 | 9.15E-03 | 817;857;2676;27 | PTK2;IGF1R;KCNH2;GJA5;CAMK2D;KCNN2;WIPF1;KCNJ5;TTN;SYNE2;GFRA3;KIF20A;TBX5;CAV1;NEURL1;PITX2 |
| GO:0001570 | vasculogenesis | http://amig | 75 | 4 | 0.21 | 18.91 | 5.83E-05 | 9.46E-03 | 857;1482;5747;6 | PTK2;NKX2-5;TBX5;CAV1 |
| GO:0006937 | regulation of muscle contraction | http://amig | 151 | 5 | 0.43 | 11.74 | 6.45E-05 | 1.03E-02 | 817;857;1482;27 | GJA5;CAMK2D;NKX2-5;HCN4;CAV1 |
| GO:0010644 | cell communication by electrical coupling | http://amig | 28 | 3 | 0.08 | 37.99 | 6.56E-05 | 1.03E-02 | 817;857;2702 | GJA5;CAMK2D;CAV1 |
| GO:0003164 | His-Purkinje system development | http://amig | 5 | 2 | 0.01 | 141.82 | 7.74E-05 | 1.19E-02 | 1482;6910 | NKX2-5;TBX5 |
| GO:0003209 | cardiac atrium morphogenesis | http://amig | 30 | 3 | 0.08 | 35.46 | 8.09E-05 | 1.23E-02 | 1482;2702;6910 | GJA5;NKX2-5;TBX5 |
| GO:0030001 | metal ion transport | http://amig | 841 | 10 | 2.37 | 4.22 | 9.58E-05 | 1.43E-02 | 817;857;1482;27 | KCNH2;GJA5;CAMK2D;KCNN2;KCNJ5;NKX2-5;HCN4;KLHL3;CAV1;KCNN3 |
| GO:0060306 | regulation of membrane repolarization | http://amig | 33 | 3 | 0.09 | 32.23 | 1.08E-04 | 1.59E-02 | 857;2702;3757 | KCNH2;GJA5;CAV1 |
| GO:0051891 | positive regulation of cardioblast differentiation | http://amig | 6 | 2 | 0.02 | 118.18 | 1.16E-04 | 1.65E-02 | 1482;6910 | NKX2-5;TBX5 |
| GO:0098914 | membrane repolarization during atrial cardiac musc | http://amig | 6 | 2 | 0.02 | 118.18 | 1.16E-04 | 1.65E-02 | 3762;3781 | KCNN2;KCNJ5 |
| GO:0003230 | cardiac atrium development | http://amig | 34 | 3 | 0.10 | 31.28 | 1.18E-04 | 1.66E-02 | 1482;2702;6910 | GJA5;NKX2-5;TBX5 |
| GO:0086010 | membrane depolarization during action potential | http://amig | 36 | 3 | 0.10 | 29.55 | 1.41E-04 | 1.94E-02 | 2702;3757;1002 | KCNH2;GJA5;HCN4 |
| GO:0042391 | regulation of membrane potential | http://amig | 414 | 7 | 1.17 | 5.99 | 1.48E-04 | 2.01E-02 | 817;857;2702;37 | KCNH2;GJA5;CAMK2D;KCNN2;KCNJ5;HCN4;CAV1 |
| GO:0038083 | peptidyl-tyrosine autophosphorylation | http://amig | 37 | 3 | 0.10 | 28.75 | 1.53E-04 | 2.04E-02 | 857;3480;5747 | PTK2;IGF1R;CAV1 |
| GO:1901844 | regulation of cell communication by electrical coupli | http://amig | 7 | 2 | 0.02 | 101.30 | 1.62E-04 | 2.13E-02 | 817;857 | CAMK2D;CAV1 |
| GO:1990573 | potassium ion import across plasma membrane | http://amig | 41 | 3 | 0.12 | 25.94 | 2.08E-04 | 2.70E-02 | 3757;3762;1002 | KCNH2;KCNJ5;HCN4 |
| GO:0051890 | regulation of cardioblast differentiation | http://amig | 8 | 2 | 0.02 | 88.64 | 2.16E-04 | 2.76E-02 | 1482;6910 | NKX2-5;TBX5 |
| GO:0034765 | regulation of ion transmembrane transport | http://amig | 441 | 7 | 1.24 | 5.63 | 2.18E-04 | 2.76E-02 | 817;857;2702;37 | KCNH2;GJA5;CAMK2D;KCNN2;KCNJ5;HCN4;CAV1 |
| GO:0003254 | regulation of membrane depolarization | http://amig | 43 | 3 | 0.12 | 24.74 | 2.40E-04 | 2.99E-02 | 817;2702;10021 | GJA5;CAMK2D;HCN4 |
| GO:0061061 | muscle structure development | http://amig | 610 | 8 | 1.72 | 4.65 | 2.74E-04 | 3.31E-02 | 463;857;1482;23 | FLNC;TTN;NKX2-5;SYNPO2L;TBX5;CAV1;NEURL1;ZFHX3 |
| GO:0086015 | SA node cell action potential | http://amig | 9 | 2 | 0.03 | 78.79 | 2.77E-04 | 3.31E-02 | 2702;10021 | GJA5;HCN4 |
| GO:0086018 | SA node cell to atrial cardiac muscle cell signaling | http://amig | 9 | 2 | 0.03 | 78.79 | 2.77E-04 | 3.31E-02 | 2702;10021 | GJA5;HCN4 |
| GO:0010107 | potassium ion import | http://amig | 46 | 3 | 0.13 | 23.12 | 2.93E-04 | 3.46E-02 | 3757;3762;1002 | KCNH2;KCNJ5;HCN4 |
| GO:0045214 | sarcomere organization | http://amig | 47 | 3 | 0.13 | 22.63 | 3.13E-04 | 3.64E-02 | 1482;7273;7993 | TTN;NKX2-5;SYNPO2L |
| GO:0009887 | animal organ morphogenesis | http://amig | 974 | 10 | 2.75 | 3.64 | 3.17E-04 | 3.65E-02 | 857;1482;2702;5 | PTK2;GJA5;TTN;NKX2-5;SYNPO2L;KLHL3;TBX5;PRRX1;CAV1;PITX2 |
| GO:0086070 | SA node cell to atrial cardiac muscle cell communic | http://amig | 10 | 2 | 0.03 | 70.91 | 3.45E-04 | 3.78E-02 | 2702;10021 | GJA5;HCN4 |
| GO:0010649 | regulation of cell communication by electrical coupli | http://amig | 10 | 2 | 0.03 | 70.91 | 3.45E-04 | 3.78E-02 | 817;857 | CAMK2D;CAV1 |
| GO:0099624 | atrial cardiac muscle cell membrane repolarization | http://amig | 10 | 2 | 0.03 | 70.91 | 3.45E-04 | 3.78E-02 | 3762;3781 | KCNN2;KCNJ5 |
| GO:0003211 | cardiac ventricle formation | http://amig | 10 | 2 | 0.03 | 70.91 | 3.45E-04 | 3.78E-02 | 1482;6910 | NKX2-5;TBX5 |
| GO:0043269 | regulation of ion transport | http://amig | 645 | 8 | 1.82 | 4.40 | 3.99E-04 | 4.31E-02 | 817;857;1482;27 | KCNH2;GJA5;CAMK2D;KCNN2;KCNJ5;NKX2-5;HCN4;CAV1 |
| GO:0016264 | gap junction assembly | http://amig | 11 | 2 | 0.03 | 64.46 | 4.21E-04 | 4.51E-02 | 857;2702 | GJA5;CAV1 |

**Online Table XI. DrugBank drugs targeting prioritized genes associated with CES.**

| Drug Name | Type | Target | Target Gene Symbol | Groups |
|---|---|---|---|---|
| 7-PYRIDIN-2-YL-N-(3,4,5-TRIMETHOXYPHI | Small molecule | Focal adhesion kinase 1 | *PTK2* | Experimental |
| Endostatin | Biotech | Focal adhesion kinase 1 | *PTK2* | Investigational |
| 2-({5-CHLORO-2-[(2-METHOXY-4-MORPHC | Small molecule | Focal adhesion kinase 1 | *PTK2* | Experimental |
| Fostamatinib | Small molecule | Focal adhesion kinase 1 | *PTK2* | Approved, Investigational |
| Mercaptoethanol | Small Molecule | Polycomb protein SCMH1 | *SCMH1* | Experimental |
| Insulin pork | Biotech | Insulin-like growth factor 1 receptor | *IGF1R* | Approved |
| Mecasermin | Biotech | Insulin-like growth factor 1 receptor | *IGF1R* | Approved, Investigational |
| ATL1101 | Small molecule | Insulin-like growth factor 1 receptor | *IGF1R* | Investigational |
| rhIGFBP-3 | Small molecule | Insulin-like growth factor 1 receptor | *IGF1R* | Investigational |
| Linsitinib | Small molecule | Insulin-like growth factor 1 receptor | *IGF1R* | Investigational |
| (4Z)-6-bromo-4-({[4-(pyrrolidin-1-ylmethyl)phe | Small molecule | Insulin-like growth factor 1 receptor | *IGF1R* | Experimental |
| Cixutumumab | Biotech | Insulin-like growth factor 1 receptor | *IGF1R* | Investigational |
| Mecasermin rinfabate | Biotech | Insulin-like growth factor 1 receptor | *IGF1R* | Approved |
| Insulin detemir | Biotech | Insulin-like growth factor 1 receptor | *IGF1R* | Approved |
| Insulin glulisine | Biotech | Insulin-like growth factor 1 receptor | *IGF1R* | Approved |
| XL228 | Small molecule | Insulin-like growth factor 1 receptor | *IGF1R* | Investigational |
| Teprotumumab | Biotech | Insulin-like growth factor 1 receptor | *IGF1R* | Approved, Investigational |
| Somatrem | Biotech | Insulin-like growth factor 1 receptor | *IGF1R* | Approved, Investigational, Withdrawn |
| Brigatinib | Small molecule | Insulin-like growth factor 1 receptor | *IGF1R* | Approved, Investigational |
| Insulin human | Biotech | Insulin-like growth factor 1 receptor | *IGF1R* | Approved, Investigational |
| Insulin lispro | Biotech | Insulin-like growth factor 1 receptor | *IGF1R* | Approved |
| Insulin glargine | Biotech | Insulin-like growth factor 1 receptor | *IGF1R* | Approved |
| Insulin aspart | Biotech | Insulin-like growth factor 1 receptor | *IGF1R* | Approved |
| Phosphoaminophosphonic Acid-Adenylate Es | Small molecule | Insulin-like growth factor 1 receptor | *IGF1R* | Experimental |
| 3-[5-(1H-IMIDAZOL-1-YL)-7-METHYL-1H-BE | Small molecule | Insulin-like growth factor 1 receptor | *IGF1R* | Experimental |
| Insulin degludec | Biotech | Insulin-like growth factor 1 receptor | *IGF1R* | Approved |
| Disopyramide | Small molecule | Potassium voltage-gated channel subfamily H member 2 | *KCNH2* | Approved |
| Alfuzosin | Small molecule | Potassium voltage-gated channel subfamily H member 2 | *KCNH2* | Approved, Investigational |
| Imipramine | Small molecule | Potassium voltage-gated channel subfamily H member 2 | *KCNH2* | Approved |
| Cisapride | Small molecule | Potassium voltage-gated channel subfamily H member 2 | *KCNH2* | Approved, Investigational, Withdrawn |
| Quinidine | Small molecule | Potassium voltage-gated channel subfamily H member 2 | *KCNH2* | Approved, Investigational |
| Doxepin | Small molecule | Potassium voltage-gated channel subfamily H member 2 | *KCNH2* | Approved, Investigational |
| Propafenone | Small molecule | Potassium voltage-gated channel subfamily H member 2 | *KCNH2* | Approved |
| Flecainide | Small molecule | Potassium voltage-gated channel subfamily H member 2 | *KCNH2* | Approved, Withdrawn |

| | | | | |
|---|---|---|---|---|
| Tecastemizole | Small molecule | Potassium voltage-gated channel subfamily H member 2 | *KCNH2* | Investigational |
| Potassium nitrate | Small molecule | Potassium voltage-gated channel subfamily H member 2 | *KCNH2* | Approved |
| Isavuconazole | Small molecule | Potassium voltage-gated channel subfamily H member 2 | *KCNH2* | Approved, Investigational |
| Pitolisant | Small molecule | Potassium voltage-gated channel subfamily H member 2 | *KCNH2* | Approved, Investigational |
| Fluvoxamine | Small molecule | Potassium voltage-gated channel subfamily H member 2 | *KCNH2* | Approved, Investigational |
| Erythromycin | Small molecule | Potassium voltage-gated channel subfamily H member 2 | *KCNH2* | Approved, Investigational, Vet approved |
| Amsacrine | Small molecule | Potassium voltage-gated channel subfamily H member 2 | *KCNH2* | Approved, Investigational |
| Loratadine | Small molecule | Potassium voltage-gated channel subfamily H member 2 | *KCNH2* | Approved, Investigational |
| Fluoxetine | Small molecule | Potassium voltage-gated channel subfamily H member 2 | *KCNH2* | Approved, Vet approved |
| Ciprofloxacin | Small molecule | Potassium voltage-gated channel subfamily H member 2 | *KCNH2* | Approved, Vet approved |
| Astemizole | Small molecule | Potassium voltage-gated channel subfamily H member 2 | *KCNH2* | Approved, Withdrawn |
| Tamoxifen | Small molecule | Potassium voltage-gated channel subfamily H member 2 | *KCNH2* | Approved |
| Thioridazine | Small molecule | Potassium voltage-gated channel subfamily H member 2 | *KCNH2* | Approved, Withdrawn |
| Carvedilol | Small molecule | Potassium voltage-gated channel subfamily H member 2 | *KCNH2* | Approved, Investigational |
| Halofantrine | Small molecule | Potassium voltage-gated channel subfamily H member 2 | *KCNH2* | Approved |
| Dronedarone | Small molecule | Potassium voltage-gated channel subfamily H member 2 | *KCNH2* | Approved |
| Azimilide | Small molecule | Potassium voltage-gated channel subfamily H member 2 | *KCNH2* | Investigational |
| Pentoxyverine | Small molecule | Potassium voltage-gated channel subfamily H member 2 | *KCNH2* | Approved, Investigational |
| Chlorobutanol | Small molecule | Potassium voltage-gated channel subfamily H member 2 | *KCNH2* | Approved, Investigational, Vet approved |
| Dofetilide | Small molecule | Potassium voltage-gated channel subfamily H member 2 | *KCNH2* | Approved, Investigational |
| Phenytoin | Small molecule | Potassium voltage-gated channel subfamily H member 2 | *KCNH2* | Approved, Vet approved |
| Ibutilide | Small molecule | Potassium voltage-gated channel subfamily H member 2 | *KCNH2* | Approved |
| Terfenadine | Small molecule | Potassium voltage-gated channel subfamily H member 2 | *KCNH2* | Approved, Withdrawn |
| Prazosin | Small molecule | Potassium voltage-gated channel subfamily H member 2 | *KCNH2* | Approved |
| Chlorpromazine | Small molecule | Potassium voltage-gated channel subfamily H member 2 | *KCNH2* | Approved, Investigational, Vet approved |
| Sotalol | Small molecule | Potassium voltage-gated channel subfamily H member 2 | *KCNH2* | Approved |
| Hydroxyzine | Small molecule | Potassium voltage-gated channel subfamily H member 2 | *KCNH2* | Approved |
| Verapamil | Small molecule | Potassium voltage-gated channel subfamily H member 2 | *KCNH2* | Approved |
| Ketoconazole | Small molecule | Potassium voltage-gated channel subfamily H member 2 | *KCNH2* | Approved, Investigational |
| Procainamide | Small molecule | Potassium voltage-gated channel subfamily H member 2 | *KCNH2* | Approved |
| Perhexiline | Small molecule | Potassium voltage-gated channel subfamily H member 2 | *KCNH2* | Approved, Investigational |
| Pimozide | Small molecule | Potassium voltage-gated channel subfamily H member 2 | *KCNH2* | Approved |
| Miconazole | Small molecule | Potassium voltage-gated channel subfamily H member 2 | *KCNH2* | Approved, Investigational, Vet approved |
| Nefazodone | Small molecule | Potassium voltage-gated channel subfamily H member 2 | *KCNH2* | Approved, Withdrawn |

| | | | | |
|---|---|---|---|---|
| Terazosin | Small molecule | Potassium voltage-gated channel subfamily H member 2 | *KCNH2* | Approved |
| Clarithromycin | Small molecule | Potassium voltage-gated channel subfamily H member 2 | *KCNH2* | Approved |
| Bepridil | Small molecule | Potassium voltage-gated channel subfamily H member 2 | *KCNH2* | Approved, Withdrawn |
| Sertindole | Small molecule | Potassium voltage-gated channel subfamily H member 2 | *KCNH2* | Approved, Investigational, Withdrawn |
| Vernakalant | Small molecule | Potassium voltage-gated channel subfamily H member 2 | *KCNH2* | Approved, Investigational |
| Amitriptyline | Small molecule | HERG human cardiac K+ channel | *KCNH2* | Approved |
| Doxazosin | Small molecule | HERG human cardiac K+ channel | *KCNH2* | Approved |
| Amiodarone | Small molecule | HERG human cardiac K+ channel | *KCNH2* | Approved, Investigational |
| Promethazine | Small molecule | Voltage-gated Potassium Channels | *KCNH2* | Approved, Investigational |
| 2-[4-[4-[(5-cyclopropyl-1H-pyrazol-3-yl)amino | Small molecule | Calcium/calmodulin-dependent protein kinase type II subunit delta | *CAMK2D* | Experimental |
| (3Z)-N,N-DIMETHYL-2-OXO-3-(4,5,6,7-TETI | Small molecule | Calcium/calmodulin-dependent protein kinase type II subunit delta | *CAMK2D* | Experimental |
| Fostamatinib | Small molecule | Calcium/calmodulin-dependent protein kinase type II subunit delta | *CAMK2D* | Approved, Investigational |
| Miconazole | Small molecule | Small conductance calcium-activated potassium channel protein 2, Sma | *KCNN2, KCNN3* | Approved, Investigational, Vet approved |
| Trimebutine | Small molecule | Calcium-activated potassium channel | *KCNN2, KCNN3* | Approved |
| Procaine | Small molecule | Calcium-activated potassium channel | *KCNN2, KCNN3* | Approved, Investigational, Vet approved |
| Copper | Small molecule | Annexin A4 | *ANXA4* | Approved, Investigational |
| Fluocinolone acetonide | Small molecule | Annexin A4 | *ANXA4* | Approved, Investigational, Vet approved |
| Arsenic trioxide | Small molecule | Cyclin-dependent kinase inhibitor 1 | *CDKN1A* | Approved, Investigational |
| Ethanol | Small molecule | G protein-activated inward rectifier potassium channel 4 | *KCNJ5* | Approved |

#### Online Table XII. Results of MTAG for previous and novel loci associated with AF. SE: Standard Error, B: beta, Z: z-score; P: p-value.

| | | | | MEGASTROKE CES | | AF2018 | | MTAG CES | | | | MTAG AF | | | | | | | |
|---|---|---|---|---|---|---|---|---|---|---|---|---|---|---|---|---|---|---|---|
| **Index Variant** | **Locus** | **Priorityzed genes** | **Novelty of the l** | **Z** | **P** | **Z** | **P** | **B** | **SE** | **Z** | **P** | **B** | **SE** | **Z** | **P** | **PPA_exc_CES** | **PPA_exc_AF** | **PPA_Pleiotropy** | **Genetics with CES** |
| rs2885697-T | 1p34.2 | *SCMH1* | Known | -2.54 | 1.09E-02 | -6.27 | 2.88E-10 | -0.009 | 0.002 | -5.67 | 1.41E-08 | -0.009 | 0.001 | -6.32 | 2.70E-10 | 3.08E-07 | 2.83E-01 | 7.15E-01 | Shared Genetics |
| rs11590635-A | 1p33 | *AGBL4* | Known | 2.48 | 1.30E-02 | 5.87 | 4.12E-09 | 0.025 | 0.005 | 5.37 | 7.89E-08 | 0.027 | 0.005 | 5.93 | 3.11E-09 | 4.47E-07 | 2.42E-01 | 7.56E-01 | Shared Genetics |
| rs146518726-A | 1p32.3 | *MIR6500* | Known | 2.17 | 2.97E-02 | 7.75 | 8.27E-15 | 0.028 | 0.004 | 6.47 | 1.01E-10 | 0.031 | 0.004 | 7.68 | 1.53E-14 | 1.02E-12 | 2.98E-01 | 7.00E-01 | Shared Genetics |
| rs79187193-A | 1q21.2 | *GJA5* | Known | -2.37 | 1.78E-02 | -7.59 | 3.15E-14 | -0.020 | 0.003 | -6.47 | 9.79E-11 | -0.023 | 0.003 | -7.56 | 4.08E-14 | 3.47E-12 | 2.83E-01 | 7.14E-01 | Shared Genetics |
| rs11264280-T | 1q21.3 | *KCNN3* | Known | 2.49 | 1.28E-02 | 18.97 | 3.07E-79 | 0.022 | 0.002 | 14.22 | 7.17E-46 | 0.028 | 0.002 | 18.44 | 5.88E-76 | 3.74E-76 | 3.40E-01 | 6.54E-01 | Shared Genetics |
| rs577676-T | 1q24.2 | *LINC01142* | Known | -3.63 | 2.71E-04 | -13.78 | 1.62E-43 | -0.017 | 0.001 | -11.36 | 6.72E-30 | -0.019 | 0.001 | -13.63 | 2.84E-42 | 3.15E-40 | 1.13E-02 | 9.88E-01 | Shared Genetics |
| rs10753933-T | 1q32.1 | *PPFIA4* | Known | 3.72 | 2.09E-04 | 9.09 | 9.84E-20 | 0.012 | 0.001 | 8.24 | 1.70E-16 | 0.013 | 0.001 | 9.16 | 5.30E-20 | 9.86E-18 | 1.38E-02 | 9.86E-01 | Shared Genetics |
| rs11125871-T | 2p15 | *USP34* | Known | -3.24 | 1.17E-03 | -5.79 | 6.42E-09 | -0.009 | 0.002 | -5.75 | 9.19E-09 | -0.009 | 0.001 | -5.95 | 2.71E-09 | 1.53E-07 | 1.25E-01 | 8.74E-01 | Shared Genetics |
| rs6747542-T | 2p13.3 | *GMCL1,ANXA4* | Known | 2.61 | 8.86E-03 | 8.27 | 1.10E-16 | 0.010 | 0.001 | 7.06 | 1.63E-12 | 0.012 | 0.001 | 8.23 | 1.82E-16 | 4.86E-14 | 2.47E-01 | 7.52E-01 | Shared Genetics |
| rs12621647-A | 2q22.3 | *TEX41* | Known | 3.25 | 1.15E-03 | 5.53 | 3.45E-08 | 0.013 | 0.002 | 5.58 | 2.45E-08 | 0.013 | 0.002 | 5.70 | 1.20E-08 | 4.67E-07 | 9.86E-02 | 9.00E-01 | Shared Genetics |
| rs56181519-T | 2q31.1 | *WIPF1* | Known | -2.73 | 6.31E-03 | -8.60 | 6.46E-18 | -0.012 | 0.002 | -7.35 | 1.98E-13 | -0.014 | 0.002 | -8.56 | 1.11E-17 | 8.99E-15 | 1.81E-01 | 8.18E-01 | Shared Genetics |
| rs2288327-A | 2q31.2 | *TTN,MIR548N,FKBP7,T* | Known | -2.36 | 1.80E-02 | -10.33 | 7.26E-25 | -0.016 | 0.002 | -8.31 | 9.70E-17 | -0.020 | 0.002 | -10.17 | 2.80E-24 | 5.54E-22 | 3.01E-01 | 6.98E-01 | Shared Genetics |
| rs1458038-T | 4q21.21 | *FGF5* | Known | 2.36 | 1.82E-02 | 6.03 | 1.74E-09 | 0.009 | 0.002 | 5.41 | 6.36E-08 | 0.009 | 0.002 | 6.06 | 1.36E-09 | 1.52E-07 | 2.19E-01 | 7.80E-01 | Shared Genetics |
| rs17042098-A | 4q25 | *PITX2* | Known | 12.72 | 3.72E-37 | 38.01 | 8,97E-318 | 0.073 | 0.002 | 32.86 | 7.73E-237 | 0.087 | 0.002 | 37.93 | 0 | 3.0416700000136e-313 | 2.40E-34 | 1.00E+00 | Shared Genetics |
| rs55754224-T | 4q26 | *CAMK2D* | Known | 2.80 | 5.05E-03 | 7.39 | 2.15E-13 | 0.011 | 0.002 | 6.57 | 4.92E-11 | 0.012 | 0.002 | 7.41 | 1.22E-13 | 2.72E-11 | 1.94E-01 | 8.05E-01 | Shared Genetics |
| rs337705-T | 5q22.3 | *KCNN2* | Known | -2.56 | 1.04E-02 | -8.29 | 1.63E-16 | -0.011 | 0.002 | -7.05 | 1.77E-12 | -0.012 | 0.001 | -8.25 | 1.57E-16 | 1.09E-13 | 2.80E-01 | 7.19E-01 | Shared Genetics |
| rs17171711-T | 5q31.2 | *WNT8A,NPY6R,MYOT,F* | Known | 3.79 | 1.56E-04 | 12.48 | 1.95E-35 | 0.020 | 0.002 | 10.57 | 4.04E-26 | 0.023 | 0.002 | 12.41 | 2.35E-35 | 2.28E-33 | 7.97E-03 | 9.92E-01 | Shared Genetics |
| rs6882776-A | 5q35.1 | *NKX2-5* | Known | -4.86 | 1.21E-06 | -9.61 | 9.64E-22 | -0.015 | 0.002 | -9.24 | 2.49E-20 | -0.015 | 0.002 | -9.80 | 1.14E-22 | 2.48E-20 | 7.95E-05 | 1.00E+00 | Shared Genetics |
| rs3176326-A | 6p21.2 | *CDKN1A,PANDAR,PI16* | Known | -3.98 | 6.66E-05 | -7.36 | 1.42E-13 | -0.013 | 0.002 | -7.23 | 5.01E-13 | -0.013 | 0.002 | -7.54 | 4.59E-14 | 1.46E-11 | 3.92E-03 | 9.96E-01 | Shared Genetics |
| **rs12211255-A** | **6q14.1** | ***FILIP1*** | **Novel** | **3.42** | **6.24E-04** | **5.44** | **5.06E-08** | **0.013** | **0.002** | **5.61** | **2.06E-08** | **0.013** | **0.002** | **5.63** | **1.78E-08** | 9.65E-05 | 1.82E-02 | 9.61E-01 | **Shared Genetics** |
| rs72966339-T | 6q22.31 | *HSF2* | Known | -2.69 | 7.27E-03 | -8.93 | 7.42E-19 | -0.012 | 0.002 | -7.55 | 4.34E-14 | -0.013 | 0.001 | -8.87 | 7.17E-19 | 5.43E-17 | 1.86E-01 | 8.14E-01 | Shared Genetics |
| rs11773845-A | 7q31.2 | *CAV1,CAV2* | Known | 4.12 | 3.89E-05 | 15.73 | 2.39E-55 | 0.019 | 0.001 | 12.95 | 2.25E-38 | 0.022 | 0.001 | 15.55 | 1.47E-54 | 2.18E-53 | 2.09E-03 | 9.98E-01 | Shared Genetics |
| rs55985730-T | 7q32.1 | *OPN1SW,CALU* | Known | -2.84 | 4.47E-03 | -5.82 | 5.24E-09 | -0.017 | 0.003 | -5.54 | 3.06E-08 | -0.018 | 0.003 | -5.92 | 3.19E-09 | 1.68E-06 | 1.21E-01 | 8.74E-01 | Shared Genetics |
| rs2269001-A | 7q36.1 | *KCNH2* | Known | -2.89 | 3.94E-03 | -6.63 | 4.01E-11 | -0.011 | 0.002 | -6.11 | 1.00E-09 | -0.012 | 0.002 | -6.70 | 2.06E-11 | 1.16E-08 | 2.60E-01 | 7.32E-01 | Shared Genetics |
| rs6994744-A | 8q24.3 | *PTK2* | Known | -2.26 | 2.34E-02 | -6.14 | 1.10E-09 | -0.008 | 0.001 | -5.42 | 5.82E-08 | -0.009 | 0.001 | -6.15 | 7.70E-10 | 1.10E-07 | 2.14E-01 | 7.84E-01 | Shared Genetics |
| rs10821415-A | 9q22.22 | *C9orf3* | Known | 3.16 | 1.57E-03 | 12.25 | 2.92E-34 | 0.015 | 0.001 | 10.06 | 8.28E-24 | 0.017 | 0.001 | 12.11 | 9.38E-34 | 8.90E-31 | 4.96E-02 | 9.46E-01 | Shared Genetics |
| rs7096385-T | 10q21.3 | *SIRT1,MYPN* | Known | 2.49 | 1.26E-02 | 5.44 | 4.87E-08 | 0.015 | 0.003 | 5.08 | 3.74E-07 | 0.013 | 0.002 | 5.51 | 3.52E-08 | 2.80E-06 | 2.25E-01 | 7.70E-01 | Shared Genetics |
| rs7915134-T | 10q22.2 | *SYNPO2L,NUDT13,MY(* | Known | -3.53 | 4.12E-04 | -12.29 | 1.42E-34 | -0.022 | 0.002 | -10.30 | 7.18E-25 | -0.025 | 0.002 | -12.20 | 3.26E-34 | 4.35E-32 | 1.83E-02 | 9.81E-01 | Shared Genetics |
| rs11001667-A | 10q22.3 | *C10orf11* | Known | -2.46 | 1.40E-02 | -6.16 | 7.55E-10 | -0.011 | 0.002 | -5.55 | 2.85E-08 | -0.012 | 0.002 | -6.20 | 5.68E-10 | 2.00E-29 | 1.59E-03 | 9.98E-01 | Shared Genetics |
| rs11598047-A | 10q24.33 | *NEURL1* | Known | -3.27 | 1.06E-03 | -17.08 | 8.95E-66 | -0.026 | 0.002 | -13.38 | 7.50E-41 | -0.032 | 0.002 | -16.73 | 7.42E-63 | 1.47E-61 | 6.45E-02 | 9.33E-01 | Shared Genetics |
| rs10749053-T | 10q25.2 | *RBM20* | Known | 4.14 | 3.34E-05 | 5.72 | 1.05E-08 | 0.013 | 0.002 | 6.21 | 5.33E-10 | 0.012 | 0.002 | 5.99 | 2.05E-09 | 7.10E-08 | 9.87E-04 | 9.99E-01 | Shared Genetics |
| rs76097649-A | 11q24.3 | *KCNJ5* | Known | 2.91 | 3.65E-03 | 9.28 | 1.26E-20 | 0.021 | 0.003 | 7.91 | 2.52E-15 | 0.022 | 0.002 | 9.24 | 2.49E-20 | 4.59E-18 | 8.07E-02 | 9.19E-01 | Shared Genetics |
| rs883079-T | 12q24.21 | *TBX5* | Known | 3.55 | 3.95E-04 | 13.26 | 2.84E-40 | 0.018 | 0.002 | 10.96 | 5.97E-28 | 0.020 | 0.002 | 13.12 | 2.60E-39 | 4.55E-38 | 2.72E-02 | 9.73E-01 | Shared Genetics |
| rs10773657-A | 12q24.31 | *HIP1R* | Known | -2.04 | 4.13E-02 | -5.58 | 2.54E-08 | -0.011 | 0.002 | -4.92 | 8.57E-07 | -0.011 | 0.002 | -5.59 | 2.23E-08 | 2.14E-06 | 2.51E-01 | 7.44E-01 | Shared Genetics |
| rs2738413-A | 14q23.2 | *SYNE2,MIR548AZ,ESR:* | Known | 2.97 | 3.03E-03 | 11.61 | 2.55E-31 | 0.014 | 0.001 | 9.52 | 1.67E-21 | 0.016 | 0.001 | 11.47 | 1.80E-30 | 5.15E-28 | 1.52E-01 | 8.46E-01 | Shared Genetics |
| rs7172038-T | 15q24.1 | *HCN4* | Known | -2.73 | 6.43E-03 | -12.58 | 4.78E-36 | -0.020 | 0.002 | -10.04 | 1.02E-23 | -0.024 | 0.002 | -12.37 | 3.76E-35 | 2.13E-33 | 2.54E-01 | 7.45E-01 | Shared Genetics |
| rs12908004-A | 15q25.1 | *ARNT2* | Known | -3.28 | 1.03E-03 | -8.13 | 4.12E-16 | -0.014 | 0.002 | -7.35 | 2.03E-13 | -0.016 | 0.002 | -8.19 | 2.65E-16 | 4.09E-13 | 4.22E-02 | 9.57E-01 | Shared Genetics |
| rs6598541-A | 15q26.3 | *IGF1R* | Known | 2.69 | 7.17E-03 | 6.35 | 2.22E-10 | 0.009 | 0.002 | 5.81 | 6.21E-09 | 0.009 | 0.001 | 6.41 | 1.48E-10 | 2.20E-07 | 2.74E-01 | 7.23E-01 | Shared Genetics |
| rs140185678-A | 16p13.3 | *RPL3L* | Known | 2.92 | 3.54E-03 | 7.61 | 2.43E-14 | 0.026 | 0.004 | 6.79 | 1.13E-11 | 0.029 | 0.004 | 7.64 | 2.13E-14 | 6.14E-12 | 1.13E-01 | 8.87E-01 | Shared Genetics |
| rs2106261-T | 16q22.3 | *ZFHX3* | Known | 6.75 | 1.63E-11 | 20.23 | 4.97E-91 | 0.034 | 0.002 | 17.48 | 2.02E-68 | 0.037 | 0.002 | 20.19 | 1.20E-90 | 3.34E-88 | 2.85E-09 | 1.00E+00 | Shared Genetics |
| rs9899183-T | 17p13.1 | *TNFSF12,TNFSF12-TNI* | Known | 2.72 | 6.67E-03 | 6.03 | 2.02E-09 | 0.009 | 0.002 | 5.61 | 2.05E-08 | 0.010 | 0.002 | 6.10 | 1.03E-09 | 2.08E-07 | 1.50E-01 | 8.48E-01 | Shared Genetics |
| rs199497-T | 17q21.31 | *WNT3* | Known | -1.88 | 6.06E-02 | -6.52 | 8.94E-11 | -0.011 | 0.002 | -5.47 | 4.61E-08 | -0.012 | 0.002 | -6.47 | 9.84E-11 | 2.73E-09 | 7.96E-03 | 9.92E-01 | Shared Genetics |
| rs2834618-T | 21q22.12 | *LINC01426* | Known | 3.31 | 9.48E-04 | 8.43 | 3.41E-17 | 0.017 | 0.002 | 7.56 | 3.96E-14 | 0.019 | 0.002 | 8.47 | 2.37E-17 | 1.43E-14 | 3.76E-02 | 9.62E-01 | Shared Genetics |
| rs284277-A | 1p36.22 | *CASZ1* | Known | -2.16 | 3.05E-02 | -6.12 | 1.25E-09 | -0.008 | 0.002 | -5.35 | 8.67E-08 | -0.009 | 0.001 | -6.12 | 9.44E-10 | 6.51E-07 | 4.66E-01 | 5.30E-01 | ? |
| rs7529220-T | 1p36.12 | *HSPG2* | Known | -0.43 | 6.68E-01 | -6.34 | 1.98E-10 | -0.010 | 0.002 | -4.52 | 6.14E-06 | -0.012 | 0.002 | -6.11 | 1.00E-09 | 2.70E-07 | 8.44E-01 | 1.53E-01 | Not Shared |
| rs1545300-T | 1p13.2 | *KCND3* | Known | 1.07 | 2.85E-01 | -7.64 | 1.48E-14 | -0.007 | 0.002 | -4.56 | 5.18E-06 | -0.011 | 0.002 | -7.17 | 7.62E-13 | 5.89E-11 | 7.84E-01 | 2.01E-01 | Not Shared |
| rs4073778-A | 1p13.1 | *CASQ2* | Known | 0.56 | 5.75E-01 | 7.25 | 4.96E-13 | 0.008 | 0.001 | 5.22 | 1.84E-07 | 0.010 | 0.001 | 7.00 | 2.52E-12 | 1.10E-09 | 8.81E-01 | 1.15E-01 | Not Shared |

| | | | | | | | | | | | | | | | | | | | |
|---|---|---|---|---|---|---|---|---|---|---|---|---|---|---|---|---|---|---|---|
| rs4951258-A | 1q32.1 | *NUCKS1,SLC41A1* | Known | -0.83 | 4.05E-01 | 5.61 | 2.10E-08 | 0.005 | 0.001 | 3.32 | 9.01E-04 | 0.008 | 0.001 | 5.26 | 1.47E-07 | 2.23E-05 | 8.37E-01 | 1.30E-01 | Not Shared |
| rs6546620-T | 2p23.3 | *KIF3C* | Known | -1.62 | 1.07E-01 | -7.00 | 3.19E-12 | -0.010 | 0.002 | -5.64 | 1.69E-08 | -0.012 | 0.002 | -6.89 | 5.43E-12 | 3.31E-09 | 7.11E-01 | 2.86E-01 | Not Shared |
| rs2723064-T | 2p14 | *CEP68* | Known | 1.60 | 1.11E-01 | 9.56 | 1.43E-21 | 0.011 | 0.002 | 7.36 | 1.86E-13 | 0.014 | 0.001 | 9.34 | 9.96E-21 | 1.39E-18 | 7.18E-01 | 2.79E-01 | Not Shared |
| rs72926475-A | 2p11.2 | *REEP1* | Known | -1.71 | 8.83E-02 | -6.70 | 2.37E-11 | -0.012 | 0.002 | -5.49 | 4.09E-08 | -0.014 | 0.002 | -6.62 | 3.71E-11 | 1.18E-08 | 6.77E-01 | 3.21E-01 | Not Shared |
| rs113949548-T | 2q14.3 | *GYPC* | Known | 0.76 | 4.45E-01 | 6.44 | 1.09E-10 | 0.011 | 0.002 | 4.78 | 1.76E-06 | 0.014 | 0.002 | 6.25 | 4.13E-10 | 1.06E-07 | 8.03E-01 | 1.94E-01 | Not Shared |
| rs56326533-T | 2q33.1 | *SPATS2L* | Known | -2.20 | 2.74E-02 | -10.07 | 6.28E-24 | -0.012 | 0.002 | -8.05 | 8.36E-16 | -0.014 | 0.001 | -9.91 | 3.95E-23 | 9.75E-21 | 4.63E-01 | 5.35E-01 | ? |
| rs6738011-T | 2q34 | *ERBB4* | Known | 1.53 | 1.26E-01 | 6.33 | 3.05E-10 | 0.009 | 0.002 | 5.14 | 2.78E-07 | 0.011 | 0.002 | 6.24 | 4.42E-10 | 8.81E-08 | 6.95E-01 | 3.02E-01 | Not Shared |
| rs4642101-T | 3p25.2 | *CAND2* | Known | -2.00 | 4.57E-02 | -10.23 | 2.95E-24 | -0.012 | 0.002 | -8.04 | 9.14E-16 | -0.015 | 0.001 | -10.03 | 1.12E-23 | 2.69E-21 | 4.95E-01 | 5.04E-01 | ? |
| rs73041705-T | 3p24.2 | *THRB* | Known | 0.34 | 7.34E-01 | 6.07 | 1.55E-09 | 0.007 | 0.002 | 4.29 | 1.79E-05 | 0.009 | 0.002 | 5.84 | 5.18E-09 | 7.83E-07 | 8.81E-01 | 1.14E-01 | Not Shared |
| rs10428132-T | 3p22.2 | *SCN10A,SCN5A* | Known | -0.60 | 5.52E-01 | -9.21 | 2.72E-20 | -0.010 | 0.001 | -6.55 | 5.62E-11 | -0.013 | 0.001 | -8.87 | 7.19E-19 | 7.60E-17 | 8.86E-01 | 1.09E-01 | Not Shared |
| rs34080181-A | 3p14.1 | *LRIG1,SLC25A26* | Known | -1.81 | 7.07E-02 | -6.46 | 1.28E-10 | -0.008 | 0.002 | -5.39 | 7.18E-08 | -0.009 | 0.001 | -6.41 | 1.50E-10 | 1.55E-07 | 6.56E-01 | 3.42E-01 | Not Shared |
| rs17005647-T | 3p14.1 | *FRMD4B* | Known | -0.89 | 3.73E-01 | 5.99 | 2.70E-09 | 0.005 | 0.002 | 3.54 | 4.03E-04 | 0.008 | 0.001 | 5.61 | 2.07E-08 | 8.37E-07 | 8.56E-01 | 1.38E-01 | Not Shared |
| rs6771054-T | 3p11.1 | *EPHA3* | Known | 0.88 | 3.81E-01 | 6.72 | 2.42E-11 | 0.008 | 0.001 | 5.03 | 4.80E-07 | 0.009 | 0.001 | 6.53 | 6.45E-11 | 1.98E-08 | 8.07E-01 | 1.89E-01 | Not Shared |
| rs10804493-A | 3q13.2 | *PHLDB2,PLCXD2* | Known | 0.72 | 4.71E-01 | 7.97 | 1.63E-15 | 0.009 | 0.002 | 5.79 | 7.05E-09 | 0.011 | 0.001 | 7.71 | 1.28E-14 | 2.78E-12 | 8.74E-01 | 1.22E-01 | Not Shared |
| rs1278493-A | 3q22.3 | *PPP2R3A* | Known | -0.23 | 8.21E-01 | -5.72 | 8.77E-09 | -0.006 | 0.001 | -3.99 | 6.56E-05 | -0.008 | 0.001 | -5.49 | 3.91E-08 | 5.42E-06 | 8.73E-01 | 1.12E-01 | Not Shared |
| rs75880040-T | 3q26.33 | *GNB4* | Known | -1.07 | 2.88E-01 | -5.84 | 5.21E-09 | -0.008 | 0.002 | -4.54 | 5.55E-06 | -0.010 | 0.002 | -5.71 | 1.12E-08 | 8.86E-07 | 8.37E-01 | 1.58E-01 | Not Shared |
| rs60902112-T | 3q29 | *XXYLT1* | Known | 1.07 | 2.84E-01 | 5.63 | 1.72E-08 | 0.008 | 0.002 | 4.41 | 1.04E-05 | 0.009 | 0.002 | 5.52 | 3.43E-08 | 1.16E-05 | 7.95E-01 | 1.66E-01 | Not Shared |
| rs3960788-T | 4q24 | *SLC9B1* | Known | -2.41 | 1.58E-02 | -5.45 | 5.98E-08 | -0.008 | 0.001 | -5.04 | 4.58E-07 | -0.008 | 0.001 | -5.51 | 3.55E-08 | 2.26E-06 | 6.58E-01 | 3.34E-01 | Not Shared |
| rs6839459-A | 4q31.23 | *ARHGAP10* | Known | -1.25 | 2.13E-01 | -6.72 | 1.64E-11 | -0.016 | 0.003 | -5.24 | 1.61E-07 | -0.019 | 0.003 | -6.58 | 4.84E-11 | 1.41E-08 | 6.74E-01 | 3.17E-01 | Not Shared |
| rs74500426-T | 4q34.1 | *HAND2,HAND2-AS1* | Known | 0.33 | 7.40E-01 | -7.25 | 4.29E-13 | -0.013 | 0.003 | -4.71 | 2.49E-06 | -0.018 | 0.003 | -6.89 | 5.70E-12 | 2.33E-10 | 8.28E-01 | 1.70E-01 | Not Shared |
| rs6596717-A | 5q21.3 | *LOC102467213* | Known | -1.15 | 2.53E-01 | -5.94 | 3.00E-09 | -0.007 | 0.002 | -4.66 | 3.16E-06 | -0.008 | 0.001 | -5.82 | 5.80E-09 | 4.95E-07 | 8.19E-01 | 1.77E-01 | Not Shared |
| rs2012809-A | 5q23.3 | *SLC27A6* | Known | -1.03 | 3.04E-01 | -6.19 | 4.92E-10 | -0.009 | 0.002 | -4.76 | 1.91E-06 | -0.010 | 0.002 | -6.05 | 1.48E-09 | 6.61E-07 | 7.41E-01 | 2.55E-01 | Not Shared |
| rs6580277-A | 5q31.1 | *NR3C1* | Known | -1.24 | 2.15E-01 | -8.48 | 1.64E-17 | -0.011 | 0.002 | -6.43 | 1.30E-10 | -0.014 | 0.002 | -8.26 | 1.45E-16 | 4.71E-14 | 8.28E-01 | 1.56E-01 | Not Shared |
| rs12188351-A | 5q34 | *SLIT3* | Known | 1.93 | 5.35E-02 | 5.97 | 2.52E-09 | 0.016 | 0.003 | 5.12 | 3.03E-07 | 0.018 | 0.003 | 5.95 | 2.75E-09 | 2.16E-07 | 5.04E-01 | 4.93E-01 | ? |
| rs73366713-A | 6p22.3 | *ATXN1* | Known | -1.70 | 8.80E-02 | -10.45 | 1.53E-25 | -0.017 | 0.002 | -8.02 | 1.03E-15 | -0.021 | 0.002 | -10.21 | 1.87E-24 | 2.12E-22 | 6.42E-01 | 3.56E-01 | Not Shared |
| rs34969716-A | 6p22.3 | *KDM1B,DEK* | Known | 1.44 | 1.50E-01 | 9.00 | 1.60E-19 | 0.011 | 0.002 | 6.89 | 5.52E-12 | 0.013 | 0.002 | 8.78 | 1.60E-18 | 1.01E-15 | 7.28E-01 | 2.68E-01 | Not Shared |
| rs13210074-A | 6q14.3 | *CGA* | Known | -1.48 | 1.38E-01 | -6.41 | 1.48E-10 | -0.008 | 0.002 | -5.17 | 2.36E-07 | -0.009 | 0.001 | -6.31 | 2.70E-10 | 1.13E-08 | 7.95E-01 | 2.03E-01 | Not Shared |
| rs4946333-A | 6q22.31 | *SLC35F1,PLN* | Known | -2.36 | 1.82E-02 | -9.68 | 5.47E-22 | -0.012 | 0.001 | -7.87 | 3.49E-15 | -0.014 | 0.001 | -9.55 | 1.29E-21 | 1.47E-19 | 4.68E-01 | 5.31E-01 | ? |
| rs117984853-T | 6q25.1 | *UST* | Known | 1.81 | 7.09E-02 | 10.23 | 1.34E-24 | 0.019 | 0.002 | 7.93 | 2.14E-15 | 0.023 | 0.002 | 10.01 | 1.41E-23 | 2.73E-21 | 5.69E-01 | 4.29E-01 | ? |
| rs12154315-T | 7p21.2 | *DGKB* | Known | 1.86 | 6.28E-02 | 7.00 | 3.38E-12 | 0.010 | 0.002 | 5.78 | 7.61E-09 | 0.011 | 0.002 | 6.92 | 4.38E-12 | 1.99E-09 | 6.60E-01 | 3.38E-01 | Not Shared |
| rs6948592-T | 7p15.1 | *CREB5* | Known | 1.06 | 2.86E-01 | 6.09 | 9.99E-10 | 0.007 | 0.002 | 4.71 | 2.46E-06 | 0.009 | 0.001 | 5.95 | 2.68E-09 | 1.01E-06 | 8.61E-01 | 1.32E-01 | Not Shared |
| rs35005436-T | 7q11.23 | *GTF2I,LOC101926943,* | Known | 0.02 | 9.84E-01 | -6.31 | 3.34E-10 | -0.009 | 0.002 | -4.25 | 2.15E-05 | -0.012 | 0.002 | -6.03 | 1.68E-09 | 8.76E-07 | 8.53E-01 | 1.41E-01 | Not Shared |
| rs56201652-A | 7q21.2 | *CDK6* | Known | -2.37 | 1.74E-02 | -7.08 | 1.74E-12 | -0.010 | 0.002 | -6.12 | 9.14E-10 | -0.011 | 0.002 | -7.07 | 1.59E-12 | 3.83E-10 | 4.30E-01 | 5.68E-01 | ? |
| rs35620480-A | 8p23.1 | *GATA4* | Known | -0.48 | 6.29E-01 | -5.87 | 5.15E-09 | -0.008 | 0.002 | -4.24 | 2.28E-05 | -0.011 | 0.002 | -5.67 | 1.43E-08 | 4.25E-06 | 7.89E-01 | 1.98E-01 | Not Shared |
| rs7508-A | 8p22 | *ASAH1* | Known | 1.97 | 4.80E-02 | 9.48 | 1.69E-21 | 0.012 | 0.002 | 7.52 | 5.57E-14 | 0.015 | 0.002 | 9.31 | 1.29E-20 | 1.16E-17 | 5.59E-01 | 4.39E-01 | ? |
| rs7834729-T | 8p21.3 | *XPO7* | Known | -1.95 | 5.19E-02 | -6.28 | 3.55E-10 | -0.012 | 0.002 | -5.34 | 9.25E-08 | -0.014 | 0.002 | -6.25 | 4.19E-10 | 5.63E-08 | 5.01E-01 | 4.95E-01 | ? |
| rs62521286-A | 8q24.13 | *FBXO32* | Known | -0.71 | 4.75E-01 | -8.90 | 4.50E-19 | -0.019 | 0.003 | -6.42 | 1.40E-10 | -0.024 | 0.003 | -8.60 | 8.10E-18 | 1.19E-15 | 8.07E-01 | 1.91E-01 | Not Shared |
| rs2274115-A | 9q34.3 | *LHX3* | Known | -2.11 | 3.51E-02 | -6.41 | 1.69E-10 | -0.009 | 0.002 | -5.52 | 3.41E-08 | -0.010 | 0.002 | -6.39 | 1.65E-10 | 1.04E-07 | 5.08E-01 | 4.90E-01 | ? |
| rs10822156-T | 10q21.3 | *REEP3,NRBF2* | Known | -1.44 | 1.50E-01 | -6.94 | 4.21E-12 | -0.008 | 0.001 | -5.50 | 3.80E-08 | -0.010 | 0.001 | -6.81 | 9.53E-12 | 7.33E-10 | 7.89E-01 | 2.05E-01 | Not Shared |
| rs4757877-A | 11p15.1 | *NAV2* | Known | 1.90 | 5.71E-02 | 9.27 | 2.93E-20 | 0.013 | 0.002 | 7.33 | 2.22E-13 | 0.015 | 0.002 | 9.10 | 9.16E-20 | 1.06E-17 | 5.72E-01 | 4.26E-01 | ? |
| rs2156664-T | 11q24.1 | *SORL1* | Known | 0.15 | 8.82E-01 | 5.86 | 4.93E-09 | 0.007 | 0.002 | 4.04 | 5.38E-05 | 0.009 | 0.002 | 5.61 | 1.98E-08 | 1.12E-06 | 8.76E-01 | 1.18E-01 | Not Shared |
| rs4963776-T | 12p12.1 | *LINC00477* | Known | -2.80 | 5.18E-03 | -10.38 | 1.84E-25 | -0.016 | 0.002 | -8.59 | 8.73E-18 | -0.019 | 0.002 | -10.27 | 9.70E-25 | 1.69E-22 | 4.54E-01 | 5.43E-01 | ? |
| rs17380837-T | 12p12.1 | *SSPN* | Known | 0.75 | 4.55E-01 | -6.96 | 4.80E-12 | -0.007 | 0.002 | -4.28 | 1.90E-05 | -0.010 | 0.002 | -6.55 | 5.61E-11 | 1.02E-08 | 8.69E-01 | 1.28E-01 | Not Shared |
| rs12809354-T | 12p11.21 | *PKP2* | Known | -1.89 | 5.91E-02 | -7.64 | 2.89E-14 | -0.013 | 0.002 | -6.23 | 4.79E-10 | -0.015 | 0.002 | -7.54 | 4.76E-14 | 6.62E-11 | 5.69E-01 | 4.25E-01 | ? |
| rs2860482-A | 12q13.3 | *NACA* | Known | 0.33 | 7.45E-01 | 7.11 | 1.21E-12 | 0.008 | 0.002 | 4.98 | 6.29E-07 | 0.011 | 0.002 | 6.83 | 8.46E-12 | 1.32E-09 | 8.80E-01 | 1.13E-01 | Not Shared |
| rs71454237-A | 12q15 | *LRRC10* | Known | -0.50 | 6.14E-01 | -7.38 | 1.78E-13 | -0.009 | 0.002 | -5.27 | 1.37E-07 | -0.012 | 0.002 | -7.12 | 1.11E-12 | 4.92E-10 | 8.22E-01 | 1.74E-01 | Not Shared |
| rs12426679-T | 12q21.2 | *PHLDA1* | Known | -0.71 | 4.77E-01 | -5.84 | 4.95E-09 | -0.006 | 0.001 | -4.34 | 1.41E-05 | -0.008 | 0.001 | -5.67 | 1.46E-08 | 6.30E-06 | 8.52E-01 | 1.30E-01 | Not Shared |
| rs6560886-T | 12q24.33 | *FBRSL1* | Known | -1.31 | 1.90E-01 | -5.67 | 1.49E-08 | -0.008 | 0.002 | -4.57 | 4.90E-06 | -0.010 | 0.002 | -5.58 | 2.39E-08 | 8.43E-06 | 6.45E-01 | 3.28E-01 | Not Shared |
| rs9506925-T | 13q12.12 | *LINC00540,LINC00621,* | Known | 2.05 | 4.03E-02 | 5.99 | 2.72E-09 | 0.009 | 0.002 | 5.20 | 1.99E-07 | 0.010 | 0.002 | 5.98 | 2.23E-09 | 2.66E-06 | 4.98E-01 | 4.94E-01 | ? |

| | | | | | | | | | | | | | | | | | | | |
|---|---|---|---|---|---|---|---|---|---|---|---|---|---|---|---|---|---|---|---|
| rs35569628-T | 13q34 | *CUL4A* | Known | 1.40 | 1.62E-01 | 5.65 | 1.38E-08 | 0.008 | 0.002 | 4.61 | 4.11E-06 | 0.009 | 0.002 | 5.58 | 2.46E-08 | 1.23E-06 | 7.36E-01 | 2.58E-01 | Not Shared |
| rs28631169-T | 14q11.2 | *MYH6,MYH7* | Known | 1.38 | 1.69E-01 | 6.21 | 5.35E-10 | 0.009 | 0.002 | 4.98 | 6.43E-07 | 0.011 | 0.002 | 6.11 | 9.76E-10 | 5.00E-07 | 8.45E-01 | 1.49E-01 | Not Shared |
| rs7140396-A | 14q12 | *AKAP6* | Known | 1.93 | 5.38E-02 | 9.20 | 2.42E-20 | 0.012 | 0.002 | 7.30 | 2.80E-13 | 0.014 | 0.002 | 9.03 | 1.66E-19 | 1.66E-16 | 6.00E-01 | 3.98E-01 | ? |
| rs73241997-T | 14q13.1 | *CFL2* | Known | 1.29 | 1.96E-01 | 7.88 | 2.94E-15 | 0.012 | 0.002 | 6.05 | 1.41E-09 | 0.016 | 0.002 | 7.70 | 1.41E-14 | 6.02E-12 | 7.59E-01 | 2.38E-01 | Not Shared |
| rs74884082-T | 14q24.2 | *DPF3* | Known | -0.22 | 8.25E-01 | -6.32 | 3.48E-10 | -0.007 | 0.002 | -4.39 | 1.11E-05 | -0.010 | 0.002 | -6.07 | 1.30E-09 | 5.41E-07 | 8.58E-01 | 1.37E-01 | Not Shared |
| rs10873298-T | 14q24.3 | *IRF2BPL* | Known | -0.15 | 8.85E-01 | -5.81 | 7.07E-09 | -0.006 | 0.002 | -4.01 | 6.17E-05 | -0.008 | 0.001 | -5.57 | 2.53E-08 | 4.94E-06 | 8.77E-01 | 1.06E-01 | Not Shared |
| rs7170477-A | 15q22.31 | *HERC1* | Known | 2.15 | 3.12E-02 | 5.46 | 4.98E-08 | 0.008 | 0.002 | 4.90 | 9.48E-07 | 0.008 | 0.002 | 5.49 | 4.05E-08 | 5.87E-06 | 5.05E-01 | 4.72E-01 | ? |
| rs7225165-A | 17p13.3 | *YWHAE,CRK,MYO1C* | Known | -0.53 | 5.98E-01 | -5.90 | 3.20E-09 | -0.010 | 0.002 | -4.28 | 1.85E-05 | -0.013 | 0.002 | -5.71 | 1.16E-08 | 1.97E-06 | 7.63E-01 | 2.28E-01 | Not Shared |
| rs55941572-T | 17p12 | *MYOCD* | Known | 1.18 | 2.39E-01 | 6.60 | 4.45E-11 | 0.012 | 0.002 | 5.12 | 3.04E-07 | 0.015 | 0.002 | 6.45 | 1.09E-10 | 7.68E-09 | 7.60E-01 | 2.38E-01 | Not Shared |
| rs7359623-T | 17q12 | *ZPBP2,GSDMB,ORMDL* | Known | -0.31 | 7.59E-01 | -6.64 | 4.39E-11 | -0.007 | 0.001 | -4.66 | 3.23E-06 | -0.009 | 0.001 | -6.38 | 1.77E-10 | 9.37E-09 | 8.94E-01 | 1.03E-01 | Not Shared |
| rs7224711-T | 17q25.3 | *CYTH1,USP36* | Known | -1.40 | 1.60E-01 | -5.53 | 3.72E-08 | -0.007 | 0.001 | -4.53 | 5.93E-06 | -0.008 | 0.001 | -5.46 | 4.69E-08 | 4.12E-06 | 7.92E-01 | 1.93E-01 | Not Shared |
| rs9953366-T | 18q21.1 | *SMAD7* | Known | -1.82 | 6.91E-02 | -6.71 | 1.82E-11 | -0.009 | 0.002 | -5.56 | 2.64E-08 | -0.010 | 0.001 | -6.65 | 3.02E-11 | 3.73E-08 | 5.97E-01 | 4.01E-01 | ? |
| rs8088085-A | 18q21.2 | *MEX3C* | Known | 0.05 | 9.65E-01 | 5.45 | 4.79E-08 | 0.005 | 0.001 | 3.71 | 2.11E-04 | 0.007 | 0.001 | 5.21 | 1.88E-07 | 2.35E-05 | 7.99E-01 | 1.71E-01 | Not Shared |
| rs464901-T | 22q11.21 | *TUBA8* | Known | -1.06 | 2.89E-01 | 7.06 | 1.53E-12 | 0.006 | 0.002 | 4.17 | 3.11E-05 | 0.010 | 0.001 | 6.61 | 3.92E-11 | 4.41E-09 | 8.35E-01 | 1.63E-01 | Not Shared |
| rs133902-T | 22q12.1 | *MYO18B* | Known | 1.91 | 5.65E-02 | 6.16 | 9.14E-10 | 0.008 | 0.001 | 5.24 | 1.60E-07 | 0.009 | 0.001 | 6.13 | 8.78E-10 | 2.35E-06 | 6.34E-01 | 3.58E-01 | Not Shared |

### Online Table XIII. Gene Ontology of biological systems results for the gene sets of prioritized genes associated with atrial fibrillation risk exclusive.

| geneSet | description | link | size | overlap | expect | enrichmentRatio | pValue | FDR | overlapId | userId |
|---|---|---|---|---|---|---|---|---|---|---|
| GO:0002027 | regulation of heart rate | http://amig | 93 | 7 | 0.34 | 20.56 | 4.58E-08 | 1.64E-04 | 845;3752;4624;4 | SCN10A;SCN5A;KCND3;MYH6;MYH7;CASQ2;YWHAE |
| GO:0044057 | regulation of system process | http://amig | 535 | 13 | 1.96 | 6.64 | 5.33E-08 | 1.64E-04 | 845;2626;3752;4 | SCN10A;SCN5A;KCND3;MYH6;MYH7;GTF2IRD2;GATA4;CASQ2;MYOCD;FBXO32;YWHAE;THRB;HAND2 |
| GO:0008016 | regulation of heart contraction | http://amig | 205 | 9 | 0.75 | 11.99 | 5.40E-08 | 1.64E-04 | 845;2626;3752;4 | SCN10A;SCN5A;KCND3;MYH6;MYH7;GATA4;CASQ2;YWHAE;THRB |
| GO:0090257 | regulation of muscle system process | http://amig | 222 | 9 | 0.81 | 11.07 | 1.07E-07 | 2.43E-04 | 845;2626;4625;6 | SCN10A;SCN5A;MYH7;GTF2IRD2;GATA4;CASQ2;MYOCD;FBXO32;HAND2 |
| GO:0060047 | heart contraction | http://amig | 231 | 9 | 0.85 | 10.64 | 1.50E-07 | 2.73E-04 | 845;2626;3752;4 | SCN10A;SCN5A;KCND3;MYH6;MYH7;GATA4;CASQ2;YWHAE;THRB |
| GO:0003015 | heart process | http://amig | 240 | 9 | 0.88 | 10.24 | 2.07E-07 | 3.14E-04 | 845;2626;3752;4 | SCN10A;SCN5A;KCND3;MYH6;MYH7;GATA4;CASQ2;YWHAE;THRB |
| GO:1903522 | regulation of blood circulation | http://amig | 248 | 9 | 0.91 | 9.91 | 2.74E-07 | 3.56E-04 | 845;2626;3752;4 | SCN10A;SCN5A;KCND3;MYH6;MYH7;GATA4;CASQ2;YWHAE;THRB |
| GO:0003012 | muscle system process | http://amig | 423 | 11 | 1.55 | 7.10 | 3.31E-07 | 3.75E-04 | 845;2626;3752;4 | SCN10A;SCN5A;KCND3;SSPN;MYH6;MYH7;GTF2IRD2;GATA4;CASQ2;MYOCD;HAND2 |
| GO:0060048 | cardiac muscle contraction | http://amig | 126 | 7 | 0.46 | 15.18 | 3.71E-07 | 3.75E-04 | 845;2626;3752;4 | SCN10A;SCN5A;KCND3;MYH6;MYH7;GATA4;CASQ2 |
| GO:0006941 | striated muscle contraction | http://amig | 160 | 7 | 0.59 | 11.95 | 1.86E-06 | 1.69E-03 | 845;2626;3752;4 | SCN10A;SCN5A;KCND3;MYH6;MYH7;GATA4;CASQ2 |
| GO:0006936 | muscle contraction | http://amig | 339 | 9 | 1.24 | 7.25 | 3.67E-06 | 3.03E-03 | 845;2626;3752;4 | SCN10A;SCN5A;KCND3;SSPN;MYH6;MYH7;GATA4;CASQ2;MYOCD |
| GO:0003231 | cardiac ventricle development | http://amig | 123 | 6 | 0.45 | 13.33 | 5.70E-06 | 4.32E-03 | 2626;4624;4625; | SCN5A;MYH6;MYH7;GATA4;MYOCD;HAND2 |
| GO:0048738 | cardiac muscle tissue development | http://amig | 200 | 7 | 0.73 | 9.56 | 8.14E-06 | 5.56E-03 | 2066;2626;4624; | MYO18B;MYH6;MYH7;GATA4;MYOCD;ERBB4;LRRC10 |
| GO:0030048 | actin filament-based movement | http://amig | 132 | 6 | 0.48 | 12.42 | 8.56E-06 | 5.56E-03 | 2626;3752;4624; | SCN5A;KCND3;MYH6;MYH7;GATA4;MYO1C |
| GO:0035051 | cardiocyte differentiation | http://amig | 141 | 6 | 0.52 | 11.62 | 1.25E-05 | 7.37E-03 | 2626;4624;9464; | MYO18B;MYH6;GATA4;MYOCD;LRRC10;HAND2 |
| GO:0043502 | regulation of muscle adaptation | http://amig | 83 | 5 | 0.30 | 16.46 | 1.30E-05 | 7.37E-03 | 4625;6331;9464; | SCN5A;MYH7;GTF2IRD2;FBXO32;HAND2 |
| GO:0006942 | regulation of striated muscle contraction | http://amig | 85 | 5 | 0.31 | 16.07 | 1.46E-05 | 7.79E-03 | 845;2626;4625;6 | SCN10A;SCN5A;MYH7;GATA4;CASQ2 |
| GO:0007512 | adult heart development | http://amig | 14 | 3 | 0.05 | 58.54 | 1.65E-05 | 8.34E-03 | 4624;4625;9464 | MYH6;MYH7;HAND2 |
| GO:0006937 | regulation of muscle contraction | http://amig | 151 | 6 | 0.55 | 10.85 | 1.85E-05 | 8.61E-03 | 845;2626;4625;6 | SCN10A;SCN5A;MYH7;GATA4;CASQ2;MYOCD |
| GO:0086009 | membrane repolarization | http://amig | 44 | 4 | 0.16 | 24.83 | 1.98E-05 | 8.61E-03 | 845;3752;6331;7 | SCN5A;KCND3;CASQ2;YWHAE |
| GO:0055002 | striated muscle cell development | http://amig | 153 | 6 | 0.56 | 10.71 | 1.99E-05 | 8.61E-03 | 845;1073;2626;4 | MYO18B;MYH6;GATA4;CASQ2;LRRC10;CFL2 |
| GO:0003205 | cardiac chamber development | http://amig | 164 | 6 | 0.60 | 9.99 | 2.94E-05 | 1.22E-02 | 2626;4624;4625; | SCN5A;MYH6;MYH7;GATA4;MYOCD;HAND2 |
| GO:0055001 | muscle cell development | http://amig | 166 | 6 | 0.61 | 9.87 | 3.15E-05 | 1.25E-02 | 845;1073;2626;4 | MYO18B;MYH6;GATA4;CASQ2;LRRC10;CFL2 |
| GO:0061337 | cardiac conduction | http://amig | 105 | 5 | 0.38 | 13.01 | 4.06E-05 | 1.51E-02 | 845;3752;6331;6 | SCN10A;SCN5A;KCND3;CASQ2;YWHAE |
| GO:0010927 | cellular component assembly involved in morphogene | http://amig | 106 | 5 | 0.39 | 12.89 | 4.25E-05 | 1.51E-02 | 845;1073;4624;9 | PHLDB2;MYH6;CASQ2;CFL2;ZPBP2 |
| GO:0051146 | striated muscle cell differentiation | http://amig | 260 | 7 | 0.95 | 7.35 | 4.42E-05 | 1.51E-02 | 845;1073;2626;4 | MYO18B;MYH6;GATA4;CASQ2;MYOCD;LRRC10;CFL2 |
| GO:0014706 | striated muscle tissue development | http://amig | 357 | 8 | 1.31 | 6.12 | 4.49E-05 | 1.51E-02 | 1073;2066;2626; | MYO18B;MYH6;MYH7;GATA4;MYOCD;ERBB4;LRRC10;CFL2 |
| GO:0030029 | actin filament-based process | http://amig | 715 | 11 | 2.62 | 4.20 | 4.94E-05 | 1.54E-02 | 845;1073;1398;2 | SCN5A;PHLDB2;KCND3;MYH6;MYH7;GATA4;CASQ2;EPHA3;CRK;MYO1C;CFL2 |
| GO:0008015 | blood circulation | http://amig | 471 | 9 | 1.72 | 5.22 | 5.01E-05 | 1.54E-02 | 845;2626;3752;4 | SCN10A;SCN5A;KCND3;MYH6;MYH7;GATA4;CASQ2;YWHAE;THRB |
| GO:0070252 | actin-mediated cell contraction | http://amig | 110 | 5 | 0.40 | 12.42 | 5.07E-05 | 1.54E-02 | 2626;3752;4624; | SCN5A;KCND3;MYH6;MYH7;GATA4 |
| GO:0055007 | cardiac muscle cell differentiation | http://amig | 111 | 5 | 0.41 | 12.31 | 5.29E-05 | 1.55E-02 | 2626;4624;8470 | MYO18B;MYH6;GATA4;MYOCD;LRRC10 |
| GO:0003013 | circulatory system process | http://amig | 480 | 9 | 1.76 | 5.12 | 5.80E-05 | 1.62E-02 | 845;2626;3752;4 | SCN10A;SCN5A;KCND3;MYH6;MYH7;GATA4;CASQ2;YWHAE;THRB |
| GO:0060537 | muscle tissue development | http://amig | 371 | 8 | 1.36 | 5.89 | 5.88E-05 | 1.62E-02 | 1073;2066;2626; | MYO18B;MYH6;MYH7;GATA4;MYOCD;ERBB4;LRRC10;CFL2 |
| GO:0086001 | cardiac muscle cell action potential | http://amig | 68 | 4 | 0.25 | 16.07 | 1.11E-04 | 2.97E-02 | 3752;6331;6336; | SCN10A;SCN5A;KCND3;YWHAE |
| GO:0055117 | regulation of cardiac muscle contraction | http://amig | 71 | 4 | 0.26 | 15.39 | 1.31E-04 | 3.40E-02 | 845;2626;6331;6 | SCN10A;SCN5A;GATA4;CASQ2 |
| GO:0007507 | heart development | http://amig | 536 | 9 | 1.96 | 4.59 | 1.35E-04 | 3.40E-02 | 2066;2626;4624; | SCN5A;MYO18B;MYH6;MYH7;GATA4;MYOCD;ERBB4;LRRC10;HAND2 |
| GO:0003208 | cardiac ventricle morphogenesis | http://amig | 72 | 4 | 0.26 | 15.18 | 1.39E-04 | 3.41E-02 | 2626;4624;4625; | MYH6;MYH7;GATA4;HAND2 |
| GO:0055013 | cardiac muscle cell development | http://amig | 76 | 4 | 0.28 | 14.38 | 1.71E-04 | 4.10E-02 | 2626;4624;8470 | MYO18B;MYH6;GATA4;LRRC10 |
| GO:0086019 | cell-cell signaling involved in cardiac conduction | http://amig | 30 | 3 | 0.11 | 27.32 | 1.77E-04 | 4.12E-02 | 845;6331;6336 | SCN10A;SCN5A;CASQ2 |
| GO:0086029 | Purkinje myocyte to ventricular cardiac muscle cell sig | http://amig | 6 | 2 | 0.02 | 91.06 | 1.96E-04 | 4.34E-02 | 845;6331 | SCN5A;CASQ2 |
| GO:0086068 | Purkinje myocyte to ventricular cardiac muscle cell co | http://amig | 6 | 2 | 0.02 | 91.06 | 1.96E-04 | 4.34E-02 | 845;6331 | SCN5A;CASQ2 |
| GO:0055006 | cardiac cell development | http://amig | 81 | 4 | 0.30 | 13.49 | 2.19E-04 | 4.63E-02 | 2626;4624;8470 | MYO18B;MYH6;GATA4;LRRC10 |
| GO:0003300 | cardiac muscle hypertrophy | http://amig | 81 | 4 | 0.30 | 13.49 | 2.19E-04 | 4.63E-02 | 2626;4624;4625; | MYH6;MYH7;GATA4;HAND2 |
| GO:0042692 | muscle cell differentiation | http://amig | 338 | 7 | 1.24 | 5.66 | 2.26E-04 | 4.67E-02 | 845;1073;2626;4 | MYO18B;MYH6;GATA4;CASQ2;MYOCD;LRRC10;CFL2 |
| GO:0060306 | regulation of membrane repolarization | http://amig | 33 | 3 | 0.12 | 24.83 | 2.35E-04 | 4.75E-02 | 845;6331;7531 | SCN5A;CASQ2;YWHAE |
| GO:0014897 | striated muscle hypertrophy | http://amig | 83 | 4 | 0.30 | 13.17 | 2.41E-04 | 4.75E-02 | 2626;4624;4625; | MYH6;MYH7;GATA4;HAND2 |
| GO:0014896 | muscle hypertrophy | http://amig | 85 | 4 | 0.31 | 12.86 | 2.64E-04 | 4.97E-02 | 2626;4624;4625; | MYH6;MYH7;GATA4;HAND2 |
| GO:0086028 | bundle of His cell to Purkinje myocyte signaling | http://amig | 7 | 2 | 0.03 | 78.05 | 2.74E-04 | 4.97E-02 | 6331;6336 | SCN10A;SCN5A |
| GO:0086043 | bundle of His cell action potential | http://amig | 7 | 2 | 0.03 | 78.05 | 2.74E-04 | 4.97E-02 | 6331;6336 | SCN10A;SCN5A |
| GO:0060371 | regulation of atrial cardiac muscle cell membrane dep | http://amig | 7 | 2 | 0.03 | 78.05 | 2.74E-04 | 4.97E-02 | 6331;6336 | SCN10A;SCN5A |

**Online Table XIV. Gene Ontology of biological systems results for the gene sets of prioritized genes associated with atrial fibrillation risk that do not overlap with those associated with CES.**

| geneSet | description | link | size | overlap | expect | enrichmentRatio | pValue | FDR | overlapId | userId |
|---|---|---|---|---|---|---|---|---|---|---|
| GO:0003231 | cardiac ventricle de | http://amigo.gen | 123 | 6 | 0.45 | 13.33 | 5.70E-06 | 4.32E-03 | 2626;4624;4625; | SCN5A;MYH6;MYH7;GATA4;MYOCD;HAND2 |
| GO:0048738 | cardiac muscle tiss | http://amigo.gen | 200 | 7 | 0.73 | 9.56 | 8.14E-06 | 5.56E-03 | 2066;2626;4624; | MYO18B;MYH6;MYH7;GATA4;MYOCD;ERBB4;LRRC10 |
| GO:0035051 | cardiocyte different | http://amigo.gen | 141 | 6 | 0.52 | 11.62 | 1.25E-05 | 7.37E-03 | 2626;4624;9464; | MYO18B;MYH6;GATA4;MYOCD;LRRC10;HAND2 |
| GO:0043502 | regulation of muscl | http://amigo.gen | 83 | 5 | 0.30 | 16.46 | 1.30E-05 | 7.37E-03 | 4625;6331;9464; | SCN5A;MYH7;GTF2IRD2;FBXO32;HAND2 |
| GO:0007512 | adult heart develop | http://amigo.gen | 14 | 3 | 0.05 | 58.54 | 1.65E-05 | 8.34E-03 | 4624;4625;9464 | MYH6;MYH7;HAND2 |
| GO:0055002 | striated muscle cell | http://amigo.gen | 153 | 6 | 0.56 | 10.71 | 1.99E-05 | 8.61E-03 | 845;1073;2626;4 | MYO18B;MYH6;GATA4;CASQ2;LRRC10;CFL2 |
| GO:0003205 | cardiac chamber de | http://amigo.gen | 164 | 6 | 0.60 | 9.99 | 2.94E-05 | 1.22E-02 | 2626;4624;4625; | SCN5A;MYH6;MYH7;GATA4;MYOCD;HAND2 |
| GO:0055001 | muscle cell develop | http://amigo.gen | 166 | 6 | 0.61 | 9.87 | 3.15E-05 | 1.25E-02 | 845;1073;2626;4 | MYO18B;MYH6;GATA4;CASQ2;LRRC10;CFL2 |
| GO:0010927 | cellular component | http://amigo.gen | 106 | 5 | 0.39 | 12.89 | 4.25E-05 | 1.51E-02 | 845;1073;4624;9 | PHLDB2;MYH6;CASQ2;CFL2;ZPBP2 |
| GO:0051146 | striated muscle cell | http://amigo.gen | 260 | 7 | 0.95 | 7.35 | 4.42E-05 | 1.51E-02 | 845;1073;2626;4 | MYO18B;MYH6;GATA4;CASQ2;MYOCD;LRRC10;CFL2 |
| GO:0014706 | striated muscle tiss | http://amigo.gen | 357 | 8 | 1.31 | 6.12 | 4.49E-05 | 1.51E-02 | 1073;2066;2626; | MYO18B;MYH6;MYH7;GATA4;MYOCD;ERBB4;LRRC10;CFL2 |
| GO:0055007 | cardiac muscle cell | http://amigo.gen | 111 | 5 | 0.41 | 12.31 | 5.29E-05 | 1.55E-02 | 2626;4624;8470 | MYO18B;MYH6;GATA4;MYOCD;LRRC10 |
| GO:0060537 | muscle tissue deve | http://amigo.gen | 371 | 8 | 1.36 | 5.89 | 5.88E-05 | 1.62E-02 | 1073;2066;2626; | MYO18B;MYH6;MYH7;GATA4;MYOCD;ERBB4;LRRC10;CFL2 |
| GO:0003208 | cardiac ventricle m | http://amigo.gen | 72 | 4 | 0.26 | 15.18 | 1.39E-04 | 3.41E-02 | 2626;4624;4625; | MYH6;MYH7;GATA4;HAND2 |
| GO:0055013 | cardiac muscle cell | http://amigo.gen | 76 | 4 | 0.28 | 14.38 | 1.71E-04 | 4.10E-02 | 2626;4624;8470 | MYO18B;MYH6;GATA4;LRRC10 |
| GO:0086029 | Purkinje myocyte to | http://amigo.gen | 6 | 2 | 0.02 | 91.06 | 1.96E-04 | 4.34E-02 | 845;6331 | SCN5A;CASQ2 |
| GO:0086068 | Purkinje myocyte to | http://amigo.gen | 6 | 2 | 0.02 | 91.06 | 1.96E-04 | 4.34E-02 | 845;6331 | SCN5A;CASQ2 |
| GO:0055006 | cardiac cell develop | http://amigo.gen | 81 | 4 | 0.30 | 13.49 | 2.19E-04 | 4.63E-02 | 2626;4624;8470 | MYO18B;MYH6;GATA4;LRRC10 |
| GO:0003300 | cardiac muscle hyp | http://amigo.gen | 81 | 4 | 0.30 | 13.49 | 2.19E-04 | 4.63E-02 | 2626;4624;4625; | MYH6;MYH7;GATA4;HAND2 |
| GO:0042692 | muscle cell differen | http://amigo.gen | 338 | 7 | 1.24 | 5.66 | 2.26E-04 | 4.67E-02 | 845;1073;2626;4 | MYO18B;MYH6;GATA4;CASQ2;MYOCD;LRRC10;CFL2 |
| GO:0014897 | striated muscle hyp | http://amigo.gen | 83 | 4 | 0.30 | 13.17 | 2.41E-04 | 4.75E-02 | 2626;4624;4625; | MYH6;MYH7;GATA4;HAND2 |
| GO:0014896 | muscle hypertrophy | http://amigo.gen | 85 | 4 | 0.31 | 12.86 | 2.64E-04 | 4.97E-02 | 2626;4624;4625; | MYH6;MYH7;GATA4;HAND2 |
| GO:0086028 | bundle of His cell to | http://amigo.gen | 7 | 2 | 0.03 | 78.05 | 2.74E-04 | 4.97E-02 | 6331;6336 | SCN10A;SCN5A |
| GO:0086043 | bundle of His cell a | http://amigo.gen | 7 | 2 | 0.03 | 78.05 | 2.74E-04 | 4.97E-02 | 6331;6336 | SCN10A;SCN5A |
| GO:0060371 | regulation of atrial c | http://amigo.gen | 7 | 2 | 0.03 | 78.05 | 2.74E-04 | 4.97E-02 | 6331;6336 | SCN10A;SCN5A |

#### 4. SUPPLEMENTAL ACKNOWLEDGMENTS

The MEGASTROKE project received funding from the sources specified at http://www.megastroke.org/acknowledgments.html.

Acknowledgement of the contributing studies and databases of the MEGASTROKE project that made the GWAS summary data available. We gratefully acknowledge the assistance and contributions of Rainer Malik, Ganesh Chauhan, Matthew Traylor, Muralidharan Sargurupremraj, Yukinori Okada, Aniket Mishra, Loes Rutten-Jacobs, Anne-Katrin Giese, Sander W. van der Laan, Solveig Gretarsdottir, Christopher D. Anderson, Michael Chong, Hieab H. H. Adams, Tetsuro Ago, Peter Almgren, Philippe Amouyel, Hakan Ay, Traci M. Bartz, Oscar R. Benavente, Steve Bevan, Giorgio B. Boncoraglio, Robert D. Brown, Jr, Adam S. Butterworth, Caty Carrera, Cara L. Carty, Daniel I. Chasman, Wei-Min Chen, John W. Cole, Adolfo Correa, Ioana Cotlarciuc, Carlos Cruchaga, John Danesh, Paul I. W. de Bakker, Anita L. DeStefano, Marcel den Hoed, Qing Duan, Stefan T. Engelter, Guido J. Falcone, Rebecca F. Gottesman, Raji P. Grewal, Vilmundur Gudnason, Stefan Gustafsson, Jeffrey Haessler, Tamara B. Harris, Ahamad Hassan, Aki S. Havulinna, Susan R. Heckbert, Elizabeth G. Holliday, George Howard, Fang-Chi Hsu, Hyacinth I. Hyacinth, M. Arfan Ikram, Erik ingelsson, Marguerite R. Irvin, Xueqiu Jian, Jordi Jimenez-Conde, Julie A. Johnson, J. Wouter Jukema, Masahiro Kanai, Keith L. Keene, Brett M. Kissela, Dawn O. Kleindorfer, Charles Kooperberg, Michiaki Kubo, Leslie A. Lange, Carl D. Langefeld, Claudia Langenberg, Lenore J. Launer, Jin-Moo Lee, Robin Lemmens, Didier Leys, Cathryn M. Lewis, Wei-Yu Lin, Arne G. Lindgren, Erik Lorentzen, Patrik K. Magnusson, Jane Maguire, Ani Manichaikul, Patrick F. McArdle, James F. Meschia, Braxton D. Mitchell, Thomas H. Mosley, Michael A. Nalls, Toshiharu Ninomiya, Martin J. O'Donnell, Bruce M. Psaty, Sara L. Pulit, Kristiina Rannikmäe, Alexander P. Reiner, Kathryn M. Rexrode, Kenneth Rice, Stephen S. Rich, Paul M. Ridker, Natalia S. Rost, Peter M. Rothwell, Jerome I. Rotter, Tatjana Rundek, Ralph L. Sacco, Saori Sakaue, Michele M. Sale, Veikko Salomaa, Bishwa R. Sapkota, Reinhold Schmidt, Carsten O. Schmidt, Ulf Schminke, Pankaj Sharma, Agnieszka Slowik, Cathie L. M. Sudlow, Christian Tanislav, Turgut Tatlisumak, Kent D. Taylor, Vincent N. S. Thijs, Gudmar Thorleifsson, Unnur Thorsteinsdottir, Steffen Tiedt, Stella Trompet, Christophe Tzourio, Cornelia M. van Duijn, Matthew Walters, Nicholas J. Wareham, Sylvia Wassertheil-Smoller, James G. Wilson, Kerri L. Wiggins, Qiong Yang, Salim Yusuf, AFGen Consortium, Cohorts for Heart and Aging Research in Genomic Epidemiology (CHARGE) Consortium, International Genomics of Blood Pressure (iGEN-BP) Consortium, INVENT Consortium, STARNET, Joshua C. Bis, Tomi Pastinen, Arno Ruusalepp, Eric E. Schadt, Simon Koplev, Johan L. M. Björkegren, Veronica Codoni, Mete Civelek, Nicholas L. Smith, David A. Tregouet, Ingrid E. Christophersen, Carolina Roselli, Steven A. Lubitz, Patrick T. Ellinor, E. Shyong Tai, Jaspal S. Kooner, Norihiro Kato, Jiang He, Pim van der Harst, Paul Elliott, John C. Chambers, Fumihiko Takeuchi, Andrew D. Johnson, BioBank Japan Cooperative Hospital Group, COMPASS Consortium, EPiC-CVD Consortium, EPiC-interAct Consortium, International Stroke Genetics Consortium (ISGC), METASTROKE Consortium, Neurology Working Group of the CHARGE Consortium, NiNDS Stroke Genetics Network (SiGN), UK Young Lacunar DNA Study, MEGASTROKE Consortium, Dharambir K. Sanghera, Olle Melander, Christina Jern, Daniel Strbian, Israel Fernandez-Cadenas, W. T. Longstreth, Jr, Arndt Rolfs, Jun Hata, Daniel Woo, Jonathan Rosand, Guillaume Pare, Jemma C. Hopewell, Danish Saleheen, Kari Stefansson, Bradford B. Worrall, Steven J. Kittner, Sudha Seshadri, Myriam Fornage, Hugh S. Markus, Joanna M. M. Howson, Yoichiro Kamatani, Stephanie Debette, and Martin Dichgans.
